# The temporal relationship between preeclampsia and heart failure

**DOI:** 10.1101/2023.07.28.23293281

**Authors:** Charlotte Baker, Peter D Smits, Patricia Rodriguez, Brianna M Goodwin Cartwright, Samuel Gratzl, Rajdeep Brar, Emily Garrett, Nicholas L Stucky

## Abstract

**Importance:** Hypertensive disorders of pregnancy (e.g., preeclampsia) occur in approximately 8 to 10% of pregnancies in the US. Preeclampsia is a significant risk factor for future cardiovascular disease but in the US there is no universal long-term clinical follow up or screening for these conditions.

**Objective:** To determine whether having preeclampsia increases the rate of heart failure at or after delivery across demographics and obstetrics related comorbidities.

**Design:** Retrospective cohort study. We examined deliveries between January 1, 2018 and March 31, 2023 and assessed the time to heart failure stratified by preeclampsia status. Index event (T0) was first recorded delivery in time frame.

**Setting:** Population based using nationally representative data from electronic health records by Truveta

**Participants:** Female. 16 to 50 years of age. First recorded delivery in time frame identified using ICD-9-CM, ICD-10-CM, and SNOMED CT codes for delivery. Exclusion criteria: missing information on sex, race, ethnicity, age; if last contact with the health care system was T0; recent history of related cardiovascular conditions; or congenital heart disease.

**Exposure:** Preeclampsia on or before delivery.

**Main Outcomes and Measures:** Diagnosis of heart failure (HF) at or after T0. Other measures included were Leonard’s obstetric comorbidity score, age, race, ethnicity, and disability. We used Cox proportional hazards models to assess time till HF.

**Results:** 718,166 individuals were included and 14,204 had preeclampsia. After adjusting for obstetric comorbidities, race, ethnicity, age and disability, Black patients had a higher hazard of HF compared to white women (HR 3.48; CI 2.97, 4.06). Black patients with preeclampsia had a higher hazard of HF than white patients without preeclampsia (HR 4.63; CI 1.65, 12.99). Black patients without preeclampsia had a risk of HF similar to other racial groups with preeclampsia.

**Conclusions and Relevance:** The risk for HF after preeclampsia was significant and should continue to be explored. Modifiable factors for both preeclampsia and HF which should be a focus include substance use disorder and pregestational diabetes. Regular long term clinical follow up or screening of women with preeclampsia for HF may be warranted.

**Key Points:** *Question:* Does having preeclampsia increase the rate of heart failure at or after delivery across demographics and obstetrics related comorbidities?

*Findings:* In this retrospective cohort study that included 718,166 deliveries, the risk of heart failure at any point after delivery was 2.82 times greater in women that had preeclampsia than those that did not, after controlling for obstetric comorbidities, race, ethnicity, and age.

*Meaning:* Regular long term clinical follow up or screening of women with preeclampsia for heart failure may be warranted.

## Introduction

There are higher rates of poor maternal health outcomes in the US than in other high income countries ^1–3^ and these geographic disparities have only increased in the last decade ^4,5^. These maternal health outcomes are driven by a number of factors including social determinants of health, community resources, clinical practices, and individual behaviors ^6–8^. Legal restriction on the availability of elective pregnancy termination ^9–11^, diminishing availability of obstetric care even in urban areas ^12–14^, and bias against patients in medicine ^15–17^ are maternal health concerns most visible in US media. There is a need to identify and address these issues of maternal health concern in order to reverse these disparities and improve overall maternal health outcomes in the US.

One specific issue of concern is the incidence and subsequent health problems related to hypertensive disorders of pregnancy (HDP), namely preeclampsia and gestational hypertension, that occur in approximately 8 to 10% of pregnancies in the US ^18,19^. Preeclampsia, a pregnancy related condition characterized by high blood pressure, occurs most often during a person’s first pregnancy ^20,21^. More than 80% of pregnancies have at least one risk factor for HDP ^18^ such as age, pregestational hypertension, and pregestational diabetes ^20,22,23^. Preeclampsia can cause maternal seizure (eclampsia), stroke, hemorrhage, kidney and cardiac injury.

A type of cardiovascular disease, heart failure (HF), has been connected to having preeclampsia ^24–27^. HF risk factors are often preventable and modifiable but HF is not typically considered a potential health problem of otherwise healthy women of childbearing age. Recent work has suggested that women with HDP during or after pregnancy are at greater risk for having HF within and after 90 days post-pregnancy ^25^.

The purpose of this study is to determine whether having a diagnosis of preeclampsia or eclampsia (referred to hereforth as preeclampsia) at or before delivery increases the rate of HF after delivery across age, race, and ethnicity demographic characteristics and obstetrics related comorbidities.

## Methods

### Data Source

Clinical data was compiled from a subset of Truveta data and consisted of de-identified medical records representing ambulatory centers, hospitals, imaging centers, and clinics and medical offices. Truveta (https://www.truveta.com/) provides access to continuously updated and linked electronic health records (EHR) and claims data including demographics, diagnoses, encounters, immunizations, medications, laboratory results, procedures, clinical notes, and images. Truveta Data used in this study was accessed on 2023-07-25. Through syntactic normalization, similar data fields from different health care organizations are mapped to a common schema referred to as the Truveta Data Model (TDM). Once organized into common fields, the values are normalized to common ontologies such as ICD-10, SNOMED-CT, LOINC, RxNorm, and CVX, through semantic normalization. The normalization process employs an expert-led, artificial intelligence driven process to accomplish high-confidence modeling at scale. This study performs analysis of de-identified EHR data accessed via Truveta Studio. Truveta Studio only contains data that has been de-identified by expert determination in accordance with HIPAA Privacy Rule, and therefore this study was exempt from Institutional Review Board approval. Truveta data has been used in previous research ^28–30^ and is available to all Truveta subscribers at (https://studio.truveta.com).

### Study design and population

This observational cohort study follows the STROBE reporting guideline ^31^.

#### Eligibility criteria

Eligible female patients had a delivery between January 1, 2018 and March 31, 2023 and were 16 to 50 years of age. If patients had multiple deliveries within the time period, only the first was included. Patients were identified using ICD-9-CM, ICD-10-CM, and SNOMED CT codes for delivery (Supplement). The first delivery in the dataset was considered the index event, or time zero (T0).

Patients were excluded if they had missing information on sex, race, ethnicity, or age. Patients were also excluded if their last contact with the health care system was on the day of delivery. Consistent with previous literature, patients were excluded if they had congenital heart disease ^32^ or a history of HF, chronic kidney disease, pulmonary embolism, myocardial infarction, cardiomyopathy, or ischemic heart disease less than a year prior to delivery ^25^ as identified by diagnostic or billing codes (Supplement).

### Outcome

The outcome of interest was HF at or after T0. A binary yes/no HF variable was defined using ICD-9-CM, ICD-10-CM, and SNOMED CT codes.

### Exposure

The exposure of interest was preeclampsia at or before T0. A binary yes/no variable for preeclampsia was defined using ICD-9-CM, ICD-10-CM, and SNOMED CT codes.

### Covariates

We used Leonard’s validated obstetric comorbidity score ^33^ to control for indicators of severe maternal morbidity during and after delivery. Included comorbidities and the method to calculate the score can be found elsewhere ^33^, but were identified using ICD-9-CM, ICD-10-CM, and SNOMED CT codes (Supplement). We modified the obstetric comorbidity score to include eclampsia in the severe preeclampsia category. Pregestational diabetes (type 1 and type 2), body mass index (BMI) *≥*40, gestational diabetes, pregestational hypertension, and age were added to the model as covariates in addition to their inclusion in the obstetric comorbidity score. Age was assessed categorically (16-17, 18-24, 25-34, 35-44, and *≥*45). Race was categorized as Black or African American (henceforth referred to as Black), white, Asian, and other. Because of small population sizes that complicate statistical modeling, the “other” is a composite category of individuals who were categorized by the health systems as “other”, Native American and Alaska Native, or Native Hawaiian or other Pacific Islanders. Ethnicity was a binary variable of Hispanic and/or Latino and not Hispanic and/or Latino. We expanded Gleason’s ICD-9-CM definitions of intellectual, physical, and sensory disability ^34^ to include ICD-10-CM and SNOMED CT codes and created a binary yes/no disability variable.

All medical definitions used in this study were reviewed and approved by a clinical informaticist.

### Study Population Flowchart

**Figure 1:**
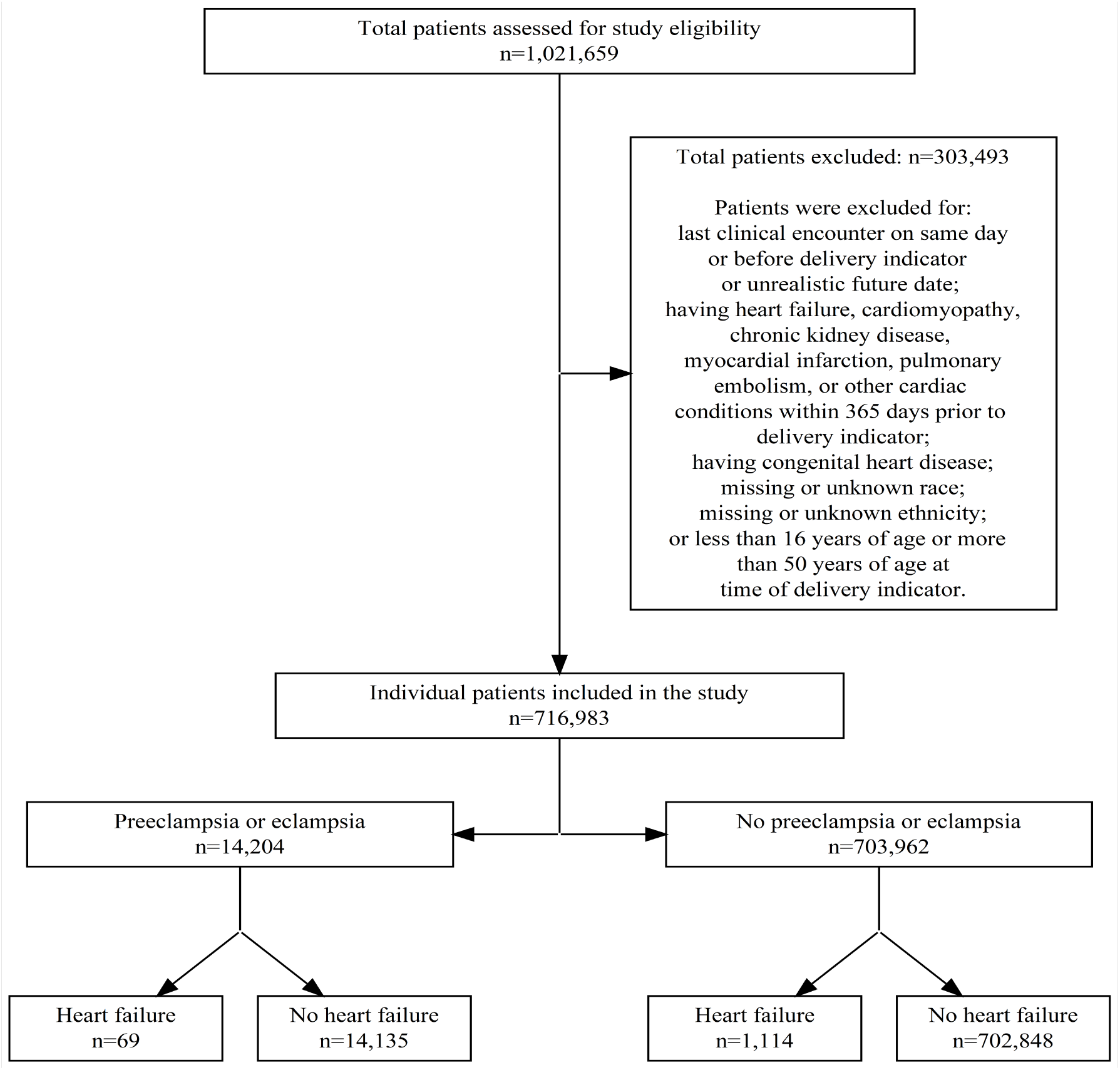
Cohort flowchart for the study of the risk of heart failure after preeclampsia or eclampsia between January 1, 2018 and March 31, 2023 in a subset of Truveta data. All medical conditions and diseases identified using ICD-9-CM, ICD-10-CM, and SNOMED CT codes (See study supplement).

### Statistical analysis

We calculated descriptive statistics to describe the study population. Due to right censoring, time to HF was assessed using Cox proportional hazards models. We completed two sets of analyses using the most recent encounter in the data as the censoring date. In the first, women were followed for 90 days until time of HF or their censoring, whichever occurred first. In the second, women were followed until the end of the available data until time of HF or their censoring, whichever occurred first. We plotted Kaplan-Meier curves of time to HF stratified by preeclampsia status and race.

For the survival analysis, all categorical covariates with more than 2 levels were transformed into multiple indicator variables, or dummy coded, with the most frequently occurring state being the reference state. Our first model included all of the covariates as additive effects, while our second model considered interaction terms between preeclampsia and race, preeclampsia and substance use disorder, preeclampsia and disability, substance use disorder and race, and disability and race.

All models were fit using the survival R package ^35,36^. We compared these models using Akaike Information Criterion (AIC)^37^, where the selected final model was that which had the lowest AIC value.

We used the final model to estimate model based hazard ratios comparing different combinations of preeclampsia, race (white or Black), substance use disorder (Yes or No), and disability status (Yes or No). These hazard ratios were estimated from marginal means calculated using the emmeans R package ^38^. This approach accounts for interaction effects which prevent the simple exponentiation of the regression coefficients and provides interpretable results. Statistical significance (*α*=0.05) of specific variables was determined using confidence intervals (not inclusive of 1.0) and p-values *≤* 0.05).

### Statistical programs and packages used

Statistical analysis was completed using Python and R software. Data preparation was completed using Python 3.10.5 ^39,40^ and packages pandas ^41,42^, NumPy ^43^, PyYAML ^44^, and matplotlib^45^. All analyses were conducted in R Version 4.1.3 ^46^ using packages dplyr^47^, ggplot2^48^, janitor^49^, gtsummary ^50^, broom ^51^, emmeans ^38^, scales^52^, MASS ^53^, xtable^54^, etm ^55^, survival^35,36^, and survminer ^56^. The study population flowchart was made using R Version 4.2.3 and packages xml2 ^57^, DiagrammeR ^58^, DiagrammeRsvg ^59^, and rsvg^60^.

## Results

### Description of study patients

There were 1,021,660 patients 16 to 50 years of age that had pregnancy indicators or a delivery on or after January 1, 2018 (Figure 1) that were considered for inclusion. Of that number, 303,493 people were excluded because of either 1) missing or unknown sex, race, ethnicity, and/or age; 2) no contact with the health system after T0 or beyond the date the data were obtained for the study (2023-07-25); 3) congenital heart disease; and/or 4) having a history of HF, chronic kidney disease, pulmonary embolism, myocardial infarction, cardiomyopathy, or ischemic heart disease between 0 and 365 days prior to T0. A total of 718,166 people met the inclusion criteria and were included in the study.

### Demographics

A total of 14,204 patients had preeclampsia at or before delivery (Table 1; Figure 1) and 69 of these patients had HF at or after delivery. A total of 703,962 patients did not have preeclampsia and 1,114 patients without preeclampsia were diagnosed with HF at or after delivery. Most people (57.3%) were between 25 and 34 years of age at the time of delivery. Black patients made up 13.7% of those without preeclampsia and 17.2% of those with preeclampsia. Asian patients made up 8.1% of those without preeclampsia and 6.4% of those with preeclampsia. White patients made up the majority of the study population (65.1%) and most of the study population was not Hispanic or Latino (81.1%).

**Table 1:** Demographic and comorbidity characteristics of people that delivered at or after January 1, 2018 to March 31, 2023 in a subset of Truveta data.

|  | No Preeclampsia/Eclampsia<br>(N=703,962) | Preeclampsia/Eclampsia<br>(N=14,204) | Overall<br>(N=718,166) |
| --- | --- | --- | --- |
| Heart Failure Event |  |  |  |
| No | 702,848 (99.8%) | 14,135 (99.5%) | 716,983 (99.8%) |
| Yes | 1,114 (0.2%) | 69 (0.5%) | 1,183 (0.2%) |
| Race |  |  |  |
| Black | 96,499 (13.7%) | 2,445 (17.2%) | 98,944 (13.8%) |
| Asian | 57,221 (8.1%) | 913 (6.4%) | 58,134 (8.1%) |
| White | 458,193 (65.1%) | 8,452 (59.5%) | 466,645 (65.0%) |
| Other Race | 92,049 (13.1%) | 2,394 (16.9%) | 94,443 (13.2%) |
| Ethnicity |  |  |  |
| Hispanic or Latino | 132,846 (18.9%) | 2,973 (20.9%) | 135,819 (18.9%) |
| Not Hispanic or Latino | 571,116 (81.1%) | 11,231 (79.1%) | 582,347 (81.1%) |
| Age Group |  |  |  |
| 0-17 | 7,372 (1.0%) | 104 (0.7%) | 7,476 (1.0%) |
| 18-24 | 121,109 (17.2%) | 2,485 (17.5%) | 123,594 (17.2%) |
| 25-34 | 403,675 (57.3%) | 7,756 (54.6%) | 411,431 (57.3%) |
| 35-44 | 160,597 (22.8%) | 3,733 (26.3%) | 164,330 (22.9%) |
| ≥45 | 11,209 (1.6%) | 126 (0.9%) | 11,335 (1.6%) |
| Bleeding Conditions |  |  |  |
| No | 692,071 (98.3%) | 13,591 (95.7%) | 705,662 (98.3%) |
| Yes | 11,891 (1.7%) | 613 (4.3%) | 12,504 (1.7%) |
| Pregestational Diabetes |  |  |  |
| No | 693,740 (98.5%) | 13,556 (95.4%) | 707,296 (98.5%) |
| Yes | 10,222 (1.5%) | 648 (4.6%) | 10,870 (1.5%) |
| Gestational Diabetes |  |  |  |
| No | 658,686 (93.6%) | 12,621 (88.9%) | 671,307 (93.5%) |
| Yes | 45,276 (6.4%) | 1,583 (11.1%) | 46,859 (6.5%) |
| Pregestational Hypertension |  |  |  |
| No | 696,451 (98.9%) | 13,076 (92.1%) | 709,527 (98.8%) |
| Yes | 7,511 (1.1%) | 1,128 (7.9%) | 8,639 (1.2%) |
| Other Preexisting Cardiac Conditions |  |  |  |
| No | 690,070 (98.0%) | 9,645 (67.9%) | 699,715 (97.4%) |
| Yes | 13,892 (2.0%) | 4,559 (32.1%) | 18,451 (2.6%) |
| Continued on next page |  |  |  |

| Feature | No Preeclampsia/Eclampsia | Preeclampsia/Eclampsia | Overall |
| --- | --- | --- | --- |
| BMI $\geq 40$ | | | |
| No | 689,056 (97.9%) | 13,424 (94.5%) | 702,480 (97.8%) |
| Yes | 14,906 (2.1%) | 780 (5.5%) | 15,686 (2.2%) |
| Substance Use Disorder |  |  |  |
| No | 639,677 (90.9%) | 12,220 (86.0%) | 651,897 (90.8%) |
| Yes | 64,285 (9.1%) | 1,984 (14.0%) | 66,269 (9.2%) |
| Disability |  |  |  |
| No | 624,955 (88.8%) | 12,036 (84.7%) | 636,991 (88.7%) |
| Yes | 79,007 (11.2%) | 2,168 (15.3%) | 81,175 (11.3%) |
| Obstetric Comorbidity Index |  |  |  |
| Mean (SD) | 19.7 (23.7) | 49.0 (47.7) | 20.2 (24.8) |
| Median [Min, Max] | 12.0 [0, 256] | 38.0 [0, 292] | 13.0 [0, 292] |

32.1% of patients with preeclampsia and 2.0% of patients without preeclampsia had a preexisting heart condition at least a year before their delivery. These conditions included but were not limited to hypertensive crisis, cardiac arrhythmia in childbirth, acute pericarditis, and hypertensive chronic kidney disease (Table 1). The prevalence of pregestational hypertension was 1.1% of patients without preeclampsia and 7.9% of patients with preeclampsia.

### Time to heart failure after delivery

The differences in prevalence seen in Table 1 carried over to analyses of time to HF after delivery. We used Kaplan-Meier curves to compare differences in patient time to HF at any point between patients with preeclampsia and patients without preeclampsia (Figure 2 A) as well as time to HF within 90 days of delivery (Figure 2 B). The prevalence of HF is small so the Y-axes in the graph exclude 0-0.98 to highlight the differences. In both cases, there is a nearly immediate separation in the curves and patients with preeclampsia have greater hazard of having the event of HF than patients without preeclampsia. There were significant differences in time to HF stratified by race (Figure 3) and disability (not shown) based on the Kaplan-Meier curves. The hazard of HF for Black patients and Asian patients with and without preeclampsia approach each other between the 4- and 5-year marks.

**Figure 2:**
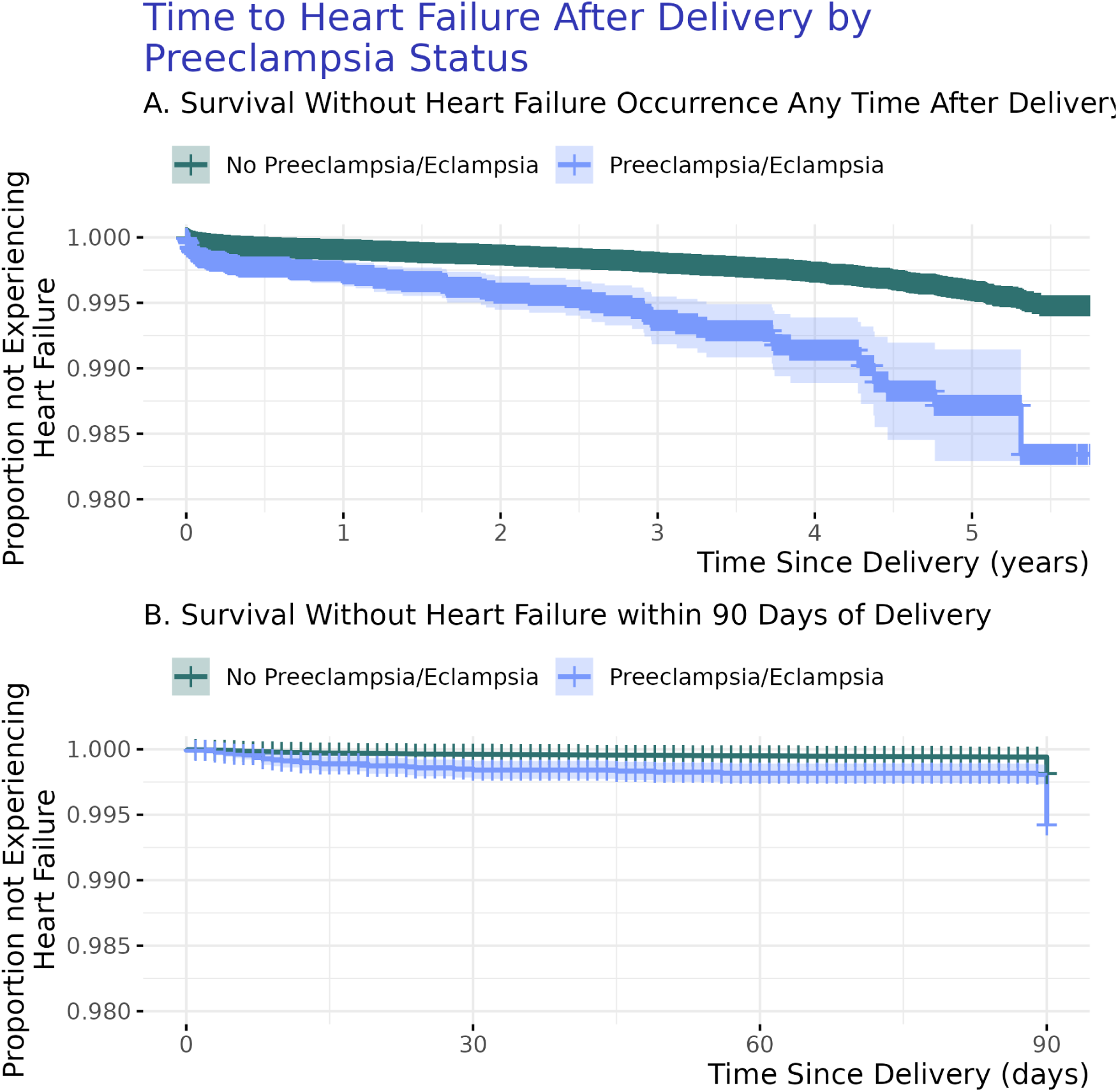
Time to Heart Failure After Delivery by Preeclampsia Status. Figure 2A displays the proportion of patients without heart failure at any point at or after delivery. Figure 2B displays the proportion of patients without heart failure within 90 days of delivery. In both figures (2A and 2B) the top line [green] is the proportion of patients without preeclampsia. The prevalence of HF is small so the Y-axes in the graph exclude 0-0.98 to highlight the differences. Graphs are not adjusted for additional factors.

**Figure 3:**
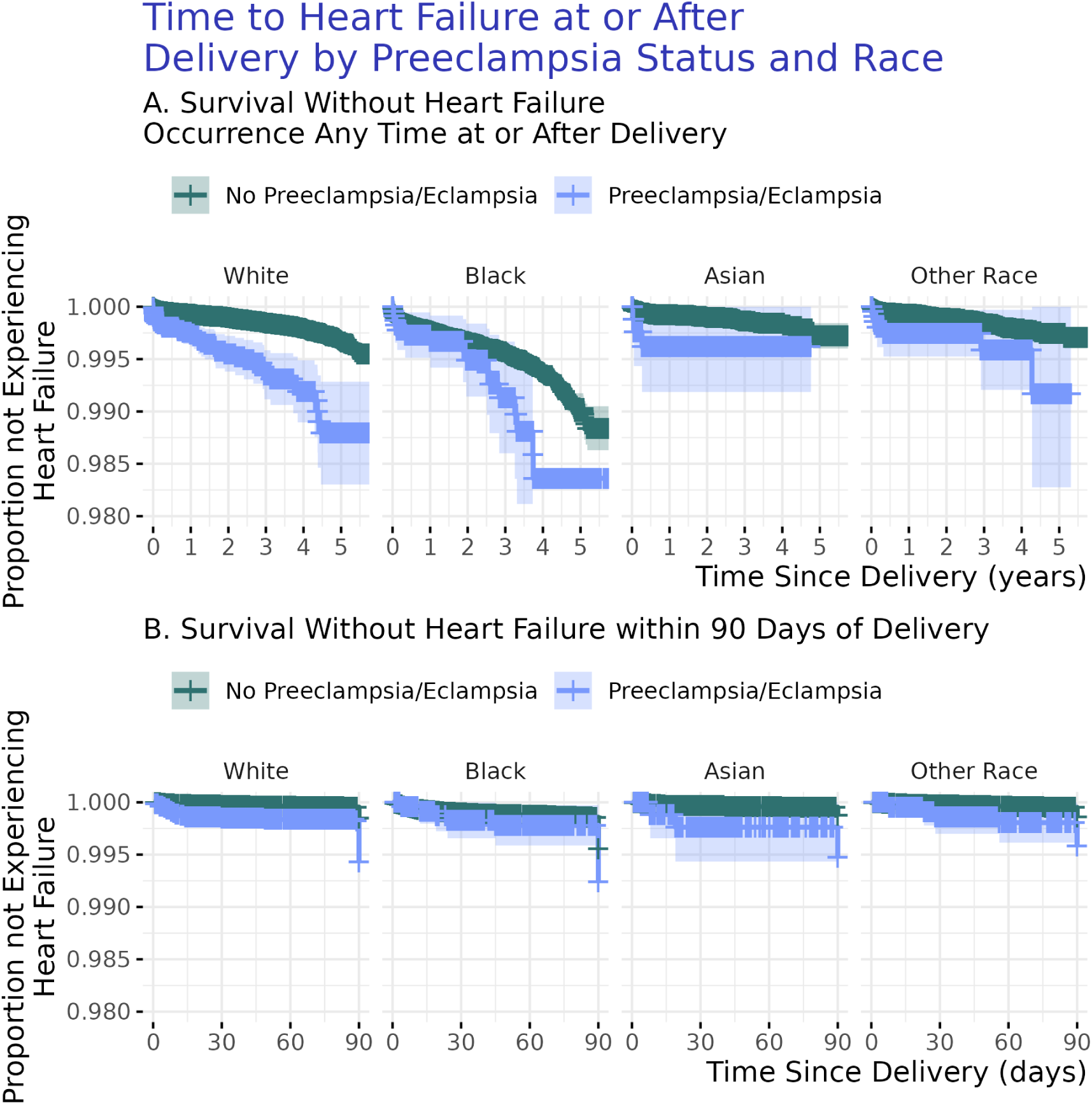
Time to Heart Failure After Delivery by Preeclampsia Status and Race. Figure 3A displays the proportion of patients without heart failure at any point at or after delivery. Figure 3B displays the proportion of patients without heart failure within 90 days of delivery. In both figures (3A and 3B) the top line [green] is the proportion of patients without preeclampsia. The prevalence of HF is small so the Y-axes in the graph exclude 0-0.98 to highlight the differences. Graphs are not adjusted for additional factors.

### Multivariate time to event analysis

We selected the Cox proportional hazards model of time till HF which featured multiple interaction terms as our final model because it had a lower AIC (29491.4) than the model which had only additive effects among the covariates (29504.9) (Supplement). The interaction model was used for all subsequent hazard ratio calculations (Table 2, Table 3, Supplement).

**Table 2:**
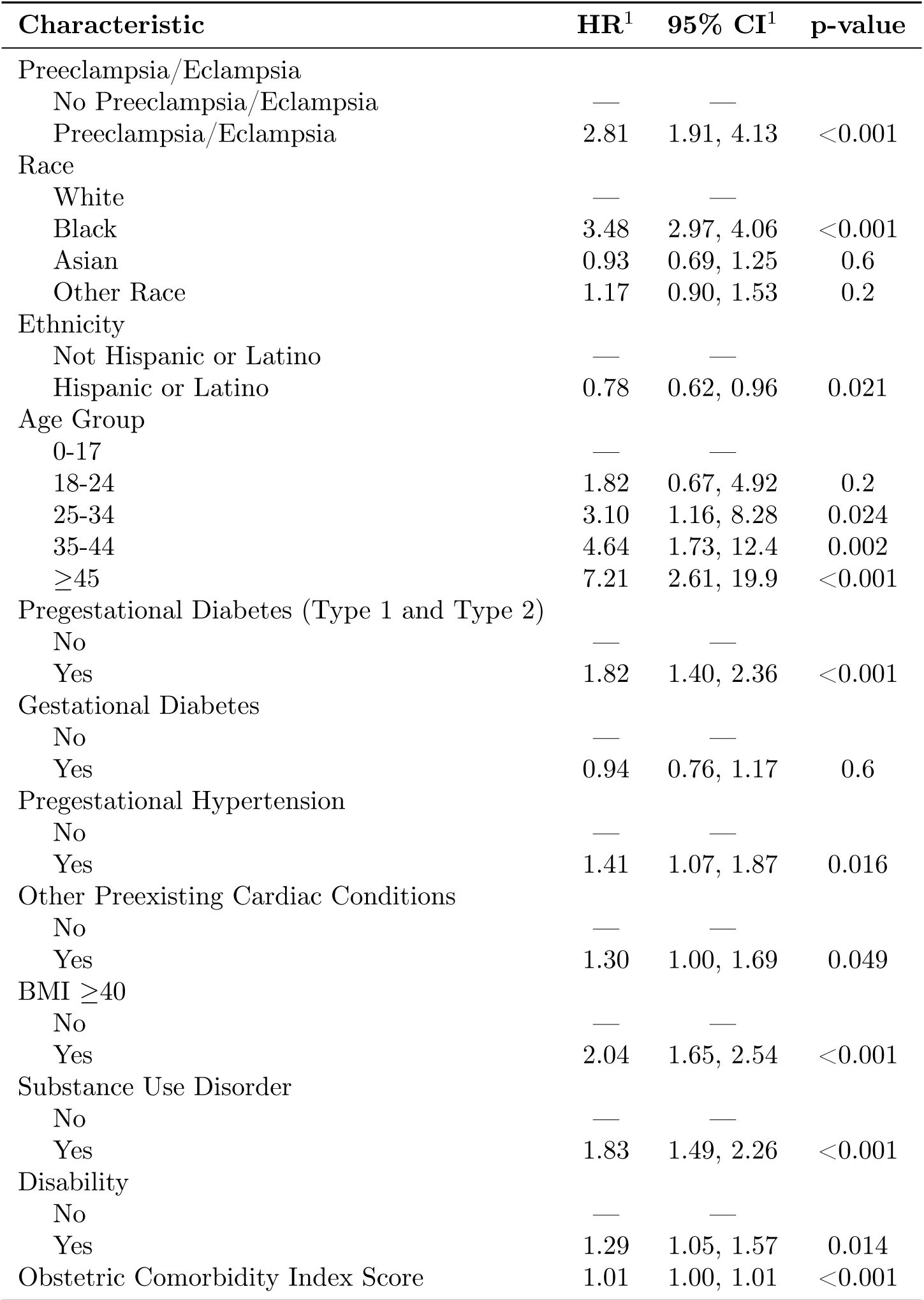
Hazard of heart failure after delivery for women with preeclampsia January 1, 2018 to March 31, 2023. ^1^HR = Hazard Ratio, CI = Confidence Interval.

**Table 3:** Hazard ratios associated comparisons between heart failure and different combinations of preclampsia/eclampsia status (Yes/No), substance use disorder (Yes/No), physical disability (Yes/No) and their interaction with race (white/Black). The state of each of these variables is shown in the [N]umerator and [D]enominator columns beneath each variable name. Hazard ratio is provided with 95% confidence intervals.

| Preeclampsia/Eclampsia |  | Race |  | Substance Use Disorder |  | Disabilities |  | Hazard Ratio | [95%] |
| --- | --- | --- | --- | --- | --- | --- | --- | --- | --- |
| N | D | N | D | N | D | N | D |  |  |
| Yes | Yes | Black | White | No | No | No | No | 1.65 | [0.53, 5.11] |
| Yes | Yes | Black | White | Yes | Yes | No | No | 0.97 | [0.28, 3.43] |
| Yes | Yes | Black | White | No | No | Yes | Yes | 1.23 | [0.35, 4.25] |
| Yes | Yes | Black | White | Yes | Yes | Yes | Yes | 0.72 | [0.20, 2.66] |
| Yes | No | Black | White | No | No | No | No | 4.63 | [1.65, 12.99] |
| Yes | No | Black | White | Yes | Yes | No | No | 1.59 | [0.33, 7.74] |
| Yes | No | Black | White | No | No | Yes | Yes | 1.92 | [0.41, 9.07] |
| Yes | No | Black | White | Yes | Yes | Yes | Yes | 0.66 | [0.11, 3.90] |
| Yes | No | White | White | No | No | No | No | 2.81 | [1.33, 5.92] |
| Yes | No | White | White | Yes | Yes | No | No | 1.63 | [0.49, 5.47] |
| Yes | No | White | White | No | No | Yes | Yes | 1.56 | [0.47, 5.23] |
| Yes | No | White | White | Yes | Yes | Yes | Yes | 0.91 | [0.23, 3.60] |
| Yes | No | Black | Black | No | No | No | No | 1.33 | [0.48, 3.70] |
| Yes | No | Black | Black | Yes | Yes | No | No | 0.78 | [0.18, 3.34] |
| Yes | No | Black | Black | No | No | Yes | Yes | 0.74 | [0.18, 3.12] |
| Yes | No | Black | Black | Yes | Yes | Yes | Yes | 0.43 | [0.08, 2.22] |
| No | No | Black | White | No | No | No | No | 3.48 | [2.57, 4.70] |
| No | No | Black | White | Yes | Yes | No | No | 2.05 | [1.10, 3.83] |
| No | No | Black | White | No | No | Yes | Yes | 2.59 | [1.43, 4.67] |
| No | No | Black | White | Yes | Yes | Yes | Yes | 1.52 | [0.76, 3.06] |

We found that the hazard of HF was greater in patients with preeclampsia during the index pregnancy compared with patients that did not have preeclampsia after adjusting for multiple factors including obstetric comorbidities, age, race, ethnicity, and BMI *≥*40 (HR 2.81; CI 1.91, 4.13) (Table 2). Black women have 3.48 (CI 2.97, 4.06) times the hazard of HF compared to white women (Table 2). Women with substance use disorder are 1.83 (CI 1.49, 2.26) times more likely than women without substance use disorder to have HF after delivery (Table 2). Women with disabilities have a higher hazard of HF (HR 1.29; CI 1.05, 1.57) than women without disabilities (Table 2).

When specifically comparing Black and white women, we found that Black women without preeclampsia have a hazard of HF 3.43 (CI 2.57, 4.70) times greater after delivery than white women without preeclampsia (Table 3, Supplement). In our population, the hazard of HF after delivery for Black women with preeclampsia compared to white women without preeclampsia was 4.63 times greater but had a wide confidence interval (CI 1.65,12.99) (Table 3, Supplement). The hazard of HF for white women with preeclampsia compared to those without was 2.81 (CI 1.35, 5.92) (Table 3, Supplement).

## Discussion

The purpose of this large retrospective cohort study was to determine whether having a diagnosis of preeclampsia at or before delivery increases the risk of HF at or after delivery across demographics, obstetrics related comorbidities, age, race and/or ethnicity. Even following resolution, preeclampsia is a significant risk factor for future cardiovascular disease, but there is no regular implementation of long term clinical follow up or screening in the US. Prior research has connected the risk of HF to preeclampsia ^24–27^, but more information is needed to assess the role of disparities in this outcome. In this study of 718,166 deliveries, the risk of HF at any point after delivery was greater in women that had preeclampsia than those that did not, after controlling for obstetrics comorbidities, race, ethnicity, and age.

The variables that we identified as the most important for the occurrence of HF are non-modifiable socio-demographic factors (e.g., race, ethnicity); and preventable or modifiable disease (e.g., substance use disorder, BMI *≥*40, pregestational diabetes)(Table 2, Table 3, Supplement). These findings are in line with previous research on preeclampsia ^18^, HF ^21^, and the combination of the preeclampsia and HF ^25^. Despite our findings, however, the etiology of the occurrence of HF after HDP, especially preeclampsia, is not completely explained. We identified pregestational diabetes, substance use disorder, and disability as important risk factors for HF after delivery (Table 2, Supplement). Despite a small number of cases of HF at or after delivery, we found that the risk for HF after preeclampsia was significant for all women and should continue to be explored.

It was clear in this analysis that race was significantly associated with both the occurrence of preeclampsia and the occurrence of HF when stratified by preeclampsia status. This was expected, especially given known disparities in maternal health outcomes in the US ^61,62^ and poor health outcomes for non-white populations in the US. Importantly, regardless of preeclampsia status, Black patients have similar risk of HF 5 or more years after delivery. Black patients without preeclampsia have risk of HF similar to other racial groups with preeclampsia. When considering HF and preeclampsia comorbidities, disparate outcomes existed for women with preeclampsia compared to those without preeclampsia within race categories. We also found that the risk of HF was higher for non-white women with substance use disorder (Supplement). There is a demonstrated connection between substance use disorder and HF hospitalizations ^63^. Programs already exist to address substance use disorder in childbearing persons and their children ^64^), but these results indicate that more can be done. Our results suggest that the combined effect of substance use disorder and preeclampsia on HF should be consistently included in further research.

### Strengths and Limitations

We did not include tobacco use, income, insurance type, urban/rural locality, parity, or previous preeclampsia, factors that are important in the discussion of HF after pregnancy. We also did not include any pregnant people that were below age 16 despite teenagers being at greater risk for preeclampsia than older persons. We did not include people that did not have a biological sex of female in the EHR. This study did not include two important clinical variables - severity of preeclampsia or gestational hypertension - outside of their inclusion in the obstetric comorbidity index. We acknowledge that the study results may be different when stratified by the severity of preeclampsia. We also measured pregnancy by using delivery as the indicator. This means that we may have undercounted the number of pregnant people, underestimated the risk of HF in people with preeclampsia, and underestimated the effect of repeat preeclampsia on risk of HF. We were unable to estimate any differences in outcome associated with caregiver type (e.g., physician, nurse practitioner, doula, midwife).

Our approach has multiple strengths. This study used a nationally representative dataset derived from EHR. The results could be considered generalizable for patients aged 16 to 50 that were white, Black, and Asian, but care should be given when applying these results to populations that are not specified here such as Native Hawaiian or Pacific Islander. Another strength of this study is the use of an obstetric specific comorbidity index over the Elixhauser or Charlson comorbidity indexes. While not containing all possible comorbidities for pregnancy, the care given to include gestational diabetes, advanced maternal age, placental abruption, anemia, and mental health issues among others makes this index better suited to account for comorbidities that are present or co-occur during pregnancy rather than those that occur with other life illnesses.

While using a large cohort is a strength, ours is subject to a variety of known limitations to EHR data. We are only able to identify events that are captured by the constituent health care systems that are a part of the Truveta member system. Events and comorbidity status may be misclassified when captured outside of member healthcare systems. These are common and well understood limitations associated with using EHR data ^65,66^. Finally, the number of exclusion criteria used in this study dramatically decreased the number of patients with preeclampsia. This directly affected the statistical outcomes of this study and may not represent the full effects of preeclampsia on HF after delivery for women with existing heart issues or newly diagnosed pregestational hypertension. Despite these limitations, our results still underscore the need to examine the relationship between preeclampsia and HF.

## Conclusion

The purpose of this large retrospective cohort study was to determine whether having a diagnosis of preeclampsia at or before delivery increases the risk of HF at or after delivery across demographics, obstetrics related comorbidities, age, race, and ethnicity. We found this to be the case for each of these factors. Regular long term clinical follow up or screening of women with preeclampsia for HF may be warranted. Future research should focus on the impact of obstetrics related comorbidities that are modifiable, the impact of society on these outcomes, the impact of parity and prior history of preeclampsia, and further examine the impact of different types of disability on the incidence of HDP and/or HF in pregnant women. We expect that each of these factors will help explain more of the disparity and provide guidance for clinical intervention over time.

## Acknowledgements

The authors thank the clinical informatics team for their assistance and input during the design and development of this study. We also thank the Truveta communications team for their ongoing support as we conduct research that saves lives with data.

## Institutional Review Board Approval

This study performs analysis of de-identified electronic health records (EHR) data accessed via Truveta Studio. Truveta Studio only contains data that has been de-identified by expert determination in accordance with HIPAA Privacy Rule, and therefore this study was exempt from Institutional Review Board approval.

## Data Availability Statement

Code necessary to generate all analyses and figures is included in a GitHub repository (https://github.com/Truveta/baker_et_al_preeclampsia_heart_failure).

