## Supplemental Material for "The temporal relationship between preeclampsia and heart failure"

#### Contents

**1 Supplemental tables** **2**
1.1 Table 1 - Akaike information criterion (AIC) Results . . . . . . . . . . 2
1.2 Multivariate Tables . . . . . . . . . . 2

**2 Data element definitions** **6**

### 1 Supplemental tables

#### 1.1 Table 1 - Akaike information criterion (AIC) Results

| | Modnames | AIC | Delta_AIC |
|---|---|---|---|
| 1 | Adjusted | 29491.38 | 0.00 |
| 2 | Unadjusted | 29504.95 | 13.56 |

#### 1.2 Multivariate Tables

Below is the fully adjusted cox proportional hazards model with exponentiated coefficients (Table 2. Table 3 provides this information in un-exponentiated form.

**Table 2. Complete model of the exponentiated hazard of heart failure after delivery for women with preeclampsia January 1, 2018 to March 31, 2023.**
[1]HR = Hazard Ratio, CI = Confidence Interval

| Characteristic | HR[1] | 95% CI[1] | p-value |
|---|---|---|---|
| Preeclampsia/Eclampsia | | | |
| No Preeclampsia/Eclampsia | — | — | |
| Preeclampsia/Eclampsia | 2.81 | 1.91, 4.13 | <0.001 |
| Race | | | |
| White | — | — | |
| Black | 3.48 | 2.97, 4.06 | <0.001 |
| Asian | 0.93 | 0.69, 1.25 | 0.6 |
| Other Race | 1.17 | 0.90, 1.53 | 0.2 |
| Ethnicity | | | |
| Not Hispanic or Latino | — | — | |
| Hispanic or Latino | 0.78 | 0.62, 0.96 | 0.021 |
| Age Group | | | |
| 0-17 | — | — | |
| 18-24 | 1.82 | 0.67, 4.92 | 0.2 |
| 25-34 | 3.10 | 1.16, 8.28 | 0.024 |
| 35-44 | 4.64 | 1.73, 12.4 | 0.002 |
| ≥45 | 7.21 | 2.61, 19.9 | <0.001 |
| Pregestational Diabetes (Type 1 and Type 2) | | | |
| No | — | — | |
| Yes | 1.82 | 1.40, 2.36 | <0.001 |
| Gestational Diabetes | | | |
| No | — | — | |
| Yes | 0.94 | 0.76, 1.17 | 0.6 |

**Table 2 – Continued on next page Table 2 – Continued**

| **Characteristic** | **HR[1]** | **95% CI[1]** | **p-value** |
|---|---|---|---|
| Pregestational Hypertension | | | |
| No | — | — | |
| Yes | 1.41 | 1.07, 1.87 | 0.016 |
| Other Preexisting Cardiac Conditions | | | |
| No | — | — | |
| Yes | 1.30 | 1.00, 1.69 | 0.049 |
| BMI ≥40 | | | |
| No | — | — | |
| Yes | 2.04 | 1.65, 2.54 | <0.001 |
| Substance Use Disorder | | | |
| No | — | — | |
| Yes | 1.83 | 1.49, 2.26 | <0.001 |
| Disability | | | |
| No | — | — | |
| Yes | 1.29 | 1.05, 1.57 | 0.014 |
| Obstetric Comorbidity Index Score | 1.01 | 1.00, 1.01 | <0.001 |
| Preeclampsia/Eclampsia * Race | | | |
| Preeclampsia/Eclampsia * Black | 0.47 | 0.26, 0.86 | 0.014 |
| Preeclampsia/Eclampsia * Asian | 1.00 | 0.34, 2.89 | >0.9 |
| Preeclampsia/Eclampsia * Other Race | 0.87 | 0.40, 1.93 | 0.7 |
| Race * Substance Use Disorder | | | |
| Black * Yes | 0.59 | 0.42, 0.83 | 0.002 |
| Asian * Yes | 1.13 | 0.44, 2.92 | 0.8 |
| Other Race * Yes | 1.33 | 0.81, 2.18 | 0.3 |
| Race * Disability | | | |
| Black * Yes | 0.74 | 0.54, 1.03 | 0.072 |
| Asian * Yes | 0.97 | 0.46, 2.03 | >0.9 |
| Other Race * Yes | 0.69 | 0.39, 1.22 | 0.2 |
| Preeclampsia/Eclampsia * Substance Use Disorder | | | |
| Preeclampsia/Eclampsia * Yes | 0.58 | 0.31, 1.08 | 0.086 |
| Preeclampsia/Eclampsia * Disability | | | |
| Preeclampsia/Eclampsia * Yes | 0.56 | 0.30, 1.03 | 0.063 |

**Table 3. Complete model of the un-exponentiated hazard of heart failure after delivery for women with preeclampsia January 1, 2018 to March 31, 2023.**

| Characteristic | log(HR)[1] | 95% CI[1] | p-value |
|---|---|---|---|
| Preeclampsia/Eclampsia | | | |
| No Preeclampsia/Eclampsia | — | — | |
| Preeclampsia/Eclampsia | 1.03 | 0.65, 1.42 | <0.001 |
| Race | | | |
| White | — | — | |
| Black | 1.25 | 1.09, 1.40 | <0.001 |
| Asian | -0.08 | -0.38, 0.23 | 0.6 |
| Other Race | 0.16 | -0.11, 0.43 | 0.2 |
| Ethnicity | | | |
| Not Hispanic or Latino | — | — | |
| Hispanic or Latino | -0.25 | -0.47, -0.04 | 0.021 |
| Age Group | | | |
| 0-17 | — | — | |
| 18-24 | 0.60 | -0.40, 1.59 | 0.2 |
| 25-34 | 1.13 | 0.15, 2.11 | 0.024 |
| 35-44 | 1.53 | 0.55, 2.52 | 0.002 |
| ≥45 | 1.98 | 0.96, 2.99 | <0.001 |
| Pregestational Diabetes (Type 1 and Type 2) | | | |
| No | — | — | |
| Yes | 0.60 | 0.34, 0.86 | <0.001 |
| Gestational Diabetes | | | |
| No | — | — | |
| Yes | -0.06 | -0.27, 0.16 | 0.6 |
| Pregestational Hypertension | | | |
| No | — | — | |
| Yes | 0.34 | 0.06, 0.62 | 0.016 |
| Other Preexisting Cardiac Conditions | | | |
| No | — | — | |
| Yes | 0.26 | 0.00, 0.53 | 0.049 |
| BMI ≥40 | | | |
| No | — | — | |
| Yes | 0.72 | 0.50, 0.93 | <0.001 |
| Substance Use Disorder | | | |
| No | — | — | |
| Yes | 0.61 | 0.40, 0.81 | <0.001 |
| Disability | | | |
| No | — | — | |
| Yes | 0.25 | 0.05, 0.45 | 0.014 |

**Table 3 – Continued on next page**

**Table 3 – Continued**

| **Characteristic** | **HR**[1] | **95% CI**[1] | **p-value** |
|---|---|---|---|
| Obstetric Comorbidity Index Score | 0.01 | 0.00, 0.01 | <0.001 |
| Preeclampsia/Eclampsia * Race | | | |
| Preeclampsia/Eclampsia * Black | -0.75 | 1.00, -0.15 | 0.014 |
| Preeclampsia/Eclampsia * Asian | 0.00 | 1.00, 1.06 | >0.9 |
| Preeclampsia/Eclampsia * Other Race | -0.13 | -0.92, 0.66 | 0.7 |
| Race * Substance Use Disorder | | | |
| Black * Yes | -0.53 | -0.87, -0.19 | 0.002 |
| Asian * Yes | 0.12 | -0.82, 1.07 | 0.8 |
| Other Race * Yes | 0.28 | -0.21, 0.78 | 0.3 |
| Race * Disability | | | |
| Black * Yes | -0.30 | -0.62, 0.03 | 0.072 |
| Asian * Yes | -0.03 | -0.77, 0.71 | >0.9 |
| Other Race * Yes | -0.37 | -0.95, 0.20 | 0.2 |
| Preeclampsia/Eclampsia * Substance Use Disorder | | | |
| Preeclampsia/Eclampsia * Yes | -0.54 | 1.00, 0.08 | 0.086 |
| Preeclampsia/Eclampsia * Disability | | | |
| Preeclampsia/Eclampsia * Yes | -0.59 | 1.00, 0.03 | 0.063 |

#### 2 Data element definitions

Below are the codes used for defining heart failure, delivery, preeclampsia/eclampsia, as well as for the comorbidities. All child codes are included even if not specified.

##### Heart Failure

ICD-10-CM: I11.0, I13.0, I13.2, I50.1, I50.20, I50.21, I50.22, I50.23, I50.30, I50.31, I50.32, I50.33, I50.40, I50.41, I50.42, I50.43, I50.81, I50.82, I50.83, I50.84, I50.89, I50.9, I50.810, I50.811, I50.812, I50.813, I50.814

SNOMED CT: 364006, 5148006, 5375005, 10091002, 10633002, 25544003, 82523003, 83105008, 84114007, 85232009, 88805009, 90727007, 92506005, 111283005, 871617000, 7411000175102, 15781000119107, 23341000119109, 67431000119105, 67441000119101, 72481000119103, 101281000119107, 120851000119104, 120861000119102, 120871000119108, 120881000119106, 120891000119109, 120901000119108, 153931000119109, 153941000119100, 153951000119103, 15629541000119106, 15629591000119103, 15629641000119107, 15629741000119102, 15964701000119109, 446221000, 471880001, 698296002, 698594003, 703272007, 703273002, 703274008, 703275009, 703276005, 717840005, 194767001, 194779001, 194781004, 195111005, 195112003, 195114002, 206586007, 314206003, 233924009, 42343007, 43736008, 44088000, 44313006, 46113002, 48447003, 56675007, 74960003, 410431009, 417996009, 418304008, 424404003, 426263006, 426611007, 441481004, 441530006, 443253003, 443254009, 443343001, 443344007, 1204206001, 1204203009, 1204204003, 1204200007, 1204462004, 1208846006, 1208848007, 1208850004, 1208843003, 1204468000, 1204460007, 5053004, 10335000, 16253001, 24841007, 33644002, 83291003, 89819002, 788950000, 7371000175103, 7381000175100, 7391000175102, 7401000175100, 7421000175106, 96311000119109, 285211000119102, 285221000119109, 16838951000119100, 13839000, 78862003, 445236007, 462172006, 462174007, 462175008, 609507007, 704242009, 722095005, 722919003, 724550005, 359617009, 367363000, 45650007, 49584005, 55565007, 60856006, 62377009, 66989003, 71892000, 79955004, 80479009, 426012001, 276514007, 277638005, 277639002, 1010444007, 1010447000, 15964661000119102, 898208007

ICD-9-CM: 402.01, 402.11, 402.91, 404.01, 404.03, 404.11, 404.13, 404.91, 404.93, 428

##### Preeclampsia or eclampsia

SNOMED CT: 15938005, 736992003, 736993008, 765182005, 198983002, 198984008, 198985009, 198986005, 198990007, 198991006, 198992004, 198993009, 198997005, 198999008, 199000005, 199002002, 199003007, 303063000, 237281009, 237282002, 237283007, 41114007, 46764007, 67359005, 69909000, 398254007, 1003729000, 10752641000119102, 95605009

ICD-10-CM: O11, O14, O15, O11.1, O11.2, O11.3, O11.4, O11.5, O11.9, O14.0, O14.00, O14.02, O14.03, O14.04, O14.05, O14.1, O14.10, O14.12, O14.13, O14.14, O14.15, O14.2, O14.20, O14.22, O14.23, O14.24, O14.25, O14.9, O14.90, O14.92, O14.93, O14.94, O14.95, O15.0, O15.00, O15.02, O15.03, O15.1, O15.2, O15.9

ICD-9-CM: 642.4, 642.5, 642.6, 642.7

#### Preeclampsia without severe features or gestational hypertension

ICD-10-CM: O13, O14.0, O14.9, O13.1, O13.2, O13.3, O13.4, O13.5, O13.9, O14.00, O14.02, O14.03, O14.04, O14.05, O14.90, O14.92, O14.93, O14.94, O14.95

ICD-9-CM: 642.3, 642.4, 642.7

SNOMED CT: 765182005, 41114007

#### Preeclampsia with severe features

ICD-10-CM: O11, O14.1, O14.2, O15, O11.1, O11.2, O11.3, O11.4, O11.5, O11.9, O14.10, O14.12, O14.13, O14.14, O14.15, O14.20, O14.22, O14.23, O14.24, O14.25, O15.0, O15.00, O15.02, O15.03, O15.1, O15.2, O15.9

SNOMED CT: 95605009, 198997005, 46764007, 198983002, 198984008, 198985009, 198986005, 198999008, 199000005, 199002002, 199003007

ICD-9-CM: 642.5, 642.60, 642.61, 642.63, 642.64

#### Cardiomyopathy

ICD-10-CM: B33.24, I25.5, I42, O90.3, I42.0, I42.1, I42.2, I42.3, I42.4, I42.5, I42.6, I42.7, I42.8, I42.9

ICD-9-CM: 414.8, 425, 674.54

SNOMED CT: 85898001, 195029002, 62377009, 1173036000, 1230303001, 1197430005, 1179293006, 1204194004, 1186866009, 1186718008, 1177176009, 1172839002, 1187642008, 1954004, 2024009, 2816000, 6022005, 8209004, 9909003, 11104006, 12563008, 15544002, 16253001, 20529002, 21430002, 24584003, 27097002, 28869005, 30496006, 30884007, 33006007, 33258008, 34342000, 83521008, 83978005, 84725008, 84733009, 86194008, 86995005, 89461002, 90202001, 90828009, 91604001, 93558006, 111285003, 1137377003, 1149109001, 1153452008, 788878000, 789037005, 860837007, 860839005, 871638006, 871639003, 871646007, 871649000, 111000119104, 108701000119102, 119731000146105, 204981000119101, 1086051000119107, 16228931000119102, 50270004, 53322007, 195030007, 445336009, 447934002, 447935001, 460500001, 471841009, 471846004, 471858004, 471870000, 471875005, 471885006, 471890009, 472100003, 472315005, 472316006, 472317002, 472318007, 472319004, 472320005, 472323007, 472324001, 472325000, 472326004, 472327008, 699668009, 700065003, 702424003, 704241002, 713523008, 715535009, 715626008, 715655000, 715865008, 717812000, 718713000, 719272007, 719380003, 719451006, 719824001, 719835006, 719839000, 720609003, 720610008, 721014007, 723993005, 724173009, 725027004, 725416005, 763280005, 766883006, 771478008, 771509001, 771513008, 774148007, 775908005, 195020003, 195021004, 195023001, 195025008, 195027000, 195031006, 195032004, 315608004, 233871002, 233873004, 233875006, 233877003, 233878008, 240077004, 253528005, 35719004, 35728003, 36471000, 39041004, 39693003, 40532006, 41964001, 42099002, 44630002, 45227007, 46696008, 46939000, 50275009, 52029003, 53043001, 53703001, 54830008, 56941009, 58629009, 58911001, 60514000, 63183009, 64041007, 64077000, 64615000, 64749001, 68602002, 69609002, 72972005, 74249003, 76931005, 79152002, 79754008, 398716006, 398754006, 399020009, 406428006, 413994008, 413995009, 415295002,

415509000, 419097006, 426300009, 426856002, 441541008, 880014009, 880038008, 880040003, 880042006, 890119003, 890120009, 890121008, 890122001

#### Chronic kidney disease

ICD-10-CM: D63.1, E08.22, E09.22, E13.22, I12, I12.0, I12.9, I13, I13.0, I13.1, I13.10, I13.11, I13.2, N18, N18.1, N18.2, N18.3, N18.4, N18.5, N18.6, N18.9, O10.211, O10.212, O10.213, O10.219, O10.22, O10.31, O10.311, O10.312, O10.313, O10.319, O10.32, O10.33, Q61.2, Q61.3, Q61.8, N18.30, N18.31, N18.32, Z94.0

ICD-9-CM: 585

SNOMED CT: 709044004, 1208934006, 16726004, 90688005, 711000119100, 721000119107, 731000119105, 741000119101, 751000119104, 771000119108, 1801000119106, 8501000119104, 71421000119105, 71701000119105, 90721000119101, 90731000119103, 90741000119107, 90751000119109, 90761000119106, 90771000119100, 90791000119104, 96441000119101, 96701000119107, 96711000119105, 96721000119103, 96731000119100, 96741000119109, 96751000119106, 104931000119100, 111411000119103, 117681000119102, 118781000119108, 120261000119101, 127991000119101, 128001000119105, 129151000119102, 129161000119100, 129171000119106, 129181000119109, 140101000119109, 140111000119107, 140121000119100, 140131000119102, 153851000119106, 153891000119101, 284961000119106, 284971000119100, 284981000119102, 284991000119104, 285001000119105, 285011000119108, 285041000119107, 285061000119106, 285081000119102, 285101000119109, 285831000119108, 285841000119104, 285851000119102, 285861000119100, 285871000119106, 285881000119109, 285911000119109, 285921000119102, 286371000119107, 434431000124103, 449631000124102, 691421000119108, 10757401000119104, 10757481000119107, 700378005, 700379002, 704667004, 707324008, 708975004, 712487000, 713313000, 714152005, 714153000, 722098007, 722149000, 722150000, 722467000, 723190009, 776416004, 236433006, 236434000, 236435004, 236436003, 236552002, 46177005, 49708008, 57557005, 425369003, 431855005, 431856006, 431857002, 433144002, 433146000, 368421000119108, 368431000119106, 368441000119102, 368451000119100, 368461000119103, 368471000119109, 897308007, 897310009, 897311008, 897312001, 1217070004

#### Ischemic heart disease

ICD-10-CM: I23.0, I23.1, I23.2, I23.3, I23.4, I23.5, I23.6, I23.8, I24.0, I24.8, I24.9, I25.5, I25.6, I25.82, I25.89, I25.9

ICD-9-CM: 414.2, 411.0, 411.1, 411.81, 411.89, 414.8, 414.9

SNOMED CT: 10365005, 28248000, 28931004, 123641001, 123642008, 78741000119103, 457592003, 461417000, 697976003, 703328007, 703330009, 712866001, 713405002, 194821006, 194862000, 194863005, 194865003, 194867006, 194868001, 315026000, 233823002, 233846000, 233847009, 233860003, 233889001, 46109009, 56276002, 63739005, 398274000, 408546009, 413439005, 413444003, 413838009, 413844008, 414545008, 414795007, 421327009, 428196007, 1208872002, 1208873007, 1163440003, 1755008, 282006, 4557003, 10273003, 15990001, 19057007, 21470009, 22298006, 25106000, 30277009, 82522008, 85284003, 87343002, 89323001, 129574000, 836293000, 836294006, 836295007, 840309000, 840310005, 840312002, 840313007, 840315000, 840316004, 840608004, 840609007, 840679006, 840680009, 846667001, 846668006, 846683001, 846684007, 868214006, 868215007, 868216008, 868217004, 868219001, 868220007, 868221006,

868222004, 868223009, 868224003, 868225002, 868226001, 10971000087107, 17531000119105, 23311000119105, 117051000119103, 285981000119103, 380001000004106, 455941000124104, 12238111000119106, 12238151000119107, 15712841000119100, 15712881000119105, 15712921000119103, 15712961000119108, 15713041000119103, 15713161000119100, 15713201000119105, 15960061000119102, 15960141000119102, 15960381000119109, 15960541000119107, 15960581000119102, 15960661000119107, 15960781000119108, 15962541000119106, 15963181000119104, 16754391000119100, 16837681000119104, 19830001000004106, 32574007, 791000119109, 15713081000119108, 15713121000119105, 472100003, 473393007, 703164000, 703165004, 703209002, 703210007, 703211006, 703212004, 703213009, 703214003, 703251009, 703252002, 703253007, 703356002, 703360004, 735566005, 194802003, 194809007, 194823009, 194828000, 194856005, 194857001, 194858006, 194866002, 304914007, 307140009, 311792005, 311793000, 311796008, 314116003, 314207007, 315025001, 371806006, 371807002, 371808007, 371809004, 371810009, 371811008, 371812001, 394659003, 394710008, 233819005, 233821000, 233825009, 233826005, 233827001, 233828006, 233829003, 233830008, 233831007, 233832000, 233833005, 233834004, 233835003, 233836002, 233837006, 233838001, 233839009, 233840006, 233841005, 233842003, 233843008, 35928006, 41334000, 42531007, 52035003, 54329005, 57054005, 58612006, 59021001, 59063002, 61490001, 62695002, 64627002, 65547006, 70211005, 70422006, 70998009, 73795002, 76593002, 79009004, 401303003, 401314000, 418044006, 426856002, 429559004, 276516009, 276517000, 300995000, 15960341000119104, 15960461000119105, 726499301000119105, 879955009, 896689003, 896690007, 896691006, 896696001, 896697005, 1204151009, 1204154001, 1204155000, 1204152002, 1204222000

#### Myocardial infarction

ICD-10-CM: I21.01, I21.02, I21.09, I21.11, I21.19, I21.21, I21.29, I21.3, I21.4, I21.9, I21.A1, I21.A9, I22.0, I22.1, I22.2, I22.8, I22.9

ICD-9-CM: 410.00, 410.01, 410.02, 410.10, 410.11, 410.12, 410.20, 410.21, 410.22, 410.30, 410.31, 410.32, 410.40, 410.41, 410.42, 410.50, 410.51, 410.52, 410.60, 410.61, 410.62, 410.70, 410.71, 410.72, 410.80, 410.81, 410.82, 410.90, 410.91, 410.92

SNOMED CT: 10273003, 15990001, 22298006, 30277009, 129574000, 32574007, 194802003, 194809007, 194856005, 304914007, 307140009, 314207007, 394710008, 233835003, 233838001, 233843008, 42531007, 52035003, 54329005, 57054005, 58612006, 62695002, 65547006, 70211005, 70422006, 73795002, 79009004, 401303003, 401314000, 418044006, 428196007, 428752002, 1208872002, 1208873007, 1163440003, 1755008, 282006, 836293000, 836294006, 836295007, 840309000, 840312002, 840316004, 840609007, 840680009, 846668006, 846683001, 868214006, 868217004, 868220007, 868224003, 868225002, 868226001, 17531000119105, 23311000119105, 285981000119103, 380001000004106, 12238111000119106, 12238151000119107, 15712841000119100, 15712881000119105, 15712921000119103, 15712961000119108, 15713041000119103, 15713161000119100, 15713201000119105, 15962541000119106, 15963181000119104, 16837681000119104, 15713081000119108, 15713121000119105, 703164000, 703165004, 703209002, 703210007, 703211006, 703212004, 703213009, 703251009, 703252002, 703253007, 703360004, 194857001, 194858006, 311792005, 311793000, 311796008, 233825009, 233826005, 233827001, 233828006, 233829003, 233830008, 233831007, 233832000, 233833005, 233834004, 233836002, 233837006, 233839009, 233840006, 233841005, 233842003, 59063002, 64627002, 70998009, 76593002, 726499301000119105, 879955009, 896689003, 896691006, 896696001, 896697005, 1204151009, 1204154001, 1204155000, 1204152002, 1204222000

#### Pulmonary embolism

ICD-10-CM: I26.01, I26.02, I26.09, I26.92, I26.93, I26.94, I26.99

SNOMED CT: 328511000119109, 15964701000119109, 49584005, 59282003, 1163244009, 2261009, 11454006, 82153002, 860680001, 860681002, 1001000119102, 133971000119108, 10759311000119104, 609480009, 703636009, 706870000, 707412000, 707413005, 707414004, 713078005, 723859005, 194883006, 200284000, 200286003, 200288002, 200290001, 200291002, 200294005, 200299000, 200301007, 200303005, 200304004, 200308001, 200310004, 200311000, 233935004, 233936003, 233937007, 233938002, 233939005, 233940007, 237342006, 51096002, 55589000, 58123006, 74315008, 438773007, 441557008, 267284008, 280964006, 15964661000119102

Truveta: 1215970

#### Other Preexisting Cardiac Conditions

ICD-9-CM: 394.0, 394.1, 394.2, 394.9, 402.00, 402.10, 402.90, 404.00, 404.01, 404.02, 404.03, 404.10, 404.11, 404.12, 404.13, 404.90, 404.91, 404.92, 404.93, 405.01, 405.09, 405.11, 405.19, 405.91, 405.99, 414.2, 411.1, 412, 413.1, 413.9, 414.01, 414.02, 414.03, 414.04, 414.05, 414.06, 414.07, 414.10, 414.11, 414.12, 414.19, 414.3, 414.4, 414.8, 414.9, 420.0, 420.90, 420.91, 420.99, 421.0, 421.1, 421.9, 422.0, 422.90, 422.91, 422.92, 422.93, 422.99, 423.0, 423.1, 423.2, 423.3, 423.8, 423.9, 424.0, 424.1, 424.2, 424.3, 424.90, 424.91, 424.99, 426.0, 426.10, 426.11, 426.12, 426.13, 426.2, 426.3, 426.4, 426.50, 426.51, 426.52, 426.53, 426.7, 426.81, 426.82, 426.89, 426.9, 427.0, 427.1, 427.2, 427.31, 427.32, 427.5, 429.2, 674.01, 674.02

ICD-10-CM: I05, I06, I07, I08, I09, I11, I12, I13, I15, I16, I20, I25, I27.8, I30, I31, I32, I33, I34, I35, I36, I37, I38, I39, I40, I41, I44, I45, I46, I47, I48, I49, O99.41, O99.42, Q20, Q21, Q22, Q23, Q24, I05.0, I05.1, I05.2, I05.8, I05.9, I06.0, I06.1, I06.2, I06.8, I06.9, I07.0, I07.1, I07.2, I07.8, I07.9, I08.0, I08.1, I08.2, I08.3, I08.8, I08.9, I09.0, I09.1, I09.2, I09.8, I09.81, I09.89, I09.9, I11.0, I11.9, I12.0, I12.9, I13.0, I13.1, I13.10, I13.11, I13.2, I15.0, I15.1, I15.2, I15.8, I15.9, I16.0, I16.1, I16.9, I20.0, I20.1, I20.8, I20.9, I25.1, I25.10, I25.11, I25.110, I25.111, I25.118, I25.119, I25.2, I25.3, I25.4, I25.41, I25.42, I25.5, I25.6, I25.7, I25.70, I25.700, I25.701, I25.708, I25.709, I25.71, I25.710, I25.711, I25.718, I25.719, I25.72, I25.720, I25.721, I25.728, I25.729, I25.73, I25.730, I25.731, I25.738, I25.739, I25.75, I25.750, I25.751, I25.758, I25.759, I25.76, I25.760, I25.761, I25.768, I25.769, I25.79, I25.790, I25.791, I25.798, I25.799, I25.8, I25.81, I25.810, I25.811, I25.812, I25.82, I25.83, I25.84, I25.89, I25.9, I27.81, I27.82, I27.83, I27.89, I30.0, I30.1, I30.8, I30.9, I31.0, I31.1, I31.2, I31.3, I31.4, I31.8, I31.9, I33.0, I33.9, I34.0, I34.1, I34.2, I34.8, I34.9, I35.0, I35.1, I35.2, I35.8, I35.9, I36.0, I36.1, I36.2, I36.8, I36.9, I37.0, I37.1, I37.2, I37.8, I37.9, I40.0, I40.1, I40.8, I40.9, I44.0, I44.1, I44.2, I44.3, I44.30, I44.39, I44.4, I44.5, I44.6, I44.60, I44.69, I44.7, I45.0, I45.1, I45.10, I45.19, I45.2, I45.3, I45.4, I45.5, I45.6, I45.8, I45.81, I45.89, I45.9, I46.2, I46.8, I46.9, I47.0, I47.1, I47.2, I47.9, I48.0, I48.1, I48.11, I48.19, I48.2, I48.20, I48.21, I48.3, I48.4, I48.9, I48.91, I48.92, I49.0, I49.01, I49.02, I49.1, I49.2, I49.3, I49.4, I49.40, I49.49, I49.5, I49.8, I49.9, O99.411, O99.412, O99.413, O99.419, Q20.0, Q20.1, Q20.2, Q20.3, Q20.4, Q20.5, Q20.6, Q20.8, Q20.9, Q21.0, Q21.1, Q21.2, Q21.3, Q21.4, Q21.8, Q21.9, Q22.0, Q22.1, Q22.2, Q22.3, Q22.4, Q22.5, Q22.6, Q22.8, Q22.9, Q23.0, Q23.1, Q23.2, Q23.3, Q23.4, Q23.8, Q23.9, Q24.0, Q24.1, Q24.2, Q24.3, Q24.4, Q24.5, Q24.6, Q24.8, Q24.9

SNOMED CT: 3238004, 12026006, 13213009, 15555002, 23685000, 31992008, 83898004, 89736004,

8501000119104, 104931000119100, 698247007, 706882009, 708121009, 724497009, 194828000, 194921005, 194989009, 195002007, 195012000, 195080001, 233916004, 44808001, 46701001, 49699002, 50920009, 55855009, 59494005, 64715009, 70320004, 410429000, 413838009, 274097009, 1179295004, 1163420005, 1204124006, 1204167003, 1204129001, 1204130006, 1204122005, 1197418004, 1230303001, 1229876001, 1208884000, 1230344000, 1222710008, 1230406008, 1230405007, 1208845005, 1208839002, 1222708006, 1208998007, 1230097004, 1162915007, 1208881008, 1208868001, 1208870005, 1208866002, 1208861007, 1208911002, 1208864004, 1208863005, 1208909006, 1186711002, 1172567004, 1172698005, 1187212004, 1163259003, 1187113001, 1186709006, 1204460007, 1208897003, 1230096008, 488007, 682004, 756003, 787001, 1131009, 1474004, 2261009, 2374000, 2554006, 2583009, 3589003, 184004, 279001, 4006006, 4082005, 4313003, 4374004, 4554005, 4557003, 4973001, 5148006, 5230009, 5370000, 5609005, 5836005, 6180003, 6285003, 6374002, 6456007, 6594005, 6624005, 6636004, 6996004, 7368005, 7484005, 7607008, 8218002, 8239009, 8676001, 8712002, 8762007, 9901000, 10058006, 10164001, 10562009, 10818008, 11157007, 11176009, 11433004, 11454006, 11849007, 12023003, 12095001, 12770006, 13395001, 13453009, 13640000, 13689005, 14455008, 14718009, 14886009, 14973001, 15016005, 15096009, 15394000, 15459006, 15676002, 16063004, 16253001, 16440002, 16567006, 16574001, 16797001, 16855001, 16876006, 17079009, 17263003, 17338001, 17366009, 17668000, 17718000, 17759006, 17869006, 18192007, 18416000, 18484008, 18546004, 18687009, 19036004, 19057007, 19249002, 20005002, 20143001, 20753005, 20852007, 20956009, 21290002, 21470009, 21604008, 21981000, 22167000, 22653005, 22966008, 23063005, 23412002, 23627006, 23717007, 23786008, 23842007, 24042004, 25380002, 25503004, 26078007, 26117009, 26201005, 26743002, 26850007, 26950008, 27015006, 27017003, 27637000, 27806003, 27885002, 28119000, 28284007, 28381002, 28386007, 28574005, 28608002, 28656008, 28773001, 28880005, 28966002, 29259002, 29320008, 29375001, 29717002, 29894000, 29928006, 29934004, 29950007, 29995000, 30328003, 31085000, 31407004, 31481000, 31881008, 31993003, 32425009, 32758004, 33413000, 33558009, 34126003, 34273003, 81376009, 81552002, 81580000, 81626002, 81681009, 81898007, 81990004, 82153002, 82226007, 82458004, 82523003, 82542004, 82838007, 83105008, 83119008, 83799000, 83916000, 84216001, 84648007, 84871007, 84939004, 85081000, 85284003, 85598007, 85971001, 86014007, 86041002, 86152005, 86234004, 86466006, 86504008, 87343002, 87648004, 88318005, 88412007, 88445007, 88782004, 88887003, 89141000, 89222008, 89242004, 89323001, 89456004, 90428001, 90453003, 91025000, 91442002, 91468009, 91529004, 91634006, 91841003, 92087008, 92289001, 92830003, 92888002, 92923008, 92941000, 92944008, 92960007, 92963009, 92969008, 92974000, 92976003, 93018000, 93051009, 93053007, 93069000, 93075009, 93076005, 93078006, 93234006, 93237004, 93247001, 93262004, 93271008, 93287006, 93782004, 93957005, 94150003, 94282006, 94484009, 94702005, 94703000, 94706008, 94822000, 94997003, 109382005, 109383000, 109425008, 109426009, 109427000, 109428005, 109429002, 109430007, 109431006, 109432004, 111284004, 111294009, 111300007, 111301006, 111319002, 111432008, 111438007, 111458008, 123599008, 123656005, 123657001, 123658006, 123659003, 123660008, 123799005, 123800009, 126733004, 126734005, 126735006, 128555001, 128556000, 128557009, 128558004, 128563000, 128567004, 128568009, 129575004, 129582000, 1141647009, 1141882007, 1142040009, 1142041008, 1142057008, 1142064005, 1142118003, 1142066007, 1142067003, 1142068008, 1142069000, 1142082008, 1142086006, 1142090008, 1142093005, 1142098001, 1142103004, 1142105006, 1142110005, 1142111009, 1142114001, 1142117008, 1142119006, 1142120000, 1142121001, 1142122008, 1142123003, 1142124009, 1142204001, 1144435009, 1144765000, 1144766004, 1144797000, 1144807000, 1148583001, 1148584007, 1148587000, 1148765006, 1149066008, 1149361009, 1149362002, 1149472000, 1149473005, 1155719000, 1155721005, 1156027008, 1156029006, 1156031002, 1156032009, 1156821008, 16415081000119104, 788290007, 788529004, 788533006, 789039008, 789693005, 827078006, 827163002, 827164008, 838316003, 838333005, 840469002, 840488009, 840495000, 840497008,

840507004, 840517009, 860676006, 860677002, 860678007, 860679004, 860680001, 860681002, 860682009, 860683004, 860686007, 860687003, 860688008, 860689000, 866118008, 870301007, 871579003, 871581001, 871583003, 871585005, 871587002, 871594004, 871597006, 871599009, 871600007, 871601006, 871602004, 871603009, 871604003, 871605002, 871606001, 871607005, 871609008, 871610003, 871611004, 871612006, 871613001, 871616009, 871618005, 871620008, 871622000, 871624004, 871625003, 871627006, 871630004, 871636005, 871640001, 871642009, 871643004, 871644005, 871645006, 871647003, 871648008, 871650000, 871652008, 871654009, 871655005, 871658007, 871659004, 871660009, 871666003, 871668002, 871672003, 871686004, 871832005, 541000119105, 5561000124105, 5761000119100, 10701000087104, 15781000119107, 21421000119109, 40511000119107, 40521000119100, 41371000119100, 63191000119107, 72181000119109, 82771000119102, 94851000119107, 96701000119107, 96711000119105, 96721000119103, 96731000119100, 96741000119109, 96751000119106, 100231000119101, 102451000119107, 108701000119102, 111411000119103, 117681000119102, 118781000119108, 120041000119109, 120261000119101, 127991000119101, 128001000119105, 129151000119102, 129161000119100, 129171000119106, 129181000119109, 132721000119104, 153851000119106, 153891000119101, 284941000119107, 284951000119109, 284961000119106, 284971000119100, 284981000119102, 284991000119104, 285001000119105, 285011000119108, 285041000119107, 285061000119106, 285081000119102, 285101000119109, 285821000119105, 285831000119108, 285841000119104, 285851000119102, 285861000119100, 285871000119106, 285881000119109, 285911000119109, 285921000119102, 286001000119104, 286011000119101, 286031000119106, 286041000119102, 286371000119107, 367821000119106, 434431000124103, 449631000124102, 451241000124108, 1089931000119105, 1240531000000103, 10629351000119108, 10743121000119108, 10743741000119103, 10745231000119102, 10749691000119103, 10750641000119101, 10752641000119102, 10753101000119102, 10760081000119108, 10754331000119108, 10757401000119104, 10757441000119102, 10757481000119107, 10759031000119106, 10759191000119106, 10759311000119104, 10759761000119100, 10760131000119108, 10760181000119109, 10805641000119102, 10809641000119100, 10811961000119109, 10835781000119105, 15960061000119102, 15960141000119102, 15960381000119109, 15960541000119107, 15960581000119102, 15960661000119107, 15960781000119108, 15964581000119107, 15964781000119101, 15964901000119107, 15964981000119104, 16754391000119100, 2829000, 9651007, 10626002, 17394001, 19092004, 22271007, 26146002, 27337007, 28189009, 30667004, 31080005, 37980001, 38274001, 39260000, 42402006, 46319007, 48194001, 54016002, 55475008, 63247009, 74390002, 80387009, 83014000, 86299006, 87442008, 95605009, 179924009, 204311009, 204319006, 204351007, 204357006, 253468007, 253496001, 253514004, 253515003, 283645003, 420938005, 421272004, 421929001, 422348008, 442917000, 460317007, 460329003, 460338001, 460352004, 460439008, 460440005, 460464006, 460620006, 698272007, 791000119109, 142951000119106, 444605001, 444851003, 444961009, 445001003, 445002005, 445003000, 445131007, 445235006, 445264002, 445268004, 445270008, 445298006, 445299003, 445310002, 445330003, 445334007, 445373007, 445435009, 445453003, 445454009, 445607003, 445636003, 445650008, 445898001, 445928005, 446326008, 446657003, 446659000, 446667008, 446670007, 446909006, 446916007, 447283008, 447284002, 447285001, 447286000, 447289007, 447661004, 447665008, 447666009, 447667000, 447669002, 447670001, 447671002, 447672009, 447673004, 447681003, 447683000, 447689001, 447690005, 447696004, 447697008, 447698003, 447774002, 447780005, 447822009, 447824005, 447825006, 447829000, 447830005, 447838003, 447839006, 447845003, 447852001, 447874007, 447875008, 447876009, 447913009, 447915002, 447919008, 447932003, 447933008, 447937009, 447938004, 447941008, 447962009, 447988007, 447998001, 447999009, 448001004, 448004007, 448007000, 448028009, 448062009, 448063004, 448064005, 448066007, 448072007, 448073002, 448074008, 448075009, 448080000, 448081001, 448082008, 448083003, 448085005, 448095003, 448103004, 448113007, 448118003, 448119006, 448120000, 448155003, 448161000, 448162007, 448181004, 448182006, 448183001, 448184007, 448277007, 448278002, 448280008, 448305002, 448308000, 448309008, 448310003, 448320008, 448331000, 448356006, 448357002, 448413002, 448415009, 448475002, 448477005, 448478000, 448497000, 448516008, 448574001, 448575000, 448604006, 448612003,

448619007, 448624005, 448625006, 448633007, 448634001, 448681000, 448729006, 448747000, 448778001, 448780007, 448786001, 448790004, 448793002, 448794008, 448809003, 448819009, 448823001, 448827000, 448842002, 448843007, 448844001, 448876006, 448887003, 448919005, 448920004, 448923002, 448946000, 448947009, 448948004, 448966009, 448975006, 449005003, 449014008, 449015009, 449016005, 449029005, 449031001, 449032008, 449037002, 449040002, 449086000, 449087009, 449098005, 449099002, 449110001, 449118008, 449123008, 449124002, 449133000, 449135007, 449138009, 449139001, 449140004, 449158005, 449178002, 449190000, 449228007, 449270002, 449315003, 449316002, 449352003, 449353008, 449354002, 449379004, 449382009, 449383004, 449385006, 449398002, 449428009, 449429001, 449436000, 449440009, 449441008, 449442001, 449455000, 449456004, 449457008, 449458003, 449459006, 449479003, 449490008, 449492000, 449511008, 449512001, 449520004, 449523002, 449532000, 449534004, 449537006, 449538001, 449539009, 449563001, 449567000, 449568005, 449587004, 449593007, 449594001, 449595000, 449596004, 449599006, 449600009, 450300002, 450301003, 450302005, 450303000, 450919004, 458039003, 459062008, 459065005, 459066006, 459067002, 459068007, 459151006, 459152004, 459153009, 459154003, 459158000, 459159008, 459178008, 459184006, 460345001, 460437005, 460438000, 460445000, 460450006, 460457009, 460471001, 460510005, 460517008, 460524009, 460531008, 460538002, 460545002, 460581004, 460582006, 460583001, 460584007, 460585008, 460586009, 460587000, 460588005, 461345005, 460614005, 460913001, 460923005, 460930004, 460937001, 460944005, 461104009, 461105005, 461107002, 461109004, 461110009, 461111008, 461112001, 461331004, 461359003, 461381008, 461382001, 461383006, 461384000, 461385004, 461420008, 461421007, 461435009, 461436005, 461438006, 461439003, 462164009, 462169004, 462170003, 471276003, 471277007, 471285003, 471286002, 471287006, 471288001, 471289009, 471290000, 471291001, 471292008, 471293003, 471294009, 471297002, 471298007, 471299004, 471841009, 472703008, 472707009, 472777008, 472778003, 472785004, 472786003, 472787007, 472794005, 472795006, 472796007, 472797003, 472799000, 472800001, 472801002, 472802009, 472809000, 472810005, 472820000, 473006003, 473362006, 473367000, 473368005, 473393007, 473443007, 473444001, 609302002, 609449004, 609457001, 609458006, 609459003, 609460008, 609465003, 609480009, 609486003, 609489005, 609497003, 609506003, 609507007, 698249005, 698250005, 698251009, 698252002, 698270004, 698271000, 698504006, 698638005, 698640000, 698971006, 699256006, 699748007, 703162001, 703178004, 703179007, 703189006, 703190002, 703191003, 703236000, 703320000, 703332001, 704667004, 706923002, 710878005, 712487000, 712859002, 713318009, 713419002, 715395008, 715535009, 715560009, 715865008, 715971003, 715987000, 716050002, 716193004, 716740009, 716773002, 717812000, 717859007, 717943008, 717959008, 718128009, 718135001, 718181001, 718216009, 718556007, 718681002, 718713000, 719379001, 719395001, 719400000, 719451006, 719456001, 719823007, 719907006, 720448006, 720507006, 720567008, 720605009, 720606005, 720610008, 720612000, 720639008, 720748007, 720858001, 721009008, 721010003, 721013001, 721014007, 721015008, 721073008, 721976003, 721978002, 722027009, 722051004, 722206009, 722461004, 723088005, 723304001, 723333000, 723336008, 723448007, 723860000, 723863003, 723865005, 723866006, 724066002, 724068001, 724486009, 725145002, 726083008, 726335002, 726704006, 733125004, 733454004, 733491005, 735682000, 735683005, 735684004, 735685003, 735768008, 737324009, 737325005, 737331008, 737332001, 737333006, 762247006, 762251008, 762252001, 762253006, 762254000, 762433009, 762460002, 762534000, 762539005, 762540007, 763066009, 763279007, 763747002, 763778003, 763834000, 763835004, 764457005, 764697003, 764732004, 764955006, 765182005, 766704005, 766881008, 766883006, 766937004, 766976003, 768552007, 770432008, 770433003, 770435005, 770784003, 771179007, 773139006, 773587008, 773749003, 780842009, 781159007, 782698000, 782699008, 783011004, 783096008, 783738002, 783773000, 784353002, 162710004, 169465000, 173574009, 174511002, 176401009, 177291008, 180906006, 181869007, 186875004, 186877007, 186878002, 187040006, 187050007, 187059008, 187195003, 194709000, 194727002, 194732001, 194733006, 194734000, 194735004, 194736003, 194741006, 194750008, 194767001, 194779001, 194780003, 194781004, 194783001, 194785008, 194788005, 194791005, 194823009, 194862000, 194905000, 194906004, 194907008, 194908003,

194909006, 194910001, 194916007, 194917003, 194918008, 194926000, 194942007, 194943002, 194944008, 194945009, 194946005, 194947001, 194948006, 194949003, 194950003, 194956009, 194957000, 194958005, 194964003, 194965002, 194970009, 194978002, 194983005, 194984004, 194990000, 194991001, 194992008, 194995005, 194997002, 195003002, 195004008, 195005009, 195033009, 195039008, 195042002, 195046004, 195060002, 195069001, 195070000, 195071001, 195072008, 195083004, 195105007, 195136004, 198650006, 198663000, 198711003, 198750004, 198941007, 198942000, 198944004, 198945003, 198946002, 198947006, 198949009, 198951008, 198952001, 198953006, 198965005, 198966006, 198967002, 198968007, 198983002, 198984008, 198985009, 198986005, 198992004, 198997005, 198999008, 199000005, 199002002, 199003007, 199005000, 199006004, 199007008, 199008003, 199192005, 199193000, 199194006, 199195007, 199266007, 199267003, 199268008, 199269000, 200054002, 200056000, 200057009, 200058004, 200059007, 200077002, 200111005, 200112003, 200113008, 200114002, 200204002, 200206000, 200207009, 200208004, 200209007, 200215007, 200216008, 200217004, 200218009, 200222004, 200224003, 200226001, 200227005, 200232006, 200233001, 200237000, 200238005, 200249004, 200251000, 200252007, 200253002, 200254008, 200255009, 200259003, 200260008, 200284000, 200286003, 200288002, 200290001, 200291002, 200294005, 200296007, 200297003, 200299000, 200301007, 200303005, 200304004, 200308001, 200310004, 200311000, 200330000, 200331001, 200332008, 200333003, 204296002, 204300001, 204306007, 204312002, 204317008, 204318003, 204330009, 204339005, 204342004, 204345002, 204346001, 204352000, 204354004, 204362007, 204363002, 204370002, 204378009, 204379001, 204383001, 204384007, 204394002, 204395001, 204397009, 204398004, 204399007, 204448004, 204470001, 204589003, 205768003, 205834002, 307632004, 308133005, 308134004, 308135003, 308136002, 308137006, 308140006, 308187004, 309809007, 311830005, 311831009, 312591002, 314116003, 314208002, 315025001, 315027009, 315615007, 360473004, 360481003, 367390009, 370141003, 371806006, 371807002, 371808007, 371809004, 371810009, 371811008, 371812001, 373131000, 373905003, 373945007, 391179008, 391180006, 395520006, 210078001, 213213007, 233819005, 233821000, 233853009, 233862006, 233867000, 233868005, 233869002, 233881003, 233883000, 233884006, 233885007, 233886008, 233887004, 233888009, 233889001, 233890005, 233891009, 233892002, 233893007, 233894001, 233895000, 233896004, 233897008, 233898003, 233899006, 233900001, 233901002, 233902009, 233903004, 233904005, 233910005, 233911009, 233913007, 233914001, 233915000, 233917008, 233918003, 233919006, 233922008, 233923003, 233927002, 234058009, 234132006, 234172002, 236529001, 236530006, 237227006, 237279007, 237281009, 237282002, 237286004, 237342006, 237343001, 237345008, 237346009, 237347000, 240298005, 240299002, 250941001, 250942008, 250983006, 250999000, 251092003, 251093008, 251094002, 251114004, 251120003, 251123001, 251124007, 251125008, 251152003, 251155001, 251161003, 251162005, 251163000, 251164006, 251165007, 251166008, 251167004, 251168009, 251170000, 251171001, 251172008, 251173003, 251174009, 251175005, 251176006, 251177002, 251178007, 251179004, 251180001, 251181002, 251182009, 251183004, 251186007, 251187003, 251188008, 253264007, 253267000, 253268005, 253269002, 253270001, 253271002, 253272009, 253274005, 253275006, 253276007, 253277003, 253278008, 253279000, 253280002, 253281003, 253282005, 253283000, 253284006, 253285007, 253286008, 253287004, 253288009, 253289001, 253290005, 253291009, 253293007, 253294001, 253295000, 253297008, 253298003, 253299006, 253300003, 253301004, 253302006, 253303001, 253304007, 253305008, 253324006, 253326008, 253327004, 253333008, 253335001, 253336000, 253337009, 253338004, 253341008, 253342001, 253343006, 253345004, 253346003, 253348002, 253349005, 253353007, 253354001, 253356004, 253357008, 253359006, 253360001, 253364005, 253371000, 253373002, 253374008, 253376005, 253377001, 253378006, 253379003, 253381001, 253382008, 253384009, 253391007, 253392000, 253393005, 253394004, 253395003, 253396002, 253397006, 253399009, 253403000, 253404006, 253405007, 253411005, 253412003, 253413008, 253414002, 253415001, 253416000, 253417009, 253418004, 253419007, 253420001, 253421002, 253422009, 253423004, 253425006, 253426007, 253439006, 253441007, 253450009, 253451008, 253458002, 253465005, 253478005, 253479002, 253480004, 253481000, 253482007, 253483002, 253484008, 253485009, 253486005, 253487001, 253489003, 253492004,

253493009, 253494003, 253495002, 253497005, 253498000, 253499008, 253500004, 253501000, 253502007, 253503002, 253504008, 253505009, 253506005, 253507001, 253508006, 253509003, 253510008, 253511007, 253512000, 253513005, 253518001, 253519009, 253520003, 253521004, 253523001, 253524007, 253525008, 253527000, 253528005, 253529002, 253530007, 253531006, 253532004, 253533009, 253534003, 253536001, 253537005, 253538000, 253539008, 253540005, 253542002, 253543007, 253544001, 253545000, 253546004, 253547008, 253548003, 253550006, 253551005, 253552003, 253553008, 253554002, 253555001, 253556000, 253559007, 253562005, 253563000, 253564006, 253565007, 253566008, 253567004, 253568009, 253569001, 253570000, 253571001, 253573003, 253574009, 253575005, 253576006, 253578007, 253580001, 253584005, 253585006, 253586007, 253587003, 253588008, 253591008, 253593006, 253596003, 253597007, 253598002, 253599005, 253600008, 253601007, 253602000, 253603005, 253607006, 253608001, 253609009, 253610004, 253611000, 253624000, 253625004, 253650001, 253700001, 253703004, 253704005, 253706007, 253707003, 253708008, 253710005, 253711009, 253712002, 253714001, 253715000, 253716004, 253717008, 253718003, 253719006, 253720000, 253725005, 253728007, 253729004, 253730009, 253731008, 253732001, 253733006, 253734000, 34694006, 35303009, 35577008, 35928006, 36078000, 36079008, 36083008, 36133000, 36221001, 36233006, 36315003, 37104009, 37139009, 37217002, 37278001, 37618003, 37639005, 37715009, 37760005, 37787009, 37925008, 38566003, 38905006, 39018007, 39097001, 39244004, 39357005, 39589002, 39727004, 40023000, 40193008, 40272001, 40312006, 40593004, 40959008, 41049003, 41114007, 41334000, 41514002, 41739008, 41863008, 41893002, 41991004, 42653000, 42999000, 43121002, 43176009, 43306002, 43736008, 43742007, 43906007, 44101005, 44103008, 44595001, 44602002, 44622003, 44918005, 44993000, 45093008, 45304001, 45503006, 46113002, 46165000, 46220003, 46619002, 46676000, 46764007, 46935006, 47292005, 47780009, 47830009, 48121000, 48228003, 48520006, 48552006, 48872007, 49044005, 49102001, 49260003, 49436004, 49710005, 49956009, 49982000, 50799005, 50902000, 51096002, 51442005, 51789008, 52048004, 52535005, 53874003, 54073003, 54225002, 54529009, 55043004, 55087008, 55482007, 55510008, 55546004, 55565007, 55589000, 55879006, 55976003, 56272000, 56309007, 56501009, 57231008, 57684003, 57734001, 57803009, 58474006, 58600007, 59021001, 59118001, 59204004, 59272004, 59464004, 59554006, 59733002, 60106004, 60232001, 60254004, 60265009, 60357009, 60423000, 60601008, 60732002, 60812006, 60899001, 61012002, 61277005, 61490001, 61558009, 61575003, 61959006, 62023000, 62026008, 62067003, 62377009, 62583006, 62657007, 63040001, 63042009, 63232000, 63287004, 63462008, 63467002, 63593006, 63795001, 64043005, 64072006, 64171002, 64190005, 64420008, 64862009, 65402008, 65447009, 65718001, 65778007, 66052004, 66131005, 66568003, 66610008, 66657009, 66704002, 67198005, 67256000, 67278007, 67359005, 67486009, 67696008, 69589006, 69730002, 69909000, 70189005, 70272006, 71023004, 71362000, 71792006, 71799002, 71874008, 71908006, 72011007, 72022006, 72252007, 72352009, 72527006, 72654001, 73410007, 73459006, 73660006, 73699003, 74021003, 74034002, 74218008, 74381009, 74451002, 74615001, 74918002, 75270000, 75372006, 75398000, 75532003, 75825001, 76257003, 76534005, 76541004, 76887001, 77221000, 77278008, 77696009, 77970009, 78031003, 78046005, 78069008, 78196008, 78240009, 78250005, 78320000, 78381004, 78485007, 78495000, 78643003, 78808002, 78975002, 79096004, 79355007, 80119007, 397829000, 397830005, 397894003, 398078006, 398147005, 398254007, 399046008, 399216004, 399228007, 402836009, 405546008, 405752007, 406461004, 410430005, 412787009, 413341007, 413342000, 413844008, 413905004, 413933004, 415810004, 417426005, 418341009, 418493005, 418818005, 419400008, 419671004, 419752005, 420002000, 420395004, 421869004, 422970001, 423168004, 423191000, 424045003, 424571008, 425582007, 425615007, 426749004, 427372006, 427443009, 427665004, 427889009, 428163005, 428575007, 429243003, 429347004, 429559004, 440028005, 440059007, 442559009, 442946007, 443379009, 443478002, 443482000, 254826008, 261195002, 263944006, 263961009, 264086008, 264162009, 264467005, 264571006, 266235007, 266237004, 266238009, 267207004, 267280004,

267282007, 267283002, 267284008, 268174004, 268180007, 270492004, 270510008, 274119009, 274947007, 275434003, 275513007, 275516004, 276512006, 276513001, 276796006, 278480000, 278482008, 278536002, 282825002, 284470004, 285830009, 286947004, 286950001, 287057009, 288250001, 300995000, 300996004, 300997008, 302943003, 302945005, 1010405004, 1010443001, 1010444007, 1010446009, 1010447000, 1052287002, 15960341000119104, 15960461000119105, 876846006, 880012008, 890220003, 890393009, 890402009, 1222539006, 1220643007

#### Essential hypertension

ICD-10-CM: I10

ICD-9-CM: 401.0, 401.1, 401.9

SNOMED CT: 1201005, 10725009, 461301000124109, 1078301000112109, 762463000, 371125006, 46481004, 48146000, 56218007, 59621000, 59720008, 65518004, 78975002, 429457004, 9901000, 18416000, 19769006, 23717007, 40511000119107, 194785008, 35303009, 63287004, 71874008, 72022006, 73410007, 74451002, 78808002

#### Physical disability

ICD-9-CM: 340, 343, 714, 725, 720, 741, 342, 344, 806, 905.1, 907, 952, 953, 438, 045.0, 045.1, 045.9, 138, 358, 359, 756.4, V46.3, V46.8, 237.7, 331, 332, 333, 334, 335, 336, 337, 341, 345, 353, 356, 357, 253, 715, 717, 721, 722, 723, 728, 732, 928, 736.89, V49.6, V49.7, 277.00, 277.01, 277.02, 277.03, 277.09, 277.1, 277.2, 277.3, 277.4, 277.5, 277.6, 277.7, 277.8, 277.9, 259.4, 742, 755, 754.51, 754.52, 754.53, 754.59, 754.6, 754.7, 756

ICD-10-CM: G35, G80, G80.0, G80.1, G80.2, G80.3, G80.4, G80.8, G80.9, M05, M06, M08.0, M08.2, M08.3, M08.4, M05.0, M05.00, M05.01, M05.011, M05.012, M05.019, M05.02, M05.021, M05.022, M05.029, M05.03, M05.031, M05.032, M05.039, M05.04, M05.041, M05.042, M05.049, M05.05, M05.051, M05.052, M05.059, M05.06, M05.061, M05.062, M05.069, M05.07, M05.071, M05.072, M05.079, M05.09, M05.1, M05.10, M05.11, M05.111, M05.112, M05.119, M05.12, M05.121, M05.122, M05.129, M05.13, M05.131, M05.132, M05.139, M05.14, M05.141, M05.142, M05.149, M05.15, M05.151, M05.152, M05.159, M05.16, M05.161, M05.162, M05.169, M05.17, M05.171, M05.172, M05.179, M05.19, M05.2, M05.20, M05.21, M05.211, M05.212, M05.219, M05.22, M05.221, M05.222, M05.229, M05.23, M05.231, M05.232, M05.239, M05.24, M05.241, M05.242, M05.249, M05.25, M05.251, M05.252, M05.259, M05.26, M05.261, M05.262, M05.269, M05.27, M05.271, M05.272, M05.279, M05.29, M05.3, M05.30, M05.31, M05.311, M05.312, M05.319, M05.32, M05.321, M05.322, M05.329, M05.33, M05.331, M05.332, M05.339, M05.34, M05.341, M05.342, M05.349, M05.35, M05.351, M05.352, M05.359, M05.36, M05.361, M05.362, M05.369, M05.37, M05.371, M05.372, M05.379, M05.39, M05.4, M05.40, M05.41, M05.411, M05.412, M05.419, M05.42, M05.421, M05.422, M05.429, M05.43, M05.431, M05.432, M05.439, M05.44, M05.441, M05.442, M05.449, M05.45, M05.451, M05.452, M05.459, M05.46, M05.461, M05.462, M05.469, M05.47, M05.471, M05.472, M05.479, M05.49, M05.5, M05.50, M05.51, M05.511, M05.512, M05.519, M05.52, M05.521, M05.522, M05.529, M05.53, M05.531, M05.532, M05.539, M05.54, M05.541, M05.542, M05.549, M05.55, M05.551, M05.552, M05.559, M05.56, M05.561, M05.562, M05.569, M05.57, M05.571, M05.572, M05.579, M05.59, M05.6, M05.60, M05.61, M05.611, M05.612, M05.619, M05.62, M05.621, M05.622,

M05.629, M05.63, M05.631, M05.632, M05.639, M05.64, M05.641, M05.642, M05.649, M05.65, M05.651, M05.652, M05.659, M05.66, M05.661, M05.662, M05.669, M05.67, M05.671, M05.672, M05.679, M05.69, M05.7, M05.70, M05.71, M05.711, M05.712, M05.719, M05.72, M05.721, M05.722, M05.729, M05.73, M05.731, M05.732, M05.739, M05.74, M05.741, M05.742, M05.749, M05.75, M05.751, M05.752, M05.759, M05.76, M05.761, M05.762, M05.769, M05.77, M05.771, M05.772, M05.779, M05.852, M05.79, M05.8, M05.80, M05.81, M05.811, M05.812, M05.819, M05.82, M05.821, M05.822, M05.829, M05.83, M05.831, M05.832, M05.839, M05.84, M05.841, M05.842, M05.849, M05.85, M05.851, M05.859, M05.86, M05.861, M05.862, M05.869, M05.87, M05.871, M05.872, M05.879, M05.89, M05.9, M06.0, M06.00, M06.01, M06.011, M06.012, M06.019, M06.02, M06.021, M06.022, M06.029, M06.03, M06.031, M06.032, M06.039, M06.04, M06.041, M06.042, M06.049, M06.05, M06.051, M06.052, M06.059, M06.06, M06.061, M06.062, M06.069, M06.07, M06.071, M06.072, M06.079, M06.08, M06.09, M06.1, M06.2, M06.20, M06.21, M06.211, M06.212, M06.219, M06.22, M06.221, M06.222, M06.229, M06.23, M06.231, M06.232, M06.239, M06.24, M06.241, M06.242, M06.249, M06.25, M06.251, M06.252, M06.259, M06.26, M06.261, M06.262, M06.269, M06.27, M06.271, M06.272, M06.279, M06.28, M06.29, M06.3, M06.30, M06.31, M06.311, M06.312, M06.319, M06.32, M06.321, M06.322, M06.329, M06.33, M06.331, M06.332, M06.339, M06.34, M06.341, M06.342, M06.349, M06.35, M06.351, M06.352, M06.359, M06.36, M06.361, M06.362, M06.369, M06.37, M06.371, M06.372, M06.379, M06.38, M06.39, M06.4, M06.8, M06.80, M06.81, M06.811, M06.812, M06.819, M06.82, M06.821, M06.822, M06.829, M06.83, M06.831, M06.832, M06.839, M06.84, M06.841, M06.842, M06.849, M06.85, M06.851, M06.852, M06.859, M06.86, M06.861, M06.862, M06.869, M06.87, M06.871, M06.872, M06.879, M06.88, M06.89, M06.9, M08.00, M08.01, M08.011, M08.012, M08.019, M08.02, M08.021, M08.022, M08.029, M08.03, M08.031, M08.032, M08.039, M08.04, M08.041, M08.042, M08.049, M08.05, M08.051, M08.052, M08.059, M08.06, M08.061, M08.062, M08.069, M08.07, M08.071, M08.072, M08.079, M08.08, M08.09, M08.20, M08.21, M08.211, M08.212, M08.219, M08.22, M08.221, M08.222, M08.229, M08.23, M08.231, M08.232, M08.239, M08.24, M08.241, M08.242, M08.249, M08.25, M08.251, M08.252, M08.259, M08.26, M08.261, M08.262, M08.269, M08.27, M08.271, M08.272, M08.279, M08.28, M08.29, M08.40, M08.41, M08.411, M08.412, M08.419, M08.42, M08.421, M08.422, M08.429, M08.43, M08.431, M08.432, M08.439, M08.44, M08.441, M08.442, M08.449, M08.45, M08.451, M08.452, M08.459, M08.46, M08.461, M08.462, M08.469, M08.47, M08.471, M08.472, M08.479, M08.48, M05.7A, M05.8A, M06.0A, M06.8A, M08.0A, M08.2A, M08.4A, M05, M06, M05.0, M05.00, M05.01, M05.011, M05.012, M05.019, M05.02, M05.021, M05.022, M05.029, M05.03, M05.031, M05.032, M05.039, M05.04, M05.041, M05.042, M05.049, M05.05, M05.051, M05.052, M05.059, M05.06, M05.061, M05.062, M05.069, M05.07, M05.071, M05.072, M05.079, M05.09, M05.1, M05.10, M05.11, M05.111, M05.112, M05.119, M05.12, M05.121, M05.122, M05.129, M05.13, M05.131, M05.132, M05.139, M05.14, M05.141, M05.142, M05.149, M05.15, M05.151, M05.152, M05.159, M05.16, M05.161, M05.162, M05.169, M05.17, M05.171, M05.172, M05.179, M05.19, M05.2, M05.20, M05.21, M05.211, M05.212, M05.219, M05.22, M05.221, M05.222, M05.229, M05.23, M05.231, M05.232, M05.239, M05.24, M05.241, M05.242, M05.249, M05.25, M05.251, M05.252, M05.259, M05.26, M05.261, M05.262, M05.269, M05.27, M05.271, M05.272, M05.279, M05.29, M05.3, M05.30, M05.31, M05.311, M05.312, M05.319, M05.32, M05.321, M05.322, M05.329, M05.33, M05.331, M05.332, M05.339, M05.34, M05.341, M05.342, M05.349, M05.35, M05.351, M05.352, M05.359, M05.36, M05.361, M05.362, M05.369, M05.37, M05.371, M05.372, M05.379, M05.39, M05.4, M05.40, M05.41, M05.411, M05.412, M05.419, M05.42, M05.421, M05.422, M05.429, M05.43, M05.431, M05.432, M05.439, M05.44, M05.441, M05.442, M05.449, M05.45, M05.451, M05.452, M05.459, M05.46, M05.461, M05.462, M05.469, M05.47, M05.471, M05.472, M05.479, M05.49, M05.5, M05.50,

M05.51, M05.511, M05.512, M05.519, M05.52, M05.521, M05.522, M05.529, M05.53, M05.531, M05.532, M05.539, M05.54, M05.541, M05.542, M05.549, M05.55, M05.551, M05.552, M05.559, M05.56, M05.561, M05.562, M05.569, M05.57, M05.571, M05.572, M05.579, M05.59, M05.6, M05.60, M05.61, M05.611, M05.612, M05.619, M05.62, M05.621, M05.622, M05.629, M05.63, M05.631, M05.632, M05.639, M05.64, M05.641, M05.642, M05.649, M05.65, M05.651, M05.652, M05.659, M05.66, M05.661, M05.662, M05.669, M05.67, M05.671, M05.672, M05.679, M05.69, M05.7, M05.70, M05.71, M05.711, M05.712, M05.719, M05.72, M05.721, M05.722, M05.729, M05.73, M05.731, M05.732, M05.739, M05.74, M05.741, M05.742, M05.749, M05.75, M05.751, M05.752, M05.759, M05.76, M05.761, M05.762, M05.769, M05.77, M05.771, M05.772, M05.779, M05.852, M05.79, M05.8, M05.80, M05.81, M05.811, M05.812, M05.819, M05.82, M05.821, M05.822, M05.829, M05.83, M05.831, M05.832, M05.839, M05.84, M05.841, M05.842, M05.849, M05.85, M05.851, M05.859, M05.86, M05.861, M05.862, M05.869, M05.87, M05.871, M05.872, M05.879, M05.89, M05.9, M06.0, M06.00, M06.01, M06.011, M06.012, M06.019, M06.02, M06.021, M06.022, M06.029, M06.03, M06.031, M06.032, M06.039, M06.04, M06.041, M06.042, M06.049, M06.05, M06.051, M06.052, M06.059, M06.06, M06.061, M06.062, M06.069, M06.07, M06.071, M06.072, M06.079, M06.08, M06.09, M06.1, M06.2, M06.20, M06.21, M06.211, M06.212, M06.219, M06.22, M06.221, M06.222, M06.229, M06.23, M06.231, M06.232, M06.239, M06.24, M06.241, M06.242, M06.249, M06.25, M06.251, M06.252, M06.259, M06.26, M06.261, M06.262, M06.269, M06.27, M06.271, M06.272, M06.279, M06.28, M06.29, M06.3, M06.30, M06.31, M06.311, M06.312, M06.319, M06.32, M06.321, M06.322, M06.329, M06.33, M06.331, M06.332, M06.339, M06.34, M06.341, M06.342, M06.349, M06.35, M06.351, M06.352, M06.359, M06.36, M06.361, M06.362, M06.369, M06.37, M06.371, M06.372, M06.379, M06.38, M06.39, M06.4, M06.8, M06.80, M06.81, M06.811, M06.812, M06.819, M06.82, M06.821, M06.822, M06.829, M06.83, M06.831, M06.832, M06.839, M06.84, M06.841, M06.842, M06.849, M06.85, M06.851, M06.852, M06.859, M06.86, M06.861, M06.862, M06.869, M06.87, M06.871, M06.872, M06.879, M06.88, M06.89, M06.9, M05.7A, M05.8A, M06.0A, M06.8A, M08.1, M45, M45.A3, M45.A5, M45.A1, M45.A6, M45.A8, M45.A2, M45.A4, M45.A0, M45.A7, M45.AB, M45.A, M45.0, M45.1, M45.2, M45.3, M45.4, M45.5, M45.6, M45.7, M45.8, M45.9, Q05, Q07.01, Q07.03, Q76.0, Q05.0, Q05.1, Q05.2, Q05.3, Q05.4, Q05.5, Q05.6, Q05.7, Q05.8, Q05.9, G81, G83, I69.06, I69.16, I69.26, I69.36, I69.86, I69.96, G81.0, G81.00, G81.01, G81.02, G81.03, G81.04, G81.1, G81.10, G81.11, G81.12, G81.13, G81.14, G81.9, G81.90, G81.91, G81.92, G81.93, G81.94, G83.0, G83.1, G83.10, G83.11, G83.12, G83.13, G83.14, G83.2, G83.20, G83.21, G83.22, G83.23, G83.24, G83.3, G83.30, G83.31, G83.32, G83.33, G83.34, G83.4, G83.5, G83.8, G83.81, G83.82, G83.83, G83.84, G83.89, G83.9, I69.061, I69.062, I69.063, I69.064, I69.065, I69.069, I69.161, I69.162, I69.163, I69.164, I69.165, I69.169, I69.261, I69.262, I69.263, I69.264, I69.265, I69.269, I69.361, I69.362, I69.363, I69.364, I69.365, I69.369, I69.861, I69.862, I69.863, I69.864, I69.865, I69.869, I69.961, I69.962, I69.963, I69.964, I69.965, I69.969, S12.9, S14, S22.009S, S22.9XXS, S24, S32.0, S32.9XXS, S34, S12.9XXA, S12.9XXD, S12.9XXS, S14.0, S14.0XXA, S14.0XXD, S14.0XXS, S14.1, S14.10, S14.101, S14.101A, S14.101D, S14.101S, S14.102, S14.102A, S14.102D, S14.102S, S14.103, S14.103A, S14.103D, S14.103S, S14.104, S14.104A, S14.104D, S14.104S, S14.105, S14.105A, S14.105D, S14.105S, S14.106, S14.106A, S14.106D, S14.106S, S14.107, S14.107A, S14.107D, S14.107S, S14.108, S14.108A, S14.108D, S14.108S, S14.109, S14.109A, S14.109D, S14.109S, S14.11, S14.111, S14.111A, S14.111D, S14.111S, S14.112, S14.112A, S14.112D, S14.112S, S14.113, S14.113A, S14.113D, S14.113S, S14.114, S14.114A, S14.114D, S14.114S, S14.115, S14.115A, S14.115D, S14.115S, S14.116, S14.116A, S14.116D, S14.116S, S14.117, S14.117A, S14.117D, S14.117S, S14.118, S14.118A, S14.118D, S14.118S, S14.119, S14.119A, S14.119D, S14.119S, S14.12, S14.121, S14.121A, S14.121D, S14.121S, S14.122, S14.122A, S14.122D, S14.122S, S14.123, S14.123A, S14.123D, S14.123S, S14.124, S14.124A,

S14.124D, S14.124S, S14.125, S14.125A, S14.125D, S14.125S, S14.126, S14.126A, S14.126D, S14.126S, S14.127, S14.127A, S14.127D, S14.127S, S14.128, S14.128A, S14.128D, S14.128S, S14.129, S14.129A, S14.129D, S14.129S, S14.13, S14.131, S14.131A, S14.131D, S14.131S, S14.132, S14.132A, S14.132D, S14.132S, S14.133, S14.133A, S14.133D, S14.133S, S14.134, S14.134A, S14.134D, S14.134S, S14.135, S14.135A, S14.135D, S14.135S, S14.136, S14.136A, S14.136D, S14.136S, S14.137, S14.137A, S14.137D, S14.137S, S14.138, S14.138A, S14.138D, S14.138S, S14.139, S14.139A, S14.139D, S14.139S, S14.14, S14.141, S14.141A, S14.141D, S14.141S, S14.142, S14.142A, S14.142D, S14.142S, S14.143, S14.143A, S14.143D, S14.143S, S14.144, S14.144A, S14.144D, S14.144S, S14.145, S14.145A, S14.145D, S14.145S, S14.146, S14.146A, S14.146D, S14.146S, S14.147, S14.147A, S14.147D, S14.147S, S14.148, S14.148A, S14.148D, S14.148S, S14.149, S14.149A, S14.149D, S14.149S, S14.15, S14.151, S14.151A, S14.151D, S14.151S, S14.152, S14.152A, S14.152D, S14.152S, S14.153, S14.153A, S14.153D, S14.153S, S14.154, S14.154A, S14.154D, S14.154S, S14.155, S14.155A, S14.155D, S14.155S, S14.156, S14.156A, S14.156D, S14.156S, S14.157, S14.157A, S14.157D, S14.157S, S14.158, S14.158A, S14.158D, S14.158S, S14.159, S14.159A, S14.159D, S14.159S, S14.2, S14.2XXA, S14.2XXD, S14.2XXS, S14.3, S14.3XXA, S14.3XXD, S14.3XXS, S14.4, S14.4XXA, S14.4XXD, S14.4XXS, S14.5, S14.5XXA, S14.5XXD, S14.5XXS, S14.8, S14.8XXA, S14.8XXD, S14.8XXS, S14.9, S14.9XXA, S14.9XXD, S14.9XXS, S24.0, S24.0XXA, S24.0XXD, S24.0XXS, S24.1, S24.10, S24.101, S24.101A, S24.101D, S24.101S, S24.102, S24.102A, S24.102D, S24.102S, S24.103, S24.103A, S24.103D, S24.103S, S24.104, S24.104A, S24.104D, S24.104S, S24.109, S24.109A, S24.109D, S24.109S, S24.11, S24.111, S24.111A, S24.111D, S24.111S, S24.112, S24.112A, S24.112D, S24.112S, S24.113, S24.113A, S24.113D, S24.113S, S24.114, S24.114A, S24.114D, S24.114S, S24.119, S24.119A, S24.119D, S24.119S, S24.13, S24.131, S24.131A, S24.131D, S24.131S, S24.132, S24.132A, S24.132D, S24.132S, S24.133, S24.133A, S24.133D, S24.133S, S24.134, S24.134A, S24.134D, S24.134S, S24.139, S24.139A, S24.139D, S24.139S, S24.14, S24.141, S24.141A, S24.141D, S24.141S, S24.142, S24.142A, S24.142D, S24.142S, S24.143, S24.143A, S24.143D, S24.143S, S24.144, S24.144A, S24.144D, S24.144S, S24.149, S24.149A, S24.149D, S24.149S, S24.15, S24.151, S24.151A, S24.151D, S24.151S, S24.152, S24.152A, S24.152D, S24.152S, S24.153, S24.153A, S24.153D, S24.153S, S24.154, S24.154A, S24.154D, S24.154S, S24.159, S24.159A, S24.159D, S24.159S, S24.2, S24.2XXA, S24.2XXD, S24.2XXS, S24.3, S24.3XXA, S24.3XXD, S24.3XXS, S24.4, S24.4XXA, S24.4XXD, S24.4XXS, S24.8, S24.8XXA, S24.8XXD, S24.8XXS, S24.9, S24.9XXA, S24.9XXD, S24.9XXS, S32.00, S32.000, S32.000A, S32.000B, S32.000D, S32.000G, S32.000K, S32.000S, S32.001, S32.001A, S32.001B, S32.001D, S32.001G, S32.001K, S32.001S, S32.002, S32.002A, S32.002B, S32.002D, S32.002G, S32.002K, S32.002S, S32.008, S32.008A, S32.008B, S32.008D, S32.008G, S32.008K, S32.008S, S32.009, S32.009A, S32.009B, S32.009D, S32.009G, S32.009K, S32.009S, S32.01, S32.010, S32.010A, S32.010B, S32.010D, S32.010G, S32.010K, S32.010S, S32.011, S32.011A, S32.011B, S32.011D, S32.011G, S32.011K, S32.011S, S32.012, S32.012A, S32.012B, S32.012D, S32.012G, S32.012K, S32.012S, S32.018, S32.018A, S32.018B, S32.018D, S32.018G, S32.018K, S32.018S, S32.019, S32.019A, S32.019B, S32.019D, S32.019G, S32.019K, S32.019S, S32.02, S32.020, S32.020A, S32.020B, S32.020D, S32.020G, S32.020K, S32.020S, S32.021, S32.021A, S32.021B, S32.021D, S32.021G, S32.021K, S32.021S, S32.022, S32.022A, S32.022B, S32.022D, S32.022G, S32.022K, S32.022S, S32.028, S32.028A, S32.028B, S32.028D, S32.028G, S32.028K, S32.028S, S32.029, S32.029A, S32.029B, S32.029D, S32.029G, S32.029K, S32.029S, S32.03, S32.030, S32.030A, S32.030B, S32.032S, S32.030D, S32.030G, S32.030K, S32.030S, S32.031, S32.031A, S32.031B, S32.031D, S32.031G, S32.031K, S32.031S, S32.032, S32.032A, S32.032B, S32.032D, S32.032G, S32.032K, S32.038, S32.038A, S32.038B, S32.038D, S32.038G, S32.038K, S32.038S, S32.039, S32.039A, S32.039B, S32.039D, S32.039G, S32.039K, S32.039S, S32.04, S32.040, S32.040A, S32.040B, S32.042S, S32.040D, S32.040G, S32.040K, S32.040S, S32.041, S32.041A, S32.041B, S32.041D, S32.041G, S32.041K, S32.041S, S32.042, S32.042A, S32.042B, S32.042D, S32.042G, S32.042K, S32.048, S32.048A, S32.048B, S32.048D,

S32.048G, S32.048K, S32.048S, S32.049, S32.049A, S32.049B, S32.049D, S32.049G, S32.049K, S32.049S, S32.05, S32.050, S32.050A, S32.050B, S32.052S, S32.050D, S32.050G, S32.050K, S32.050S, S32.051, S32.051A, S32.051B, S32.051D, S32.051G, S32.051K, S32.051S, S32.052, S32.052A, S32.052B, S32.052D, S32.052G, S32.052K, S32.058, S32.058A, S32.058B, S32.058D, S32.058G, S32.058K, S32.058S, S32.059, S32.059A, S32.059B, S32.059D, S32.059G, S32.059K, S32.059S, S34.0, S34.01, S34.01XA, S34.01XD, S34.01XS, S34.02, S34.02XA, S34.02XD, S34.02XS, S34.1, S34.10, S34.101, S34.101A, S34.101D, S34.101S, S34.102, S34.102A, S34.102D, S34.102S, S34.103, S34.103A, S34.103D, S34.103S, S34.104, S34.104A, S34.104D, S34.104S, S34.105, S34.105A, S34.105D, S34.105S, S34.109, S34.109A, S34.109D, S34.109S, S34.11, S34.111, S34.111A, S34.111D, S34.111S, S34.112, S34.112A, S34.112D, S34.112S, S34.113, S34.113A, S34.113D, S34.113S, S34.114, S34.114A, S34.114D, S34.114S, S34.115, S34.115A, S34.115D, S34.115S, S34.119, S34.119A, S34.119D, S34.119S, S34.12, S34.121, S34.121A, S34.121D, S34.121S, S34.122, S34.122A, S34.122D, S34.122S, S34.123, S34.123A, S34.123D, S34.123S, S34.124, S34.124A, S34.124D, S34.124S, S34.125, S34.125A, S34.125D, S34.125S, S34.129, S34.129A, S34.129D, S34.129S, S34.13, S34.131, S34.131A, S34.131D, S34.131S, S34.132, S34.132A, S34.132D, S34.132S, S34.139, S34.139A, S34.139D, S34.139S, S34.2, S34.21, S34.21XA, S34.21XD, S34.21XS, S34.22, S34.22XA, S34.22XD, S34.22XS, S34.3, S34.3XXA, S34.3XXD, S34.3XXS, S34.4, S34.4XXA, S34.4XXD, S34.4XXS, S34.5, S34.5XXA, S34.5XXD, S34.5XXS, S34.6, S34.6XXA, S34.6XXD, S34.6XXS, S34.8, S34.8XXA, S34.8XXD, S34.8XXS, S34.9, S34.9XXA, S34.9XXD, S34.9XXS, I69.9, I69.90, I69.91, I69.910, I69.911, I69.912, I69.913, I69.914, I69.915, I69.918, I69.919, I69.92, I69.920, I69.921, I69.922, I69.923, I69.928, I69.93, I69.931, I69.932, I69.933, I69.934, I69.939, I69.94, I69.941, I69.942, I69.943, I69.944, I69.949, I69.95, I69.951, I69.952, I69.953, I69.954, I69.959, I69.96, I69.961, I69.962, I69.963, I69.964, I69.965, I69.969, I69.99, I69.990, I69.991, I69.992, I69.993, I69.998, A80.0, A80.1, A80.2, A80.3, A80.9, B91, G14, A80.30, A80.39, G70, G70.0, G70.00, G70.01, G70.1, G70.2, G70.8, G70.80, G70.81, G70.89, G70.9, G71, G71.0, G71.00, G71.01, G71.02, G71.09, G71.1, G71.11, G71.12, G71.13, G71.14, G71.19, G71.2, G71.3, G71.8, G71.9, G71.20, G71.21, G71.22, G71.220, G71.228, G71.29, Q77.0, Q77.1, Z99.3, Z99.89, Q85.0, Q85.00, Q85.01, Q85.02, Q85.03, Q85.09, G31.89, G20, G21, G21.0, G21.1, G21.11, G21.19, G21.2, G21.3, G21.4, G21.8, G21.9, G10, G23, G24, G25, G80.3, G23.0, G23.1, G23.2, G23.8, G23.9, G24.0, G24.01, G24.02, G24.09, G24.1, G24.2, G24.3, G24.4, G24.5, G24.8, G24.9, G25.0, G25.1, G25.2, G25.3, G25.4, G25.5, G25.6, G25.61, G25.69, G25.7, G25.70, G25.71, G25.79, G25.8, G25.81, G25.82, G25.83, G25.89, G25.9, G11, G32.81, G11.0, G11.1, G11.2, G11.3, G11.4, G11.8, G11.9, G11.10, G11.11, G11.19, G12, G12.0, G12.1, G12.2, G12.20, G12.21, G12.22, G12.23, G12.24, G12.25, G12.29, G12.8, G12.9, G32.0, G95, G95.0, G95.1, G95.11, G95.19, G95.2, G95.20, G95.29, G95.8, G95.81, G95.89, G95.9, G90, G99, G90.0, G90.01, G90.09, G90.1, G90.2, G90.3, G90.4, G90.5, G90.50, G90.51, G90.511, G90.512, G90.513, G90.519, G90.52, G90.521, G90.522, G90.523, G90.529, G90.59, G90.8, G90.9, G99.0, G99.2, G99.8, G36.0, G37, G37.0, G37.1, G37.2, G37.3, G37.4, G37.5, G37.8, G37.9, G40, G40.0, G40.00, G40.001, G40.009, G40.01, G40.011, G40.019, G40.1, G40.10, G40.101, G40.109, G40.11, G40.111, G40.119, G40.2, G40.20, G40.201, G40.209, G40.21, G40.50, G40.211, G40.219, G40.3, G40.30, G40.301, G40.309, G40.31, G40.311, G40.319, G40.4, G40.40, G40.401, G40.409, G40.41, G40.411, G40.419, G40.5, G40.501, G40.509, G40.8, G40.80, G40.801, G40.802, G40.803, G40.804, G40.81, G40.811, G40.812, G40.813, G40.814, G40.82, G40.821, G40.822, G40.823, G40.824, G40.89, G40.9, G40.90, G40.901, G40.909, G40.91, G40.911, G40.919, G40.A, G40.A0, G40.A01, G40.A09, G40.A1, G40.A11, G40.A19, G40.B, G40.B0, G40.B01, G40.B09, G40.B1, G40.B11, G40.B19, G40.42, G40.83, G40.833, G40.834, G54, G56, G54.0, G54.1, G54.2, G54.3, G54.4, G54.5, G54.6, G54.7, G54.8, G54.9, G56.0, G56.00, G56.01, G56.02, G56.03, G56.1, G56.10, G56.11, G56.12, G56.13, G56.2, G56.20, G56.21, G56.22, G56.23, G56.3, G56.30, G56.31, G56.32, G56.33, G56.4,

G56.40, G56.41, G56.42, G56.43, G56.8, G56.80, G56.81, G56.82, G56.83, G56.9, G56.90, G56.91, G56.92, G56.93, G60, G60.0, G60.1, G60.2, G60.3, G60.8, G60.9, E08.42, E09.42, E10.42, E11.42, E13.42, G61, G62, G63, G61.0, G61.1, G61.8, G61.81, G61.82, G61.89, G61.9, G62.0, G62.1, G62.2, G62.8, G62.81, G62.82, G62.89, G62.9, E22, E22.0, E22.1, E22.2, E22.8, E22.9, M15, M16, M17, M18, M19, M15.0, M15.1, M15.2, M15.3, M15.4, M15.8, M15.9, M16.0, M16.1, M16.10, M16.11, M16.12, M16.2, M16.3, M16.30, M16.31, M16.32, M16.4, M16.5, M16.50, M16.51, M16.52, M16.6, M16.7, M16.9, M17.0, M17.1, M17.10, M17.11, M17.12, M17.2, M17.3, M17.30, M17.31, M17.32, M17.4, M17.5, M17.9, M18.0, M18.1, M18.10, M18.11, M18.12, M18.2, M18.3, M18.30, M18.31, M18.32, M18.4, M18.5, M18.50, M18.51, M18.52, M18.9, M19.0, M19.01, M19.011, M19.012, M19.019, M19.02, M19.021, M19.022, M19.029, M19.03, M19.031, M19.032, M19.039, M19.04, M19.041, M19.042, M19.049, M19.07, M19.071, M19.072, M19.079, M19.1, M19.11, M19.111, M19.112, M19.119, M19.12, M19.121, M19.122, M19.129, M19.13, M19.131, M19.132, M19.139, M19.14, M19.141, M19.142, M19.149, M19.17, M19.171, M19.172, M19.179, M19.2, M19.21, M19.211, M19.212, M19.219, M19.22, M19.221, M19.222, M19.229, M19.23, M19.231, M19.232, M19.239, M19.24, M19.241, M19.242, M19.249, M19.27, M19.271, M19.272, M19.279, M19.9, M19.90, M19.91, M19.92, M19.93, M19.09, M19.19, M19.29, M23, M23.0, M23.00, M23.000, M23.001, M23.002, M23.003, M23.004, M23.005, M23.006, M23.007, M23.009, M23.01, M23.011, M23.012, M23.019, M23.02, M23.021, M23.022, M23.029, M23.03, M23.031, M23.032, M23.039, M23.04, M23.041, M23.042, M23.049, M23.05, M23.051, M23.052, M23.059, M23.06, M23.061, M23.062, M23.069, M23.2, M23.20, M23.200, M23.201, M23.202, M23.203, M23.204, M23.205, M23.206, M23.207, M23.209, M23.21, M23.211, M23.212, M23.219, M23.22, M23.221, M23.222, M23.229, M23.23, M23.231, M23.232, M23.239, M23.24, M23.241, M23.242, M23.249, M23.25, M23.251, M23.252, M23.259, M23.26, M23.261, M23.262, M23.269, M23.3, M23.30, M23.300, M23.301, M23.302, M23.303, M23.304, M23.305, M23.306, M23.307, M23.309, M23.31, M23.311, M23.312, M23.319, M23.32, M23.321, M23.322, M23.329, M23.33, M23.331, M23.332, M23.339, M23.34, M23.341, M23.342, M23.349, M23.35, M23.351, M23.352, M23.359, M23.36, M23.361, M23.362, M23.369, M23.4, M23.40, M23.41, M23.42, M23.5, M23.50, M23.51, M23.52, M23.6, M23.60, M23.601, M23.602, M23.609, M23.61, M23.611, M23.612, M23.619, M23.62, M23.621, M23.622, M23.629, M23.63, M23.631, M23.632, M23.639, M23.64, M23.641, M23.642, M23.649, M23.67, M23.671, M23.672, M23.679, M23.8, M23.8X, M23.8X1, M23.8X2, M23.8X9, M23.9, M23.90, M23.91, M23.92, M47, M48, M47.0, M47.01, M47.011, M47.012, M47.013, M47.014, M47.015, M47.016, M47.019, M47.02, M47.021, M47.022, M47.029, M47.1, M47.10, M47.11, M47.12, M47.13, M47.14, M47.15, M47.16, M47.2, M47.20, M47.21, M47.22, M47.23, M47.24, M47.25, M47.26, M47.27, M47.28, M47.8, M47.81, M47.811, M47.812, M47.813, M47.814, M47.815, M47.816, M47.817, M47.818, M47.819, M47.89, M47.891, M47.892, M47.893, M47.894, M47.895, M47.896, M47.897, M47.898, M47.899, M47.9, M48.0, M48.00, M48.01, M48.02, M48.03, M48.04, M48.05, M48.06, M48.061, M48.062, M48.07, M48.08, M48.1, M48.10, M48.11, M48.12, M48.13, M48.14, M48.15, M48.16, M48.17, M48.18, M48.19, M48.2, M48.20, M48.21, M48.22, M48.23, M48.24, M48.25, M48.26, M48.27, M48.3, M48.30, M48.31, M48.32, M48.33, M48.34, M48.35, M48.36, M48.37, M48.38, M48.4, M48.40, M48.40XA, M48.40XD, M48.40XG, M48.40XS, M48.41, M48.41XA, M48.41XD, M48.41XG, M48.41XS, M48.42, M48.42XA, M48.42XD, M48.42XG, M48.42XS, M48.43, M48.43XA, M48.43XD, M48.43XG, M48.43XS, M48.44, M48.44XA, M48.44XD, M48.44XG, M48.44XS, M48.45, M48.45XA, M48.45XD, M48.45XG, M48.45XS, M48.46, M48.46XA, M48.46XD, M48.46XG, M48.46XS, M48.47, M48.47XA, M48.47XD, M48.47XG, M48.47XS, M48.48, M48.48XA, M48.48XD, M48.48XG, M48.48XS, M48.5, M48.50, M48.50XA, M48.50XD, M48.50XG, M48.50XS, M48.51, M48.51XA, M48.51XD, M48.51XG, M48.51XS, M48.52, M48.52XA, M48.52XD, M48.52XG, M48.52XS, M48.53,

M48.53XA, M48.53XD, M48.53XG, M48.53XS, M48.54, M48.54XA, M48.54XD, M48.54XG, M48.54XS, M48.55, M48.55XA, M48.55XD, M48.55XG, M48.55XS, M48.56, M48.56XA, M48.56XD, M48.56XG, M48.56XS, M48.57, M48.57XA, M48.57XD, M48.57XG, M48.57XS, M48.58, M48.58XA, M48.58XD, M48.58XG, M48.58XS, M48.8, M48.8X, M48.8X1, M48.8X2, M48.8X3, M48.8X4, M48.8X5, M48.8X6, M48.8X7, M48.8X8, M48.8X9, M48.9, M50, M51, M50.0, M50.00, M50.01, M50.02, M50.020, M50.021, M50.022, M50.023, M50.03, M50.1, M50.10, M50.11, M50.12, M50.120, M50.121, M50.122, M50.123, M50.13, M50.2, M50.20, M50.21, M50.22, M50.220, M50.221, M50.222, M50.223, M50.23, M50.3, M50.30, M50.31, M50.32, M50.320, M50.321, M50.322, M50.323, M50.33, M50.8, M50.80, M50.81, M50.82, M50.820, M50.821, M50.822, M50.823, M50.83, M50.9, M50.90, M50.91, M50.92, M50.920, M50.921, M50.922, M50.923, M50.93, M51.0, M51.04, M51.05, M51.06, M51.1, M51.14, M51.15, M51.16, M51.17, M51.2, M51.24, M51.25, M51.26, M51.27, M51.3, M51.34, M51.35, M51.36, M51.37, M51.4, M51.44, M51.45, M51.46, M51.47, M51.8, M51.84, M51.85, M51.86, M51.87, M51.9, M48.02, M53.0, M53.1, M54.2, M24.0, M35.7, M43.6, M53.82, M54.02, M54.12, M54.13, M60, M60.0, M60.1, M60.2, M61, M62, M63, M67.88, M72, M24.00, M24.01, M24.011, M24.012, M24.019, M24.02, M24.021, M24.022, M24.029, M24.03, M24.031, M24.032, M24.039, M24.04, M24.041, M24.042, M24.049, M24.05, M24.051, M24.052, M24.059, M24.07, M24.071, M24.072, M24.073, M24.074, M24.075, M24.076, M24.08, M60.00, M60.000, M60.001, M60.002, M60.003, M60.004, M60.005, M60.009, M60.01, M60.011, M60.012, M60.019, M60.02, M60.021, M60.022, M60.029, M60.03, M60.031, M60.032, M60.039, M60.04, M60.041, M60.042, M60.043, M60.044, M60.045, M60.046, M60.05, M60.051, M60.052, M60.059, M60.06, M60.061, M60.062, M60.069, M60.07, M60.070, M60.071, M60.072, M60.073, M60.074, M60.075, M60.076, M60.077, M60.078, M60.08, M60.09, M60.10, M60.11, M60.111, M60.112, M60.119, M60.12, M60.121, M60.122, M60.129, M60.13, M60.131, M60.132, M60.139, M60.14, M60.141, M60.142, M60.149, M60.15, M60.151, M60.152, M60.159, M60.16, M60.161, M60.162, M60.169, M60.17, M60.171, M60.172, M60.179, M60.18, M60.19, M60.20, M60.21, M60.211, M60.212, M60.219, M60.22, M60.221, M60.222, M60.229, M60.23, M60.231, M60.232, M60.239, M60.24, M60.241, M60.242, M60.249, M60.25, M60.251, M60.252, M60.259, M60.26, M60.261, M60.262, M60.269, M60.27, M60.271, M60.272, M60.279, M60.28, M60.8, M60.80, M60.81, M60.811, M60.812, M60.819, M60.82, M60.821, M60.822, M60.829, M60.83, M60.831, M60.832, M60.839, M60.84, M60.841, M60.842, M60.849, M60.85, M60.851, M60.852, M60.859, M60.86, M60.861, M60.862, M60.869, M60.87, M60.871, M60.872, M60.879, M60.88, M60.89, M60.9, M61.0, M61.00, M61.01, M61.011, M61.012, M61.019, M61.02, M61.021, M61.022, M61.029, M61.03, M61.031, M61.032, M61.039, M61.04, M61.041, M61.042, M61.049, M61.05, M61.051, M61.052, M61.059, M61.06, M61.061, M61.062, M61.069, M61.07, M61.071, M61.072, M61.079, M61.08, M61.09, M61.1, M61.10, M61.11, M61.111, M61.112, M61.119, M61.12, M61.121, M61.122, M61.129, M61.13, M61.131, M61.132, M61.139, M61.14, M61.141, M61.142, M61.143, M61.144, M61.145, M61.146, M61.15, M61.151, M61.152, M61.159, M61.16, M61.161, M61.162, M61.169, M61.17, M61.171, M61.172, M61.173, M61.174, M61.175, M61.176, M61.177, M61.178, M61.179, M61.18, M61.19, M61.2, M61.20, M61.21, M61.211, M61.212, M61.219, M61.22, M61.221, M61.222, M61.229, M61.23, M61.231, M61.232, M61.239, M61.24, M61.241, M61.242, M61.249, M61.25, M61.251, M61.252, M61.259, M61.26, M61.261, M61.262, M61.269, M61.27, M61.271, M61.272, M61.279, M61.28, M61.29, M61.3, M61.30, M61.31, M61.311, M61.312, M61.319, M61.32, M61.321, M61.322, M61.329, M61.33, M61.331, M61.332, M61.339, M61.34, M61.341, M61.342, M61.349, M61.35, M61.351, M61.352, M61.359, M61.36, M61.361, M61.362, M61.369, M61.37, M61.371, M61.372, M61.379, M61.38, M61.39, M61.4, M61.40, M61.41, M61.411, M61.412, M61.419, M61.42, M61.421, M61.422, M61.429, M61.43, M61.431, M61.432, M61.439, M61.44, M61.441, M61.442, M61.449, M61.45, M61.451, M61.452, M61.459, M61.46, M61.461,

M61.462, M61.469, M61.47, M61.471, M61.472, M61.479, M61.48, M61.49, M61.5, M61.50, M61.51, M61.511, M61.512, M61.519, M61.52, M61.521, M61.522, M61.529, M61.53, M61.531, M61.532, M61.539, M61.54, M61.541, M61.542, M61.549, M61.55, M61.551, M61.552, M61.559, M61.56, M61.561, M61.562, M61.569, M61.57, M61.571, M61.572, M61.579, M61.58, M61.59, M61.9, M62.0, M62.00, M62.01, M62.011, M62.012, M62.019, M62.02, M62.021, M62.022, M62.029, M62.03, M62.031, M62.032, M62.039, M62.04, M62.041, M62.042, M62.049, M62.05, M62.051, M62.052, M62.059, M62.06, M62.061, M62.062, M62.069, M62.07, M62.071, M62.072, M62.079, M62.08, M62.1, M62.10, M62.11, M62.111, M62.112, M62.119, M62.12, M62.121, M62.122, M62.129, M62.13, M62.131, M62.132, M62.139, M62.14, M62.141, M62.142, M62.149, M62.15, M62.151, M62.152, M62.159, M62.16, M62.161, M62.162, M62.169, M62.17, M62.171, M62.172, M62.179, M62.18, M62.2, M62.20, M62.21, M62.211, M62.212, M62.219, M62.22, M62.221, M62.222, M62.229, M62.23, M62.231, M62.232, M62.239, M62.24, M62.241, M62.242, M62.249, M62.25, M62.251, M62.252, M62.259, M62.26, M62.261, M62.262, M62.269, M62.27, M62.271, M62.272, M62.279, M62.28, M62.3, M62.4, M62.40, M62.41, M62.411, M62.412, M62.419, M62.42, M62.421, M62.422, M62.429, M62.43, M62.431, M62.432, M62.439, M62.44, M62.441, M62.442, M62.449, M62.45, M62.451, M62.452, M62.459, M62.46, M62.461, M62.462, M62.469, M62.47, M62.471, M62.472, M62.479, M62.48, M62.49, M62.5, M62.50, M62.51, M62.511, M62.512, M62.519, M62.52, M62.521, M62.522, M62.529, M62.53, M62.531, M62.532, M62.539, M62.54, M62.541, M62.542, M62.549, M62.55, M62.551, M62.552, M62.559, M62.56, M62.561, M62.562, M62.569, M62.57, M62.571, M62.572, M62.579, M62.58, M62.59, M62.8, M62.81, M62.82, M62.83, M62.830, M62.831, M62.838, M62.84, M62.89, M62.9, M63.8, M63.80, M63.81, M63.811, M63.812, M63.819, M63.82, M63.821, M63.822, M63.829, M63.83, M63.831, M63.832, M63.839, M63.84, M63.841, M63.842, M63.849, M63.85, M63.851, M63.852, M63.859, M63.86, M63.861, M63.862, M63.869, M63.87, M63.871, M63.872, M63.879, M63.88, M63.89, M72.0, M72.1, M72.2, M72.4, M72.6, M72.8, M72.9, M42, M91, M92, M93, M42.0, M42.00, M42.01, M42.02, M42.03, M42.04, M42.05, M42.06, M42.07, M42.08, M42.09, M42.1, M42.10, M42.11, M42.12, M42.13, M42.14, M42.15, M42.16, M42.17, M42.18, M42.19, M42.9, M91.0, M91.1, M91.10, M91.11, M91.12, M91.2, M91.20, M91.21, M91.22, M91.3, M91.30, M91.31, M91.32, M91.4, M91.40, M91.41, M91.42, M91.8, M91.80, M91.81, M91.82, M91.9, M91.90, M91.91, M91.92, M92.0, M92.00, M92.01, M92.02, M92.1, M92.10, M92.11, M92.12, M92.2, M92.20, M92.201, M92.202, M92.209, M92.21, M92.211, M92.212, M92.219, M92.22, M92.221, M92.222, M92.229, M92.29, M92.291, M92.292, M92.299, M92.3, M92.30, M92.31, M92.32, M92.4, M92.40, M92.41, M92.42, M92.5, M92.50, M92.51, M92.52, M92.6, M92.60, M92.61, M92.62, M92.7, M92.70, M92.71, M92.72, M92.8, M92.9, M93.0, M93.00, M93.001, M93.002, M93.003, M93.01, M93.011, M93.012, M93.013, M93.02, M93.021, M93.022, M93.023, M93.03, M93.031, M93.032, M93.033, M93.1, M93.2, M93.20, M93.21, M93.211, M93.212, M93.219, M93.22, M93.221, M93.222, M93.229, M93.23, M93.231, M93.232, M93.239, M93.24, M93.241, M93.242, M93.249, M93.25, M93.251, M93.252, M93.259, M93.26, M93.261, M93.262, M93.269, M93.27, M93.271, M93.272, M93.279, M93.28, M93.29, M93.8, M93.80, M93.81, M93.811, M93.812, M93.819, M93.82, M93.821, M93.822, M93.829, M93.83, M93.831, M93.832, M93.839, M93.84, M93.841, M93.842, M93.849, M93.85, M93.851, M93.852, M93.859, M93.86, M93.861, M93.862, M93.869, M93.87, M93.871, M93.872, M93.879, M93.88, M93.89, M93.9, M93.90, M93.91, M93.911, M93.912, M93.919, M93.92, M93.921, M93.922, M93.929, M93.93, M93.931, M93.932, M93.939, M93.94, M93.941, M93.942, M93.949, M93.95, M93.951, M93.952, M93.959, M93.96, M93.961, M93.962, M93.969, M93.97, M93.971, M93.972, M93.979, M93.98, M93.99, M92.501, M92.502, M92.503, M92.509, M92.511, M92.512, M92.513, M92.519, M92.521, M92.522, M92.523, M92.529, M92.59, M92.591, M92.592, M92.593, M92.599, S77, S87, S97, S77.11XD, S77.0, S77.00, S77.00XA, S77.00XD, S77.00XS, S77.01, S77.01XA,

S77.01XD, S77.01XS, S77.02, S77.02XA, S77.02XD, S77.02XS, S77.1, S77.10, S77.10XA, S77.10XD, S77.10XS, S77.11, S77.11XA, S77.11XS, S77.12, S77.12XA, S77.12XD, S77.12XS, S77.2, S77.20, S77.20XA, S77.20XD, S77.20XS, S77.21, S77.21XA, S77.21XD, S77.21XS, S77.22, S77.22XA, S77.22XD, S77.22XS, S87.81XD, S87.81XS, S87.82, S87.82XA, S87.82XD, S87.0, S87.00, S87.00XA, S87.00XD, S87.00XS, S87.01, S87.01XA, S87.01XD, S87.01XS, S87.02, S87.02XA, S87.02XD, S87.02XS, S87.8, S87.80, S87.80XA, S87.80XD, S87.80XS, S87.81, S87.81XA, S87.82XS, S97.00XS, S97.01, S97.01XA, S97.01XD, S97.01XS, S97.02, S97.02XA, S97.0, S97.00, S97.00XA, S97.00XD, S97.02XD, S97.02XS, S97.1, S97.10, S97.101, S97.101A, S97.101D, S97.101S, S97.102, S97.102A, S97.102D, S97.102S, S97.109, S97.109A, S97.109D, S97.109S, S97.11, S97.111, S97.111A, S97.111D, S97.111S, S97.112, S97.112A, S97.112D, S97.112S, S97.119, S97.119A, S97.119D, S97.119S, S97.12, S97.121, S97.121A, S97.121D, S97.121S, S97.122, S97.122A, S97.122D, S97.122S, S97.129, S97.129A, S97.129D, S97.129S, S97.8, S97.80, S97.80XA, S97.80XD, S97.80XS, S97.81, S97.81XA, S97.81XD, S97.81XS, S97.82, S97.82XA, S97.82XD, S97.82XS, M21, Z89, M21.0, M21.00, M21.02, M21.021, M21.022, M21.029, M21.05, M21.051, M21.052, M21.059, M21.06, M21.061, M21.062, M21.069, M21.07, M21.071, M21.072, M21.079, M21.1, M21.10, M21.12, M21.121, M21.122, M21.129, M21.15, M21.151, M21.152, M21.159, M21.16, M21.161, M21.162, M21.169, M21.17, M21.171, M21.172, M21.179, M21.2, M21.20, M21.21, M21.211, M21.212, M21.219, M21.22, M21.221, M21.222, M21.229, M21.23, M21.231, M21.232, M21.239, M21.24, M21.241, M21.242, M21.249, M21.25, M21.251, M21.252, M21.259, M21.26, M21.261, M21.262, M21.269, M21.27, M21.271, M21.272, M21.279, M21.3, M21.33, M21.331, M21.332, M21.339, M21.37, M21.371, M21.372, M21.379, M21.4, M21.40, M21.41, M21.42, M21.5, M21.51, M21.511, M21.512, M21.519, M21.52, M21.521, M21.522, M21.529, M21.53, M21.531, M21.532, M21.539, M21.54, M21.541, M21.542, M21.549, M21.6, M21.61, M21.611, M21.612, M21.619, M21.62, M21.621, M21.622, M21.629, M21.6X, M21.6X1, M21.6X2, M21.6X9, M21.7, M21.70, M21.72, M21.721, M21.722, M21.729, M21.73, M21.731, M21.732, M21.733, M21.734, M21.739, M21.75, M21.751, M21.752, M21.759, M21.76, M21.761, M21.762, M21.763, M21.764, M21.769, M21.8, M21.80, M21.82, M21.821, M21.822, M21.829, M21.83, M21.831, M21.832, M21.839, M21.85, M21.851, M21.852, M21.859, M21.86, M21.861, M21.862, M21.869, M21.9, M21.90, M21.92, M21.921, M21.922, M21.929, M21.93, M21.931, M21.932, M21.939, M21.94, M21.941, M21.942, M21.949, M21.95, M21.951, M21.952, M21.959, M21.96, M21.961, M21.962, M21.969, Z89.0, Z89.01, Z89.011, Z89.012, Z89.019, Z89.02, Z89.021, Z89.022, Z89.029, Z89.1, Z89.11, Z89.111, Z89.112, Z89.119, Z89.12, Z89.121, Z89.122, Z89.129, Z89.2, Z89.20, Z89.201, Z89.202, Z89.209, Z89.21, Z89.211, Z89.212, Z89.219, Z89.22, Z89.221, Z89.222, Z89.229, Z89.23, Z89.231, Z89.232, Z89.239, Z89.4, Z89.41, Z89.411, Z89.412, Z89.419, Z89.42, Z89.421, Z89.422, Z89.429, Z89.43, Z89.431, Z89.432, Z89.439, Z89.44, Z89.441, Z89.442, Z89.449, Z89.5, Z89.51, Z89.511, Z89.512, Z89.519, Z89.52, Z89.521, Z89.522, Z89.529, Z89.6, Z89.61, Z89.611, Z89.612, Z89.619, Z89.62, Z89.621, Z89.622, Z89.629, Z89.9, E84.0, E84.11, E84.19, E84.8, E84.9, E34.3, G90.1, Q01, Q02, Q03, Q04, Q06, Q07.8, Q07.9, Q01.0, Q01.1, Q01.2, Q01.8, Q01.9, Q03.0, Q03.1, Q03.8, Q03.9, Q04.0, Q04.1, Q04.2, Q04.3, Q04.4, Q04.5, Q04.6, Q04.8, Q04.9, Q06.0, Q06.1, Q06.2, Q06.3, Q06.4, Q06.8, Q06.9, Q70, Q71, Q72, Q74.8, Q70.0, Q70.00, Q70.01, Q70.02, Q70.03, Q70.1, Q70.10, Q70.11, Q70.12, Q70.13, Q70.2, Q70.20, Q70.21, Q70.22, Q70.23, Q70.3, Q70.30, Q70.31, Q70.32, Q70.33, Q70.4, Q70.9, Q71.0, Q71.00, Q71.01, Q71.02, Q71.03, Q71.1, Q71.10, Q71.11, Q71.12, Q71.13, Q71.2, Q71.20, Q71.21, Q71.22, Q71.23, Q71.3, Q71.30, Q71.31, Q71.32, Q71.33, Q71.4, Q71.40, Q71.41, Q71.42, Q71.43, Q71.5, Q71.50, Q71.51, Q71.52, Q71.53, Q71.6, Q71.60, Q71.61, Q71.62, Q71.63, Q71.8, Q71.81, Q71.811, Q71.812, Q71.813, Q71.819, Q71.89, Q71.891, Q71.892, Q71.893, Q71.899, Q71.9, Q71.90, Q71.91, Q71.92, Q71.93, Q72.0, Q72.00, Q72.01, Q72.02, Q72.03, Q72.1, Q72.10, Q72.11, Q72.12, Q72.13, Q72.2, Q72.20, Q72.21, Q72.22, Q72.23, Q72.3, Q72.30, Q72.31, Q72.32,

Q72.33, Q72.4, Q72.40, Q72.41, Q72.42, Q72.43, Q72.5, Q72.50, Q72.51, Q72.52, Q72.53, Q72.6, Q72.60, Q72.61, Q72.62, Q72.63, Q72.7, Q72.70, Q72.71, Q72.72, Q72.73, Q72.8, Q72.81, Q72.811, Q72.812, Q72.813, Q72.819, Q72.89, Q72.891, Q72.892, Q72.893, Q72.899, Q72.9, Q72.90, Q72.91, Q72.92, Q72.93, Q66, Q66.0, Q66.00, Q66.01, Q66.02, Q66.1, Q66.10, Q66.11, Q66.12, Q66.2, Q66.21, Q66.211, Q66.212, Q66.219, Q66.22, Q66.221, Q66.222, Q66.229, Q66.3, Q66.30, Q66.31, Q66.32, Q66.4, Q66.40, Q66.41, Q66.42, Q66.5, Q66.50, Q66.51, Q66.52, Q66.6, Q66.7, Q66.70, Q66.71, Q66.72, Q66.8, Q66.80, Q66.81, Q66.82, Q66.89, Q66.9, Q66.90, Q66.91, Q66.92, Q68.8, Q75.0, Q75.2, Q75.9, Q76.0, Q76.1, Q76.2, Q76.419, Q76.49, Q76.5, Q76.6, Q76.7, Q77.1, Q77.2, Q77.4, Q77.6, Q77.8, Q78.0, Q78.1, Q78.2, Q78.3, Q78.4, Q78.6, Q78.8, Q78.9, Q79.0, Q79.1, Q79.2, Q79.3, Q79.4, Q79.51, Q79.59, Q79.6, Q79.8, Q79.9, Q79.60, Q79.61, Q79.62, Q79.63, Q79.69

SNOMED CT: 1186728004, 16092000, 18353007, 24700007, 816984002, 724778008, 766246000, 155023009, 192926004, 192927008, 192928003, 192930001, 230372003, 230373008, 230374002, 425500002, 426373005, 428700003, 438511000, 439567002, 733028000, 192929006, 1178005, 128188000, 43486001, 702314005, 702315006, 702316007, 702317003, 702318008, 702319000, 702320006, 702321005, 192947000, 192950002, 192958009, 192961005, 194546004, 307756005, 371079004, 230772000, 230773005, 230776002, 230777006, 230778001, 230780007, 230781006, 48721008, 56409008, 58193001, 75019001, 270488006, 275466008, 275467004, 278512001, 16171003, 329741000119109, 330521000119103, 716335003, 783005002, 44395000, 275468009, 12396006, 781206002, 160413008, 161567008, 201766009, 201769002, 201771002, 201775006, 201777003, 201778008, 201779000, 239791005, 239792003, 429192004, 287006005, 287007001, 1222679006, 1162319009, 1162313005, 1162303002, 1162362005, 1162364006, 1073601000119101, 1073611000119103, 1156774008, 1156777001, 15691081000119100, 15691121000119103, 15691841000119102, 15691921000119100, 15691961000119105, 15692001000119101, 15692041000119104, 16606681000119101, 318841000119100, 318851000119103, 318891000119108, 318901000119107, 318911000119105, 318921000119103, 318961000119108, 318971000119102, 319011000119103, 319041000119104, 319051000119102, 319061000119100, 319071000119106, 319091000119107, 319121000119106, 319131000119109, 319141000119100, 319151000119103, 318871000119107, 318881000119105, 318941000119109, 318951000119106, 319031000119108, 319081000119109, 319111000119104, 1073681000119109, 1073691000119107, 1073711000119105, 1073721000119103, 1073731000119100, 1073741000119109, 1073751000119106, 1073761000119108, 1073771000119102, 1073791000119101, 1073801000119100, 1073811000119102, 1073821000119109, 1073831000119107, 1105151000119108, 15686281000119101, 15686321000119106, 15687201000119107, 15687321000119109, 15687841000119108, 15691161000119108, 15691721000119102, 15691761000119107, 15691801000119104, 1073701000119107, 1073781000119104, 201764007, 201770001, 201772009, 201773004, 201774005, 201780002, 201781003, 201783000, 201784006, 201785007, 308143008, 410796000, 15691241000119101, 15691321000119101, 15691881000119107, 16050071000119108, 16606721000119107, 318861000119101, 318931000119100, 69896004, 1162336001, 1222679006, 1162319009, 1162313005, 1162303002, 1162362005, 1162364006, 7607008, 10713006, 28880005, 33719002, 82939003, 86219005, 129563009, 1073601000119101, 1073611000119103, 1149218009, 1149219001, 1156774008, 1156777001, 15691081000119100, 15691121000119103, 15691841000119102, 15691921000119100, 15691961000119105, 15692001000119101, 15692041000119104, 16606681000119101, 318841000119100, 318851000119103, 318891000119108, 318901000119107, 318911000119105, 318921000119103, 318961000119108, 318971000119102, 319011000119103, 319041000119104, 319051000119102, 319061000119100, 319071000119106, 319091000119107, 319121000119106, 319131000119109, 319141000119100, 319151000119103, 1162677006, 143441000119108, 318871000119107, 318881000119105, 318941000119109, 318951000119106, 319031000119108, 319081000119109, 319111000119104, 319841000119107, 319861000119106, 319871000119100, 319941000119103, 319951000119101, 459911000124100, 459921000124108, 1073681000119109, 1073691000119107, 1073711000119105, 1073721000119103, 1073731000119100, 1073741000119109,

1073751000119106, 1073761000119108, 1073771000119102, 1073791000119101, 1073801000119100, 1073811000119102, 1073821000119109, 1073831000119107, 1105151000119108, 1567361000119100, 1567352100011910 1, 1568592100011910 2, 1568596100011910 7, 1568600100011910 4, 1568628100011910 1, 1568632100011910 6, 1568720100011910 7, 1568732100011910 9, 1568784100011910 8, 1569116100011910 8, 1569172100011910 2, 1569176100011910 7, 1569180100011910 4, 57160007, 781206002, 1073701000119107, 1073781000119104, 735599007, 735600005, 201764007, 201766009, 201767000, 201768005, 201769002, 201770001, 201771002, 201772009, 201773004, 201774005, 201775006, 201776007, 201777003, 201778008, 201779000, 201780002, 201781003, 201783000, 201784006, 201785007, 201789001, 201791009, 308143008, 239791005, 239792003, 239795001, 239943002, 38877003, 52661003, 54867000, 59165007, 77522006, 398640008, 398726004, 399923009, 400054000, 402426007, 402427003, 402428008, 402431009, 402432002, 402433007, 402434001, 410796000, 427770001, 429192004, 287006005, 287007001, 287008006, 15691241000119101, 15691321000119101, 15691881000119107, 16050071000119108, 16606721000119107, 318861000119101, 318931000119100, 9631008, 786077003, 162930007, 239805001, 239806000, 239810002, 239811003, 76481000119102, 783724009, 67531005, 429466000, 1197212001, 1162826002, 26568002, 30620003, 32232003, 81526008, 82058009, 93557001, 4061000119104, 17761000119109, 17771000119103, 103771000119101, 445308004, 444860006, 445307009, 732927000, 735749005, 763889002, 768939009, 203934001, 203935000, 203936004, 203941007, 203942000, 203943005, 203944004, 203946002, 203948001, 203949009, 203950009, 203951008, 203954000, 203955004, 203957007, 203974007, 203975008, 203976009, 203985009, 203986005, 203987001, 203990007, 203991006, 203992004, 203994003, 203996001, 203997005, 203998000, 204003007, 204004001, 204005000, 204006004, 204008003, 204010001, 204011002, 204012009, 204013004, 253111001, 253113003, 253114009, 253115005, 253117002, 253118007, 253119004, 253120005, 38116000, 40130009, 53318002, 58557008, 61819007, 70534000, 76916001, 77224008, 414667000, 425687007, 427216002, 268143001, 268146009, 1003445004, 1003448002, 1010643006, 1010644000, 1052326006, 890422008, 894125007, 192974004, 267591005, 1734006, 1156538002, 9341001, 90584004, 446835008, 698500002, 212355006, 212357003, 230616000, 50448004, 405753002, 405754008, 405759003, 262521009, 262715008, 262716009, 262738006, 34011000087104, 1187144005, 1162281004, 1162290006, 1162289002, 1162283001, 1162329002, 1162310008, 496002, 3217005, 3446000, 4260009, 8183009, 8303001, 8459005, 8758001, 8840000, 8939001, 9048001, 9966008, 10310006, 10575009, 10771002, 11008006, 11413003, 11782000, 11807002, 12462000, 12519004, 13456001, 13463001, 13498003, 14363008, 15701003, 15724005, 16695002, 17214002, 17463000, 18960007, 20132009, 20377001, 21573009, 22848002, 22973003, 23283004, 24392008, 25331007, 26037005, 26738009, 28425007, 28753006, 31033002, 31062001, 31882001, 33132004, 33147008, 80980003, 81442004, 81501004, 81642009, 82419009, 83635002, 83716008, 83982007, 83991006, 85262003, 87804006, 87900003, 88405003, 88743001, 89130002, 89825003, 90631008, 110150000, 111232005, 111465000, 111622009, 111633007, 111648001, 125606003, 125607007, 125608002, 125609005, 125871005, 125872003, 129161004, 129162006, 129163001, 129164007, 129165008, 129166009, 1145242001, 1145243006, 1148990002, 146251000146109, 860614004, 866087009, 870364001, 811000119108, 821000119101, 5291000124100, 5301000124104, 36241000119108, 292971000119105, 430681000124108, 430691000124106, 430701000124106, 433721000124109, 433731000124107, 673581000119104, 10642191000119108, 10642311000119101, 10938941000119103, 11310521000119109, 11310561000119104, 11310601000119104, 11310641000119102, 11310841000119101, 11310881000119106, 11310921000119104, 11310961000119109, 11311001000119101, 11311041000119104, 11311081000119109, 11311121000119106, 11311161000119101, 11311361000119103, 11311401000119107, 11311441000119109, 11311481000119104, 11311521000119104, 11311561000119109, 11311601000119109, 11311641000119106, 11311681000119101, 11311721000119107, 11311761000119102, 11311801000119105, 11311841000119107, 11311881000119102, 11311921000119109, 11311961000119104, 11312001000119108, 11312041000119105, 11312081000119100, 11312121000119103, 11312161000119108, 11312201000119103, 18960001000004105, 21920001000004109,

28490001000004108, 29110001000004106, 29120001000004103, 29580001000004102, 31580001000004106, 13194006, 14493003, 21978005, 27937002, 31235003, 41455002, 43405004, 49497005, 53868003, 123534001, 123535000, 445423005, 445734009, 445779008, 446644006, 447464004, 447575003, 448788000, 473435002, 473436001, 473438000, 608829007, 697975004, 698077007, 698404009, 698499006, 698783004, 699332009, 699980004, 721342001, 721343006, 721344000, 721345004, 721346003, 721347007, 721348002, 721349005, 721350005, 721351009, 721352002, 721353007, 721354001, 721355000, 721356004, 721357008, 721358003, 721359006, 721380000, 721360001, 721361002, 721362009, 721363004, 721364005, 721365006, 721366007, 721367003, 721368008, 721369000, 721370004, 721371000, 721372007, 721373002, 721374008, 721375009, 721376005, 721377001, 721378006, 721379003, 721381001, 721382008, 721383003, 721384009, 721385005, 721386006, 721387002, 721388007, 721389004, 721390008, 721391007, 721392000, 721393005, 721394004, 721395003, 721396002, 721397006, 721415001, 721416000, 721417009, 721418004, 721419007, 721420001, 721421002, 721422009, 721423004, 721424005, 721425006, 721426007, 721428008, 721429000, 721430005, 721431009, 721432002, 721433007, 721434001, 721435000, 721436004, 721437008, 721438003, 721439006, 721440008, 721441007, 721442000, 721443005, 721444004, 721445003, 721446002, 721447006, 721448001, 721449009, 721450009, 721451008, 721452001, 721453006, 721454000, 721455004, 721456003, 721457007, 721458002, 721459005, 721460000, 721461001, 721462008, 721463003, 721464009, 721465005, 721466006, 721467002, 721468007, 721469004, 721470003, 721471004, 721472006, 721473001, 721474007, 721475008, 721476009, 721477000, 721478005, 721479002, 721480004, 721494003, 721495002, 721496001, 721497005, 721498000, 721499008, 721500004, 721501000, 721502007, 721503002, 721504008, 721505009, 721506005, 721507001, 721508006, 721509003, 721510008, 721511007, 721512000, 721513005, 721514004, 721515003, 721516002, 721517006, 721518001, 721519009, 721520003, 721521004, 721522006, 721529002, 722641001, 722653002, 722654008, 722900008, 722901007, 722985001, 722986000, 722994007, 722995008, 723144001, 723145000, 723155001, 724892008, 724901008, 733196009, 733197000, 735743006, 737233008, 737234002, 737236000, 737237009, 737238004, 737239007, 737260001, 737263004, 739665005, 739666006, 763869003, 766775007, 767262002, 770094004, 193120004, 195373009, 202745004, 202749005, 202753007, 202831001, 203450003, 203451004, 203452006, 203630007, 206221000, 206223002, 206224008, 207884008, 207895002, 207896001, 207897005, 207898000, 207899008, 207900003, 207901004, 207902006, 207903001, 207904007, 207905008, 207906009, 207907000, 207908005, 207909002, 207910007, 207921003, 207922005, 207923000, 207924006, 207925007, 207926008, 207927004, 207928009, 207929001, 207930006, 207931005, 207932003, 207933008, 207934002, 207935001, 207936000, 207938004, 207939007, 207940009, 207941008, 207942001, 207943006, 207944000, 207945004, 207949005, 207950005, 207951009, 207952002, 207953007, 207954001, 207955000, 207956004, 207957008, 207958003, 207959006, 207960001, 207961002, 207962009, 207963004, 207964005, 207965006, 207966007, 207967003, 207968008, 207969000, 207970004, 207971000, 207972007, 207974008, 207975009, 207976005, 207977001, 207978006, 207979003, 207983003, 207984009, 207985005, 207998001, 208000003, 208001004, 208003001, 208006009, 208007000, 208009002, 208012004, 208014003, 208017005, 208020002, 208022005, 208023000, 208028009, 208029001, 208031005, 208034002, 208035001, 208037009, 208042001, 208043006, 208045004, 208048002, 208049005, 208051009, 208056004, 208057008, 208058003, 208059006, 208060001, 208063004, 208064005, 208065006, 208066007, 208067003, 208071000, 208078006, 208198004, 208206004, 208215006, 209048009, 209049001, 209050001, 209051002, 209054005, 209064001, 209066004, 209067008, 209068003, 209069006, 209071006, 209077005, 209078000, 209079008, 209080006, 209082003, 209083008, 209084002, 209085001, 209090003, 209092006, 209093001, 209094007, 209095008, 209097000, 209098005, 209100005, 209101009, 209102002, 209112009, 209145009, 209146005, 209147001, 209148006, 209151004, 209153001, 209154007, 209155008, 209156009, 209157000, 209158005, 209159002, 209160007, 209162004, 209163009, 209164003, 209165002, 209166001, 209173006, 209174000, 209175004, 209176003, 209178002, 209179005, 209180008, 209181007, 209182000, 209185003, 209186002, 209188001, 209189009, 209190000, 209191001, 209193003, 307137009, 307211004, 308145001,

311806008, 311814002, 311815001, 311848001, 311862008, 311864009, 311890007, 311891006, 312786001, 312787005, 312788000, 312792007, 312837005, 312838000, 312839008, 313304007, 314667001, 314668006, 371561008, 371562001, 372477008, 209194009, 209195005, 209196006, 209197002, 209206007, 209208008, 209548004, 209549007, 209550007, 209553009, 209559008, 209560003, 209561004, 209565008, 209719007, 209720001, 209721002, 209723004, 209725006, 209729000, 209731009, 209818005, 210512002, 211492005, 212169004, 212171004, 212172006, 212173001, 212174007, 212177000, 212178005, 212179002, 212180004, 212183002, 212185009, 212186005, 212187001, 212188006, 212192004, 212193009, 212194003, 212197005, 212198000, 212199008, 212200006, 212201005, 212202003, 212203008, 212205001, 212206000, 212207009, 212208004, 212209007, 212210002, 212211003, 212212005, 212213000, 212214006, 212215007, 212216008, 212217004, 212218009, 212219001, 212220007, 212221006, 212222004, 212223009, 212224003, 212225002, 212226001, 212227005, 212356007, 212452005, 212453000, 212454006, 212455007, 213350006, 213361003, 213383004, 230579006, 230614002, 230741004, 234007007, 240198002, 240212007, 241992009, 34996005, 35403005, 38170003, 39058009, 40047007, 41078002, 41133002, 42157000, 42942008, 44237008, 44264009, 44434003, 44701004, 45387006, 46549005, 46931002, 46995009, 47518006, 48956000, 49747005, 49815007, 49891002, 50087006, 50888000, 51258005, 51514006, 52289006, 52622006, 53810004, 54317001, 54957006, 58919004, 61386002, 62297002, 63081009, 63819001, 63903007, 64306000, 64469005, 64728002, 66020004, 66158003, 68577004, 69593000, 72346008, 72513001, 74177004, 74297002, 74360009, 76435008, 78085003, 78157002, 78211006, 78396000, 78687007, 79125007, 79591004, 405561009, 405755009, 405756005, 405757001, 405758006, 405760008, 405761007, 405764004, 405765003, 426646004, 427027005, 427571000, 427887006, 428144009, 429665006, 430948002, 430962009, 431947002, 432249006, 441881007, 442544003, 262522002, 262523007, 262524001, 262696004, 262697008, 262698003, 262699006, 262700007, 262705002, 262706001, 262707005, 262708000, 262709008, 262710003, 262711004, 262712006, 262713001, 262714007, 262718005, 262721007, 262723005, 262724004, 262725003, 262726002, 262727006, 262728001, 262729009, 262730004, 262731000, 262732007, 262733002, 262734008, 262735009, 262736005, 262737001, 262740001, 262741002, 262742009, 262743004, 262744005, 262745006, 262746007, 262747003, 262748008, 262944000, 262975009, 262995003, 263068000, 263069008, 263070009, 263071008, 263072001, 263178004, 263179007, 263180005, 269061001, 269062008, 269063003, 269064009, 269065005, 269066006, 269067002, 269068007, 269069004, 269070003, 269071004, 269072006, 269073001, 269074007, 269075008, 269076009, 269077000, 269122007, 269123002, 269124008, 269125009, 269126005, 269127001, 269128006, 270507001, 270508006, 274146004, 274153008, 274154002, 274155001, 274156000, 274157009, 274158004, 274162005, 277326006, 281598004, 281907005, 281908000, 281909008, 281910003, 281911004, 281912006, 281913001, 281914007, 281915008, 281916009, 281917000, 281918005, 281919002, 281920008, 281921007, 281922000, 281923005, 281924004, 281925003, 281926002, 281927006, 281928001, 281929009, 281930004, 281931000, 281932007, 281933002, 281934008, 281935009, 281936005, 281937001, 281938006, 281940001, 281941002, 282785008, 302661000119103, 302681000119107, 302701000119105, 302711000119108, 723083001, 1231168008, 672371000119103, 672381000119100, 672421000119109, 672481000119108, 672501000119104, 672521000119108, 672541000119102, 672561000119103, 674091000119108, 674111000119100, 674131000119105, 674161000119102, 674361000119104, 674381000119108, 674401000119108, 16703761000119102, 672441000119103, 672461000119104, 31097004, 110270004, 446312004, 240460008, 138371000119104, 446311006, 446957000, 446958005, 447262002, 447378002, 186476008, 186478009, 186479001, 186480003, 64764001, 425561001, 429297003, 770596007, 193206003, 230683002, 257277002, 1179294000, 1208615009, 1222704008, 16415791000119104, 5127009, 26609002, 31839002, 80976008, 82178003, 87842000, 91637004, 1156827007, 787172004, 77461000119109, 77471000119103, 292991000119106, 15729961000119108, 16005191000119106, 16005231000119102, 29145002, 56989000, 58756001, 230299004, 230300007, 230301006, 608874000, 717266001, 722432000, 763352005, 763688008, 771261002, 774153002, 774154008, 782752005, 193207007, 205294008, 363222009, 230669004, 230670003,

230671004, 230672006, 230673001, 230674007, 230675008, 230676009, 230677000, 230678005, 230679002, 230682007, 230684008, 230685009, 230686005, 230687001, 55051001, 63135006, 65242001, 67725009, 414927004, 423753005, 277190002, 16005311000119104, 722060007, 723407009, 73297009, 77097004, 1172688004, 1187251009, 1179295004, 1230273004, 1217226000, 1177122009, 1172694007, 1179297007, 1172703004, 111500006, 111501005, 111503008, 111505001, 111506000, 111508004, 129596006, 129620000, 1156836006, 1156848009, 787037000, 77956009, 78468005, 111502003, 111504002, 240048000, 698846009, 702343002, 702383005, 715317001, 715340002, 715341003, 715429006, 718176005, 718177001, 718178006, 718179003, 718180000, 718572004, 718850008, 719985001, 719986000, 719987009, 719988004, 719989007, 719990003, 720522001, 720523006, 723308003, 725042001, 725043006, 725420009, 725907002, 726107008, 726614009, 726615005, 726616006, 726617002, 726618007, 732929002, 732930007, 732931006, 733489002, 763314009, 763718009, 763776004, 763829004, 764812008, 764859001, 764944006, 765197008, 770792007, 771267003, 771272007, 771334000, 773729007, 782675008, 782772000, 783148005, 783166000, 783174004, 783175003, 783176002, 783554002, 784352007, 785298001, 193225000, 193227008, 193230001, 387732009, 240046001, 240047005, 240049008, 240050008, 240051007, 240052000, 240053005, 240054004, 240055003, 240058001, 240060004, 240061000, 240062007, 240063002, 240064008, 240065009, 240067001, 240068006, 240069003, 240070002, 240071003, 240072005, 240073000, 240074006, 240075007, 240076008, 240077004, 240078009, 240104008, 55016009, 58795000, 67747009, 76670001, 399091004, 277373000, 1010712009, 890368007, 890395002, 1418007, 378007, 53974002, 67988000, 1230239009, 8032007, 15788006, 23061007, 92489003, 92490007, 92491006, 92492004, 92844009, 92845005, 92846006, 92847002, 92875000, 93022005, 93038004, 93441003, 93442005, 93443000, 93444006, 95467005, 95468000, 7259005, 7720002, 46041001, 238044004, 715568002, 719041000, 719213009, 722284009, 722434004, 724146008, 724208006, 725104005, 725904009, 733416004, 766821006, 204558002, 205480005, 205481009, 304576008, 306949002, 306950002, 306951003, 306953000, 389264005, 389268008, 389271000, 389272007, 232381002, 253737007, 253742004, 253743009, 42666000, 70690000, 254044004, 254140006, 254142003, 254144002, 268274005, 1003427004, 1003440009, 105501005, 105502003, 105503008, 105504002, 788900007, 931000119107, 60631000119109, 60651000119103, 79021000119104, 79031000119101, 89201000119106, 89241000119108, 152921000119101, 11000731000119102, 11000771000119104, 16320631000119104, 16419651000119103, 444932008, 713655003, 428937001, 428982002, 429075005, 429091008, 429487005, 443143006, 443596009, 898185002, 1208340009, 19133005, 81669005, 92503002, 92824003, 781641005, 715344006, 403815003, 403816002, 403817006, 403819009, 403820003, 403821004, 403822006, 254241004, 1003465006, 1010653007, 880093002, 28634005, 59103002, 68107009, 49049000, 230292008, 1230308005, 1186798007, 1176989007, 1176991004, 1186836002, 4223005, 19972008, 1156822001, 870288002, 870295006, 15782007, 72820004, 230291001, 715345007, 716107009, 716662004, 718685006, 719717006, 722556003, 722557007, 722558002, 722599008, 722965000, 724546003, 724761004, 724762006, 725146001, 766872002, 783012006, 230293003, 230294009, 230295005, 230296006, 38523005, 427488005, 58756001, 76349003, 1186856001, 1167373005, 1172584005, 1217230002, 1197059004, 1208478005, 1230308005, 1220573009, 1208341008, 1177177000, 1186798007, 1186861004, 1187533007, 1172603005, 1176989007, 1176991004, 1186836002, 1187545003, 2992000, 3456001, 4223005, 5822000, 11265003, 15244003, 15802004, 16171003, 16576004, 18842008, 19091006, 19350004, 19903002, 19972008, 22451001, 25297005, 29003001, 29315008, 29618004, 31283000, 32798002, 83465002, 83759003, 89576007, 129608008, 1156764005, 1156822001, 788908000, 788911004, 789657008, 816067005, 838276009, 840501003, 870288002, 870295006, 3331000119108, 135811000119107, 15631251000119109, 15631291000119104, 15631331000119105, 15985791000119101, 15985831000119107, 5158005, 13950001, 15782007, 23772009, 41713005, 45814002, 49049000, 72820004, 80098002, 88125005, 230291001, 230299004, 230300007, 230301006, 312991009, 698279003, 444980006, 445006008, 1230018005, 609218006, 609221008, 699184009, 702314005, 702315006, 702316007, 702317003, 702323008, 702376003, 702377007, 702448007, 712647003,

713956000, 715345007, 715483009, 715534008, 715564000, 715737004, 715768000, 715777007, 715827001, 716107009, 716662004, 716664003, 717054001, 717942003, 718174008, 718576001, 718685006, 719276005, 719278006, 719516000, 719521002, 719717006, 720466001, 721092005, 721228006, 722435003, 722556003, 722557007, 722558002, 722599008, 722763000, 722964001, 722965000, 723992000, 724072002, 724172004, 724383002, 724546003, 724761004, 724762006, 724763001, 724764007, 724765008, 724822003, 724823008, 725146001, 725392005, 733065003, 737582007, 763065008, 763349002, 766872002, 768553002, 768554008, 770722002, 770751003, 770757004, 770939009, 771223000, 771469002, 782695002, 782718007, 782743001, 783005002, 783012006, 783179009, 783242003, 784371009, 163665004, 192859002, 192958009, 307355007, 229692002, 230235007, 230236008, 230292008, 230293003, 230294009, 230295005, 230296006, 230309008, 230310003, 230314007, 230315008, 230316009, 230317000, 230318005, 230319002, 230320008, 230321007, 230322000, 230323005, 230324004, 230325003, 230326002, 230327006, 230328001, 230329009, 230330004, 230332007, 230500006, 230780007, 230781006, 38523005, 41009006, 42369001, 42769004, 43105007, 43647007, 44913001, 48090007, 49386006, 49949003, 52008007, 52274002, 53194005, 55776008, 56509006, 59026006, 60935008, 61200008, 61337004, 62904009, 63576006, 66637005, 66881004, 67305007, 67761004, 68116008, 72585000, 73495003, 74333002, 75019001, 75046006, 76156000, 77375009, 77553008, 80180004, 80381005, 423753005, 425492002, 427232004, 427488005, 427945008, 431034009, 433493000, 439732004, 443352005, 265377002, 278512001, 300993007, 1010642001, 85102008, 91502009, 763597000, 230232005, 1187643003, 1217230002, 1217379007, 1222655009, 1208512000, 1208513005, 1204415006, 1177169004, 1186850007, 5822000, 22443004, 24059009, 80734006, 90791004, 91953003, 91954009, 91955005, 91956006, 129609000, 1156768008, 1156796002, 787174003, 789657008, 827172005, 434311000124104, 10394003, 32680009, 46808003, 91952008, 768663003, 784343003, 444980006, 447351004, 448045004, 699328003, 702354007, 702441001, 702445005, 711158005, 715366004, 715369006, 715371006, 715415005, 715465001, 715483009, 715726000, 715748006, 715751004, 715752006, 715753001, 715754007, 715755008, 715776003, 715824008, 715825009, 715826005, 715984007, 716724006, 717266001, 717332007, 718752007, 718753002, 718754008, 718755009, 718756005, 718769009, 718770005, 718771009, 718772002, 718774001, 718845002, 718849008, 719102004, 719207000, 719208005, 719209002, 719210007, 719249005, 719250005, 719251009, 719252002, 719253007, 719254001, 719255000, 719300001, 719301002, 719302009, 719816006, 719817002, 719818007, 720517001, 720634003, 720750004, 721207002, 721846006, 722064003, 722293005, 723441001, 724065003, 724227000, 724283004, 724765008, 724768005, 724769002, 724770001, 725394006, 725408001, 725433003, 726031001, 733033001, 733065003, 734020000, 734021001, 734023003, 763312008, 763344007, 763348005, 763351003, 763669001, 764095005, 765091006, 766814006, 766818009, 768556005, 770898002, 771469002, 771514002, 773492007, 773495009, 773498006, 773769008, 782690007, 782696001, 782719004, 782721009, 782753000, 783060009, 783203003, 784347002, 784380009, 785300001, 785301002, 785302009, 192871008, 192874000, 193165008, 361272001, 230227009, 230228004, 230229007, 230230002, 230231003, 230233000, 230234006, 230235007, 230236008, 230237004, 230238009, 230239001, 230240004, 230242007, 230243002, 237611007, 253176002, 37960002, 43977004, 45897005, 60146005, 60935008, 67761004, 68116008, 68504005, 73495003, 420932006, 421182009, 421455009, 254092004, 300992002, 1208516002, 1208339007, 5262007, 85672005, 37340000, 1172588008, 1201947005, 1201961000, 1208615009, 1197523001, 1197524007, 1172689007, 7379000, 31097004, 1204350002, 1204351003, 81211007, 84590007, 85505000, 86044005, 88923002, 95647008, 128212001, 838276009, 866051002, 64383006, 699866005, 703524005, 711406009, 711483003, 715565004, 717964007, 718555006, 719836007, 722987009, 723612001, 763067000, 763533003, 766764008, 770430000, 770630005, 770727008, 771081007, 771238004, 771302009, 771475006, 772129007, 783618006, 784341001, 784391002, 785809005, 305719002, 1201950008, 1204334005, 1204349002, 230246005, 230247001, 230248006, 230249003, 230250003, 230251004, 230252006, 230253001, 230254007, 230255008, 230257000, 230258005, 230264003, 230274000, 230784003, 249892007, 44395000, 46251005, 49793008, 54280009, 54304004,

398432008, 1201863001, 1208412003, 60576007, 15241006, 22722001, 1172838005, 5112009, 8627007, 20725005, 21615003, 24225004, 26609002, 29159009, 33209009, 84438001, 86489003, 88219000, 109917000, 109918005, 109919002, 126979000, 128123007, 128205005, 128206006, 129618003, 1144894004, 789430001, 789431002, 838319005, 838320004, 838321000, 838322007, 838330008, 838347009, 838348004, 860807003, 860809000, 860810005, 860811009, 860812002, 860813007, 860814001, 860883001, 866068003, 9631000119102, 290391000119106, 350471000119102, 434761000124100, 14070001000004105, 15729961000119108, 16005191000119106, 16005231000119102, 16026591000119103, 16026631000119103, 204739008, 204740005, 204741009, 271730003, 360434004, 399040002, 703535000, 444024002, 444197004, 472697007, 472698002, 699190008, 702363009, 712882000, 712883005, 713702000, 713703005, 713704004, 715952000, 717822006, 717825008, 717826009, 717827000, 719833004, 719838008, 719972004, 721221000, 721222007, 721223002, 721536001, 721537005, 721608001, 721609009, 722377004, 722518004, 722853000, 722854006, 722996009, 722997000, 722998005, 723147008, 723183004, 723940009, 724806004, 724811002, 724812009, 724813004, 724814005, 724815006, 724816007, 725097006, 735751009, 735752002, 735753007, 737199001, 737240009, 737241008, 770626007, 783198006, 783550006, 783737007, 785723001, 164018003, 188321006, 191952007, 191953002, 191962000, 191972002, 191973007, 192915005, 192918007, 204745000, 371109001, 386537007, 212239002, 212242008, 212243003, 212244009, 230297002, 230537000, 230540000, 230553002, 230556005, 230557001, 230618004, 230655004, 230656003, 230657007, 230658002, 230659005, 230660000, 230661001, 230664009, 230665005, 230666006, 230667002, 231517009, 231518004, 38205001, 40620001, 45294007, 45299002, 46091002, 50330009, 50620007, 51723007, 51881002, 52723009, 61830005, 62985007, 64370005, 64518008, 72585000, 79366000, 397734008, 398148000, 409661008, 425892007, 430042004, 268650001, 277879009, 282746001, 282747005, 282748000, 282792003, 282793008, 282794002, 282795001, 282796000, 282797009, 282799007, 282800006, 282801005, 282803008, 368141000119106, 679871000119102, 679881000119104, 679891000119101, 890118006, 6118003, 25044007, 1186728004, 1179508005, 6807001, 22255007, 24700007, 83942000, 90099008, 816984002, 2701000124105, 182961000119101, 432451000124103, 434321000124107, 16058431000119104, 16415361000119105, 49692006, 230380005, 420554003, 420718004, 733193001, 445967004, 447351004, 703537008, 703621006, 703622004, 713487008, 713543002, 724565006, 724778008, 724780002, 724781003, 724782005, 724783000, 724784006, 724991000, 733028000, 762352004, 766246000, 192926004, 192927008, 192929006, 308634000, 359686005, 386766007, 230372003, 230377009, 230379007, 230564004, 72986009, 425500002, 426373005, 428700003, 438511000, 439567002, 84757009, 1172593006, 1187042007, 1163529009, 1163527006, 1197587003, 1179359005, 1208939001, 1228857005, 1222659003, 1231282002, 1187278006, 1179282009, 1167371007, 2198002, 241006, 6204001, 7033004, 13973009, 15523002, 19598007, 28055006, 95208000, 109478007, 111498005, 788417006, 860804005, 860806007, 860815000, 3371000119106, 5321000124109, 8291000119107, 14401000119109, 20121000119105, 21391000119102, 47391000119107, 82381000119103, 84161000119100, 84171000119106, 84181000119109, 84191000119107, 84201000119105, 84211000119108, 84221000119101, 84231000119103, 100941000119100, 119001000119108, 187931000119106, 290671000119100, 290681000119102, 290691000119104, 290711000119101, 290721000119108, 290731000119106, 290741000119102, 290761000119103, 290871000119101, 290881000119103, 291311000119108, 329991000119104, 430811000124100, 430821000124108, 431071000124107, 431081000124105, 431091000124108, 431101000124102, 431111000124104, 431131000124105, 431141000124100, 431151000124103, 431161000124101, 431461000124109, 431471000124102, 431961000124104, 431971000124106, 431991000124107, 432031000124101, 432151000124107, 432161000124109, 432171000124102, 432291000124105, 432461000124101, 432471000124108, 432541000124104, 434201000124105, 434211000124108, 434291000124103, 434491000124104, 434501000124107, 434511000124105, 434541000124109, 434551000124106, 435321000124106, 435331000124109, 435341000124104, 435361000124100, 436011000124108, 438411000124101, 438421000124109, 440211000124100, 442151000124108, 442161000124105, 444441000124109, 444451000124106, 10750951000119106, 15985351000119105, 322112361000132104, 44145005, 230387008, 230388003, 230438007, 431121000124107, 445355009,

698021005, 698760002, 698761003, 698762005, 698763000, 698764006, 698767004, 699688008, 702326000, 703524005, 710046001, 715534008, 715629001, 716278005, 716706009, 717223008, 717225001, 720519003, 721088003, 722386009, 723125008, 723676007, 724274009, 724549005, 724576005, 724785007, 724786008, 724787004, 724788009, 724789001, 724988000, 724989008, 724990004, 724991000, 724992007, 725163002, 725413002, 726702005, 733031004, 733032006, 733082001, 733195008, 733623005, 734434007, 763349002, 763534009, 763622006, 763632004, 763802009, 763827002, 764453009, 764522009, 765093009, 765170001, 765216006, 766044005, 766815007, 767254005, 768666006, 770405003, 770431001, 770438007, 770622009, 770623004, 770624005, 770643005, 770758009, 770898002, 771141002, 771142009, 771223000, 771448004, 771469002, 773421009, 773497001, 773498006, 773548008, 773643006, 778001003, 778047006, 778063003, 780827006, 782737003, 782772000, 782825008, 783055005, 783062001, 783064000, 783139000, 783739005, 784342008, 784372002, 784377008, 785726009, 163594004, 192845009, 192979009, 192990004, 192999003, 193000002, 193002005, 193003000, 193008009, 193009001, 193010006, 193021002, 193022009, 307356008, 307357004, 352818000, 361123003, 361268000, 230191005, 230381009, 230382002, 230383007, 230384001, 230386004, 230389006, 230390002, 230391003, 230392005, 230393000, 230394006, 230395007, 230396008, 230397004, 230398009, 230399001, 230400008, 230401007, 230403005, 230404004, 230405003, 230406002, 230407006, 230408001, 230410004, 230411000, 230412007, 230413002, 230414008, 230415009, 230416005, 230417001, 230418006, 230419003, 230420009, 230421008, 230422001, 230423006, 230425004, 230426003, 230427007, 230428002, 230429005, 230430000, 230435005, 230437002, 230439004, 230440002, 230441003, 230443000, 230444006, 230445007, 230447004, 230448009, 230450001, 230452009, 230453004, 230454005, 230455006, 230456007, 230457003, 230458008, 230459000, 230460005, 237612000, 36803009, 37356005, 38281008, 39194005, 39745004, 40816002, 49776008, 50866000, 65120008, 71831005, 75023009, 79745005, 413101007, 422513000, 422724001, 422873003, 425054007, 425237009, 425349008, 441678004, 442481002, 442512002, 267581004, 267592003, 278510009, 1222656005, 1179360000, 1230376005, 1179547007, 1222662000, 193119005, 22722001, 65017003, 1187621006, 1172684002, 1187566006, 1187643003, 1187620007, 1187618009, 1172634009, 1187565005, 1187564009, 1187567002, 1187619001, 1187563003, 1220595008, 1222646006, 1222704008, 1187617004, 1187128001, 1172585006, 1172838005, 1172899000, 1177165005, 4183003, 4463009, 5134006, 11442006, 20725005, 26609002, 29159009, 33209009, 86489003, 111499002, 128203003, 128204009, 128205005, 128206006, 1156835005, 1156852009, 860807003, 860809000, 860810005, 860811009, 860812002, 860813007, 860814001, 67661000119100, 350471000119102, 16005191000119106, 16005231000119102, 9133005, 25362006, 41590007, 230552007, 398040009, 398187000, 398229007, 702439002, 715645004, 715665006, 715666007, 715795005, 715796006, 715797002, 715798007, 715799004, 715800000, 715801001, 715802008, 715803003, 717008005, 717010007, 717011006, 717012004, 717013009, 717014003, 717016001, 717825008, 717826009, 717827000, 719510006, 719511005, 719512003, 719513008, 719514002, 719515001, 719833004, 719838008, 719979008, 719980006, 719981005, 720637005, 720638000, 720852000, 722294004, 723497003, 724769002, 725047007, 725048002, 733071009, 763135001, 763136000, 763345008, 763347000, 763400005, 763455008, 763457000, 763458005, 763460007, 763798008, 764730007, 764850002, 764854006, 765046002, 765047006, 765744006, 765745007, 765746008, 765747004, 766977007, 770759001, 771143004, 771144005, 771261002, 771307003, 773308001, 773330000, 773393001, 773414009, 774147002, 778003000, 782742006, 782826009, 782829002, 782881002, 783198006, 783550006, 785723001, 192915005, 230537000, 230540000, 230553002, 230556005, 230557001, 230558006, 230559003, 230655004, 230656003, 230657007, 230658002, 40632002, 42295001, 42986003, 45853006, 53857003, 62985007, 397734008, 398100001, 398148000, 402460000, 267601009, 80849007, 74107003, 2041006, 12331003, 86073008, 789187001, 81780002, 719826004, 773645004, 75968004, 267386008, 82304009, 201826000, 201847001, 37895003, 77994009, 396275006, 268054009, 1162336001, 312351000119105, 1162523002, 1162525009, 1162319009, 1162303002, 1162342002, 1162302007, 1162808000, 1162362005, 4168008, 7126001, 8847002, 10196009, 10948005, 11023001, 18834007,

20075001, 22193007, 25343008, 26241001, 26538006, 28736004, 33262002, 33420007, 33952002, 34427002, 86787000, 90448008, 90860001, 91240008, 109668000, 111242007, 123798002, 1148444001, 1148911004, 1148918005, 1149278003, 1149280009, 1153331004, 1153332006, 1153334007, 1153376004, 1153378003, 1162304008, 789000004, 789001000, 789002007, 4211000119108, 107881000119100, 112961000119103, 112971000119109, 112981000119107, 112991000119105, 113001000119106, 113011000119109, 113021000119102, 113031000119104, 113041000119108, 120401000119101, 122691000119106, 137791000119100, 303001000119101, 303041000119104, 303481000119104, 303551000119107, 303881000119102, 303901000119100, 303921000119109, 303941000119103, 303961000119104, 304121000119108, 304261000119109, 304281000119100, 304301000119101, 304321000119105, 304341000119104, 306131000119100, 306151000119106, 306171000119102, 306191000119101, 306211000119100, 306371000119100, 306551000119102, 306571000119106, 306591000119107, 307371000119102, 307391000119101, 307451000119100, 307471000119109, 318641000119101, 318651000119104, 318661000119102, 318671000119108, 318681000119106, 318691000119109, 318701000119109, 318711000119107, 318721000119100, 318731000119102, 321311000119108, 321341000119107, 323161000119107, 323171000119101, 323251000119103, 323261000119101, 323291000119108, 323301000119109, 323311000119107, 323321000119100, 431731000124106, 433711000124101, 1074061000119108, 1074071000119102, 1074081000119104, 1074091000119101, 1074101000119106, 1074111000119109, 1074121000119102, 1074131000119104, 1074141000119108, 1074151000119105, 1074161000119107, 1074171000119101, 1074181000119103, 1074191000119100, 1074201000119102, 1074211000119104, 1074371000119104, 1074381000119101, 1074391000119103, 1074401000119101, 1074411000119103, 1074421000119105, 1074431000119108, 1074441000119104, 1074451000119102, 1074461000119100, 1074471000119106, 1074481000119109, 1074491000119107, 1074501000119100, 1074511000119102, 1074521000119109, 1074531000119107, 1074561000119104, 1074571000119105, 1075001000119108, 1075021000119104, 10624571000119106, 12241631000119102, 12241671000119104, 12367361000119109, 12367411000119102, 14380001000004102, 15636211000119100, 15637471000119109, 15637511000119100, 15637551000119104, 15686041000119102, 15686081000119107, 15686121000119109, 15705201000119109, 15705241000119106, 15705281000119101, 15705361000119101, 15720481000119102, 15722441000119106, 15722521000119101, 15722561000119106, 15722961000119101, 15749161000119108, 16005991000119109, 16008111000119108, 16008191000119104, 16008271000119106, 16010951000119105, 16015391000119105, 16016151000119105, 16016191000119100, 16016511000119102, 16016551000119101, 16016631000119106, 16039071000119104, 16039111000119106, 16039151000119107, 16039191000119102, 16039231000119106, 16039311000119109, 16039391000119100, 16039431000119105, 16039631000119108, 16039791000119101, 16039831000119107, 16040031000119108, 16040071000119106, 16046631000119100, 16046951000119104, 16076051000119104, 16076131000119106, 16583281000119101, 16583321000119106, 16583361000119101, 82300000, 239872002, 239881008, 270505009, 428857003, 303861000119106, 431011000124103, 12367461000119104, 445477009, 445478004, 445496008, 446496001, 450521003, 699261008, 699262001, 699655001, 699942000, 700293006, 700294000, 700295004, 700304006, 700305007, 700307004, 700312003, 700313008, 700314002, 700315001, 700316000, 700317009, 700318004, 700324005, 700325006, 700326007, 700328008, 700331009, 700332002, 700333007, 700335000, 700336004, 703050003, 703051004, 703052006, 703053001, 703054007, 703627005, 712504007, 713482002, 713826008, 713869000, 713911007, 718602007, 721090002, 724604003, 724605002, 726628003, 735598004, 735599007, 735602002, 735603007, 735604001, 735605000, 735606004, 735607008, 735608003, 735609006, 735610001, 735611002, 735914007, 737057000, 785808002, 190828008, 190842000, 193120004, 201625003, 201627006, 201630004, 201631000, 201633002, 201634008, 201637001, 201639003, 201642009, 201643004, 201645006, 201646007, 201819000, 201829007, 201831003, 201834006, 201835007, 201837004, 201849003, 201852006, 201855008, 201857000, 201858005, 201921009, 201924001, 201925000, 201927008, 201929006, 202657000, 202658005, 202659002, 202670009, 202671008, 202672001, 202674000, 202675004, 202676003, 202677007, 202678002, 202679005, 202680008, 202687006, 202688001, 202689009, 202690000, 202692008, 202693003,

202694009, 202695005, 202696006, 202747007, 202751009, 202755000, 202771000, 308143008, 309246000, 310640003, 313257005, 313258000, 313259008, 313294001, 373623009, 387800004, 387802007, 394991004, 225655006, 232439007, 237898004, 238850005, 238867003, 239828000, 239832006, 239833001, 239837000, 239842008, 239849004, 239862000, 239863005, 239865003, 239866002, 239867006, 239868001, 239869009, 239870005, 239871009, 239873007, 239874001, 239876004, 239877008, 239878003, 239880009, 239883006, 239884000, 240211000, 36071006, 36540006, 39838007, 42944009, 43132002, 43829003, 48210000, 50921008, 52334005, 53332000, 60644007, 60937000, 63198006, 67315001, 67437007, 68675004, 68859000, 69195002, 72275000, 73877009, 74885006, 75468006, 77547008, 78675000, 428027008, 428705008, 428768008, 429419004, 429420005, 432733005, 442884002, 442925003, 442928001, 442941002, 442942009, 443001001, 443002008, 443524000, 254779008, 267889007, 267890003, 267970006, 275324008, 281365008, 1010664005, 1010666007, 1010668008, 1055297002, 1055298007, 16039471000119108, 16039511000119104, 1228860003, 16039591000119108, 16039671000119106, 63643000, 5313005, 9771006, 21333004, 82632005, 111222003, 111224002, 296171000119106, 296181000119109, 296191000119107, 296201000119105, 296211000119108, 296221000119101, 297181000119107, 297191000119105, 297201000119108, 297211000119106, 300891000119108, 300901000119107, 305831000119104, 450691000124105, 450701000124105, 1223891000119108, 12238951000119109, 15636251000119104, 15637391000119101, 15637431000119106, 16016991000119102, 16017031000119102, 16017071000119104, 16017151000119107, 16017431000119105, 16017471000119108, 16017591000119108, 16017631000119108, 16018471000119107, 16018511000119103, 733186000, 202078001, 202079009, 202081006, 202082004, 202093002, 202096005, 202099003, 309744003, 313294001, 239721001, 70690000, 77860008, 267969005, 36427004, 1162292003, 310491000119108, 310501000119101, 310511000119103, 310521000119105, 310551000119102, 310561000119100, 310571000119106, 305961000119100, 1162301000, 2304001, 20021007, 23882000, 26538006, 33132004, 34139004, 84857004, 85216006, 91240008, 1145238004, 1145241008, 1145242001, 1145243006, 1153372002, 1153396007, 1162355001, 866081005, 13271000119101, 13291000119100, 22531000119104, 311191000119109, 311211000119105, 311221000119103, 311231000119100, 431731000124106, 433721000124109, 433731000124107, 433741000124102, 448581000124108, 448591000124106, 448601000124103, 448701000124105, 29574001, 45181002, 86804002, 445332006, 472979002, 472980004, 608829007, 697975004, 699332009, 699456009, 715057009, 724604003, 735604001, 735605000, 735606004, 735607008, 735608003, 735609006, 735610001, 735611002, 193119005, 202707000, 202708005, 202728009, 202729001, 202730006, 202731005, 202732003, 202733008, 202734002, 202735001, 202736000, 202744000, 202745004, 202746003, 202747007, 202748002, 202749005, 202750005, 202751009, 202752002, 202753007, 202754001, 202755000, 202756004, 202757008, 311804006, 212452005, 212453000, 212454006, 212455007, 240211000, 240212007, 240213002, 240215009, 240216005, 41022003, 41733009, 43132002, 43898003, 44983007, 46960006, 60490008, 60937000, 64787009, 68675004, 69195002, 70218004, 73589001, 75467001, 77547008, 425878001, 428778006, 443700006, 267974002, 274137005, 281371002, 302935008, 128049000, 305121000119107, 310431000119109, 1217225001, 314351000119104, 8281000119109, 310491000119108, 310501000119101, 320981000119109, 320991000119107, 321031000119101, 321091000119102, 1163556006, 496002, 2438005, 3217005, 8303001, 8459005, 8840000, 8939001, 9048001, 10310006, 10771002, 11008006, 11049006, 11782000, 11807002, 13463001, 13498003, 15701003, 20132009, 20377001, 21573009, 24392008, 28753006, 32232003, 33147008, 80980003, 81442004, 81501004, 83561009, 83991006, 85216006, 87804006, 89130002, 91877005, 91879008, 91880006, 92055005, 92489003, 92494003, 92806000, 92836009, 92844009, 92849004, 92890001, 92900005, 92999004, 93239001, 93307005, 93315008, 93372003, 93373008, 93437002, 93441003, 93480006, 93564004, 93573007, 93619007, 93629000, 93751003, 94250008, 94791006, 95282002, 95289006, 95293000, 95410001, 95671001, 111234006, 111465000, 111622009, 111648001, 125606003, 125609005, 126553002, 1145242001, 1156266006, 1156962001, 1156963006, 1156964000, 1156965004, 1156966003, 1156967007, 1156968002, 1156969005, 301991000119105, 830196006, 9971000119105,

19441000119107, 22531000119104, 31481000119106, 36241000119108, 200021000119109, 301111000119103, 302311000119107, 302321000119100, 305111000119100, 310421000119106, 314341000119101, 314631000119101, 321151000119106, 321311000119108, 322111000119104, 430681000124108, 430691000124106, 430701000124106, 448581000124108, 673061000119104, 11311521000119104, 11311561000119109, 11312081000119100, 11312121000119103, 15978711000119106, 15978831000119108, 16426681000119106, 29110001000004106, 29120001000004103, 32100001000004100, 5601008, 21978005, 31235003, 43405004, 53868003, 78141002, 89902003, 203198001, 203327008, 444550009, 444844007, 445018004, 445242006, 445332006, 712524006, 712549005, 713741003, 715462003, 719097002, 721342001, 721343006, 721344000, 721345004, 721346003, 721347007, 721348002, 721349005, 721350005, 721351009, 721352002, 721353007, 721354001, 721355000, 721356004, 721357008, 721358003, 721359006, 721380000, 721360001, 721361002, 721362009, 721363004, 721364005, 721365006, 721366007, 721367003, 721368008, 721369000, 721370004, 721371000, 721372007, 721373002, 721374008, 721375009, 721376005, 721377001, 721378006, 721379003, 721381001, 721382008, 721383003, 721384009, 721385005, 721386006, 721387002, 721388007, 721389004, 721390008, 721391007, 721392000, 721393005, 721394004, 721395003, 721396002, 721397006, 721398001, 721399009, 721400002, 721401003, 721402005, 721403000, 721404006, 721405007, 721406008, 722641001, 722900008, 722901007, 723144001, 723145000, 724604003, 724616009, 725012004, 733196009, 733197000, 735604001, 735605000, 737215002, 737233008, 737234002, 767817009, 770094004, 770408001, 187916000, 187989003, 188925008, 189024004, 201764007, 202657000, 202658005, 202659002, 202661006, 202662004, 202663009, 202664003, 202683005, 202684004, 202685003, 202686002, 202687006, 202729001, 202733008, 202744000, 202745004, 202746003, 202747007, 202757008, 202760001, 202761002, 202762009, 202771000, 202812007, 202813002, 202814008, 202815009, 202830000, 203104007, 203157008, 203193005, 203217000, 203256006, 203263006, 203296008, 203452006, 203927003, 203928008, 203934001, 203941007, 203948001, 203969004, 203974007, 203980004, 203985009, 203990007, 203996001, 204003007, 204010001, 205427006, 205428001, 207884008, 207895002, 207896001, 207897005, 207898000, 207899008, 207900003, 207901004, 207902006, 207903001, 207904007, 207905008, 207906009, 207907000, 207908005, 207909002, 207910007, 207921003, 207922005, 207923000, 207924006, 207925007, 207926008, 207927004, 207928009, 207929001, 207930006, 207931005, 207932003, 207933008, 207934002, 207935001, 207936000, 207983003, 207984009, 207985005, 207998001, 208000003, 208001004, 208003001, 208006009, 208007000, 208009002, 208012004, 208014003, 208017005, 208020002, 208022005, 208023000, 209037002, 209046008, 209048009, 209049001, 209050001, 209051002, 209054005, 209064001, 209066004, 209067008, 209068003, 209069006, 209071006, 209083008, 209134006, 209136008, 209137004, 209138009, 209139001, 209140004, 209141000, 209142007, 209143002, 209145009, 209146005, 209147001, 209148006, 209149003, 209151004, 209153001, 209154007, 209155008, 209156009, 209157000, 209158005, 209159002, 209160007, 209162004, 209163009, 209164003, 209165002, 209166001, 308145001, 308757003, 311806008, 311848001, 312660000, 312786001, 312837005, 314195000, 387800004, 387801000, 388981000, 209559008, 209560003, 209561004, 209729000, 212197005, 212198000, 212199008, 212200006, 212201005, 212202003, 212203008, 212205001, 212356007, 212455007, 240215009, 34996005, 35403005, 40047007, 43132002, 43898003, 44264009, 44701004, 45387006, 46931002, 46995009, 50087006, 50888000, 51514006, 52622006, 54404000, 54957006, 58919004, 60490008, 61386002, 62297002, 64469005, 65260001, 66020004, 66158003, 69195002, 69593000, 72346008, 74297002, 74360009, 75467001, 76435008, 78044008, 78157002, 78396000, 78687007, 79125007, 79665007, 405561009, 405754008, 405755009, 405756005, 405757001, 405758006, 405759003, 405760008, 405761007, 405764004, 405765003, 424463006, 425687007, 425871007, 425878001, 427500003, 427787004, 428799009, 430904007, 430948002, 262522002, 262697008, 262701006, 262706001, 262711004, 262724004, 262729009, 262734008, 262740001, 262745006, 263014005, 263015006, 263040004, 263041000, 263042007, 263069008, 263070009, 263180005, 267970006, 267974002, 269062008, 269063003, 269064009, 269065005, 269066006, 269067002, 269068007, 269069004, 269070003, 269071004, 269072006, 269073001, 269074007, 269075008,

269076009, 269077000, 269115004, 269116003, 269117007, 269118002, 269119005, 269120004, 269121000, 269122007, 269123002, 269124008, 269125009, 269126005, 269127001, 269128006, 269483007, 274153008, 274156000, 279030006, 281908000, 281910003, 281911004, 281912006, 281913001, 281914007, 281915008, 281916009, 281917000, 281918005, 281919002, 281920008, 298157007, 298392006, 298393001, 298394007, 298396009, 890422008, 894125007, 318541000119102, 302051000119106, 302661000119103, 268006001, 77881008, 701003, 1418007, 13907006, 19579005, 31249007, 84062004, 89807005, 92489003, 92490007, 92491006, 92492004, 92844009, 92845005, 92846006, 92847002, 93441003, 93442005, 93443000, 93444006, 109347009, 109348004, 1142092000, 353661000119102, 353671000119108, 354621000119105, 11307481000119106, 11307521000119106, 15681801000119103, 15692081000119109, 15692121000119106, 15692161000119101, 7720002, 29574001, 46041001, 86804002, 203395009, 203418008, 702604009, 710211000, 713038003, 719041000, 719213009, 722434004, 724146008, 725104005, 725904009, 733416004, 735605000, 735608003, 735611002, 766821006, 202142008, 202144009, 203368004, 203382004, 203521008, 205480005, 205481009, 304576008, 367356000, 386032009, 389272007, 240134003, 240177002, 240178007, 240179004, 240180001, 240181002, 240182009, 54615002, 58392004, 60937000, 62100001, 70736000, 79362003, 79534009, 429766001, 430505004, 430506003, 254044004, 254142003, 254144002, 268022000, 268023005, 268024004, 268025003, 268274005, 276480007, 276481006, 287062005, 9560007, 125665001, 39595001, 11812741000119105, 11810341000119108, 11810301000119106, 11812781000119100, 2591000, 4975008, 5450005, 6015002, 10380004, 10959001, 11730002, 18386006, 19180002, 32734003, 33847006, 81428007, 90465004, 111691006, 111693009, 129144002, 11775091000119104, 11775131000119102, 11776451000119109, 11776491000119104, 11776691000119101, 11776731000119108, 11776771000119106, 11776811000119106, 11786801000119101, 11786841000119104, 11787841000119101, 11787881000119106, 11787921000119104, 11787961000119109, 11788001000119106, 11788041000119108, 11788081000119103, 11788121000119101, 11788161000119106, 11790721000119101, 11790761000119106, 11791121000119106, 11791161000119101, 11791521000119104, 11791561000119109, 11829551000119104, 11829591000119109, 11837241000119109, 11851981000119101, 11852021000119105, 11852061000119100, 11852101000119102, 11852141000119100, 11852181000119105, 11853731000119103, 11853771000119100, 11853891000119109, 11853931000119101, 11864641000119105, 11864681000119100, 11864881000119101, 11864921000119108, 699001001, 721333008, 723137005, 735792000, 737235001, 737263004, 211537004, 211544008, 211550003, 211551004, 211552006, 211553001, 211554007, 211555008, 211556009, 211557000, 211558005, 211559002, 211560007, 211561006, 211563009, 211564003, 211565002, 211566001, 211567005, 211569008, 211571008, 211572001, 211573006, 211574000, 211575004, 211576003, 211577007, 211578002, 211579005, 211582000, 211583005, 211584004, 211585003, 211586002, 211587006, 211589009, 211590000, 211591001, 211592008, 211593003, 211595005, 211596006, 211597002, 211598007, 211599004, 211602009, 211604005, 211605006, 211606007, 211607003, 213373003, 213374009, 213375005, 230623004, 236431008, 38128008, 40368002, 40874009, 43422002, 47814001, 48944001, 50001009, 50793006, 56697009, 59038002, 60464005, 64417000, 65504006, 65896005, 67070005, 68565009, 69271009, 70000009, 71458007, 72739001, 73825003, 74247001, 74270009, 74682007, 78055008, 78198009, 78508009, 79555006, 397182009, 402181006, 428199000, 262576002, 262928009, 262932003, 262970004, 262989004, 263146007, 269224001, 269225000, 275420003, 283851005, 283852003, 283853008, 283854002, 283855001, 283856000, 283857009, 283858004, 283859007, 283860002, 283861003, 283862005, 283863000, 283979000, 283980002, 283981003, 284571002, 284572009, 286548001, 17230005, 397117006, 400136002, 323051000119106, 323111000119109, 15741561000119104, 15713601000119108, 15713361000119102, 15741641000119102, 490831000124100, 510831000124107, 490841000124105, 1539003, 5321004, 6654000, 8215004, 9896002, 11228000, 12067001, 12663001, 13995008, 16123003, 16579006, 16979000, 23425005, 23574008, 26475003, 26517000, 29066007, 29087002, 33260005, 83450003, 86380000, 86900005, 87562003, 88312006, 90374001, 90521005, 110470001, 111259002, 111261006, 111263009, 111271008, 125583000, 125584006, 107527100011910 8,

1075281000119106, 1075291000119109, 1075301000119105, 1156014009, 1156015005, 15713321000119107, 15713521000119103, 15713881000119101, 15714081000119105, 15714121000119107, 15714161000119102, 15741681000119107, 15742161000119100, 300351000119107, 300391000119102, 323041000119109, 323061000119108, 323081000119104, 323101000119106, 323121000119102, 323141000119108, 787159000, 787161009, 787213005, 787399008, 787400001, 788625001, 551000119107, 99701000119102, 295041000119108, 295051000119105, 296041000119103, 300261000119100, 300331000119101, 300361000119109, 300371000119103, 300401000119100, 303071000119106, 303081000119109, 304721000119109, 304731000119107, 304741000119103, 304751000119101, 304761000119104, 304771000119105, 308101000119108, 308111000119106, 313521000119101, 313531000119103, 320661000119106, 320671000119100, 322901000119100, 322961000119104, 323401000119101, 323411000119103, 323431000119108, 323441000119104, 323481000119109, 323491000119107, 323501000119100, 323511000119102, 323521000119109, 323531000119107, 446311000124106, 446321000124103, 446331000124100, 446341000124105, 446351000124107, 451951000124100, 451961000124103, 452351000124109, 452361000124106, 452391000124103, 452401000124101, 452411000124103, 452421000124106, 452431000124109, 452441000124104, 452511000124102, 1075091000119102, 1075101000119107, 1075121000119103, 1075131000119100, 1075141000119109, 1075151000119106, 1075161000119108, 1075171000119102, 1075181000119104, 1075191000119101, 1075201000119103, 1075211000119100, 1075221000119107, 1075231000119105, 1075241000119101, 1075251000119104, 1075261000119102, 10816301000119102, 10818101000119104, 10823451000119107, 10823531000119102, 10823571000119104, 10830741000119109, 10834461000119108, 10934521000119109, 10934561000119104, 10934601000119104, 10934641000119102, 10934681000119107, 10934721000119101, 10934761000119106, 10934801000119103, 11874301000119109, 11874581000119108, 12239791000119107, 12239831000119101, 15632171000119101, 15632211000119104, 15636171000119102, 15636291000119109, 15637351000119106, 15638851000119101, 15703281000119100, 15703321000119105, 15703361000119100, 15703401000119109, 15703441000119106, 15713441000119108, 15713561000119108, 15713641000119105, 15713681000119100, 15713721000119106, 15713761000119101, 15713921000119108, 15713961000119103, 15714001000119102, 15714041000119100, 15714201000119107, 15714241000119109, 15714281000119104, 15714321000119109, 15714361000119104, 15714401000119108, 15714441000119105, 15714481000119100, 15714641000119107, 15714681000119102, 15714721000119108, 15741521000119109, 15741601000119104, 15741721000119101, 15741761000119106, 15741841000119101, 15741881000119106, 15741961000119109, 15742041000119103, 15742081000119108, 15742121000119105, 15744921000119101, 15848421000119103, 15934101000119103, 15934141000119101, 15934181000119106, 15934221000119103, 15934261000119108, 15934541000119106, 15934581000119101, 15934621000119101, 15934981000119109, 15935021000119105, 15935061000119100, 16898361000119108, 16898401000119104, 5029005, 16866008, 40658002, 88562000, 210707000, 322891000119104, 322911000119102, 322921000119109, 322951000119101, 322971000119105, 322981000119108, 446271000124106, 15741801000119103, 449375005, 449376006, 449712003, 700195008, 704028006, 704030008, 704093007, 704183003, 704237001, 704238006, 704239003, 712650000, 716276009, 721056006, 721059004, 721060009, 721061008, 723311002, 723312009, 723314005, 723315006, 723689001, 723726002, 723731000, 723840008, 724198005, 724199002, 724200004, 724201000, 724285006, 724377000, 724378005, 724595003, 724596002, 726651003, 732212008, 732213003, 732214009, 732934003, 737242001, 764305006, 773819005, 773821000, 774121006, 774122004, 774123009, 774124003, 180011006, 180015002, 180016001, 180018000, 180030006, 180038004, 180040009, 180086006, 180157006, 203533003, 203557003, 203560005, 203562002, 203564001, 203572004, 203575002, 203576001, 203577005, 203582003, 203583008, 203584002, 203585001, 203586000, 203588004, 203589007, 203590003, 203591004, 203595008, 203596009, 203601000, 203602007, 208443000, 208459000, 208476001, 208492006, 314038003, 359724001, 360388005, 367393006, 371186005, 387684008, 209724005, 209775008, 210586009, 211570009, 212442006, 213041003, 214046003, 239574009, 239575005, 239576006, 239577002, 239578007, 239579004, 239580001, 239581002, 239583004, 239584005, 239585006, 239586007, 239587003, 239588008, 239589000,

239590009, 239591008, 239592001, 239593006, 239594000, 239595004, 239596003, 240229005, 240230000, 240231001, 240232008, 240234009, 240235005, 240236006, 240238007, 240240002, 240241003, 240244006, 240251002, 240252009, 240253004, 240254005, 240255006, 240257003, 34685000, 36710006, 37718006, 38014004, 38162008, 39633000, 40531004, 42251007, 43234007, 44364007, 44480001, 45636002, 45657005, 45673004, 45858002, 46028000, 46101007, 47381006, 49482007, 52012001, 52054003, 53704007, 54583007, 54914001, 58660009, 64298006, 64925008, 65358001, 65362007, 66327008, 67321002, 71526000, 71742005, 73570001, 76020000, 76956004, 78785006, 79599002, 79733001, 396767001, 397163000, 397164006, 397166008, 397167004, 397168009, 397218006, 397219003, 397220009, 403306001, 403308000, 405441000, 405443002, 413429004, 438478002, 442558001, 443025009, 265736004, 265737008, 265738003, 265739006, 268036005, 268038006, 275095009, 281476006, 281478007, 281479004, 281482009, 281484005, 281491008, 281492001, 281493006, 281494000, 281495004, 281756004, 281757008, 285566004, 287644000, 287647007, 288245007, 298049006, 298050006, 302296000, 305591000119103, 305601000119105, 895470004, 895471000, 895473002, 895474008, 895522007, 895523002, 895524008, 895525009, 81423003, 86092005, 86555001, 190905008, 235978006, 707766007, 716088000, 720401009, 725052002, 762269004, 762270003, 762271004, 426705001, 237836003, 1177173001, 1220595008, 1217229007, 1208746001, 1197592001, 1208348002, 1220596009, 1217076005, 1220568003, 1220597000, 1217372003, 1197586007, 1174000008, 1177179002, 1177178005, 1187195007, 1187303004, 1187277001, 1177175008, 2391001, 2593002, 4434006, 7530009, 7903009, 12045002, 13109008, 14870002, 14921002, 17192009, 18200000, 20900005, 21265004, 22567005, 22764001, 24629003, 26015003, 27270004, 28557005, 28681006, 29248006, 29352008, 80660001, 86268005, 89454001, 91187007, 95243004, 111307005, 1153392009, 787171006, 789777007, 133221000119100, 368331000119100, 2355008, 4530000, 7720002, 15069006, 17818006, 33979003, 40354009, 69463008, 82699004, 719172003, 699316006, 699870002, 702400006, 702829000, 703508009, 703523004, 711156009, 715242008, 715428003, 715471007, 715472000, 715482004, 715625007, 715652002, 715672007, 715673002, 715674008, 715862006, 715988005, 716024001, 716090004, 716195006, 716334004, 717264003, 717330004, 717823001, 717887003, 717914000, 717929003, 717930008, 718577005, 718717004, 718764004, 718765003, 719013004, 719046005, 719101006, 719162001, 719165004, 719166003, 719171005, 719201004, 719202006, 719211006, 719212004, 719213009, 719378009, 719395001, 719397009, 719405005, 719466009, 719471002, 719685004, 719688002, 719689005, 719944006, 720573009, 720574003, 720612000, 720826006, 720851007, 720863002, 721072003, 721073008, 721074002, 721089006, 721148005, 721841001, 721843003, 721975004, 722033000, 722108000, 722210007, 722381004, 722433005, 722450007, 722453009, 723404002, 723830005, 723995003, 724097003, 724140002, 724141003, 724179008, 725100001, 725417001, 725461009, 726621009, 726672000, 726734001, 732248005, 733050004, 733066002, 733069009, 733072002, 733092009, 733097003, 733419006, 735643002, 762499005, 762500001, 762501002, 763130006, 763320005, 763616002, 763618001, 763631006, 763830009, 763863002, 763868006, 763885008, 763890006, 763893008, 764460003, 764942005, 764956007, 764959000, 764995008, 765204000, 765471005, 765758008, 765761009, 766717008, 766750008, 766753005, 766817004, 766819001, 766820007, 766821006, 766874001, 768935003, 770564004, 770565003, 770603000, 770909004, 770948004, 771074000, 771077007, 771148008, 771179007, 771182002, 771262009, 771515001, 772085001, 772086000, 773300008, 773302000, 773303005, 773404000, 773419004, 773552008, 773625007, 773665006, 773693005, 773768000, 773769008, 774155009, 782755007, 782821004, 782914000, 782941005, 782951006, 783003009, 783096008, 783098009, 783551005, 783559007, 783789002, 162849005, 205468002, 205473008, 205808005, 304576008, 312214005, 313339007, 313426007, 367460001, 389157002, 389158007, 389161008, 389162001, 389163006, 389165004, 216693007, 234533006, 237614004, 237837007, 237838002, 237839005, 239025003, 36914006, 38196001, 38494008, 39401000, 41483000, 42725006, 45582004, 51371005, 51952004, 57536002, 57917004, 58263000, 58561002, 59708000, 67854007, 71003000, 74257000, 74928006, 76520005, 403765001, 422065006, 429362002, 431038007, 254049009, 254060000, 254061001, 254062008,

254064009, 254065005, 254066006, 254067002, 254068007, 254079002, 254084008, 254085009, 254088006, 254090007, 254101001, 254102008, 254103003, 267410008, 302869005, 1003379004, 1010609002, 874931001, 890233009, 890237005, 890380004, 897570002, 1228886008, 1229882003, 1228844002, 1197589000, 88425004, 1172594000, 1179296003, 1216942009, 1197357008, 1208346003, 1230020008, 1230014007, 1197365006, 1229876001, 1222710008, 1231206001, 1228871002, 1197212001, 1208727002, 1208998007, 1217228004, 1222704008, 1177169004, 1186656004, 1162826002, 1187304005, 1187195007, 1172624000, 1172628002, 1172626003, 1162839003, 1169358003, 1162462009, 1538006, 2438005, 2593002, 4945003, 5102002, 5645008, 5842009, 6537000, 6729006, 7199000, 7611002, 8301004, 9740002, 10007009, 11197005, 11701009, 14447001, 15671007, 16026008, 19109004, 19133005, 19276002, 21086008, 22133005, 22471005, 23024003, 23150001, 23880008, 25129008, 25397008, 26568002, 26595007, 27642008, 29956001, 30023002, 30620003, 31429000, 32232003, 81042008, 81526008, 82058009, 84461004, 84873005, 85641006, 89369001, 92503002, 92904001, 92905000, 93249003, 93557001, 93559003, 95477007, 95478002, 95498007, 95499004, 95502000, 95652003, 109561000, 111337001, 111338006, 111505001, 128124001, 128533009, 1149087003, 1187215002, 1208720000, 789120001, 816068000, 838339009, 838441009, 860951008, 866053004, 4061000119104, 17761000119109, 17771000119103, 23931000119104, 23941000119108, 35111000119109, 103771000119101, 104431000119107, 111641000119102, 127551000119100, 330041000119103, 335331000119104, 340941000119101, 348151000119103, 348601000119109, 430711000124109, 12275031000119106, 15665521000119105, 15863931000119108, 15986111000119101, 15986151000119100, 15986191000119105, 15986231000119101, 15986271000119103, 15986311000119103, 16054391000119102, 16054431000119107, 16054471000119105, 15182000, 19886006, 21634003, 30278004, 32219008, 40354009, 54411001, 60318001, 76880004, 79385002, 80651009, 92824003, 111504002, 128082002, 128083007, 128084001, 204061006, 204062004, 204739008, 204740005, 204741009, 253184003, 253185002, 253186001, 266673001, 360434004, 445308004, 703535000, 781641005, 444860006, 445116003, 445307009, 448227009, 449904002, 449905001, 460880006, 609528003, 609529006, 698765007, 698766008, 698999002, 699812002, 699867001, 700063005, 702433001, 702450004, 702611008, 702628006, 703284009, 703285005, 703286006, 703369003, 703526007, 703536004, 703542000, 715344006, 715369006, 715406003, 715422002, 715436007, 715463008, 715482004, 715533002, 715727009, 715780008, 715817007, 715819005, 715820004, 715821000, 715822007, 715905006, 715951007, 715952000, 715981004, 715990006, 716112005, 716169009, 716198008, 716335003, 716996008, 716997004, 716998009, 716999001, 717003001, 717185008, 717632002, 717822006, 717859007, 717939009, 717943008, 717977003, 718095000, 718556007, 718576001, 718608006, 718605009, 718606005, 718607001, 718609003, 718610008, 718611007, 718719001, 718720007, 718759003, 719021005, 719043002, 719136005, 719139003, 719162001, 719377004, 719378009, 719379001, 719380003, 719394002, 719395001, 719819004, 719824001, 719833004, 719947004, 719972004, 720494009, 720498007, 720518006, 720632004, 720813007, 720815000, 720816004, 720819006, 720825005, 720855003, 720954000, 721008000, 721015008, 721072003, 721221000, 721222007, 721223002, 721229003, 721231007, 721297008, 721608001, 721609009, 721847002, 721862000, 721873007, 721903007, 721974000, 721975004, 722006004, 722027009, 722033000, 722036008, 722055008, 722056009, 722201004, 722213009, 722281001, 722282008, 722380003, 722385008, 722390006, 722432000, 722451006, 722453009, 722455002, 722493007, 722604002, 722853000, 722854006, 722989007, 722992006, 722996009, 723131006, 723183004, 723304001, 723309006, 723404002, 723405001, 723501008, 723612001, 723830005, 724067006, 724091002, 724141003, 724226009, 724999003, 725461009, 726031001, 726083008, 726702005, 726704006, 732926009, 732927000, 733028000, 733031004, 733033001, 733049004, 733068001, 733072002, 733086003, 733091002, 733092009, 733094005, 733417008, 733418003, 733469003, 733472005, 733604003, 733637001, 734017008, 734031008, 735749005, 762295002, 763130006, 763344007, 763350002, 763722004, 763797003, 763798008, 763821001, 763834000, 763837007, 763861000, 763889002, 764732004, 765089003, 765325002, 765331004, 765434008, 765757003, 765758008, 766709000, 766710005, 766753005, 766871009, 766931003,

766934006, 768939009, 770404004, 770564004, 770565003, 770604006, 770625006, 770655004, 770721009, 770725000, 771074000, 771076003, 771142009, 771146007, 771147003, 771148008, 771179007, 771234002, 771303004, 771336003, 771473004, 772224009, 772225005, 773280009, 773305003, 773307006, 773394007, 773497001, 773552008, 773554009, 773610007, 773627004, 773665006, 773672007, 773737004, 774070008, 774150004, 774208009, 775907000, 778005007, 778070003, 782720005, 782737003, 782739000, 782753000, 782754006, 782755007, 782757004, 782825008, 782879004, 782884005, 782945001, 782951006, 783176002, 783701002, 783703004, 784344009, 785298001, 785299009, 785306007, 785307003, 786076007, 172069000, 203927003, 203928008, 203934001, 203935000, 203936004, 203941007, 203942000, 203943005, 203944004, 203946002, 203948001, 203949009, 203950009, 203951008, 203954000, 203955004, 203957007, 203969004, 203974007, 203975008, 203976009, 203980004, 203981000, 203982007, 203985009, 203986005, 203987001, 203990007, 203991006, 203992004, 203994003, 203996001, 203997005, 203998000, 204003007, 204004001, 204005000, 204006004, 204008003, 204010001, 204011002, 204012009, 204013004, 204021005, 204022003, 204036008, 204040004, 204042007, 204043002, 204044008, 204046005, 204047001, 204049003, 204067005, 204068000, 204072001, 204074000, 204081007, 204745000, 205425003, 205749001, 205750001, 307355007, 307359001, 359824007, 360527003, 360530005, 363034005, 371076006, 373587001, 373650004, 373661007, 770560008, 230502003, 230504002, 230506000, 230530003, 230560008, 230562000, 230794008, 233718008, 234139002, 238905009, 247204001, 247209006, 253098009, 253101008, 253103006, 253104000, 253106003, 253107007, 253108002, 253109005, 253111001, 253113003, 253114009, 253115005, 253117002, 253118007, 253119004, 253120005, 253124001, 253125000, 253128003, 253133004, 253135006, 253136007, 253137003, 253138008, 253139000, 253140003, 253143001, 253145008, 253146009, 253147000, 253148005, 253149002, 253150002, 253151003, 253152005, 253153000, 253154006, 253156008, 253158009, 253159001, 253160006, 253166000, 253167009, 253168004, 253169007, 253170008, 253171007, 253172000, 253174004, 253175003, 253178001, 253179009, 253180007, 253181006, 253182004, 253183009, 253187005, 253188000, 253189008, 253190004, 253192007, 253199003, 253203003, 253238001, 253239009, 36010004, 36025004, 38116000, 38353004, 38632003, 39150004, 39574006, 40130009, 41283003, 42376006, 43427008, 44295002, 44621005, 45163000, 46659004, 46829007, 47032000, 50429003, 52330001, 53318002, 53633000, 53776005, 54794009, 55999004, 56155002, 56531003, 57148006, 57917004, 58557008, 59636002, 60505005, 61819007, 62158001, 64370005, 65144005, 65455002, 67531005, 67876003, 69664005, 70534000, 71178008, 73331006, 74012004, 75076004, 75968004, 76916001, 77224008, 77817004, 78071008, 78572006, 78693004, 78784005, 79665007, 403775003, 403815003, 403816002, 403817006, 403819009, 403820003, 403821004, 403822006, 403856000, 410060009, 414063002, 414667000, 415713009, 416792008, 417658006, 422474003, 424728002, 425687007, 427216002, 429466000, 438583008, 442300000, 254132000, 254241004, 254243001, 254255007, 254256008, 268143001, 268146009, 277921008, 277922001, 277949001, 277950001, 281899002, 287080001, 302882002, 1172705006, 1003373003, 1003374009, 1003378007, 1003422005, 1003429001, 1003430006, 1003444000, 1003445004, 1003447007, 1003448002, 1003461002, 1003462009, 1003463004, 1003464005, 1003881009, 1010464002, 1010465001, 1010604007, 1010630006, 1010643006, 1010644000, 1010663004, 1052326006, 879919001, 890118006, 890285006, 890286007, 890287003, 890288008, 890396001, 890422008, 890430009, 890432001, 890433006, 890434000, 894125007, 817968741000119105, 1208338004, 1208935007, 32113001, 34048007, 81793007, 84773003, 367506006, 373413006, 62218008, 63387002, 66510004, 268262006, 1187115008, 1162716000, 1172633003, 16007351000119108, 16007311000119107, 304961000119101, 304971000119107, 1230354001, 1230024004, 1230384009, 1230385005, 1230271002, 16008831000119109, 16009071000119106, 1078201000119106, 1078191000119108, 1208482007, 15672241000119102, 15672201000119104, 16467301000119103, 16467261000119106, 16466741000119102, 16466581000119101, 1197591008, 1230295000, 1163579007, 1172635005, 1187114007, 1187303004, 1187277001, 1187132007, 1163260008, 1239002, 2560006, 2749000, 3699000, 4136000, 5968001, 6688006, 7037003, 7458004, 7921007, 9252005, 9634000, 9989000, 10155006, 13280000,

13624003, 15453007, 16585004, 16603000, 16856000, 16904009, 17268007, 17601009, 18132009, 20136007, 20491006, 20834007, 20948006, 21893008, 22099008, 22199006, 22868008, 23407003, 23568008, 23876003, 27173008, 27774009, 29155003, 29581008, 30526003, 30592006, 32339005, 32455002, 33313004, 33534005, 33754009, 34111000, 81330005, 81539007, 81700000, 82785005, 83015004, 84598000, 84918006, 85280007, 85589009, 86818009, 89409002, 89434002, 89689008, 91846008, 91872004, 91873009, 91878000, 91888004, 91891004, 91898005, 91899002, 91905002, 91906001, 91907005, 91908000, 91913001, 91914007, 92491006, 92497005, 92838005, 92842008, 92843003, 92846006, 92851000, 92854008, 92855009, 92859003, 92860008, 92862000, 92867006, 92868001, 92874001, 92878003, 92879006, 92880009, 92882001, 92892009, 92897003, 92898008, 92902002, 92906004, 92909006, 92910001, 92915006, 92917003, 92920006, 92928004, 92929007, 92940004, 92942007, 92945009, 92950003, 92951004, 92954007, 92956009, 92964003, 92981007, 92982000, 92984004, 92985003, 92986002, 92987006, 92989009, 92991001, 93006009, 93008005, 93017005, 93021003, 93023000, 93096002, 93100001, 93102009, 93104005, 93108008, 93109000, 93115000, 93116004, 93118003, 93122008, 93123003, 93124009, 93126006, 93241000, 93246005, 93248006, 93250003, 93255008, 93256009, 93264003, 93266001, 93269008, 93272001, 93273006, 93286002, 93288001, 93295005, 93296006, 93298007, 93309008, 93311004, 93313001, 93321007, 93322000, 93326002, 93327006, 93331000, 93335009, 93336005, 93346007, 93348008, 93350000, 93356006, 93357002, 93362001, 93364000, 93377009, 93379007, 93380005, 93397004, 93398009, 93399001, 93402000, 93403005, 93404004, 93405003, 93406002, 93408001, 93409009, 93410004, 93411000, 93412007, 93439004, 93443000, 93482003, 93566002, 93570005, 93571009, 93575000, 93577008, 93579006, 93580009, 93584000, 93588002, 93592009, 93593004, 93599000, 93600002, 93602005, 93606008, 93607004, 93608009, 93610006, 93621002, 93626007, 93627003, 93631009, 93633007, 93635000, 95245006, 95284001, 95287008, 95291003, 95299001, 95300009, 95301008, 95465002, 95469008, 109419009, 109422006, 109423001, 111315008, 123557007, 123558002, 123559005, 123560000, 123561001, 123562008, 123563003, 123566006, 123568007, 123569004, 123570003, 123571004, 126762003, 128061008, 128544001, 134219008, 1155640009, 1156184000, 1156185004, 1156186003, 15666721000119108, 15666761000119103, 15666801000119106, 1230357008, 1231289006, 1230356004, 838310009, 838313006, 838441009, 840451006, 840475006, 840477003, 840481003, 840491009, 840512003, 871626002, 2111000119106, 12011000119105, 12121000119102, 112871000119104, 127701000119109, 304931000119109, 305201000119107, 305211000119105, 313271000119103, 323751000119109, 323761000119106, 323771000119100, 450451000124101, 1078131000119109, 1078141000119100, 1078151000119103, 1078161000119101, 1078171000119107, 1078181000119105, 1078251000119105, 1078261000119107, 1078271000119101, 1078291000119100, 1078301000119104, 1078311000119101, 1078321000119108, 1078331000119106, 1078361000119103, 1078371000119109, 1079651000119104, 1079681000119106, 1079691000119109, 1079721000119100, 12235781000119107, 12235821000119102, 12235861000119107, 12235901000119101, 12235941000119104, 12235981000119109, 12236021000119104, 12236061000119109, 15630081000119103, 15630121000119101, 15630161000119106, 15630411000119102, 15637111000119107, 15665921000119100, 15666281000119104, 15666321000119109, 15666481000119100, 15666521000119100, 15666561000119105, 15666601000119105, 15666641000119107, 15666681000119102, 15667041000119103, 15667081000119108, 15667121000119105, 15667201000119105, 15667281000119102, 15667401000119106, 15667441000119108, 15667481000119103, 15667521000119103, 15667561000119108, 15667601000119108, 15667721000119106, 15667761000119101, 15667801000119109, 15667841000119106, 15667881000119101, 15667921000119108, 15667961000119103, 15668001000119101, 15668041000119104, 15668081000119109, 15668161000119101, 15668201000119106, 15668241000119108, 15668281000119103, 15668321000119108, 15668361000119103, 15668441000119109, 15668481000119104, 15668561000119109, 15668601000119109, 15668681000119101, 15668721000119107, 15668761000119102, 15668801000119105, 15668841000119107, 15668921000119109,

15668961000119104, 15669121000119101, 15669161000119106, 15669241000119104, 15669321000119104, 15669521000119108, 15669561000119103, 15669601000119103, 15669641000119101, 15669681000119106, 15669721000119100, 15669761000119105, 15669801000119102, 15669881000119105, 15670161000119108, 15670241000119101, 15670481000119105, 15670521000119105, 15670561000119100, 15670601000119100, 15670881000119107, 15671041000119100, 15671281000119104, 15671321000119109, 15671361000119104, 15671681000119102, 15671721000119108, 15671761000119103, 15671921000119105, 15715361000119103, 15715401000119107, 15864481000119102, 15864521000119102, 15864561000119107, 15864601000119107, 15864641000119109, 15864681000119104, 15916651000119107, 15934341000119103, 15934461000119109, 15934861000119107, 15935701000119103, 15935781000119106, 15973181000119106, 15973221000119103, 15973301000119100, 15973541000119106, 15973621000119101, 16004071000119105, 16004111000119103, 16004151000119102, 16004631000119101, 16004711000119104, 16004751000119103, 16004791000119108, 16004831000119102, 16007591000119106, 16007631000119106, 16009191000119108, 16018591000119107, 16018671000119105, 16018711000119109, 16018751000119105, 16018791000119100, 16026031000119109, 16026151000119104, 16090091000119100, 1967001, 4530000, 19092004, 38371006, 40158001, 42162004, 58010002, 66758006, 72922008, 93300007, 403767009, 724071009, 11873751000119106, 444462006, 445175006, 445216006, 445307009, 449682004, 449683009, 449692007, 449871005, 473394001, 473395000, 699049007, 699256006, 699298009, 699867001, 702313004, 702362004, 702407009, 702425002, 709105005, 715242008, 715440003, 715442006, 715474004, 715523005, 715531000, 715535009, 715704001, 715707008, 715710001, 715723008, 715724002, 715725001, 715861004, 715991005, 716006003, 716091000, 716092007, 716094008, 716099003, 716238003, 716249009, 716636008, 716741008, 717850006, 717914000, 717943008, 718222000, 718679004, 718720007, 719041000, 719138006, 719158007, 719159004, 719162001, 719298001, 719305006, 719685004, 719842006, 719843001, 719907006, 719946008, 719950001, 719951002, 720412009, 720415006, 720456009, 720463009, 720467005, 720498007, 720515009, 720568003, 720572004, 720600004, 720601000, 720602007, 720603002, 720633009, 720752007, 720757001, 720815000, 720818003, 720819006, 720952001, 720953006, 720954000, 721010003, 721013001, 721017000, 721069005, 721075001, 721086004, 721148005, 721221000, 721229003, 721874001, 721882001, 721883006, 721902002, 721970009, 721976003, 721978002, 721979005, 722019000, 722032005, 722211006, 722376008, 722383001, 722452004, 722463001, 723365002, 723442008, 723446006, 723556008, 723580007, 723581006, 723611008, 723888006, 723973002, 724066002, 724170007, 724212000, 724224007, 724617000, 724618005, 724619002, 725097006, 725140007, 725409009, 726032008, 726083008, 726116007, 726117003, 726722009, 726735000, 732250002, 732251003, 732927000, 732928005, 732957009, 733030003, 733032006, 733033001, 733045005, 733067006, 733068001, 733070005, 733087007, 733088002, 733093004, 733094005, 733117001, 733425005, 733454004, 733456002, 733466005, 733606001, 737216001, 737217005, 737219008, 737266007, 737581000, 763066009, 763128009, 763535005, 763624007, 763665007, 763691008, 763742008, 763755009, 763886009, 763889002, 763893008, 764106001, 764437006, 766819001, 766999004, 767000001, 767001002, 767002009, 767003004, 767004005, 767005006, 770409009, 770438007, 770543003, 770544009, 770788000, 771146007, 771177009, 771180005, 771182002, 771261002, 771264005, 771335004, 771338002, 773279006, 773307006, 773416006, 773556006, 773583007, 773733000, 774070008, 774150004, 782332007, 782758009, 782913006, 782937006, 783003009, 783553008, 783562005, 783700001, 783738002, 783774006, 784351000, 786039009, 177504007, 201698009, 203944004, 203951008, 204013004, 205052001, 205057007, 205063003, 205064009, 205065005, 205066006, 205067002, 205068007, 205073001, 205082007, 205083002, 205091006, 205092004, 205093009, 205097005, 205108007, 205109004, 205110009, 205111008, 205122007, 205123002, 205124008, 205125009, 205126005, 205127001, 205129003, 205130008, 205131007, 205132000, 205133005, 205135003, 205139009, 205140006, 205142003, 205143008, 205144002, 205145001, 205146000, 205148004, 205149007, 205161004, 205163001, 205164007, 205171002, 205174005, 205175006, 205176007, 205177003, 205178008, 205179000, 205180002, 205183000, 205184006, 205187004, 205188009, 205189001, 205190005, 205191009, 205203006, 205204000,

205206003, 205207007, 205211001, 205212008, 205213003, 205215005, 205216006, 205217002, 205218007, 205219004, 205221009, 205222002, 205223007, 205225000, 205227008, 205228003, 205230001, 205253000, 205254006, 205255007, 205258009, 205260006, 205261005, 205262003, 205264002, 205265001, 205266000, 205267009, 205269007, 205270008, 205271007, 205273005, 205274004, 205276002, 205277006, 205278001, 205280007, 205285002, 205286001, 205288000, 205291000, 205292007, 205297001, 205299003, 205304002, 205306000, 205308004, 205311003, 205312005, 205313000, 205315007, 205316008, 205322004, 205328000, 205329008, 205330003, 205342008, 205343003, 205346006, 205347002, 205348007, 205349004, 205351000, 205352007, 205353002, 205354008, 205358006, 205361007, 205362000, 205364004, 205365003, 205366002, 205367006, 205368001, 205369009, 205370005, 205371009, 205372002, 205373007, 205378003, 205379006, 205382001, 205385004, 205386003, 205387007, 205395006, 205397003, 205398008, 205399000, 205425003, 205484001, 205557000, 205835001, 310798000, 310800007, 360526007, 361213004, 371080001, 371189003, 371191006, 371197005, 371199008, 371629001, 373427001, 373937002, 389276005, 389277001, 394910004, 395693008, 770561007, 229815001, 229816000, 229845003, 229846002, 229847006, 231250005, 232373003, 239076000, 36172001, 36574005, 37221009, 38859008, 41878001, 41895009, 42808000, 44444001, 45482001, 45615004, 45806008, 46434000, 46829007, 47276000, 47713000, 48008009, 48301005, 48334007, 49226005, 51219000, 51635000, 52022007, 52415006, 52474000, 52781008, 52837007, 53842005, 55379003, 55852007, 59068006, 59070002, 59352006, 59693007, 60108003, 61293001, 61900003, 62964007, 63340009, 63391007, 63711009, 64008009, 65573001, 65976001, 66078008, 66345008, 66783006, 69381005, 70123009, 70410008, 70642007, 70690000, 70794004, 70902004, 71016000, 71084008, 71464000, 71901000, 74245009, 74388003, 74820003, 74928006, 75950002, 76744005, 76842000, 77595004, 77608001, 78018008, 78393008, 79120002, 79168008, 79177001, 79214007, 79969004, 400018004, 400085009, 403281007, 403282000, 403768004, 423095007, 423368008, 427103005, 432293006, 432993002, 439258003, 440391004, 440624006, 441686004, 441944007, 443905007, 253916007, 253917003, 253918008, 253919000, 253920006, 253924002, 253925001, 253926000, 253929007, 253930002, 253932005, 253933000, 253936008, 253939001, 253940004, 253941000, 253942007, 253943002, 253944008, 253945009, 253946005, 253947001, 253955008, 253958005, 253959002, 253961006, 253963009, 253965002, 253966001, 253967005, 253968000, 253969008, 253972001, 254039006, 254051008, 254052001, 254090007, 254137006, 254138001, 254139009, 265798000, 268243008, 268251006, 268264007, 268265008, 270518001, 270963009, 271542005, 278532000, 281372009, 288248009, 297195000, 302297009, 302298004, 302299007, 302955009, 302956005, 302957001, 302958006, 302959003, 1003324007, 1003330007, 1003337005, 1003339008, 1003431005, 1003432003, 1003442001, 1003446003, 1003509004, 1003510009, 1003513006, 1003515004, 1003516003, 1003518002, 1003519005, 1003546000, 1003549007, 1003554003, 1003557005, 1003559008, 1003571002, 1003574005, 1003579000, 1003581003, 1003582005, 1003620005, 1003621009, 1003644007, 1003704006, 1003877009, 1003916008, 1003918009, 1003939002, 1010212005, 1010213000, 1010215007, 1010217004, 1010609002, 1010618000, 1010709006, 15973381000119108, 15973421000119104, 308561000119101, 876847002, 876853002, 890190003, 890191004, 890221004, 890222006, 890223001, 890347006, 890352001, 890353006, 890356003, 890357007, 890359005, 890360000, 890361001, 890364009, 890366006, 890367002, 890373001, 890380004, 890384008, 890397005, 890399008, 890401002, 890403004, 890404005, 890405006, 890409000, 890411009, 890413007, 890416004, 890419006, 890425005, 890429004, 890438002, 890439005, 897523003, 897531008, 897532001, 897533006, 897573000, 897575007, 897577004, 897579001, 897586009, 897588005, 1201777007, 1201776003, 16007391000119103, 1230355000, 1197589000, 1231288003, 16009151000119103, 15670201000119103, 253966001, 304961000119101, 304971000119107, 16467301000119103, 16466741000119102, 1197591008, 1230295000, 1172635005, 1187303004, 1187132007, 4136000, 7037003, 7458004, 9989000, 20491006, 22099008, 23407003, 23568008, 27173008, 29581008, 30592006, 32113001, 81539007, 82785005, 85280007, 89434002, 91872004, 91899002, 91913001, 91914007, 92842008, 92868001, 92878003, 92879006, 92897003,

92929007, 92950003, 92951004, 93109000, 93122008, 93123003, 93246005, 93273006, 93295005, 93296006, 93311004, 93336005, 93356006, 93357002, 93409009, 93410004, 93570005, 93593004, 93606008, 93607004, 93626007, 95287008, 95299001, 95301008, 95465002, 95469008, 109422006, 109423001, 123566006, 123568007, 123569004, 126762003, 128061008, 128544001, 134219008, 1155640009, 1156184000, 1156185004, 1156186003, 838310009, 840451006, 840475006, 840491009, 840512003, 871626002, 12011000119105, 112871000119104, 323751000119109, 323761000119106, 323771000119100, 450451000124101, 1078131000119109, 1078141000119100, 1078151000119103, 1078161000119101, 1078171000119107, 1078181000119105, 1078311000119101, 1078321000119108, 1078331000119106, 12235781000119107, 12235821000119102, 12235861000119107, 12235901000119101, 12235941000119104, 12235981000119109, 15630411000119102, 15637111000119107, 15665921000119100, 15666481000119100, 15666521000119100, 15666561000119105, 15666601000119105, 15666641000119107, 15666681000119102, 15667201000119105, 15667281000119102, 15667401000119106, 15667441000119108, 15667481000119103, 15667961000119103, 15668001000119101, 15668041000119104, 15668081000119109, 15668161000119101, 15668201000119106, 15668241000119108, 15668281000119103, 15668321000119108, 15668361000119103, 15668441000119109, 15668481000119104, 15668561000119109, 15668601000119109, 15668681000119101, 15668721000119107, 15669721000119100, 15669761000119105, 15669801000119102, 15670481000119105, 15671281000119104, 15671321000119109, 15671361000119104, 15671681000119102, 15671721000119108, 15671761000119103, 15864481000119102, 15864521000119102, 15864561000119107, 15864601000119107, 15864641000119109, 15864681000119104, 15916651000119107, 15934861000119107, 40158001, 42162004, 66758006, 11873751000119106, 445175006, 699049007, 702425002, 709105005, 715535009, 717914000, 718679004, 719305006, 720953006, 721075001, 721086004, 721979005, 722032005, 722452004, 722463001, 723581006, 723611008, 723888006, 723973002, 724212000, 725140007, 726117003, 732957009, 733030003, 733033001, 733045005, 733067006, 733070005, 763128009, 766999004, 767000001, 767001002, 770788000, 771180005, 773307006, 773583007, 782758009, 783700001, 783774006, 177504007, 205073001, 205082007, 205083002, 205091006, 205092004, 205097005, 205132000, 205133005, 205144002, 205145001, 205146000, 205148004, 205149007, 205187004, 205188009, 205189001, 205190005, 205191009, 205204000, 205219004, 205221009, 205222002, 205223007, 205225000, 205227008, 205228003, 205230001, 205342008, 205343003, 205346006, 205347002, 205348007, 205349004, 205351000, 205352007, 205353002, 205354008, 205358006, 205361007, 205362000, 205372002, 205378003, 205379006, 310800007, 371191006, 371197005, 395693008, 229845003, 229846002, 229847006, 231250005, 239076000, 37221009, 38859008, 49226005, 53842005, 59070002, 62218008, 66345008, 79969004, 400018004, 400085009, 403282000, 439258003, 253943002, 253944008, 253955008, 253958005, 253961006, 253965002, 253967005, 253968000, 253969008, 254090007, 302297009, 302298004, 302299007, 302958006, 1003330007, 1003337005, 1003509004, 1003519005, 1003571002, 1003582005, 1003644007, 15973381000119108, 15973421000119104, 308561000119101, 876853002, 890190003, 890191004, 890352001, 890353006, 890359005, 890364009, 890367002, 890384008, 890401002, 890429004, 890439005, 897573000, 897575007, 897577004, 897579001, 73573004, 1187359009, 1187358001, 1187292000, 1172688004, 1177173001, 1187115008, 1162716000, 1172633003, 1220595008, 1228858000, 16007351000119108, 16007311000119107, 1197754004, 1217629000, 1217627003, 1217624005, 1217625006, 1208413008, 1209168005, 1202023003, 1208416000, 1201964008, 305121000119107, 1230239009, 1078201000119106, 1078191000119108, 1217229007, 1217373008, 1230098009, 1217210001, 1216940001, 1217225001, 1216943004, 1230273004, 16467301000119103, 16467261000119106, 16466741000119102, 16466581000119101, 16466821000119102, 1197053003, 1208348002, 1222710008, 1231181001, 1231206001, 1197018005, 1197212001, 1220568003, 1208488006, 1217372003, 1197586007, 1217228004, 1197588008, 1201861004, 1208415001, 1202024009, 1208418004, 1229999001, 1201862006, 1208414002, 1202025005, 1208417009, 1163579007, 1162826002, 1172898008, 1177179002, 1179299005, 1177166006, 1169364005, 1187642008, 1179283004, 1169362009, 1169363004, 1172689007, 1187303004, 1187212004, 1187277001,

1172632008, 1177175008, 1187132007, 1162837001, 1172692006, 1163260008, 1163556006, 1187113001, 1231182008, 734001, 841002, 1232006, 1239002, 1418007, 1926006, 2391001, 2438005, 2560006, 2749000, 2884008, 3733009, 4170004, 4749004, 5187006, 5968001, 6688006, 7134007, 7603007, 7727004, 7763004, 7921007, 8032007, 8447006, 8649001, 9147009, 9252005, 9634000, 9723006, 10033001, 10155006, 10623005, 10653003, 11144004, 12252009, 12322003, 13236000, 13280000, 13449007, 14061004, 14870002, 15080006, 15453007, 15552004, 15671007, 15788006, 15843004, 15969009, 16603000, 16856000, 17128000, 17144009, 17190001, 17527002, 17568006, 17601009, 18132009, 18355000, 19441002, 20136007, 20491006, 20834007, 20944008, 21893008, 22006008, 22199006, 22567005, 22660004, 22764001, 22868008, 23061007, 23150001, 23568008, 23876003, 23939000, 24140005, 24297000, 24629003, 24752008, 24897005, 24963004, 25129008, 25642007, 26568002, 27173008, 27262009, 27272007, 27299009, 27774009, 27837003, 28070007, 28557005, 28681006, 28740008, 28861008, 29155003, 29248006, 29271008, 29307005, 29352008, 29581008, 30028006, 30592006, 30620003, 31798004, 32003007, 32232003, 32339005, 32455002, 32958008, 32985001, 33010005, 33313004, 33534005, 33754009, 34513009, 34566007, 1230021007, 1197057002, 80712009, 81526008, 81539007, 82058009, 82077006, 82663009, 82785005, 83015004, 84461004, 84918006, 85280007, 85445006, 85589009, 85697009, 86268005, 86470003, 86610004, 86667008, 86818009, 87074006, 87294007, 88386004, 89434002, 89444000, 89454001, 89497006, 89647000, 89689008, 89859004, 89886004, 89988002, 90145001, 90693008, 91846008, 91847004, 91869006, 91870007, 91871006, 91872004, 91873009, 91877005, 91878000, 91879008, 91880006, 91883008, 91885001, 91887009, 91888004, 91889007, 91891004, 91892006, 91893001, 91894007, 91895008, 91896009, 91897000, 91898005, 91899002, 91900007, 91901006, 91902004, 91903009, 91904003, 91905002, 91906001, 91907005, 91908000, 91909008, 91910003, 91911004, 91913001, 91914007, 91915008, 91916009, 91917000, 91919002, 91920008, 91921007, 92489003, 92490007, 92491006, 92492004, 92494003, 92495002, 92496001, 92497005, 92498000, 92499008, 92500004, 92806000, 92822004, 92834007, 92836009, 92837000, 92838005, 92839002, 92840000, 92841001, 92842008, 92843003, 92844009, 92845005, 92846006, 92847002, 92849004, 92850004, 92851000, 92852007, 92853002, 92854008, 92855009, 92857001, 92859003, 92860008, 92861007, 92862000, 92863005, 92865003, 92866002, 92867006, 92868001, 92869009, 92870005, 92871009, 92872002, 92873007, 92874001, 92875000, 92876004, 92877008, 92878003, 92879006, 92880009, 92881008, 92882001, 92883006, 92884000, 92887007, 92890001, 92891002, 92892009, 92893004, 92895006, 92896007, 92897003, 92898008, 92900005, 92901009, 92902002, 92903007, 92906004, 92908003, 92909006, 92910001, 92912009, 92915006, 92916007, 92917003, 92919000, 92920006, 92922003, 92926000, 92927009, 92928004, 92929007, 92930002, 92935007, 92936008, 92938009, 92939001, 92940004, 92942007, 92943002, 92945009, 92947001, 92948006, 92949003, 92950003, 92951004, 92954007, 92955008, 92956009, 92958005, 92959002, 92964003, 92980008, 92981007, 92982000, 92983005, 92984004, 92985003, 92986002, 92987006, 92988001, 92989009, 92991001, 92992008, 92995005, 92996006, 92998007, 92999004, 93002006, 93003001, 93004007, 93005008, 93006009, 93007000, 93008005, 93009002, 93010007, 93011006, 93014003, 93015002, 93017005, 93019008, 93020002, 93021003, 93022005, 93023000, 93024006, 93025007, 93026008, 93027004, 93028009, 93029001, 93035001, 93036000, 93037009, 93038004, 93070004, 93080000, 93082008, 93083003, 93084009, 93085005, 93086006, 93087002, 93088007, 93089004, 93093005, 93096002, 93098001, 93100001, 93101002, 93102009, 93103004, 93104005, 93105006, 93106007, 93107003, 93108008, 93109000, 93110005, 93111009, 93112002, 93113007, 93114001, 93115000, 93116004, 93117008, 93118003, 93119006, 93120000, 93121001, 93122008, 93123003, 93124009, 93125005, 93126006, 93127002, 93128007, 93131008, 93167001, 93236008, 93239001, 93240004, 93241000, 93242007, 93243002, 93244008, 93246005, 93248006, 93250003, 93253001, 93255008, 93256009, 93258005, 93264003,

93265002, 93266001, 93267005, 93269008, 93270009, 93272001, 93273006, 93274000, 93281007, 93282000, 93284004, 93285003, 93286002, 93288001, 93291001, 93293003, 93294009, 93295005, 93296006, 93298007, 93299004, 93302004, 93303009, 93307005, 93308000, 93309008, 93310003, 93311004, 93313001, 93315008, 93321007, 93322000, 93326002, 93327006, 93331000, 93333002, 93335009, 93336005, 93346007, 93348008, 93349000, 93350000, 93352008, 93356006, 93357002, 93360009, 93362001, 93364000, 93372003, 93373008, 93374002, 93375001, 93377009, 93378004, 93379007, 93380005, 93381009, 93382002, 93383007, 93397004, 93398009, 93399001, 93402000, 93403005, 93404004, 93405003, 93406002, 93407006, 93408001, 93409009, 93410004, 93411000, 93412007, 93413002, 93429005, 93430000, 93437002, 93438007, 93439004, 93440002, 93441003, 93442005, 93443000, 93444006, 93460005, 93480006, 93481005, 93482003, 93483008, 93557001, 93562000, 93564004, 93565003, 93566002, 93567006, 93568001, 93569009, 93570005, 93571009, 93573007, 93574001, 93575000, 93576004, 93577008, 93578003, 93579006, 93580009, 93582001, 93584000, 93585004, 93587007, 93588002, 93589005, 93590001, 93591002, 93592009, 93593004, 93594005, 93595006, 93596007, 93597003, 93598008, 93599000, 93600002, 93601003, 93602005, 93603000, 93604006, 93605007, 93606008, 93607004, 93608009, 93609001, 93610006, 93611005, 93612003, 93615001, 93617009, 93619007, 93620001, 93621002, 93622009, 93624005, 93625006, 93626007, 93627003, 93629000, 93630005, 93631009, 93632002, 93633007, 93634001, 93635000, 95241002, 95243004, 95245006, 95265004, 95282002, 95283007, 95284001, 95285000, 95287008, 95289006, 95290002, 95291003, 95292005, 95293000, 95295007, 95298009, 95299001, 95300009, 95301008, 95302001, 95304000, 95463009, 95464003, 95465002, 95467005, 95468000, 109393007, 109394001, 109395000, 109397008, 109398003, 109399006, 109400004, 109402007, 109403002, 109404008, 109406005, 109407001, 109408006, 109409003, 109410008, 109411007, 109413005, 109414004, 109415003, 109416002, 109417006, 109418001, 109419009, 109420003, 109422006, 109423001, 109513007, 109514001, 109515000, 109517008, 109518003, 109519006, 109520000, 109521001, 109523003, 109524009, 109525005, 109526006, 109527002, 109528007, 109529004, 109714003, 111304003, 111315008, 111501005, 111503008, 111505001, 111508004, 123557007, 123558002, 123559005, 123560000, 123561001, 123562008, 123563003, 123566006, 123568007, 123569004, 123570003, 123571004, 123572006, 126762003, 127328006, 128219005, 128223002, 128224008, 128544001, 129621001, 129622008, 134219008, 1155640009, 1156184000, 1156185004, 1156186003, 1156306006, 15666721000119108, 15666761000119103, 15666801000119106, 787037000, 787407003, 787408008, 788875002, 789777007, 818959006, 838310009, 838313006, 838339009, 840476007, 840481003, 840483000, 871626002, 2111000119106, 4061000119104, 8611000119100, 12121000119102, 17761000119109, 17771000119103, 22301000119102, 23971000119101, 25201000119104, 103771000119101, 104431000119107, 127701000119109, 255581000119100, 305111000119100, 305141000119101, 305201000119107, 305211000119105, 313271000119103, 315271000119104, 318761000119105, 1078131000119109, 1078141000119100, 1078151000119103, 1078161000119101, 1078171000119107, 1078181000119105, 1078361000119103, 1078371000119109, 12235781000119107, 12235821000119102, 15630081000119103, 15630121000119101, 15630161000119106, 15637111000119107, 15664961000119109, 15665001000119108, 15665921000119100, 15666041000119100, 15666081000119105, 15666481000119100, 15666521000119100, 15666561000119105, 15666601000119105, 15666641000119107, 15666681000119102, 15667041000119103, 15667081000119108, 15667121000119105, 15667201000119105, 15667281000119102, 15667401000119106, 15667441000119108, 15667481000119103, 15667521000119103, 15667561000119108, 15667601000119108, 15667721000119106, 15667761000119101, 15667801000119109, 15667841000119106, 15667881000119101, 15667921000119108, 15667961000119103, 15668001000119101, 15668041000119104, 15668081000119109, 15668161000119101, 15668201000119106, 15668241000119108, 15668281000119103, 15668441000119109, 15668481000119104, 15668561000119109, 15668601000119109, 15668681000119101, 15668841000119107, 15668961000119104, 15669041000119108, 15669081000119103, 15669721000119100, 15669761000119105,

15669801000119102, 15669881000119105, 15670161000119108, 15670241000119101, 15670481000119105, 15670521000119105, 15670561000119100, 15670601000119100, 15670881000119107, 15671041000119100, 15671921000119105, 15671961000119100, 15672001000119109, 15715361000119103, 15715401000119107, 15864601000119107, 15864641000119109, 15864681000119104, 15934341000119103, 15934461000119109, 15934861000119107, 15935701000119103, 15935781000119106, 16004071000119105, 16004111000119103, 16004151000119102, 16004631000119101, 16004711000119104, 16004751000119103, 16004791000119108, 16004831000119102, 16018591000119107, 16018671000119105, 1393001, 1967001, 4530000, 5601008, 7720002, 17025000, 17484005, 17818006, 18756002, 19092004, 20766005, 25606004, 29145002, 30652003, 33979003, 35031005, 38371006, 40158001, 41069008, 41656005, 46041001, 48763007, 51409009, 55711009, 58010002, 58112007, 58833000, 62311004, 65764006, 66042003, 66758006, 67202007, 71322004, 72922008, 75049004, 77956009, 82203000, 82236004, 82351006, 82699004, 83470009, 83586000, 86345004, 93132001, 93300007, 93623004, 105986008, 111502003, 111504002, 205418005, 237918004, 238026007, 240048000, 254083002, 254099008, 403767009, 403778001, 422348008, 445308004, 447821002, 719172003, 724071009, 763346009, 444462006, 444550009, 444844007, 444860006, 445116003, 445306000, 445307009, 445468002, 449777000, 449821007, 450808003, 699185005, 699251001, 699313003, 699381006, 699700006, 699867001, 699870002, 702312009, 702339001, 702342007, 702343002, 702346005, 702347001, 702361006, 702382000, 702400006, 702432006, 702443003, 702613006, 703132007, 703523004, 703528008, 703542000, 703544004, 708669006, 709105005, 709490002, 709604005, 711039004, 715242008, 715318006, 715409005, 715419004, 715421009, 715428003, 715429006, 715434005, 715440003, 715462003, 715470008, 715471007, 715472000, 715474004, 715487005, 715505002, 715523005, 715524004, 715531000, 715532007, 715533002, 715568002, 715631005, 715646003, 715652002, 715653007, 715654001, 715670004, 715672007, 715673002, 715674008, 715710001, 715720006, 715721005, 715722003, 715861004, 715862006, 715867000, 715986009, 715991005, 716006003, 716022002, 716023007, 716089008, 716090004, 716092007, 716094008, 716170005, 716189005, 716195006, 716231009, 716232002, 716238003, 716337006, 716696006, 716741008, 716868003, 717221005, 717264003, 717330004, 717771007, 717785002, 717813005, 717814004, 717823001, 717977003, 718099006, 718211004, 718395009, 718556007, 718559000, 718572004, 718576001, 718679004, 718713000, 718720007, 718763005, 718764004, 718765003, 718766002, 719021005, 719041000, 719046005, 719069008, 719096006, 719097002, 719138006, 719162001, 719165004, 719166003, 719171005, 719201004, 719202006, 719203001, 719204007, 719205008, 719213009, 719258003, 719268008, 719298001, 719299009, 719304005, 719378009, 719397009, 719398004, 719404009, 719405005, 719409004, 719466009, 719471002, 719475006, 719685004, 719688002, 719689005, 719800009, 719812008, 719813003, 719815005, 719837003, 719842006, 719843001, 719946008, 719947004, 719948009, 719949001, 719950001, 719951002, 720408003, 720412009, 720414005, 720415006, 720416007, 720418008, 720419000, 720427009, 720429007, 720430002, 720457000, 720467005, 720492008, 720568003, 720598005, 720599002, 720600004, 720601000, 720602007, 720603002, 720606005, 720612000, 720635002, 720640005, 720746006, 720752007, 720753002, 720754008, 720755009, 720757001, 720812002, 720813007, 720815000, 720816004, 720817008, 720818003, 720819006, 720826006, 720851007, 720858001, 720859009, 720860004, 720861000, 720863002, 720952001, 720953006, 720954000, 720955004, 720957007, 720958002, 720984008, 720986005, 721008000, 721009008, 721010003, 721013001, 721014007, 721017000, 721069005, 721072003, 721075001, 721082002, 721086004, 721090002, 721094006, 721095007, 721105004, 721148005, 721223002, 721227001, 721229003, 721296004, 721835008, 721836009, 721845005, 721874001, 721882001, 721883006, 721887007, 721973006, 721974000, 721975004, 722002002, 722003007, 722019000, 722032005, 722051004, 722107005, 722108000, 722109008, 722110003, 722111004, 722113001, 722114007, 722117000, 722201004, 722207000, 722210007, 722211006, 722281001, 722282008, 722283003, 722284009, 722298001, 722383001, 722429003, 722432000, 722433005, 722434004, 722452004, 722456001, 722459008, 722461004, 722463001, 722476007, 722478008, 723308003, 723309006, 723403008, 723404002, 723439002, 723442008, 723556008, 723578001, 723580007, 723610009,

723676007, 723827003, 723991007, 723995003, 723998001, 723999009, 724064004, 724068001, 724069009, 724140002, 724145007, 724146008, 724147004, 724208006, 724212000, 724226009, 724284005, 724349009, 724615008, 724616009, 724617000, 724619002, 725029001, 725030006, 725050005, 725096002, 725097006, 725098001, 725099009, 725100001, 725101002, 725104005, 725141006, 725142004, 725164008, 725165009, 725166005, 725417001, 725420009, 725461009, 725904009, 726029005, 726032008, 726083008, 726116007, 726621009, 726722009, 726724005, 732248005, 732249002, 732250002, 732263008, 732926009, 732927000, 732928005, 732954002, 732955001, 732956000, 732957009, 733030003, 733031004, 733034007, 733038005, 733045005, 733066002, 733067006, 733068001, 733093004, 733095006, 733110004, 733117001, 733118006, 733416004, 733418003, 733419006, 733425005, 733454004, 733456002, 733457006, 733466005, 733468006, 733606001, 733621007, 734173003, 735105001, 735107009, 735749005, 736781007, 736782000, 737215002, 737216001, 737219008, 737581000, 762580003, 762581004, 762582006, 762583001, 762584007, 762585008, 762586009, 762587000, 762588005, 762660007, 763128009, 763134002, 763215008, 763314009, 763315005, 763318007, 763320005, 763353000, 763498007, 763499004, 763535005, 763616002, 763619009, 763658004, 763684005, 763727005, 763728000, 763735008, 763755009, 763774001, 763778003, 763830009, 763863002, 763868006, 763885008, 763886009, 763889002, 763893008, 764437006, 764460003, 764525006, 764697003, 764812008, 764857004, 764942005, 764944006, 764945007, 764956007, 764957003, 764959000, 765197008, 765204000, 765750001, 765761009, 766717008, 766750008, 766819001, 766820007, 766821006, 766870005, 766874001, 766881008, 766884000, 766992008, 767002009, 767003004, 767004005, 767005006, 767326000, 767327009, 768935003, 768939009, 770406002, 770603000, 770625006, 770627003, 770755007, 770786001, 770787005, 770788000, 770909004, 770943008, 770945001, 770946000, 770948004, 771077007, 771146007, 771177009, 771180005, 771185000, 771186004, 771261002, 771264005, 771265006, 771266007, 771267003, 771272007, 771301002, 771338002, 771511005, 773276004, 773278003, 773279006, 773300008, 773302000, 773303005, 773304004, 773306002, 773307006, 773332008, 773404000, 773406003, 773556006, 773557002, 773578004, 773584001, 773586004, 773588003, 773622005, 773625007, 773626008, 773628009, 773667003, 773672007, 773693005, 773702002, 773730002, 773733000, 773773006, 773985008, 774150004, 774155009, 777998000, 778008009, 778022009, 778067002, 778068007, 778069004, 782332007, 782689003, 782758009, 782772000, 782781006, 782782004, 782783009, 782785002, 782786001, 782820003, 782821004, 782882009, 782912001, 782913006, 782914000, 782937006, 782940006, 782941005, 782945001, 782951006, 783003009, 783004003, 783005002, 783097004, 783137003, 783140003, 783159001, 783165001, 783174004, 783175003, 783176002, 783181006, 783204009, 783553008, 783559007, 783620009, 783700001, 783723003, 783774006, 783789002, 784006008, 784010006, 784350004, 784381008, 785298001, 786039009, 193222002, 203923004, 203927003, 203928008, 203934001, 203935000, 203936004, 203941007, 203942000, 203943005, 203944004, 203946002, 203948001, 203949009, 203950009, 203951008, 203954000, 203955004, 203957007, 203969004, 203974007, 203975008, 203976009, 203980004, 203981000, 203982007, 203985009, 203986005, 203987001, 203990007, 203991006, 203992004, 203994003, 203996001, 203997005, 203998000, 204003007, 204004001, 204005000, 204006004, 204008003, 204010001, 204011002, 204012009, 204013004, 204021005, 204022003, 204216001, 204217005, 204558002, 205052001, 205057007, 205063003, 205064009, 205065005, 205066006, 205067002, 205068007, 205082007, 205092004, 205093009, 205101001, 205111008, 205142003, 205143008, 205148004, 205149007, 205171002, 205179000, 205180002, 205183000, 205184006, 205211001, 205212008, 205213003, 205215005, 205216006, 205217002, 205218007, 205219004, 205221009, 205222002, 205223007, 205225000, 205253000, 205254006, 205255007, 205258009, 205260006, 205269007, 205270008, 205271007, 205273005, 205274004, 205276002, 205277006, 205278001, 205280007, 205281006, 205285002, 205286001, 205308004, 205313000, 205317004, 205322004, 205328000, 205329008, 205330003, 205347002, 205348007, 205349004, 205354008, 205361007, 205362000, 205364004, 205365003, 205366002, 205367006, 205368001, 205378003, 205379006, 205395006, 205397003, 205398008, 205399000, 205425003, 205427006, 205428001, 205429009, 205430004, 205431000, 205432007, 205435009, 205436005, 205437001, 205438006, 205439003,

205456006, 205460009, 205465004, 205468002, 205473008, 205480005, 205481009, 205482002, 205483007, 205484001, 205496008, 205497004, 205503007, 205506004, 205532005, 205797000, 205798005, 205812004, 304576008, 306949002, 306950002, 306951003, 306953000, 310800007, 313339007, 327877008, 360507004, 363032009, 363036007, 367489004, 370481004, 371629001, 373427001, 373643003, 373937002, 385482004, 385483009, 387732009, 388981000, 389157002, 389158007, 389159004, 389160009, 389161008, 389162001, 389163006, 389165004, 389166003, 389167007, 389168002, 389169005, 389170006, 389171005, 389191003, 389193000, 389195007, 389199001, 389207000, 389214003, 389216001, 389236000, 389237009, 389239007, 389260001, 389261002, 389262009, 389263004, 389264005, 389268008, 389271000, 389272007, 389273002, 389276005, 389277001, 391982004, 229815001, 229816000, 229847006, 232112009, 232113004, 232373003, 232381002, 234143003, 234988003, 236529001, 236705005, 236796004, 237513002, 237888005, 237889002, 237890006, 238025006, 238027003, 238047006, 238855000, 238861002, 238866007, 240047005, 240049008, 240050008, 240051007, 240052000, 240053005, 240055003, 240058001, 240060004, 240061000, 240062007, 240063002, 240064008, 240071003, 240072005, 240073000, 240074006, 240075007, 240076008, 240078009, 240081004, 240082006, 240083001, 240084007, 240085008, 240086009, 240087000, 240104008, 240156000, 253101008, 253103006, 253104000, 253106003, 253107007, 253108002, 253109005, 253111001, 253113003, 253114009, 253115005, 253117002, 253118007, 253119004, 253120005, 253247009, 253737007, 253742004, 253743009, 253817004, 34643004, 35691006, 36010004, 36172001, 36517007, 36601008, 37281006, 38116000, 38494008, 38776003, 38827001, 38859008, 38998004, 39401000, 39540007, 40130009, 41132007, 41517009, 41895009, 42190000, 42376006, 42432003, 42666000, 42725006, 42808000, 43036001, 43063000, 43152001, 43226001, 43814000, 44444001, 44621005, 44697002, 45482001, 45987002, 46434000, 46698009, 46804001, 47276000, 47713000, 47880003, 48008009, 48069004, 48334007, 48777005, 49226005, 49347007, 49946005, 50108000, 50123005, 50751005, 50847000, 50869007, 51655004, 51780007, 51952004, 52022007, 52330001, 52415006, 52474000, 52616002, 52781008, 52837007, 53318002, 53776005, 53842005, 53974002, 54008006, 54036001, 54954004, 55016009, 55133004, 55379003, 55852007, 55999004, 56692003, 57219006, 57917004, 58557008, 58561002, 59068006, 59070002, 59178007, 59352006, 59399004, 59708000, 59763006, 60108003, 60192008, 60399005, 61172008, 61293001, 61367005, 61819007, 61900003, 62501005, 62628008, 62803002, 62964007, 63122002, 63387002, 63391007, 63413008, 63711009, 63890001, 64404003, 64716005, 65144005, 65455002, 65524005, 65573001, 65976001, 66038001, 66102006, 67531005, 67988000, 68186003, 68359008, 70123009, 70199000, 70410008, 70534000, 70610001, 70690000, 70794004, 70974002, 71178008, 71311003, 71901000, 72239002, 72424001, 72535009, 72855002, 73284007, 74245009, 74388003, 74820003, 74928006, 75072002, 75311005, 75491005, 75654009, 75950002, 76098004, 76520005, 76556008, 76670001, 76744005, 76842000, 76916001, 77097004, 77224008, 77471004, 77542002, 77608001, 78044008, 78314001, 78675000, 79120002, 79168008, 79177001, 79191007, 79214007, 79244005, 79665007, 79969004, 398114001, 398719004, 398958000, 399045007, 399091004, 399249008, 400946004, 400947008, 403409009, 403410004, 403411000, 403768004, 403856000, 405772002, 410060009, 413906003, 414667000, 415713009, 417658006, 423095007, 423368008, 424286001, 425687007, 425871007, 427103005, 427216002, 429466000, 429753001, 431768007, 432993002, 440350001, 440391004, 441686004, 441944007, 443905007, 253916007, 253917003, 253930002, 253940004, 253941000, 253942007, 253943002, 253947001, 253958005, 253961006, 253965002, 253969008, 253975004, 253977007, 253979005, 253980008, 253989009, 254020001, 254021002, 254022009, 254023004, 254025006, 254026007, 254027003, 254033007, 254034001, 254039006, 254040008, 254041007, 254043005, 254044004, 254045003, 254046002, 254047006, 254048001, 254049009, 254050009, 254051008, 254052001, 254054000, 254055004, 254058002, 254059005, 254060000, 254061001, 254062008, 254064009, 254065005, 254066006, 254067002, 254068007, 254069004, 254078005, 254079002, 254080004, 254082007, 254084008, 254085009, 254087001, 254088006, 254090007, 254091006, 254092004, 254093009, 254094003, 254095002, 254096001,

254097005, 254098000, 254100000, 254101001, 254102008, 254103003, 254104009, 254110009, 254111008, 254112001, 254113006, 254114000, 254116003, 254117007, 254120004, 254121000, 254122007, 254123002, 254124008, 254125009, 254129003, 254130008, 254131007, 254132000, 254133005, 254134004, 254135003, 254137006, 254138001, 254139009, 254140006, 254142003, 254144002, 254146000, 254149007, 254150007, 254151006, 254152004, 254153009, 268143001, 268146009, 268239009, 268240006, 268243008, 268262006, 268274005, 268276007, 270518001, 270519009, 271018002, 271542005, 271574003, 277807007, 277949001, 277950001, 278708009, 278710006, 278712003, 278713008, 278715001, 278832007, 278833002, 278834008, 278991002, 279081001, 279082008, 279309008, 280159008, 281372009, 281373004, 287087003, 288248009, 302298004, 302299007, 302957001, 1172705006, 1003324007, 1003379004, 1003384005, 1003427004, 1003431005, 1003432003, 1003440009, 1003442001, 1003445004, 1003446003, 1003448002, 1003505005, 1003513006, 1003515004, 1003516003, 1003518002, 1003519005, 1003520004, 1003546000, 1003549007, 1003554003, 1003557005, 1003558000, 1003559008, 1003563001, 1003566009, 1003568005, 1003571002, 1003573004, 1003576007, 1003577003, 1003578008, 1003581003, 1003582005, 1003704006, 1003858007, 1003860009, 1003862001, 1003877009, 1003916008, 1003918009, 1010212005, 1010213000, 1010215007, 1010217004, 1010609002, 1010643006, 1010644000, 1010664005, 1010666007, 1010668008, 1010685005, 1010709006, 1052326006, 15973381000119108, 15973421000119104, 874931001, 876847002, 876853002, 890190003, 890191004, 890233009, 890237005, 890347006, 890351008, 890352001, 890353006, 890361001, 890367002, 890368007, 890380004, 890395002, 890397005, 890399008, 890404005, 890409000, 890410005, 890411009, 890413007, 890419006, 890422008, 890425005, 890437007, 890438002, 890439005, 894125007, 897523003, 897530009, 897531008, 897532001, 897533006, 897537007, 897560004, 897586009, 897590006, 1229895008, 1222661007, 1228860003, 1208342001, 16007391000119103, 1197589000, 15670201000119103, 1222706005

HCPCS: L5250, L5270, L5331, L6050, L6055, L6200, L6205, L6300, L6310, L6320, L6450, L6550, L6920, L6925, L6940, L6945, L6960, L6965

ICD10PCS: 0X6L0Z0, 0X6L0Z1, 0X6L0Z2, 0X6L0Z3, 0X6M0Z0, 0X6M0Z1, 0X6M0Z2, 0X6M0Z3

#### Intellectual disability

SNOMED CT: 86765009, 110359009, 82371000119101, 698626001, 1187042007, 1172594000, 1186734006, 1173036000, 1179408008, 1187644009, 1169355000, 1187249005, 1208936008, 1217229007, 1222658006, 1217371005, 1217382002, 1197593006, 1208746001, 1220589007, 1230273004, 1217381009, 1222710008, 1217379007, 1208987006, 1220594007, 1208481000, 1222657001, 1220568003, 1220597000, 1197148005, 1208488006, 1208727002, 1197591008, 1208341008, 1217228004, 1197588008, 1229872004, 1208614008, 1177169004, 1187278006, 1179301003, 1172627007, 1186730002, 1172630000, 1186711002, 1186721005, 1187642008, 1187304005, 1172685001, 1179283004, 1187195007, 1167375003, 1187114007, 1187038009, 1172889005, 1173998003, 1187303004, 1172698005, 1172624000, 1172629005, 1179282009, 1187277001, 1177175008, 1187041000, 1172626003, 1187247007, 1187122000, 1172697000, 1186713004, 1172691004, 1186729007, 1187210007, 1177167002, 1172899000, 1167371007, 1162462009, 9527009, 10007009, 31216003, 33982008, 1208344000, 109478007, 1156584007, 1169359006, 1187250005, 1208720000, 787093004, 787171006, 787174003, 787175002, 788417006, 788584007, 816067005, 838441009, 613003, 5619004, 17827007, 21634003, 68618008, 76880004, 79385002, 702356009, 702412005, 702416008, 703389002, 703535000, 722037004, 770907002, 699297004, 699298009, 699316006, 699669001, 702327009, 702344008, 702354007, 702423009, 702441001, 702816000, 703526007, 715409005, 715428003, 715441004, 715628009, 715989002, 716024001, 716089008, 716096005, 716107009, 716112005, 716191002, 716334004,

716706009, 716709002, 716996008, 717157006, 717222003, 717763008, 717822006, 717887003, 717913006, 717945001, 718226002, 718573009, 718577005, 718680001, 718681002, 718766002, 718845002, 718847005, 718848000, 718896000, 718897009, 718900002, 718905007, 718908009, 718909001, 718910006, 718911005, 718912003, 718914002, 719017003, 719009006, 719010001, 719011002, 719012009, 719013004, 719016007, 719018008, 719020006, 719042007, 719046005, 719069008, 719097002, 719101006, 719102004, 719136005, 719138006, 719139003, 719140001, 719155005, 719156006, 719157002, 719160009, 719161008, 719162001, 719202006, 719212004, 719378009, 719380003, 719450007, 719466009, 719583002, 719599008, 719600006, 719800009, 719808002, 719810000, 719811001, 719812008, 719825000, 719826004, 719834005, 719842006, 719909009, 719947004, 720401009, 720468000, 720501007, 720502000, 720517001, 720573009, 720635002, 720639008, 720746006, 720748007, 720825005, 720855003, 720954000, 720955004, 720957007, 720979002, 720981000, 720982007, 721007005, 721008000, 721017000, 721073008, 721086004, 721087008, 721089006, 721146009, 721207002, 721208007, 721224008, 721841001, 721843003, 721875000, 721883006, 721973006, 721974000, 722002002, 722003007, 722031003, 722033000, 722035007, 722055008, 722056009, 722065002, 722075004, 722105002, 722106001, 722107005, 722110003, 722111004, 722209002, 722213009, 722281001, 722282008, 722378009, 722379001, 722380003, 722454003, 722455002, 722456001, 722459008, 722477003, 722478008, 723304001, 723332005, 723333000, 723336008, 723365002, 723403008, 723410002, 723441001, 723454008, 723501008, 723504000, 723621000, 723676007, 723994004, 724001005, 724039002, 724137002, 724178000, 724207001, 724228005, 725140007, 725163002, 725289009, 725589005, 725906006, 725908007, 725912001, 726031001, 726621009, 726669007, 726670008, 726672000, 726709001, 726727003, 726732002, 732246009, 732251003, 732954002, 732957009, 732958004, 732961003, 733031004, 733032006, 733049004, 733050004, 733062000, 733072002, 733086003, 733088002, 733097003, 733110004, 733116005, 733117001, 733417008, 733419006, 733455003, 733472005, 733522005, 734017008, 734173003, 734349003, 763136000, 763186006, 763278004, 763320005, 763344007, 763350002, 763353000, 763404001, 763615003, 763618001, 763626009, 763665007, 763722004, 763741001, 763742008, 763743003, 763744009, 763745005, 763773007, 763795006, 763797003, 763837007, 763861000, 764455002, 764732004, 764861005, 764950001, 764959000, 765089003, 765170001, 765434008, 765471005, 765758008, 765761009, 766753005, 766824003, 766870005, 766871009, 768473009, 768677000, 768843007, 770404004, 770431001, 770564004, 770565003, 770604006, 770679002, 770719004, 770721009, 770723007, 770725000, 770750002, 770751003, 770755007, 770756008, 770790004, 770793002, 770794008, 770898002, 770901001, 770941005, 771074000, 771077007, 771148008, 771149000, 771179007, 771262009, 771336003, 771448004, 771470001, 771472009, 771476007, 771477003, 771512003, 772127009, 772224009, 772225005, 773230003, 773274001, 773303005, 773307006, 773329005, 773400009, 773404000, 773405004, 773416006, 773418007, 773419004, 773493002, 773494008, 773498006, 773547003, 773548008, 773551001, 773552008, 773553003, 773554009, 773556006, 773578004, 773581009, 773583007, 773587008, 773621003, 773670004, 773692000, 773699009, 773735007, 773769008, 773772001, 773984007, 774068004, 774070008, 774102003, 774149004, 774203000, 776204008, 777998000, 778009001, 778011005, 778025006, 780827006, 782723007, 782736007, 782753000, 782755007, 782757004, 782772000, 782886007, 782911008, 782941005, 782945001, 783005002, 783061008, 783089006, 783174004, 783619003, 783702009, 783703004, 785298001, 785726009, 232059000, 234146006, 236529001, 253176002, 40700009, 57917004, 59252009, 61152003, 77287004, 412787009, 422437002, 432091002, 1010630006, 879939002, 890118006, 1229895008, 1229875002, 1228886008, 1229882003, 1228890005, 1229873009, 1229883008, 1229891004, 1222672002, 1220600004, 1222656005, 1222706005, 1230376005, 77128003, 460436001, 767263007, 190991007, 39951000119105, 191689008, 191690004, 373618009, 231536004, 35919005, 43614003, 71961003, 408856003, 408857007, 442314000, 870260008, 870261007, 870262000, 870263005, 870264004, 870265003, 870266002, 870267006, 870268001, 870269009, 870270005, 870280009, 870282001, 870303005, 870304004, 870305003, 870306002, 870307006, 870308001, 23560001, 68618008, 702450004, 702732007, 708037001, 712884004, 718393002, 719600006, 722287002, 723332005, 733623005, 766824003,

770790004, 771448004, 771512003, 783089006, 191692007, 191693002, 432091002, 192780002, 21111006, 613003, 254265001, 254266000, 254268004, 156995003, 157019002, 157021007, 157022000, 205614001, 205617008, 205618003, 205621001, 205622008, 205625005, 205626006, 205673000, 205674006, 20598008, 41040004, 51500006, 254263008, 254267009, 254274004, 268344000, 268357008, 1216940001, 1162460001, 1162478006, 1162440009, 4135001, 4242009, 4325000, 6002006, 15841002, 16129004, 17122004, 19419002, 19550003, 24786004, 29257000, 29379007, 30361005, 31981007, 81304006, 1153430004, 1153583005, 787171006, 840505007, 125501000119105, 125511000119108, 125521000119101, 63247009, 449818005, 449819002, 699254009, 699305004, 699306003, 699307007, 699308002, 699310000, 702346005, 702357000, 709469005, 711489004, 715215007, 715438008, 716381003, 716387004, 716456000, 716515000, 717633007, 718227006, 718615003, 718687003, 718688008, 719046005, 719047001, 719574007, 719575008, 719576009, 719577000, 719580004, 719584008, 719597005, 719599008, 719600006, 719646006, 719649004, 719650004, 719651000, 719652007, 719657001, 719658006, 719659003, 719660008, 719661007, 719662000, 719663005, 719664004, 719666002, 719686003, 719808002, 721158009, 724070005, 724098008, 724147004, 724282009, 724643004, 724644005, 726365007, 726366008, 726367004, 726368009, 726369001, 726370000, 726371001, 726372008, 726373003, 726374009, 726375005, 726376006, 726377002, 726378007, 726379004, 726380001, 726381002, 726382009, 726383004, 726384005, 726385006, 726386007, 726387003, 726388008, 726389000, 726390009, 726391008, 726392001, 726393006, 726394000, 726395004, 726396003, 726397007, 726398002, 726399005, 726705007, 726733007, 732259001, 733194007, 733300002, 733519008, 733520002, 733521003, 733626002, 734029004, 734030009, 734349003, 763062006, 763527007, 763528002, 763529005, 763530000, 764440006, 764696007, 764725008, 764739008, 765171002, 766050000, 766716004, 766766005, 767263007, 768471006, 768555009, 770401007, 770410004, 770411000, 770566002, 770629000, 770665005, 770719004, 770754006, 770756008, 770760006, 770902008, 770905005, 770944002, 771072001, 771340007, 771439009, 773326003, 773346008, 773493002, 773494008, 773547003, 778000002, 778007004, 782673001, 782674007, 782694003, 783061008, 783164002, 783619003, 205615000, 205616004, 205619006, 205620000, 205623003, 205624009, 205630009, 371045000, 36369001, 36608002, 37506004, 43420005, 56653005, 62995000, 70173007, 270889005, 270890001, 1003322006, 1003358004, 1003364006, 1003380001, 1003391008, 1003392001, 1003393006, 1003394000, 1003396003, 1003417005, 1003418000, 1003419008, 1003420002, 1003449005, 1003894006, 1003895007, 1003896008, 1003897004, 1003898009, 1003899001, 1003900006, 1003901005, 1003902003, 1003903008, 1003904002, 1003910002, 1003911003, 1003912005, 1003913000, 1003914006, 1003915007, 1003929008, 1003930003, 1003931004, 1003932006, 1003933001, 1003934007, 1003935008, 1003937000, 879939002, 880078001, 880081006, 880086001, 880093002, 890117001, 890123006, 890124000, 890125004, 890127007, 890128002, 890130000, 890180006, 897524009, 897526006, 897527002, 897528007, 897529004, 897536003, 897542004, 897561000, 897564008, 897566005, 897568006, 897569003, 1229895008, 1229875002, 1228886008, 1229882003, 1228890005, 1229873009, 1228844002, 1229891004, 7199000, 765331004, 233718008, 36025004, 254243001, 722859001, 78572006, 6729006, 7199000, 19133005, 23150001, 92503002, 19886006, 54411001, 58037000, 92824003, 234138005, 781641005, 698765007, 698766008, 703284009, 703285005, 703286006, 715344006, 716862002, 722385008, 722451006, 724091002, 733469003, 763867001, 765331004, 771473004, 233718008, 234139002, 36025004, 46659004, 53633000, 403775003, 403815003, 403816002, 403817006, 403819009, 403820003, 403821004, 403822006, 403856000, 254241004, 254243001, 1003465006, 1010653007, 880093002, 22199006, 19092004, 40354009, 205808005, 43929004, 1177173001, 1220595008, 1187303004, 1177175008, 2391001, 2593002, 4434006, 7903009, 14870002, 14921002, 22567005, 22764001, 24629003, 28681006, 29248006, 86268005, 89454001, 95243004, 789777007, 4530000, 7720002, 15069006, 17818006, 33979003, 82699004, 719172003, 699870002, 702400006, 702829000, 703523004, 711156009, 715242008, 715471007, 715472000, 715652002, 715672007, 715673002, 715674008, 715862006, 716195006, 717264003, 717330004, 717823001, 719165004, 719166003, 719171005, 719201004, 719202006, 719397009, 719405005, 719471002, 719685004, 719688002,

719689005, 720826006, 720863002, 721148005, 721975004, 722210007, 722433005, 723995003, 725417001, 733419006, 763868006, 763885008, 763893008, 764460003, 764942005, 765204000, 765761009, 766717008, 766750008, 766820007, 766821006, 766874001, 768935003, 770603000, 770909004, 770948004, 773300008, 773302000, 773303005, 773404000, 773693005, 782821004, 782941005, 782951006, 783789002, 205468002, 205473008, 304576008, 313339007, 313426007, 389161008, 389162001, 389163006, 389165004, 38494008, 41483000, 42725006, 45582004, 51952004, 58561002, 59708000, 74928006, 76520005, 403765001, 254049009, 254060000, 254061001, 254062008, 254064009, 254065005, 254066006, 254067002, 254068007, 254079002, 254084008, 254085009, 254088006, 1010609002, 874931001, 890233009, 890237005, 890380004, 609437000, 205788004

ICD-10-CM: F70, F71, F72, F73, F78, F79, F78.A1, F78.A, F78.A9, D82.1, F84, F84.0, F84.2, F84.3, F84.5, F84.8, F84.9, E75.0, E75.1, E75.23, E75.25, E75.26, E75.29, E75.4, F84.2, G31.81, G31.82, G31.9, G93.89, G93.9, E75.00, E75.01, E75.02, E75.09, E75.10, E75.11, E75.19, Q90, Q91, Q92, Q93, Q95.2, Q95.3, Q99.2, Q99.8, Q99.9, Q90.0, Q90.1, Q90.2, Q90.9, Q91.0, Q91.1, Q91.2, Q91.3, Q91.4, Q91.5, Q91.6, Q91.7, Q92.0, Q92.1, Q92.2, Q92.5, Q92.6, Q92.61, Q92.62, Q92.7, Q92.8, Q92.9, Q93.0, Q93.1, Q93.2, Q93.3, Q93.4, Q93.5, Q93.51, Q93.59, Q93.7, Q93.8, Q93.81, Q93.82, Q93.88, Q93.89, Q93.9, Q85.1, Q85.8, Q85.9, E78.72, Q87.1, Q87.2, Q87.3, Q87.5, Q87.8, Q89.7, Q89.8, Q87.11, Q87.19, Q87.81, Q87.82, Q87.89, P04.3, Q86.0

ICD-9-CM: 317, 318, 319, 279.11, 299, 330, 758.0, 758.1, 758.2, 758.3, 758.5, 758.89, 758.9, 759.5, 759.6, 759.7, 759.81, 759.83, 759.89, 760.71

##### Sensory disability

SNOMED CT: 1955003, 11144004, 15188001, 21905007, 457061000124107, 1085711000119101, 1089891000119102, 15864131000119107, 15864171000119105, 15864251000119108, 15864291000119103, 15983431000119106, 15983471000119109, 15984551000119102, 15984591000119107, 29715005, 43353004, 609125008, 700453005, 155255001, 204256004, 204258003, 232217008, 35547002, 40320008, 43557002, 44057004, 49224008, 57436000, 60700002, 71619006, 75231006, 75355004, 268172000, 275259005, 1169356004, 1173035001, 1220595008, 1197059004, 1230014007, 1208480004, 1179361001, 1230344000, 1222655009, 1228871002, 1230412003, 1222649004, 1208614008, 1229999001, 1204415006, 1186656004, 1172635005, 1187512003, 1172605003, 1167372000, 1172604004, 1179555000, 2061000, 290006, 3820005, 8531006, 10170007, 15123008, 15190000, 16512005, 22138001, 24559001, 26061003, 26718008, 30169000, 84063009, 85571008, 89496002, 92894005, 92918008, 92937004, 93345006, 93466004, 95515009, 95820000, 95821001, 95828007, 109393007, 109395000, 109397008, 111339003, 129629004, 129630009, 129631008, 1142112002, 788873009, 840480002, 840482005, 3541000119107, 3551000119109, 3561000119106, 18701000119107, 18711000119105, 21451000119101, 23631000119109, 23641000119100, 457051000124105, 1083811000119108, 1084051000119104, 1084151000119100, 1088611000119103, 1088891000119106, 1088931000119103, 1089221000119100, 1091221000119100, 1091541000119108, 1091831000119101, 1092791000119103, 15864091000119105, 15864211000119107, 15979191000119107, 15979271000119100, 15983231000119107, 15983311000119105, 15983511000119100, 15984431000119109, 15984471000119107, 15984511000119103, 15984631000119107, 15984671000119105, 15984711000119109, 15984751000119105, 16888881000119105, 16889021000119105, 16889061000119100, 1271009, 47434006, 70348004, 70694009, 77507001, 93623004, 240554006, 473421004, 473422006, 473423001, 473424007, 698614006, 698615007, 699238006, 699316006, 699866005, 700112007, 700454004, 700489002, 702360007, 702362004, 702441001, 702443003, 711409002, 715239002, 715483009, 715504003, 715506001, 715527006, 715529009, 715666007,

715952000, 716007007, 716110002, 716112005, 716170005, 716192009, 716238003, 716239006, 716243005, 716245003, 717183001, 717761005, 717766000, 717767009, 717768004, 717791000, 717826009, 717909004, 717944002, 718574003, 718681002, 718714006, 718763005, 719102004, 719272007, 719305006, 719468005, 719689005, 719800009, 719817002, 719838008, 720506002, 720515009, 720517001, 720518006, 720519003, 720567008, 720601000, 720612000, 720634003, 720639008, 720640005, 720748007, 720749004, 720859009, 720955004, 720957007, 721084001, 721085000, 721086004, 721087008, 721089006, 721092005, 721207002, 721221000, 721294001, 721584005, 721836009, 721888002, 721973006, 722006004, 722019000, 722035007, 722054007, 722108000, 722111004, 722203001, 722213009, 722285005, 722293005, 722378009, 722389002, 722449007, 722453009, 722468005, 722476007, 723442008, 723452007, 723497003, 723504000, 723611008, 723721007, 723993005, 724001005, 724092009, 724093004, 724139004, 724173009, 724282009, 724636005, 725096002, 725137007, 726106004, 726621009, 726669007, 726672000, 726709001, 732248005, 732951005, 732953008, 733029008, 733050004, 733065003, 733069009, 733071009, 733089005, 733419006, 734022008, 737047001, 737048006, 737049003, 737050003, 737344003, 737377004, 738164003, 762658005, 762662004, 763130006, 763136000, 763279007, 763688008, 763774001, 763815000, 763860004, 764097002, 764098007, 764810000, 765178008, 765325002, 766249007, 766870005, 770414008, 770751003, 770784003, 770944002, 771308008, 773398005, 773406003, 773575001, 773583007, 773610007, 773647007, 773648002, 773665006, 773667003, 773735007, 773750003, 774155009, 777998000, 778006008, 780818005, 780819002, 782690007, 782752005, 782942003, 783097004, 783555001, 783614008, 783700001, 783742004, 783774006, 162344009, 164060004, 164061000, 164062007, 164063002, 164068006, 164070002, 186570004, 194387002, 194413008, 194414002, 194415001, 194416000, 194417009, 194419007, 194424005, 194426007, 194429000, 204223000, 204245004, 204250005, 204252002, 204257008, 343087000, 361265002, 373905003, 232057003, 232058008, 232262007, 232298004, 232299007, 232300004, 232301000, 232302007, 232325008, 232326009, 232328005, 232329002, 232330007, 232331006, 232332004, 232333009, 232335002, 236529001, 236532003, 237612000, 237617006, 237619009, 238047006, 253204009, 253251006, 253252004, 253253009, 253254003, 253255002, 35045004, 37635004, 38919006, 40897007, 42538001, 42853003, 44315004, 44621005, 46050004, 47111006, 48758008, 49526009, 54036001, 54386000, 55647004, 56943007, 57838006, 61743004, 61947007, 63702009, 65680009, 66597000, 67817003, 67985002, 68467004, 69056000, 70583001, 70967007, 71055004, 71973003, 72964008, 73119000, 73310008, 73371001, 73415002, 75311005, 75922002, 76223006, 79442007, 79471008, 79665007, 1179362008, 403805009, 427644005, 427772009, 428470000, 428887009, 429967001, 430977001, 430985005, 433147009, 441944007, 254173004, 262764000, 267677007, 268166000, 272033007, 275482009, 275879008, 277424008, 277494001, 277495000, 277663005, 282038006, 1010229008, 1010230003, 1010236009, 1010238005, 1010439000, 1010440003, 1010441004, 1010442006, 1010606009, 1010610007, 1010636000, 1010638004, 1052205008, 1052206009, 1119386008, 1119387004, 878795009, 878796005, 878797001, 897530009, 897539005, 897557006, 897572005, 897574006, 897591005, 340411000119108, 334811000119101, 332941000119101, 1361009, 1847009, 2532009, 122003, 5425007, 5743005, 5962000, 6141006, 11422002, 11547003, 12371008, 16553002, 17657000, 19721008, 20842008, 23986001, 24596005, 25277000, 25859000, 25913001, 26468004, 27871007, 28998008, 29555009, 29668007, 32022003, 34037000, 81796004, 82576008, 84884003, 85478004, 87807004, 93478000, 95251001, 95500008, 95695004, 95709007, 95722004, 105597003, 111512005, 128306009, 128329001, 128473001, 816119002, 816120008, 833278008, 334791000119100, 336801000119107, 340391000119108, 344481000119109, 345731000119104, 347471000119101, 347531000119100, 347541000119109, 347591000119101, 347601000119108, 430801000124103, 678601000119100, 678611000119102, 678621000119109, 680781000119108, 680791000119106, 680801000119107, 4855003, 22681004, 193570009, 445922006, 700280001, 733318004, 735646005, 762357005, 762358000, 762436001, 762617003, 155120009, 155126003, 193379006, 193504001, 193506004, 193507008, 193508003, 193509006, 193510001, 193511002, 193514005, 193515006, 193517003, 193518008, 193525001, 193526000, 194068002, 194069005, 194070006, 194071005,

194608001, 194631001, 312770008, 312918002, 359766000, 370941004, 371756009, 212141007, 212142000, 231948009, 231949001, 231951002, 231962002, 232067008, 247037009, 247136003, 247200005, 253235003, 35323005, 37761009, 39450006, 42059000, 43727009, 45953007, 47269005, 49857001, 52276000, 53854005, 53866004, 54320009, 55456003, 56311003, 57441008, 57534004, 61267008, 62585004, 62795009, 63317005, 66485003, 68241007, 68574006, 68575007, 68633000, 69820004, 70397008, 70992005, 73757007, 74469006, 75614007, 75895005, 77971008, 78778007, 78875003, 80499003, 400958004, 400959007, 404631002, 406446000, 415297005, 418801006, 267622003, 267623008, 267624002, 345751000119105, 338551000119108, 344321000119100, 1172594000, 1177028005, 1179296003, 10878931000119103, 10878881000119108, 1217651008, 1217650009, 1217661001, 1220595008, 1197357008, 1229998009, 1217688006, 1230020008, 1217674007, 1231725002, 15731241000119101, 15731321000119101, 1231258007, 1231280005, 1231278004, 1231275001, 1197365006, 1231183003, 1231271005, 1231270006, 1196868007, 677301000119106, 677311000119109, 1208838005, 1217647006, 1216988003, 1208840000, 820063141000119101, 377096861000119104, 1217370006, 342031000119105, 15665881000119107, 341461000119103, 341411000119101, 338911000119103, 340991000119109, 340541000119108, 15993351000119107, 339971000119103, 675341000119102, 675351000119100, 1231253003, 1217645003, 1197315002, 1231227006, 1231228001, 1222665003, 1220599002, 1197180007, 15997831000119104, 15997791000119105, 1220597000, 336421000119100, 15665841000119102, 335851000119101, 335801000119100, 333301000119107, 335381000119103, 334931000119102, 15993311000119106, 334371000119100, 333401000119104, 332951000119104, 15663481000119106, 15734921000119102, 334961000119105, 332081000119107, 332051000119100, 332031000119106, 15685481000119107, 331821000119106, 1222888000, 1208341008, 1217667002, 1229938006, 1208829008, 1204415006, 349231000119108, 630352961000119108, 15993471000119105, 345571000119106, 790731321000119100, 675331000119106, 345341000119107, 561258721000119104, 345241000119101, 1217652001, 1231243004, 1196911006, 15997991000119107, 1176990003, 1163479009, 1163460006, 1172683008, 1172900005, 1187543005, 1172605003, 1179282009, 1177029002, 1177018001, 1177031006, 1187639002, 1172636006, 1217638003, 1231150005, 1231225003, 539009, 776009, 849000, 875001, 903008, 943003, 1079004, 1207009, 1412008, 1493002, 1654001, 1674008, 1833005, 1869002, 1909000, 1979001, 2167004, 2251002, 2463005, 2790002, 2951009, 3426005, 3506008, 3577000, 3598000, 264008, 4040006, 4041005, 4178006, 4229009, 4490000, 4646004, 4927003, 5310008, 5318001, 5361003, 5587004, 6131008, 6313001, 6395007, 6869001, 6962006, 7089008, 7219007, 7366009, 7469001, 7611002, 7758003, 8207002, 8231007, 8247009, 8554002, 8656007, 9091006, 9585003, 9633006, 9941009, 10613001, 10698009, 10877007, 10970008, 11096003, 11255009, 11391004, 11603001, 11623000, 12195004, 12411003, 12603003, 12630008, 13045009, 13282008, 13608004, 13706005, 13734003, 13937002, 13978000, 14046000, 14427009, 14781008, 14894002, 15010004, 15059000, 15228007, 15236003, 15374009, 15552004, 15847003, 16150008, 16169003, 16350000, 17140000, 17480001, 18121009, 18377001, 18410006, 19031009, 19144002, 19309007, 19620000, 19871000, 20555001, 21011008, 21117005, 21328003, 21571006, 21590003, 21709007, 21928008, 21939006, 22017009, 22067002, 22135003, 22385004, 22455005, 22950006, 23320001, 23653003, 23672003, 23798003, 24005006, 24210004, 24225004, 24911006, 24950005, 25412000, 25506007, 26214006, 27339005, 27735002, 28143002, 28157000, 28391008, 28550007, 28578008, 28835009, 28900006, 28946006, 29050005, 29297002, 29369005, 29504002, 29538005, 29590001, 29887003, 30041005, 30069002, 30080002, 30272003, 30337003, 30537002, 31079007, 31264007, 31341008, 31550006, 32893002, 33382000, 33647009, 34361001, 34533008, 1208932005, 345941000119106, 1231274002, 868431021000119100, 337691000119103, 337661000119105, 337641000119106, 80734006, 80744008, 82617003, 82696006, 83405000, 83785004, 84333006, 84494001, 85366002, 86103006, 86191000, 86219005, 86266009, 86643000, 86923008, 87194000, 87224000, 87614000, 87764000, 88032003, 88290000, 88538005, 88768004, 89114005, 89182000, 89188001, 89215000, 91268007, 91408006, 91570002, 92060009,

92321003, 92566008, 92699002, 92820007, 92829008, 93435005, 93470007, 93756008, 93987004, 94255003, 94516003, 94796001, 95027001, 95207005, 95213001, 95221007, 95223005, 95250000, 95255005, 95479005, 95484004, 95485003, 95486002, 95488001, 95493003, 95494009, 95495005, 95496006, 95497002, 95498007, 95680001, 95682009, 95683004, 95685006, 95686007, 95688008, 95689000, 95690009, 95691008, 95692001, 95693006, 95694000, 95696003, 95697007, 95699005, 95700006, 95701005, 95702003, 95703008, 95710002, 95711003, 95712005, 95714006, 95715007, 95716008, 95717004, 95720007, 95723009, 95724003, 95737000, 95774001, 95800001, 110998005, 110999002, 111000003, 111510002, 111511003, 111513000, 111514006, 111515007, 111518009, 111519001, 111521006, 111522004, 118166004, 123614006, 123670005, 126998003, 127002001, 128080005, 128331005, 128533009, 128985003, 129623003, 1081591000119101, 1081601000119108, 1137552000, 1137553005, 1137554004, 1137571003, 1137633000, 1144419002, 1144978007, 1148920008, 1153347002, 1156246002, 1156843000, 1156844006, 1156845007, 1156846008, 15991471000119106, 15991511000119102, 353061000119107, 353081000119103, 1176981005, 1176982003, 339011000119105, 1187229001, 15986471000119104, 1187232003, 787051000, 787052007, 788718000, 788913001, 788945006, 788946007, 788947003, 788948008, 788963006, 788967007, 788995007, 789008006, 789116000, 789220007, 789675009, 789676005, 789700005, 792908009, 816177009, 816178004, 816961009, 816962002, 823016000, 829979008, 829980006, 830270002, 830271003, 838350007, 840456001, 840489001, 860721006, 860798008, 860836003, 866099002, 866114005, 866133000, 870199008, 870273007, 870312007, 870329005, 870330000, 870331001, 870332008, 870333003, 870335005, 870346008, 870420005, 870421009, 870529009, 871778008, 871781003, 391000119105, 1491000119102, 1501000119109, 1551000119108, 8041000119108, 11211000119108, 15681000119102, 24151000119106, 28331000119107, 31181000119100, 31191000119102, 35091000119101, 35531000119107, 35881000119106, 40121000119107, 41911000119107, 60951000119105, 60961000119107, 60971000119101, 60981000119103, 60991000119100, 82541000119100, 82551000119103, 82571000119107, 82581000119105, 83231000119101, 97331000119101, 97341000119105, 103841000119102, 103981000119101, 104941000119109, 104951000119106, 104961000119108, 109171000119104, 117831000119104, 124681000119104, 126591000119108, 133131000119107, 133361000119101, 138881000119106, 138891000119109, 138901000119108, 138911000119106, 138921000119104, 138941000119105, 140961000119107, 217791000119107, 220101000119108, 331071000119101, 331101000119105, 331111000119108, 331151000119109, 331371000119103, 331381000119100, 331471000119105, 331781000119101, 331791000119103, 331801000119102, 331811000119104, 331831000119109, 331931000119100, 331981000119104, 332091000119105, 332101000119100, 332111000119102, 332341000119100, 332961000119102, 332371000119107, 332381000119105, 332391000119108, 332411000119108, 332421000119101, 332431000119103, 332441000119107, 332481000119102, 332491000119104, 332521000119102, 332711000119103, 332751000119102, 332771000119106, 332831000119101, 332851000119107, 332871000119103, 332921000119107, 332971000119108, 333041000119107, 333071000119100, 333081000119102, 333091000119104, 333111000119107, 333151000119108, 333161000119105, 333171000119104, 333231000119107, 333241000119103, 333251000119101, 333261000119104, 333281000119108, 333391000119101, 333531000119105, 333661000119101, 333751000119107, 333881000119109, 333971000119101, 334311000119108, 334321000119101, 334691000119106, 334821000119108, 334831000119106, 334891000119105, 334901000119109, 335061000119105, 335161000119109, 335201000119104, 335351000119105, 335531000119109, 335541000119100, 335551000119103, 335831000119107, 335841000119103, 335861000119104, 335871000119105, 335961000119106, 335971000119100, 336031000119105, 336041000119101, 336051000119104, 336061000119102, 336071000119108, 336081000119106, 336091000119109, 336101000119104, 336191000119105, 336201000119108, 336221000119104, 336251000119107, 336331000119108, 336501000119105, 336511000119108, 336531000119103, 336541000119107, 336611000119109, 336631000119104, 336791000119106, 336811000119105, 336851000119106, 336911000119104, 336921000119106, 336981000119105, 336991000119108, 337081000119109, 337401000119107, 337411000119105, 337421000119103, 337431000119100, 337451000119106, 337551000119107, 337601000119109, 337701000119103, 337711000119100, 337721000119107, 337951000119101, 337981000119108,

337991000119106, 338001000119102, 338021000119106, 338031000119109, 338041000119100, 338051000119103, 338091000119108, 338101000119103, 338131000119105, 338321000119109, 338361000119104, 338381000119108, 338441000119105, 338461000119109, 338481000119100, 338531000119102, 338571000119104, 338581000119101, 338651000119109, 338681000119102, 338691000119104, 338701000119104, 338721000119108, 338761000119103, 338771000119109, 338781000119107, 338841000119108, 338851000119105, 338861000119107, 338871000119101, 338891000119100, 339001000119107, 339141000119108, 339261000119101, 339351000119108, 339481000119109, 339571000119105, 339911000119106, 339921000119104, 340291000119103, 340421000119101, 340441000119107, 340501000119106, 340511000119109, 340671000119102, 340781000119109, 340811000119106, 340961000119102, 341141000119106, 341151000119108, 341161000119105, 341441000119102, 341451000119100, 341471000119109, 341481000119107, 341571000119108, 341581000119106, 341641000119100, 341651000119103, 341661000119101, 341671000119107, 341681000119105, 341691000119108, 341701000119108, 341711000119106, 341801000119101, 341811000119103, 341831000119108, 341861000119100, 341941000119108, 342111000119101, 342121000119108, 342141000119102, 342151000119100, 342221000119104, 342241000119105, 342291000119102, 342381000119109, 342581000119102, 342741000119103, 342751000119101, 342821000119103, 342831000119100, 342911000119104, 343021000119100, 343031000119102, 343041000119106, 343051000119108, 343091000119103, 343101000119108, 343161000119109, 343191000119102, 343201000119104, 343221000119108, 343231000119106, 343341000119108, 343411000119100, 343461000119102, 343471000119108, 343481000119106, 343491000119109, 343721000119103, 343751000119106, 343761000119108, 343771000119102, 343781000119104, 343791000119101, 343801000119100, 343811000119102, 343821000119109, 343841000119103, 343861000119104, 343871000119105, 344091000119101, 344131000119104, 344151000119105, 344211000119104, 344231000119109, 344251000119103, 344301000119109, 344341000119106, 344351000119108, 344421000119105, 344451000119102, 344461000119100, 344471000119106, 344491000119107, 344541000119103, 344551000119101, 344611000119101, 344621000119108, 344631000119106, 344641000119102, 344671000119109, 344751000119109, 344891000119109, 345021000119104, 345111000119105, 345211000119100, 345291000119109, 345351000119109, 345611000119102, 345771000119101, 345811000119101, 346231000119100, 346241000119109, 346251000119106, 346261000119108, 346341000119103, 346401000119108, 346411000119106, 346421000119104, 346691000119104, 346701000119104, 346711000119101, 346721000119108, 346731000119106, 346791000119105, 346801000119106, 346861000119107, 346871000119101, 346901000119101, 346911000119103, 346921000119105, 346931000119108, 347011000119102, 347021000119109, 347041000119103, 347071000119105, 347121000119105, 347151000119102, 347211000119108, 347251000119109, 347261000119106, 347271000119100, 347281000119102, 347301000119103, 347311000119100, 347381000119106, 347401000119106, 347461000119107, 347481000119103, 347521000119103, 347561000119108, 347581000119104, 347991000119105, 348071000119106, 348081000119109, 348151000119103, 348311000119101, 348351000119100, 348361000119103, 348381000119107, 348431000119100, 348521000119104, 348531000119101, 348601000119109, 348761000119102, 348801000119105, 348811000119108, 348831000119103, 349061000119107, 349111000119108, 349121000119101, 349271000119106, 349281000119109, 349301000119108, 349311000119106, 351311000119107, 352941000119102, 354071000119104, 368711000119106, 368721000119104, 432141000124105, 434391000124109, 450641000124101, 450921000124103, 675361000119103, 675371000119109, 675381000119107, 677381000119103, 677391000119100, 677401000119103, 677421000119107, 677431000119105, 677501000119102, 677511000119104, 677521000119106, 677531000119109, 677541000119100, 677551000119103, 677571000119107, 677581000119105, 677591000119108, 677601000119101, 677611000119103, 677741000119109, 677771000119102, 677781000119104, 677791000119101, 677801000119100, 677811000119102, 677821000119109, 677831000119107, 677851000119101, 677871000119105, 677891000119106, 677901000119105, 677911000119108, 677921000119101, 677931000119103, 677941000119107, 677951000119109, 677961000119106, 677971000119100, 677981000119102, 678051000119107, 678061000119109, 678071000119103, 678081000119100, 678091000119102, 678101000119107, 678161000119108, 678171000119102, 678181000119104, 678191000119101, 678201000119103, 678211000119100, 678281000119106, 678291000119109, 678301000119105, 678311000119108, 678321000119101, 678331000119103, 678351000119109, 678361000119106,

678381000119102, 678391000119104, 678401000119102, 678411000119104, 678421000119106, 678471000119107, 678481000119105, 678491000119108, 678501000119101, 678511000119103, 678581000119109, 678591000119107, 678671000119105, 678681000119108, 678691000119106, 678821000119108, 678831000119106, 678841000119102, 678881000119107, 678891000119105, 678901000119109, 679121000119108, 679131000119106, 679171000119109, 679181000119107, 679191000119105, 679211000119106, 679231000119101, 679251000119107, 679911000119104, 679931000119109, 680291000119102, 680531000119103, 680821000119103, 680831000119100, 680841000119109, 680941000119100, 681041000119106, 1078791000119109, 1078801000119105, 1092761000119105, 1099031000119105, 1099041000119101, 1099051000119104, 1099241000119105, 10762111000119102, 10762151000119101, 10762191000119106, 10942751000119100, 10942791000119105, 10999281000119103, 10999321000119108, 10999361000119103, 12237911000119109, 12237951000119105, 12239101000119100, 12239141000119103, 12239181000119108, 12239301000119102, 12239421000119101, 12239461000119106, 12239501000119106, 13015741000119105, 13015781000119100, 13015821000119105, 15630771000119104, 15630811000119104, 15630851000119103, 15631531000119100, 15631611000119106, 15632931000119101, 15632971000119103, 15633011000119103, 15633051000119102, 15633091000119107, 15633131000119109, 15633171000119107, 15633201000119106, 15633241000119108, 15633281000119103, 15633321000119108, 15638451000119104, 15638491000119109, 15638531000119109, 15639131000119107, 15639181000119108, 15639231000119102, 15639281000119101, 15639321000119106, 15639361000119101, 15639401000119105, 15639441000119107, 15639481000119102, 15640441000119104, 15663561000119101, 15664241000119100, 15664281000119105, 15665321000119101, 15665361000119106, 15665401000119102, 15665521000119105, 15665681000119108, 15665721000119102, 15665761000119107, 15673001000119103, 15673041000119101, 15673081000119106, 15677681000119107, 15677721000119101, 15677761000119106, 15677801000119103, 15677841000119101, 15677881000119106, 15678041000119108, 15678081000119103, 15678441000119103, 15678481000119108, 15678521000119108, 15678561000119103, 15678601000119103, 15678641000119101, 15678681000119106, 15678761000119105, 15678801000119102, 15679241000119108, 15679321000119108, 15679361000119103, 15679401000119107, 15679441000119109, 15679481000119104, 15679561000119109, 15679681000119101, 15679721000119107, 15679761000119102, 15679801000119105, 15681561000119104, 15681681000119107, 15681721000119101, 15685321000119103, 15685361000119108, 15685401000119104, 15685441000119102, 15685561000119102, 15685641000119100, 15685801000119101, 15685841000119104, 15685881000119109, 15692601000119109, 15692641000119106, 15692681000119101, 15699201000119104, 15699241000119102, 15699281000119107, 15705441000119107, 15705481000119102, 15705521000119102, 15705561000119107, 15708081000119103, 15708121000119101, 15708161000119106, 15709961000119104, 15710001000119105, 15710041000119107, 15710081000119102, 15710121000119100, 15710161000119105, 15710881000119109, 15710921000119102, 15710961000119107, 15731281000119106, 15731521000119105, 15731561000119100, 15732881000119102, 15732921000119109, 15732961000119104, 15733001000119107, 15733041000119109, 15733081000119104, 15733121000119102, 15733161000119107, 15733201000119102, 15733241000119100, 15733281000119105, 15733321000119100, 15733361000119105, 15733401000119101, 15733441000119104, 15733601000119104, 15733641000119102, 15733681000119107, 15734121000119100, 15734161000119105, 15734321000119103, 15734361000119108, 15734401000119104, 15734521000119107, 15734561000119102, 15734641000119100, 15734681000119105, 15734801000119101, 15734841000119104, 15734881000119109, 15735041000119108, 15735081000119103, 15735121000119101, 15735161000119106, 15735201000119101, 15735241000119104, 15735281000119109, 15735321000119104, 15735361000119109, 15735401000119100, 15735721000119100, 15735801000119102, 15736441000119108, 15736481000119103, 15736521000119103, 15736561000119108, 15736601000119108, 15736641000119105, 15736681000119100, 15736721000119106, 15736761000119101, 15736801000119109, 15736841000119106, 15736881000119101, 15737041000119101, 15737081000119106, 15737121000119108, 15737161000119103, 15737201000119108, 15737241000119105, 15737281000119100, 15737321000119105, 15737361000119100, 15737641000119108, 15737681000119103,

15737721000119109, 15737761000119104, 15737801000119107, 15737841000119109, 15737921000119106, 15737961000119101, 15738161000119104, 15738201000119109, 15738241000119106, 15738281000119101, 15738321000119106, 15738361000119101, 15738401000119105, 15738441000119107, 15738481000119102, 15738521000119102, 15738561000119107, 15738761000119100, 15738841000119105, 15738881000119100, 15738921000119107, 15739041000119106, 15739081000119101, 15739121000119104, 15739161000119109, 15739201000119104, 15739241000119102, 15739281000119107, 15739321000119102, 15739361000119107, 15739401000119103, 15739441000119101, 15739481000119106, 15739561000119101, 15739641000119104, 15739681000119109, 15739721000119103, 15739761000119108, 15739801000119100, 15739881000119108, 15740001000119101, 15740041000119104, 15740081000119109, 15911041000119105, 15911111000119105, 15911321000119101, 15911391000119104, 15913031000119100, 15921411000119106, 15930301000119107, 15930341000119109, 15930381000119104, 15986071000119108, 15986111000119101, 15986151000119100, 15986191000119105, 15986231000119101, 15986271000119103, 15986311000119103, 15986511000119108, 15986551000119109, 15986591000119104, 15986791000119106, 15986951000119103, 15986991000119108, 15987031000119108, 15987071000119106, 15987111000119104, 15987391000119105, 15987431000119100, 15989831000119105, 15989991000119102, 15990031000119108, 15990111000119104, 15990391000119105, 15990431000119100, 15990471000119102, 15991071000119102, 15991111000119109, 15991151000119105, 15991711000119108, 15991791000119104, 15992631000119101, 15992671000119103, 15992711000119104, 15992911000119101, 15992951000119100, 15992991000119105, 15993391000119102, 15993671000119108, 15993711000119107, 15993751000119108, 15996831000119101, 15997471000119103, 15997511000119107, 15997551000119108, 15997591000119103, 15997631000119103, 15997671000119100, 15997911000119103, 15997951000119102, 15998311000119103, 15998351000119102, 15998391000119107, 15999591000119107, 15999791000119100, 15999831000119106, 16000071000119103, 16000111000119105, 16000151000119106, 16064571000119103, 16077531000119101, 16229071000119108, 16279401000119108, 16279441000119105, 16320791000119106, 16320831000119100, 16320871000119102, 16376191000119109, 16419701000119108, 16419741000119105, 16419781000119100, 16477021000119103, 16711031000119109, 16711071000119107, 16711111000119100, 16711191000119109, 16711231000119100, 16711271000119102, 16711351000119101, 16711391000119106, 16711431000119101, 16711551000119108, 16711591000119103, 16711631000119103, 16716281000119108, 1135000, 4416007, 9936001, 11226001, 15509004, 21404001, 25362006, 26291002, 27971006, 35545005, 36524008, 42031001, 43959009, 46497009, 50801008, 51445007, 56787009, 56852002, 70099003, 75070005, 75739009, 79385002, 80651009, 89062002, 90036004, 95501007, 190615004, 193410003, 193413001, 193489006, 193497004, 193687000, 193731001, 193839007, 206456003, 206459005, 231930000, 231944006, 231974003, 232030000, 247132001, 271730003, 312912001, 312927001, 312946000, 312956001, 314015001, 314019007, 314429009, 314537004, 399868002, 399871005, 410480004, 410481000, 410485009, 410494003, 410495002, 417065002, 417115008, 420308006, 420756003, 421460008, 421706001, 421920002, 422189002, 699861000, 774010000, 766834007, 768792007, 768793002, 768794008, 768798006, 768799003, 769181007, 769185003, 769186002, 769187006, 769188001, 769190000, 769219006, 769220000, 769221001, 769222008, 770097006, 770323005, 770765001, 783787000, 335471000119108, 341081000119102, 368741000119105, 680521000119101, 15685601000119102, 15705401000119105, 15705601000119107, 15737881000119104, 677291000119105, 444248002, 444863008, 445213003, 445257004, 445921004, 446466006, 446474007, 447010005, 449866003, 698840003, 698847000, 698848005, 698881006, 698882004, 698883009, 699515009, 699516005, 699520009, 699521008, 699755009, 699757001, 699758006, 699759003, 699761007, 699763005, 702359002, 702398007, 703542000, 704272003, 706894000, 707608003, 711162004, 711407000, 711482008, 711566009, 711567000, 711616006, 711617002, 711644007, 712506009, 712507000, 712508005, 712733005, 713345005, 713457002, 715144004, 715240000, 715429006, 715465001, 715484003, 715528001, 715562001, 715628009, 715647007, 715908008, 715984007, 715988005, 715989002, 716099003, 716166002, 716170005, 716197003, 716663009, 716717005, 716998009, 717286002, 717761005,

717785002, 717812000, 717887003, 717893006, 717894000, 717920004, 718579008, 718680001, 718766002, 718851007, 719042007, 719102004, 719205008, 719266007, 719282008, 719296002, 719297006, 719431007, 719455002, 719517009, 719520001, 719808002, 719944006, 719976001, 720410001, 720460007, 720467005, 720468000, 720609003, 720636001, 720749004, 720750004, 720850008, 720856002, 720983002, 720987001, 721015008, 721076000, 721233005, 721546004, 721547008, 721862000, 721898008, 721974000, 722003007, 722062004, 722066001, 722108000, 722110003, 722282008, 722321001, 722329004, 722378009, 722379001, 722380003, 722381004, 722382006, 722437006, 722450007, 722457005, 722463001, 722604002, 722606000, 722607009, 722999002, 723023003, 723131006, 723169001, 723364003, 723408004, 723450004, 723452007, 723502001, 723503006, 723512008, 723582004, 723584003, 723612001, 723621000, 723828008, 723999009, 724000006, 724001005, 724002003, 724065003, 724175002, 724371004, 724384008, 724769002, 724809006, 725168006, 725289009, 725590001, 726078000, 726404007, 726405008, 726619004, 726704006, 726709001, 732246009, 732247000, 732251003, 732952003, 733049004, 733086003, 733110004, 733113002, 733116005, 733317009, 733319007, 733320001, 733321002, 733322009, 733323004, 733325006, 733326007, 733337008, 733455003, 733626002, 735648006, 735649003, 735659002, 735660007, 735661006, 735916009, 735919002, 735920008, 735922000, 735925003, 736321008, 736838001, 737006004, 737271000, 737272007, 737273002, 737579002, 762360003, 762361004, 762362006, 762363001, 762416002, 762417006, 762418001, 762419009, 762420003, 762523003, 762524009, 762525005, 762526006, 762663009, 763134002, 763387005, 763669001, 764452004, 764939004, 764969006, 765178008, 765191009, 766933000, 768797001, 768932000, 769182000, 769183005, 769184004, 769191001, 769217008, 769218003, 769244003, 769245002, 770324004, 770361008, 770362001, 770434009, 770437002, 770581008, 770582001, 770594005, 770599000, 770600002, 770626007, 770667002, 770766000, 770776002, 770777006, 770791000, 771235001, 771342004, 771476007, 772224009, 773328002, 773398005, 773548008, 773576000, 773577009, 773583007, 773627004, 773648002, 773690008, 773727009, 773991005, 773992003, 773993008, 774152007, 774212003, 778021002, 778042000, 778049009, 782724001, 782754006, 782822006, 782914000, 782949007, 783090002, 783160006, 783203003, 783246000, 783612007, 785884003, 785885002, 786049007, 786050007, 786054003, 786055002, 786076007, 786077003, 786078008, 162808005, 162815002, 163946009, 163960006, 163986001, 163988000, 163989008, 163990004, 163991000, 164024009, 186263000, 186266008, 186854007, 187039009, 187049007, 187058000, 187194004, 188263008, 188264002, 189151003, 192788009, 193300006, 193319001, 193320007, 193322004, 193323009, 193324003, 193325002, 193326001, 193327005, 193329008, 193338005, 193341001, 193342008, 193349004, 193350004, 193357001, 193370005, 193387007, 193400007, 193409008, 193411004, 193415008, 193428001, 193434008, 193436005, 193438006, 193442009, 193443004, 193444005, 193445006, 193446007, 193458007, 193459004, 193463006, 193466003, 193468002, 193474002, 193487008, 193488003, 193493000, 193501009, 193523008, 193531003, 193533000, 193534006, 193548006, 193552006, 193553001, 193555008, 193556009, 193561006, 193562004, 193576003, 193577007, 193589009, 193590000, 193598007, 193600001, 193602009, 193609000, 193610005, 193615000, 193683001, 193690006, 193691005, 193699007, 193703005, 193704004, 193707006, 193708001, 193709009, 193711000, 193712007, 193714008, 193716005, 193718006, 193719003, 193720009, 193722001, 193725004, 193727007, 193728002, 193732008, 193735005, 193736006, 193738007, 193741003, 193742005, 193745007, 193748009, 193749001, 193751002, 193795008, 193837009, 193880000, 193881001, 194168004, 197367007, 204113001, 204125003, 204127006, 204128001, 204138006, 204139003, 204148008, 204149000, 204152008, 204154009, 204159004, 204171005, 204173008, 204179007, 204181009, 204189006, 206248004, 206249007, 206446002, 206439006, 206442000, 206443005, 206444004, 206447006, 206448001, 206453006, 206454000, 206455004, 206457007, 206458002, 206469004, 206470003, 206471004, 303019009, 309594000, 309596003, 311782002, 311789006, 312491004, 312701008, 312702001, 312703006, 312704000, 312720005, 312898002, 312899005, 312901001, 312902008, 312903003, 312904009, 312905005, 312906006, 312907002, 312908007, 312909004, 312910009, 312911008, 312913006, 312914000, 312915004, 312916003, 312917007, 312920004, 312921000, 312922007, 312923002, 312924008, 312925009, 312928006,

312929003, 312930008, 312931007, 312932000, 312933005, 312934004, 312935003, 312937006, 312938001, 312940006, 312941005, 312943008, 312947009, 312949007, 312950007, 312951006, 312955002, 312957005, 312958000, 312959008, 312960003, 312961004, 312962006, 312963001, 312997008, 312998003, 312999006, 313000007, 313001006, 313002004, 313003009, 313164002, 313165001, 314000002, 314006008, 314007004, 314008009, 314009001, 314010006, 314011005, 314014002, 314017009, 314026007, 314028008, 314029000, 314030005, 314031009, 314032002, 314033007, 314265001, 314266000, 314267009, 314268004, 314269007, 314407005, 314418005, 314419002, 314420008, 314427006, 314428001, 314437001, 314467007, 314493000, 314494006, 314544008, 314545009, 314546005, 314547001, 314548006, 314766004, 314780006, 314784002, 314787009, 315288007, 315352000, 315353005, 359628008, 359668007, 359695002, 359698000, 359702001, 359707007, 360455002, 361122008, 361130009, 361199007, 363465007, 363677007, 367649002, 370483001, 370504007, 370506009, 370891001, 370967009, 371060002, 371098002, 371099005, 373426005, 373614006, 373650004, 373661007, 373667006, 384675002, 387712008, 388979002, 390834004, 392029006, 392030001, 392133001, 392288006, 392291006, 392300000, 392352004, 392461003, 392481002, 1187230006, 340571000119101, 1187231005, 1231259004, 225569001, 230504002, 230506000, 230513000, 230514006, 230515007, 230519001, 230526001, 231882004, 231883009, 231889008, 231898006, 231907006, 231908001, 231909009, 231912007, 231913002, 231915009, 231916005, 231917001, 231918006, 231919003, 231920009, 231924000, 231925004, 231926003, 231927007, 231928002, 231929005, 231931001, 231933003, 231934009, 231947004, 231950001, 231952009, 231953004, 231954005, 231955006, 231957003, 231959000, 231960005, 231961009, 231963007, 231964001, 231967008, 231968003, 231970007, 231971006, 231972004, 231981005, 231983008, 231984002, 231985001, 231986000, 231988004, 231990003, 231992006, 231994007, 231995008, 231996009, 231997000, 231999002, 232000008, 232001007, 232002000, 232003005, 232004004, 232005003, 232006002, 232007006, 232008001, 232009009, 232010004, 232011000, 232012007, 232013002, 232014008, 232015009, 232016005, 232017001, 232018006, 232020009, 232021008, 232022001, 232023006, 232024000, 232025004, 232027007, 232028002, 232029005, 232031001, 232032008, 232033003, 232034009, 232035005, 232036006, 232037002, 232038007, 232039004, 232040002, 232066004, 232041003, 232042005, 232043000, 232044006, 232045007, 232046008, 232048009, 232049001, 232050001, 232051002, 232052009, 232053004, 232054005, 232055006, 232056007, 232057003, 232058008, 232061009, 232062002, 232063007, 232064001, 232065000, 232068003, 232069006, 232071006, 232072004, 232073009, 232074003, 232075002, 232076001, 232078000, 232080006, 232081005, 232082003, 232083008, 232086000, 232087009, 232088004, 232090003, 232119000, 232120006, 232121005, 232122003, 232142007, 232143002, 232144008, 232146005, 232148006, 232150003, 233753001, 236531005, 237611007, 241973007, 246877005, 246927006, 246928001, 246937001, 246938006, 247034002, 247041008, 247043006, 247097006, 247098001, 247099009, 247100001, 247121001, 247124009, 247126006, 247127002, 247131008, 247133006, 247134000, 247137007, 247138002, 247139005, 247140007, 247141006, 247142004, 247146001, 247147005, 247149008, 247155003, 247157006, 247158001, 247159009, 247161000, 247165009, 247167001, 247168006, 247169003, 247170002, 247171003, 247172005, 247173000, 247174006, 247176008, 247177004, 247181004, 247182006, 247183001, 247184007, 247186009, 247187000, 247192003, 247194002, 247198004, 247204001, 247209006, 247210001, 247212009, 247213004, 247219000, 247233000, 248506002, 253176002, 253223002, 253224008, 253225009, 253226005, 253227001, 253231007, 253232000, 253233005, 253234004, 253238001, 253239009, 34623005, 34711008, 35055000, 35472004, 35558006, 35629000, 35691006, 36535003, 36676002, 36794003, 36982001, 36994005, 37075008, 37155002, 37231002, 37565002, 38465001, 38513001, 38579007, 38583007, 38599001, 38742007, 39139002, 39194005, 39302008, 39501002, 39662004, 39832008, 39914007, 39944005, 40024006, 40714009, 40787005, 40933005, 41250005, 41445001, 41564009, 41708006, 41720003, 41799005, 42492000, 43193009, 43300008, 43363007, 43521007, 43645004, 43972005, 44115007, 44219007, 44268007, 44295002, 44381003, 44674000, 45283008, 45623002, 45688009, 45795007, 46085004, 46168003, 46627006, 46674002, 46916006, 47444008, 47673003, 48225000, 48233004, 48249002,

48742005, 49001006, 49089006, 49381001, 50165004, 50455002, 50485007, 50821009, 50975002, 51044000, 51174006, 51226000, 51485001, 51579006, 51886007, 51930008, 51987004, 51995000, 52002008, 52357004, 52421005, 52539004, 52593001, 53667005, 53808001, 53885001, 53889007, 54122009, 54184008, 54359000, 54388004, 55031000, 55713007, 55819001, 55828000, 56039006, 56125009, 56202001, 56211001, 56370005, 56530002, 56921004, 57100005, 57129007, 57138009, 57286009, 57838006, 58749007, 58937003, 59017008, 59044003, 59252009, 59276001, 59412005, 59523007, 59753003, 60189009, 60217008, 60258001, 60505005, 60676002, 61517002, 61536007, 61922005, 62425004, 62660000, 62955002, 62977003, 63251006, 63297008, 63385005, 63511007, 63600008, 63702009, 64015001, 64634000, 64741003, 64959007, 65074000, 65194006, 65460003, 65545003, 65593009, 65720003, 65956007, 66104007, 66281009, 66499004, 66629002, 66725002, 66747002, 67638006, 67841000, 68158006, 68216000, 68478007, 68777001, 69134001, 69278003, 69316005, 69347004, 69811008, 70449006, 70461003, 71044004, 71047006, 71111008, 71676008, 71719003, 72020003, 72124005, 72180004, 72494008, 72620002, 73119000, 73181007, 73221001, 73749009, 73876000, 74460005, 75000006, 75133003, 75229002, 75241009, 75315001, 75663006, 75724008, 76309006, 76562003, 76975009, 77075001, 77103006, 77479002, 77489003, 77628002, 77676001, 77797009, 77873007, 77939001, 78307003, 78348000, 78584008, 78675000, 78768009, 78769001, 79004009, 79410001, 79556007, 80328002, 1217634001, 15685521000119107, 337441000119109, 677711000119105, 397568004, 397572000, 398155003, 398264003, 399120006, 399305009, 399336001, 399463004, 399660006, 399862001, 399863006, 399864000, 399865004, 399866003, 399869005, 399870006, 399872003, 399873008, 399874002, 399875001, 399876000, 399877009, 400962005, 400963000, 400965007, 403435005, 403766000, 404630001, 404633004, 404634005, 404647000, 404648005, 404649002, 404658009, 404663008, 404667009, 404668004, 404669007, 404670008, 404692007, 404694008, 405270006, 405722004, 408311002, 408312009, 408313004, 408409007, 408410002, 408411003, 408412005, 408413000, 408414006, 408847006, 408848001, 408849009, 408850009, 408851008, 410470003, 410471004, 410472006, 410473001, 410476009, 410477000, 410482007, 410484008, 410496001, 410497005, 410498000, 410501000, 410556000, 410568009, 410692006, 413583003, 413665004, 413716003, 413746000, 413851004, 413921009, 413924001, 414173003, 414297000, 414303007, 414491002, 414600000, 414689001, 414783007, 414784001, 414875008, 414894003, 414910007, 414996002, 415176004, 415182001, 415218004, 415344002, 415359008, 415501002, 415584007, 415657000, 415737009, 415843000, 416069001, 416071001, 416165005, 416195002, 416241001, 416296007, 416328002, 416351002, 416449002, 416463008, 416589006, 416633008, 416650005, 416666007, 416703007, 416834004, 416864007, 416877003, 416901002, 416913007, 416960004, 416994002, 417019000, 417020006, 417125003, 417145006, 417183007, 417303004, 417321001, 417344003, 417395001, 417405005, 417544004, 417547006, 417647002, 417651000, 417677008, 417939004, 418054005, 418106004, 418254004, 418402006, 418435001, 418541005, 418572009, 418590005, 418839003, 418871005, 418918009, 418961007, 419039007, 419051009, 419074008, 419096002, 419197009, 419207008, 419237004, 419378004, 419395007, 419459000, 419476001, 419596007, 419648001, 419737003, 419900000, 419982005, 419990005, 420030000, 420183005, 420342002, 420485005, 420486006, 420545001, 420789003, 420805002, 421029004, 421278000, 421293001, 421328004, 421506003, 421689001, 421735009, 421779007, 421973002, 422001004, 422034002, 422061002, 422217003, 422233000, 422338006, 422471006, 423084001, 423248004, 423341008, 423488006, 423673009, 424338005, 424353002, 424358006, 424497004, 424578002, 424669003, 424728002, 424757003, 424909003, 424956000, 425062004, 425217005, 425356002, 425622004, 425961009, 427055009, 428212003, 428674000, 428676003, 429397000, 429449002, 430888006, 432789001, 441622000, 441854007, 443142001, 443201006, 443369007, 443437004, 254092004, 254150007, 254979004, 255012009, 255013004, 255015006, 255016007, 255017003, 255018008, 255019000, 255020006, 255026000, 255027009, 260030003, 262756006, 264443002, 264633009, 267611002, 267612009, 267613004, 267618008, 267619000, 267625001, 267626000, 267638009, 267639001, 267718000, 267727004, 267728009, 268160006, 274571007, 274572000, 275360003,

275477002, 275899002, 276436007, 276549000, 276550000, 278805002, 302174000, 302888003, 302889006, 302890002, 302893000, 302894006, 302895007, 302897004, 302902003, 302904002, 1003408005, 1003424006, 1003425007, 1003428009, 1003488007, 1003489004, 1003490008, 1003491007, 1003530008, 1003532000, 1003534004, 1003535003, 1003605004, 1003614009, 1003615005, 1003616006, 1003617002, 1003657004, 1003884001, 1003921006, 1010610007, 1010664005, 1010666007, 1010668008, 1119266006, 1119281000, 15628611000119101, 15678721000119100, 15678841000119100, 15679281000119103, 15734081000119102, 15991751000119109, 15995551000119104, 16000231000119105, 16000271000119108, 16697421000119103, 16697471000119102, 16697521000119104, 16745051000119107, 16745131000119100, 16745211000119100, 16745411000119104, 16745531000119108, 16745611000119102, 16745651000119101, 16745891000119105, 16745931000119102, 16746131000119105, 16746261000119108, 16746341000119103, 16746661000119106, 16746741000119102, 16746901000119101, 16746941000119104, 16747021000119109, 16747101000119101, 16747141000119104, 16747341000119101, 16747661000119109, 16747741000119100, 16747901000119104, 16748141000119100, 16748741000119101, 16749661000119102, 16749781000119101, 332041000119102, 332361000119101, 332451000119109, 332811000119106, 333741000119105, 334381000119102, 334761000119107, 335021000119100, 337651000119108, 337971000119105, 338061000119101, 338421000119104, 339341000119106, 339981000119100, 340361000119101, 340631000119100, 343421000119107, 343741000119109, 343831000119107, 344191000119100, 345101000119107, 345701000119106, 346071000119107, 368151000119108, 677861000119104, 677881000119108, 678431000119109, 678441000119100, 678451000119103, 679111000119101, 679871000119102, 679881000119104, 679891000119101, 680491000119103, 680501000119105, 680511000119108, 890350009, 890396001, 890423003, 897559009, 897585008, 15989951000119107, 15989911000119106, 817968741000119105, 350781000119106, 1231276000, 354211000119101, 1222546002, 677721000119103, 1217670003, 677731000119100, 1208910001, 1231260009, 1231279007, 1231448006, 1208900005, 15665801000119104, 1231232007, 15986751000119101, 1208895006, 1208890001, 346661000119106, 345161000119108, 15997751000119100, 728667671000119108, 592513751000119104, 345881000119107, 1196922005, 338561000119105, 15663521000119106, 15735001000119106, 1196864009, 544102301000119103, 343431000119105, 344331000119102, 1196923000

ICD-10-CM: H90, H91, Q16, Q17.3, Q17.4, Q17.8, Q17.9, H90.0, H90.1, H90.11, H90.12, H90.2, H90.3, H90.4, H90.41, H90.42, H90.5, H90.6, H90.7, H90.71, H90.72, H90.8, H90.A, H90.A1, H90.A11, H90.A12, H90.A2, H90.A21, H90.A22, H90.A3, H90.A31, H90.A32, H91.0, H91.01, H91.02, H91.03, H91.09, H91.1, H91.10, H91.11, H91.12, H91.13, H91.2, H91.20, H91.21, H91.22, H91.23, H91.3, H91.8, H91.8X, H91.8X1, H91.8X2, H91.8X3, H91.8X9, H91.9, H91.90, H91.91, H91.92, H91.93, Q16.0, Q16.1, Q16.2, Q16.3, Q16.4, Q16.5, Q16.9, E11.3, E11.36, H20, H21, H25, H26, H30, H31, H34, H35, H40, H44, H47.6, H54, S04.0, S04.04, E11.31, E11.311, E11.319, E11.32, E11.321, E11.3211, E11.3212, E11.3213, E11.3219, E11.329, E11.3291, E11.3292, E11.3293, E11.3299, E11.33, E11.331, E11.3311, E11.3312, E11.3313, E11.3319, E11.339, E11.3391, E11.3392, E11.3393, E11.3399, E11.34, E11.341, E11.3411, E11.3412, E11.3413, E11.3419, E11.349, E11.3491, E11.3492, E11.3493, E11.3499, E11.35, E11.351, E11.3511, E11.3512, E11.3513, E11.3519, E11.352, E11.3521, E11.3522, E11.3523, E11.3529, E11.353, E11.3531, E11.3532, E11.3533, E11.3539, E11.354, E11.3541, E11.3542, E11.3543, E11.3549, E11.355, E11.3551, E11.3552, E11.3553, E11.3559, E11.359, E11.3591, E11.3592, E11.3593, E11.3599, E11.37, E11.37X1, E11.37X2, E11.37X3, E11.37X9, E11.39, H20.0, H20.00, H20.01, H20.011, H20.012, H20.013, H20.019, H20.02, H20.021, H20.022, H20.023, H20.029, H20.03, H20.031, H20.032, H20.033, H20.039, H20.04, H20.041, H20.042, H20.043, H20.049, H20.05, H20.051, H20.052, H20.053, H20.059, H20.1, H20.10, H20.11, H20.12, H20.13, H20.2, H20.20, H20.21, H20.22, H20.23, H20.8, H20.81, H20.811, H20.812, H20.813, H20.819, H20.82, H20.821, H20.822, H20.823, H20.829, H20.9, H21.0, H21.00, H21.01, H21.02, H21.03, H21.1, H21.1X, H21.1X1, H21.1X2, H21.1X3, H21.1X9, H21.2, H21.21, H21.211, H21.212, H21.213, H21.219, H21.22, H21.221, H21.222,

H21.223, H21.229, H21.23, H21.231, H21.232, H21.233, H21.239, H21.24, H21.241, H21.242, H21.243, H21.249, H21.25, H21.251, H21.252, H21.253, H21.259, H21.26, H21.261, H21.262, H21.263, H21.269, H21.27, H21.271, H21.272, H21.273, H21.279, H21.29, H21.3, H21.30, H21.301, H21.302, H21.303, H21.309, H21.31, H21.311, H21.312, H21.313, H21.319, H21.32, H21.321, H21.322, H21.323, H21.329, H21.33, H21.331, H21.332, H21.333, H21.339, H21.34, H21.341, H21.342, H21.343, H21.349, H21.35, H21.351, H21.352, H21.353, H21.359, H21.4, H21.40, H21.41, H21.42, H21.43, H21.5, H21.50, H21.501, H21.502, H21.503, H21.509, H21.51, H21.511, H21.512, H21.513, H21.519, H21.52, H21.521, H21.522, H21.523, H21.529, H21.53, H21.531, H21.532, H21.533, H21.539, H21.54, H21.541, H21.542, H21.543, H21.549, H21.55, H21.551, H21.552, H21.553, H21.559, H21.56, H21.561, H21.562, H21.563, H21.569, H21.8, H21.81, H21.82, H21.89, H21.9, H25.0, H25.01, H25.011, H25.012, H25.013, H25.019, H25.03, H25.031, H25.032, H25.033, H25.039, H25.04, H25.041, H25.042, H25.043, H25.049, H25.09, H25.091, H25.092, H25.093, H25.099, H25.1, H25.10, H25.11, H25.12, H25.13, H25.2, H25.20, H25.21, H25.22, H25.23, H25.8, H25.81, H25.811, H25.812, H25.813, H25.819, H25.89, H25.9, H26.0, H26.00, H26.001, H26.002, H26.003, H26.009, H26.01, H26.011, H26.012, H26.013, H26.019, H26.03, H26.031, H26.032, H26.033, H26.039, H26.04, H26.041, H26.042, H26.043, H26.049, H26.05, H26.051, H26.052, H26.053, H26.059, H26.06, H26.061, H26.062, H26.063, H26.069, H26.09, H26.1, H26.10, H26.101, H26.102, H26.103, H26.109, H26.11, H26.111, H26.112, H26.113, H26.119, H26.12, H26.121, H26.122, H26.123, H26.129, H26.13, H26.131, H26.132, H26.133, H26.139, H26.2, H26.20, H26.21, H26.211, H26.212, H26.213, H26.219, H26.22, H26.221, H26.222, H26.223, H26.229, H26.23, H26.231, H26.232, H26.233, H26.239, H26.3, H26.30, H26.31, H26.32, H26.33, H26.4, H26.40, H26.41, H26.411, H26.412, H26.413, H26.419, H26.49, H26.491, H26.492, H26.493, H26.499, H26.8, H26.9, H30.0, H30.00, H30.001, H30.002, H30.003, H30.009, H30.01, H30.011, H30.012, H30.013, H30.019, H30.02, H30.021, H30.022, H30.023, H30.029, H30.03, H30.031, H30.032, H30.033, H30.039, H30.04, H30.041, H30.042, H30.043, H30.049, H30.1, H30.10, H30.101, H30.102, H30.103, H30.109, H30.11, H30.111, H30.112, H30.113, H30.119, H30.12, H30.121, H30.122, H30.123, H30.129, H30.13, H30.131, H30.132, H30.133, H30.139, H30.14, H30.141, H30.142, H30.143, H30.149, H30.2, H30.20, H30.21, H30.22, H30.23, H30.8, H30.81, H30.811, H30.812, H30.813, H30.819, H30.89, H30.891, H30.892, H30.893, H30.899, H30.9, H30.90, H30.91, H30.92, H30.93, H31.0, H31.00, H31.001, H31.002, H31.003, H31.009, H31.01, H31.011, H31.012, H31.013, H31.019, H31.02, H31.021, H31.022, H31.023, H31.029, H31.09, H31.091, H31.092, H31.093, H31.099, H31.1, H31.10, H31.101, H31.102, H31.103, H31.109, H31.11, H31.111, H31.112, H31.113, H31.119, H31.12, H31.121, H31.122, H31.123, H31.129, H31.2, H31.20, H31.21, H31.22, H31.23, H31.29, H31.3, H31.30, H31.301, H31.302, H31.303, H31.309, H31.31, H31.311, H31.312, H31.313, H31.319, H31.32, H31.321, H31.322, H31.323, H31.329, H31.4, H31.40, H31.401, H31.402, H31.403, H31.409, H31.41, H31.411, H31.412, H31.413, H31.419, H31.42, H31.421, H31.422, H31.423, H31.429, H31.8, H31.9, H34.0, H34.00, H34.01, H34.02, H34.03, H34.1, H34.10, H34.11, H34.12, H34.13, H34.2, H34.21, H34.211, H34.212, H34.213, H34.219, H34.23, H34.231, H34.232, H34.233, H34.239, H34.8, H34.81, H34.811, H34.8110, H34.8111, H34.8112, H34.812, H34.8120, H34.8121, H34.8122, H34.813, H34.8130, H34.8131, H34.8132, H34.819, H34.8190, H34.8191, H34.8192, H34.82, H34.821, H34.822, H34.823, H34.829, H34.83, H34.831, H34.8310, H34.8311, H34.8312, H34.832, H34.8320, H34.8321, H34.8322, H34.833, H34.8330, H34.8331, H34.8332, H34.839, H34.8390, H34.8391, H34.8392, H34.9, H35.0, H35.00, H35.01, H35.011, H35.012, H35.013, H35.019, H35.02, H35.021, H35.022, H35.023, H35.029, H35.03, H35.031, H35.032, H35.033, H35.039, H35.04, H35.041, H35.042, H35.043, H35.049, H35.05, H35.051, H35.052, H35.053, H35.059, H35.06, H35.061, H35.062, H35.063, H35.069, H35.07, H35.071, H35.072, H35.073, H35.079, H35.09, H35.1, H35.10, H35.101, H35.102, H35.103, H35.109, H35.11, H35.111, H35.112, H35.113, H35.119, H35.12, H35.121, H35.122, H35.123, H35.129, H35.13, H35.131, H35.132, H35.133, H35.139, H35.14, H35.141,

H35.142, H35.143, H35.149, H35.15, H35.151, H35.152, H35.153, H35.159, H35.16, H35.161, H35.162, H35.163, H35.169, H35.17, H35.171, H35.172, H35.173, H35.179, H35.2, H35.20, H35.21, H35.22, H35.23, H35.3, H35.30, H35.31, H35.311, H35.3110, H35.3111, H35.3112, H35.3113, H35.3114, H35.312, H35.3120, H35.3121, H35.3122, H35.33, H35.3123, H35.3124, H35.313, H35.3130, H35.3131, H35.3132, H35.3133, H35.3134, H35.319, H35.3190, H35.3191, H35.3192, H35.3193, H35.3194, H35.32, H35.321, H35.3210, H35.34, H35.3211, H35.3212, H35.3213, H35.322, H35.3220, H35.3221, H35.3222, H35.3223, H35.323, H35.3230, H35.3231, H35.3232, H35.3233, H35.329, H35.3290, H35.3291, H35.3292, H35.3293, H35.341, H35.342, H35.343, H35.349, H35.35, H35.351, H35.352, H35.353, H35.359, H35.36, H35.361, H35.362, H35.363, H35.369, H35.37, H35.371, H35.372, H35.373, H35.379, H35.38, H35.381, H35.382, H35.383, H35.389, H35.4, H35.40, H35.41, H35.411, H35.412, H35.413, H35.419, H35.42, H35.421, H35.422, H35.423, H35.429, H35.43, H35.431, H35.432, H35.433, H35.439, H35.44, H35.441, H35.442, H35.443, H35.449, H35.45, H35.451, H35.452, H35.453, H35.459, H35.46, H35.461, H35.462, H35.463, H35.469, H35.5, H35.50, H35.51, H35.52, H35.53, H35.54, H35.6, H35.60, H35.61, H35.62, H35.63, H35.7, H35.70, H35.71, H35.711, H35.712, H35.713, H35.719, H35.72, H35.721, H35.722, H35.723, H35.729, H35.73, H35.731, H35.732, H35.733, H35.739, H35.8, H35.81, H35.82, H35.89, H35.9, H40.0, H40.00, H40.001, H40.002, H40.003, H40.009, H40.01, H40.011, H40.012, H40.013, H40.019, H40.02, H40.021, H40.022, H40.023, H40.029, H40.03, H40.031, H40.032, H40.033, H40.039, H40.04, H40.041, H40.042, H40.043, H40.049, H40.05, H40.051, H40.052, H40.053, H40.059, H40.06, H40.061, H40.062, H40.063, H40.069, H40.1, H40.10, H40.10X0, H40.10X1, H40.10X2, H40.10X3, H40.10X4, H40.11, H40.111, H40.1110, H40.1111, H40.1112, H40.1113, H40.1114, H40.112, H40.1120, H40.1121, H40.1122, H40.1123, H40.1124, H40.113, H40.1130, H40.1131, H40.1132, H40.1133, H40.1134, H40.119, H40.1190, H40.1191, H40.1192, H40.1193, H40.1194, H40.12, H40.121, H40.1210, H40.1211, H40.1212, H40.1213, H40.1214, H40.122, H40.1220, H40.1221, H40.1222, H40.1223, H40.1224, H40.123, H40.1230, H40.1231, H40.1232, H40.1233, H40.1234, H40.129, H40.1290, H40.1291, H40.1292, H40.1293, H40.1294, H40.13, H40.131, H40.1310, H40.1311, H40.1312, H40.1313, H40.1314, H40.132, H40.1320, H40.1321, H40.1322, H40.1323, H40.1324, H40.133, H40.1330, H40.1331, H40.1332, H40.1333, H40.1334, H40.139, H40.1390, H40.1391, H40.1392, H40.1393, H40.1394, H40.14, H40.141, H40.1410, H40.1411, H40.1412, H40.1413, H40.1414, H40.142, H40.1420, H40.1421, H40.1422, H40.1423, H40.1424, H40.143, H40.1430, H40.1431, H40.1432, H40.1433, H40.1434, H40.149, H40.1490, H40.1491, H40.1492, H40.1493, H40.1494, H40.15, H40.151, H40.152, H40.153, H40.159, H40.2, H40.20, H40.20X0, H40.20X1, H40.20X2, H40.20X3, H40.20X4, H40.21, H40.211, H40.212, H40.213, H40.219, H40.22, H40.221, H40.2210, H40.2211, H40.2212, H40.2213, H40.2214, H40.222, H40.2220, H40.2221, H40.2222, H40.2223, H40.2224, H40.223, H40.2230, H40.2231, H40.2232, H40.2233, H40.2234, H40.229, H40.2290, H40.2291, H40.2292, H40.2293, H40.2294, H40.23, H40.231, H40.232, H40.233, H40.239, H40.24, H40.241, H40.242, H40.243, H40.249, H40.3, H40.30, H40.30X0, H40.30X1, H40.30X2, H40.30X3, H40.30X4, H40.31, H40.31X0, H40.31X1, H40.31X2, H40.31X3, H40.31X4, H40.32, H40.32X0, H40.32X1, H40.32X2, H40.32X3, H40.32X4, H40.33, H40.33X0, H40.33X1, H40.33X2, H40.33X3, H40.33X4, H40.4, H40.40, H40.40X0, H40.40X1, H40.40X2, H40.40X3, H40.40X4, H40.41, H40.41X0, H40.41X1, H40.41X2, H40.41X3, H40.41X4, H40.42, H40.42X0, H40.42X1, H40.42X2, H40.42X3, H40.42X4, H40.43, H40.43X0, H40.43X1, H40.43X2, H40.43X3, H40.43X4, H40.5, H40.50, H40.50X0, H40.50X1, H40.50X2, H40.50X3, H40.50X4, H40.51, H40.51X0, H40.51X1, H40.51X2, H40.51X3, H40.51X4, H40.52, H40.52X0, H40.52X1, H40.52X2, H40.52X3, H40.52X4, H40.53, H40.53X0, H40.53X1, H40.53X2, H40.53X3, H40.53X4, H40.6, H40.60, H40.60X0, H40.60X1, H40.60X2, H40.60X3, H40.60X4, H40.61, H40.61X0, H40.61X1, H40.61X2, H40.61X3, H40.61X4, H40.62, H40.62X0, H40.62X1, H40.62X2, H40.62X3, H40.62X4, H40.63, H40.63X0, H40.63X1, H40.63X2, H40.63X3, H40.63X4, H40.8, H40.81, H40.811, H40.812, H40.813, H40.819,

H40.82, H40.821, H40.822, H40.823, H40.829, H40.83, H40.831, H40.832, H40.833, H40.839, H40.89, H40.9, H44.0, H44.00, H44.001, H44.002, H44.003, H44.009, H44.01, H44.011, H44.012, H44.013, H44.019, H44.02, H44.021, H44.022, H44.023, H44.029, H44.1, H44.11, H44.111, H44.112, H44.113, H44.119, H44.12, H44.121, H44.122, H44.123, H44.129, H44.13, H44.131, H44.132, H44.133, H44.139, H44.19, H44.2, H44.20, H44.21, H44.22, H44.23, H44.2A, H44.2A1, H44.2A2, H44.2A3, H44.2A9, H44.2B, H44.2B1, H44.2B2, H44.2B3, H44.2B9, H44.2C, H44.2C1, H44.2C2, H44.2C3, H44.2C9, H44.2D, H44.2D1, H44.2D2, H44.2D3, H44.2D9, H44.2E, H44.2E1, H44.2E2, H44.2E3, H44.2E9, H44.3, H44.30, H44.31, H44.311, H44.312, H44.313, H44.319, H44.32, H44.321, H44.322, H44.323, H44.329, H44.39, H44.391, H44.392, H44.393, H44.399, H44.4, H44.40, H44.41, H44.411, H44.412, H44.413, H44.419, H44.42, H44.421, H44.422, H44.423, H44.429, H44.43, H44.431, H44.432, H44.433, H44.439, H44.44, H44.441, H44.442, H44.443, H44.449, H44.5, H44.50, H44.51, H44.511, H44.512, H44.513, H44.519, H44.52, H44.521, H44.522, H44.523, H44.529, H44.53, H44.531, H44.532, H44.533, H44.539, H44.6, H44.60, H44.601, H44.602, H44.603, H44.609, H44.61, H44.611, H44.612, H44.613, H44.619, H44.62, H44.621, H44.622, H44.623, H44.629, H44.63, H44.631, H44.632, H44.633, H44.639, H44.64, H44.641, H44.642, H44.643, H44.649, H44.65, H44.651, H44.652, H44.653, H44.659, H44.69, H44.691, H44.692, H44.693, H44.699, H44.7, H44.70, H44.701, H44.702, H44.703, H44.709, H44.71, H44.711, H44.712, H44.713, H44.719, H44.72, H44.721, H44.722, H44.723, H44.729, H44.73, H44.731, H44.732, H44.733, H44.739, H44.74, H44.741, H44.742, H44.743, H44.749, H44.75, H44.751, H44.752, H44.753, H44.759, H44.79, H44.791, H44.792, H44.793, H44.799, H44.8, H44.81, H44.811, H44.812, H44.813, H44.819, H44.82, H44.821, H44.822, H44.823, H44.829, H44.89, H44.9, H47.61, H47.611, H47.612, H47.619, H47.62, H47.621, H47.622, H47.629, H47.63, H47.631, H47.632, H47.639, H47.64, H47.641, H47.642, H47.649, H54.0, H54.0X, H54.0X3, H54.0X33, H54.0X34, H54.0X35, H54.0X4, H54.0X43, H54.0X44, H54.0X45, H54.0X5, H54.0X53, H54.0X54, H54.0X55, H54.1, H54.10, H54.11, H54.113, H54.1131, H54.1132, H54.114, H54.1141, H54.1142, H54.115, H54.1151, H54.1152, H54.12, H54.121, H54.1213, H54.1214, H54.1215, H54.122, H54.1223, H54.1224, H54.1225, H54.2, H54.2X, H54.2X1, H54.2X11, H54.2X12, H54.2X2, H54.2X21, H54.2X22, H54.3, H54.4, H54.40, H54.41, H54.413, H54.413A, H54.414, H54.414A, H54.415, H54.415A, H54.42, H54.42A, H54.42A3, H54.42A4, H54.42A5, H54.5, H54.50, H54.51, H54.511, H54.511A, H54.512A, H54.52, H54.52A, H54.52A1, H54.52A2, H54.6, H54.60, H54.61, H54.62, H54.7, H54.8, S04.01, S04.011, S04.011A, S04.011D, S04.011S, S04.012, S04.012A, S04.012D, S04.012S, S04.019, S04.019A, S04.019D, S04.019S, S04.02, S04.02XA, S04.02XD, S04.02XS, S04.03, S04.031, S04.031A, S04.031D, S04.031S, S04.032, S04.032A, S04.032D, S04.032S, S04.039, S04.039A, S04.039D, S04.039S, S04.041, S04.041A, S04.041D, S04.041S, S04.042, S04.042A, S04.042D, S04.042S, S04.049, S04.049A, S04.049D, S04.049S

ICD-9-CM: 389, 744.0, 744.21, 744.24, 744.29, 744.3, 360, 362, 363, 364, 365, 366, 369, 377.7, 950

#### Congenital heart disease

ICD-10-CM: Q21.22, Q21.20, Q21.21, Q21.23, Q20, Q20.1, Q20.2, Q20.3, Q20.4, Q20.5, Q20.6, Q20.8, Q20.9, Q21, Q21.0, Q21.1, Q21.2, Q21.3, Q21.4, Q21.8, Q21.9, Q22, Q22.1, Q22.2, Q22.4, Q22.5, Q22.8, Q22.9, Q23, Q23.0, Q23.1, Q23.2, Q23.3, Q23.4, Q23.8, Q23.9, Q24, Q24.0, Q24.3, Q24.4, Q24.5, Q24.6, Q24.8, Q24.9, Q20.0, Q22.0, Q22.3, Q22.6, Q24.1, Q24.2

ICD-9-CM: 745, 746

SNOMED CT: 4374004, 7368005, 7484005, 13213009, 15096009, 17024001, 18546004, 27637000, 28574005, 28656008, 29928006, 30288003, 82458004, 86299006, 204357006, 204296002, 204370002, 253277003, 253281003, 253335001, 253414002, 46619002, 59494005, 62067003, 67278007, 70142008, 70320004, 73660006, 1204130006, 1230303001, 1229876001, 1230344000, 1222710008, 1222708006, 1208998007, 1162915007, 1187212004, 1163259003, 1187113001, 1186709006, 1131009, 2583009, 4313003, 5230009, 6996004, 8239009, 8712002, 10818008, 11433004, 12770006, 13453009, 13689005, 14886009, 15459006, 16567006, 17718000, 19249002, 21981000, 23063005, 26201005, 29375001, 29934004, 81990004, 83119008, 83799000, 84648007, 85081000, 88445007, 89222008, 91634006, 92830003, 92888002, 92923008, 92941000, 92944008, 92960007, 92963009, 92969008, 92974000, 92976003, 93051009, 93053007, 93069000, 93075009, 93076005, 93078006, 93234006, 93237004, 93247001, 93262004, 93271008, 93287006, 94150003, 94702005, 94703000, 94706008, 95268002, 109379000, 109425008, 109426009, 109427000, 109428005, 109429002, 109430007, 109431006, 109432004, 111319002, 123656005, 123657001, 123658006, 123659003, 123660008, 128555001, 128556000, 128557009, 128558004, 128563000, 128567004, 128568009, 129582000, 1141882007, 788533006, 827078006, 838333005, 840469002, 840488009, 840495000, 840497008, 840507004, 840517009, 866118008, 871579003, 871581001, 871583003, 871585005, 871594004, 871597006, 871599009, 871600007, 871601006, 871602004, 871603009, 871604003, 871605002, 871606001, 871607005, 871609008, 871610003, 871611004, 871612006, 871613001, 871616009, 871618005, 871620008, 871622000, 871624004, 871625003, 871627006, 871630004, 871636005, 871643004, 871645006, 871647003, 871648008, 871650000, 871652008, 871654009, 871655005, 871658007, 871659004, 871660009, 871666003, 871668002, 871672003, 41371000119100, 63191000119107, 15964981000119104, 2829000, 17394001, 19092004, 26146002, 31080005, 42402006, 63247009, 80387009, 204311009, 204319006, 204351007, 253468007, 253496001, 253514004, 253515003, 422348008, 444851003, 444961009, 445001003, 445002005, 445003000, 445131007, 445235006, 445264002, 445268004, 445270008, 445298006, 445299003, 445310002, 445330003, 445334007, 445373007, 445435009, 445453003, 445454009, 445607003, 445636003, 445650008, 445898001, 445928005, 446326008, 446657003, 446659000, 446667008, 446670007, 446909006, 446916007, 447283008, 447284002, 447285001, 447286000, 447289007, 447661004, 447665008, 447666009, 447667000, 447669002, 447670001, 447671002, 447672009, 447673004, 447681003, 447683000, 447689001, 447690005, 447696004, 447697008, 447698003, 447700007, 447701006, 447702004, 447773008, 447774002, 447780005, 447822009, 447824005, 447825006, 447829000, 447830005, 447838003, 447839006, 447845003, 447852001, 447874007, 447875008, 447876009, 447901004, 447913009, 447915002, 447919008, 447932003, 447933008, 447937009, 447938004, 447941008, 447962009, 447988007, 447998001, 447999009, 448001004, 448004007, 448007000, 448028009, 448062009, 448063004, 448064005, 448066007, 448072007, 448073002, 448074008, 448075009, 448080000, 448081001, 448082008, 448083003, 448085005, 448095003, 448103004, 448113007, 448118003, 448119006, 448120000, 448155003, 448161000, 448162007, 448181004, 448182006, 448183001, 448184007, 448208003, 448277007, 448278002, 448280008, 448305002, 448308000, 448309008, 448310003, 448320008, 448331000, 448356006, 448357002, 448413002, 448414008, 448415009, 448475002, 448477005, 448478000, 448497000, 448516008, 448574001, 448575000, 448604006, 448612003, 448619007, 448624005, 448625006, 448633007, 448634001, 448681000, 448729006, 448747000, 448778001, 448780007, 448786001, 448790004, 448793002, 448794008, 448809003, 448819009, 448823001, 448827000, 448842002, 448843007, 448844001, 448876006, 448887003, 448919005, 448920004, 448923002, 448946000, 448947009, 448948004, 448966009, 448975006, 449005003, 449014008, 449015009, 449016005, 449029005, 449031001, 449032008, 449037002, 449040002, 449086000, 449087009, 449098005, 449099002, 449110001, 449118008, 449123008, 449124002, 449133000, 449135007, 449138009, 449139001, 449140004, 449158005, 449178002, 449190000, 449228007, 449270002, 449315003, 449316002, 449351005, 449352003, 449353008, 449354002, 449379004, 449382009, 449383004, 449385006, 449398002, 449428009, 449429001, 449436000, 449440009, 449441008, 449442001, 449455000, 449456004, 449457008, 449458003, 449459006, 449479003,

449490008, 449492000, 449511008, 449512001, 449520004, 449523002, 449532000, 449534004, 449537006, 449538001, 449539009, 449563001, 449567000, 449568005, 449587004, 449593007, 449594001, 449595000, 449596004, 449599006, 449600009, 450300002, 450301003, 450302005, 458039003, 459062008, 459065005, 459066006, 459173004, 460437005, 460438000, 460471001, 460510005, 460517008, 460524009, 460531008, 460538002, 460545002, 460581004, 460582006, 460583001, 460584007, 460585008, 460586009, 460587000, 460588005, 461345005, 460614005, 460913001, 460923005, 460930004, 460937001, 460944005, 461104009, 461105005, 461107002, 461109004, 461110009, 461111008, 461112001, 461331004, 461359003, 461381008, 461382001, 461383006, 461384000, 461385004, 461420008, 461421007, 461435009, 461436005, 461438006, 461439003, 471276003, 471277007, 471278002, 471279005, 471280008, 471285003, 471286002, 471287006, 471288001, 471289009, 471290000, 471291001, 471292008, 471293003, 471297002, 471298007, 471299004, 471301006, 472703008, 472769009, 472785004, 472786003, 472787007, 472794005, 472795006, 472796007, 472797003, 472799000, 472800001, 472801002, 472802009, 472807003, 472820000, 473362006, 473393007, 473443007, 473444001, 698971006, 715535009, 715987000, 716193004, 716740009, 716773002, 717812000, 717859007, 717943008, 718128009, 718135001, 718181001, 718216009, 718556007, 718681002, 718713000, 719379001, 719395001, 719400000, 719451006, 719456001, 720567008, 720605009, 720606005, 720610008, 720612000, 720639008, 720748007, 720858001, 721009008, 721010003, 721013001, 721014007, 721015008, 721073008, 721976003, 721978002, 722027009, 722051004, 722206009, 722461004, 723089002, 723304001, 723333000, 723336008, 723448007, 724066002, 725145002, 726083008, 726335002, 726704006, 733491005, 735571003, 735579001, 735580003, 735581004, 762251008, 762252001, 762253006, 762254000, 762433009, 762460002, 763066009, 763279007, 763747002, 763778003, 763834000, 763835004, 764697003, 764955006, 766881008, 766976003, 768552007, 770432008, 770433003, 770435005, 773139006, 773749003, 780842009, 781159007, 782698000, 782699008, 783011004, 783096008, 783738002, 783773000, 784353002, 194863005, 204300001, 204306007, 204312002, 204315000, 204317008, 204318003, 204330009, 204339005, 204342004, 204345002, 204346001, 204352000, 204354004, 204362007, 204363002, 204378009, 204379001, 204394002, 204395001, 204397009, 204398004, 204399007, 204448004, 205768003, 205834002, 360473004, 360481003, 373131000, 373905003, 233846000, 233862006, 234132006, 236529001, 236530006, 237227006, 250941001, 250942008, 250983006, 253264007, 253267000, 253268005, 253269002, 253270001, 253271002, 253272009, 253274005, 253275006, 253276007, 253278008, 253279000, 253280002, 253282005, 253283000, 253284006, 253285007, 253286008, 253287004, 253288009, 253289001, 253290005, 253291009, 253293007, 253294001, 253295000, 253297008, 253298003, 253299006, 253300003, 253301004, 253302006, 253303001, 253304007, 253305008, 253324006, 253326008, 253327004, 253333008, 253336000, 253337009, 253338004, 253341008, 253342001, 253343006, 253345004, 253346003, 253348002, 253349005, 253353007, 253354001, 253356004, 253357008, 253359006, 253360001, 253364005, 253366007, 253369000, 253370004, 253371000, 253373002, 253374008, 253376005, 253377001, 253378006, 253379003, 253381001, 253382008, 253384009, 253391007, 253392000, 253393005, 253394004, 253395003, 253396002, 253397006, 253399009, 253403000, 253404006, 253405007, 253411005, 253412003, 253413008, 253415001, 253416000, 253417009, 253418004, 253419007, 253420001, 253421002, 253422009, 253423004, 253425006, 253426007, 253439006, 253441007, 253450009, 253451008, 253458002, 253465005, 253478005, 253479002, 253480004, 253481000, 253482007, 253483002, 253484008, 253485009, 253486005, 253487001, 253489003, 253492004, 253493009, 253494003, 253495002, 253497005, 253498000, 253499008, 253500004, 253501000, 253502007, 253503002, 253504008, 253505009, 253506005, 253507001, 253508006, 253509003, 253510008, 253511007, 253512000, 253513005, 253518001, 253519009, 253520003, 253521004, 253523001, 253524007, 253525008, 253527000, 253529002, 253530007, 253531006, 253532004, 253533009, 253534003, 253536001, 253537005, 253538000, 253539008, 253540005, 253542002, 253543007, 253544001, 253545000, 253546004, 253547008, 253548003, 253550006, 253551005, 253552003, 253553008, 253554002, 253555001, 253556000, 253559007, 253562005, 253563000, 253564006, 253565007, 253566008, 253567004, 253568009, 253569001, 253570000, 253571001, 253573003, 253574009,

253575005, 253576006, 253578007, 253580001, 253584005, 253585006, 253586007, 253587003, 253588008, 253591008, 253593006, 253596003, 253597007, 253598002, 253599005, 253600008, 253601007, 253602000, 253603005, 253607006, 253608001, 253609009, 253610004, 253611000, 253624000, 253625004, 253650001, 253700001, 253703004, 253704005, 253706007, 253707003, 253708008, 253710005, 253711009, 253712002, 253714001, 253715000, 253716004, 253717008, 253718003, 253719006, 253720000, 253725005, 253728007, 253729004, 253730009, 253732001, 35577008, 36079008, 36133000, 36233006, 37104009, 37639005, 39589002, 40272001, 41049003, 41514002, 41893002, 43176009, 45503006, 48121000, 48520006, 51442005, 51789008, 54073003, 55510008, 55546004, 56309007, 59554006, 60106004, 60232001, 60732002, 61959006, 63042009, 64862009, 72252007, 72352009, 73699003, 74034002, 74218008, 75270000, 75372006, 75398000, 76257003, 77696009, 78196008, 78250005, 78320000, 78485007, 78495000, 399046008, 399216004, 399228007, 405752007, 412787009, 413905004, 424045003, 443379009, 263944006, 263961009, 264086008, 264162009, 264467005, 264571006, 268174004, 268180007, 270510008, 274947007, 302943003, 876846006, 880003006, 880062003, 890220003, 890393009, 890402009

#### Preexisting anemias

ICD-10-CM: D50, D55, D56, D57.1, D57.20, D57.3, D57.40, D57.80, D58, D59, O99.01, O99.02, D55.29, D55.21, D50.0, D50.1, D50.8, D50.9, D55.0, D55.1, D55.2, D55.3, D55.8, D55.9, D56.0, D56.1, D56.2, D56.3, D56.4, D56.5, D56.8, D56.9, D58.0, D58.1, D58.2, D58.8, D58.9, D59.0, D59.1, D59.2, D59.3, D59.4, D59.5, D59.6, D59.8, D59.9, O99.011, O99.012, O99.013, O99.019, D59.10, D59.11, D59.12, D59.13, D59.19

SNOMED CT: 4854004, 16402000, 27342004, 87522002, 111577008, 10741751000119100, 38911009, 40108008, 416180004, 417048006, 417683006, 934007, 3978000, 4939006, 5300004, 5315003, 5994005, 6398009, 8857001, 10564005, 11781007, 13172003, 15121005, 15276008, 16360009, 16427007, 16964007, 18323000, 19442009, 22098000, 23371004, 24975009, 25251008, 25266006, 26682008, 29177004, 32648007, 32855007, 33183004, 33905008, 34194007, 81711008, 84188003, 85557000, 86225009, 86715000, 89459006, 89810003, 91217009, 111469006, 111578003, 111579006, 127041004, 127042006, 127043001, 127044007, 127045008, 127046009, 127049002, 127050002, 127052005, 127053000, 127055007, 127056008, 127057004, 127058009, 127060006, 127061005, 127319005, 1142030006, 1142062009, 1142083003, 1148853004, 1148855006, 1148856007, 1148861009, 1148880006, 1148893001, 1148895008, 1148897000, 1148899002, 1153358006, 1153399000, 1153403009, 788944005, 863953007, 72321000119107, 72331000119105, 119641000119103, 119691000119106, 119701000119106, 350601000119109, 10759401000119105, 10806241000119108, 10812081000119108, 5603006, 5967006, 15326009, 22933009, 24661004, 27080008, 28147001, 32858009, 33491002, 34852006, 39586009, 47084006, 56205004, 62609001, 69216008, 71832003, 73190000, 76366001, 80126007, 80963002, 82003006, 85422000, 86242003, 86859003, 86986002, 91411007, 111572002, 127054006, 240453002, 387705004, 421851008, 447117006, 713533000, 715342005, 716682000, 717948004, 718196002, 718716008, 719402008, 720982007, 722005000, 724556004, 724557008, 725057008, 732960002, 732962005, 732963000, 732965007, 732966008, 734349003, 766982000, 774083009, 191128004, 191169008, 191170009, 191178002, 191179005, 191187006, 191189009, 191210005, 191211009, 191212002, 191213007, 191216004, 191217008, 191218003, 191222008, 191228007, 191229004, 199248002, 307333004, 307334005, 307335006, 307336007, 307337003, 307338008, 307343001, 309742004, 363041004, 371315009, 373420004, 234380002, 234381003, 234383000, 234385007, 234386008, 234387004, 234388009, 234389001, 234390005, 234392002, 234410008, 35703006, 36467003, 36472007, 36568005, 38689004, 41387000, 42484009, 44206008, 44641000, 47047009, 47516005, 47526003, 48553001, 50253007, 52212006, 52413004, 54006005, 55995005, 59644002, 61395005, 61777009, 62074008,

62268000, 62403005, 62871001, 65959000, 66055002, 66262001, 68361004, 68913001, 69981004, 73073009, 73162004, 74703006, 75331009, 75443009, 75451007, 76336008, 77413008, 77663007, 78129009, 78908001, 79592006, 398937006, 402653004, 402654005, 413603009, 416826005, 417626001, 438476003, 439007008, 441322009, 269175006, 273985002, 276580005, 290246007, 301317008, 302873008, 350611000119107

ICD-9-CM: 280, 282, 282.3, 282.4, 282.41, 282.5, 282.61, 282.63, 282.68, 283, 648.24

#### Placenta accreta spectrum

ICD-10-CM: O43.2, O43.21, O43.211, O43.212, O43.213, O43.219, O43.22, O43.221, O43.222, O43.223, O43.229, O43.23, O43.231, O43.232, O43.233, O43.239

SNOMED CT: 237296008, 11763009, 25585008, 33058001, 1230040001, 237297004, 70129008

ICD-9-CM: 667

#### Chronic renal disease

ICD-10-CM: I12, I12.9, I13, N03, N04, N05, N07, N08, N11.1, N11.8, N11.9, N18, N25.0, N25.1, N25.81, N25.89, N25.9, N26.9, O26.83, I12.0, I13.0, I13.1, I13.10, I13.11, I13.2, N03.0, N03.1, N03.2, N03.3, N03.4, N03.5, N03.6, N03.7, N03.8, N03.9, N04.0, N04.1, N04.2, N04.3, N04.4, N04.5, N04.6, N04.7, N04.8, N04.9, N05.0, N05.1, N05.2, N05.3, N05.4, N05.5, N05.6, N05.7, N05.8, N05.9, N07.0, N07.1, N07.2, N07.3, N07.4, N07.5, N07.6, N07.7, N07.8, N07.9, N18.1, N18.2, N18.3, N18.4, N18.5, N18.6, N18.9, O26.831, O26.832, O26.833, O26.839, N03.A, N04.A, N05.A, N07.A, N18.30, N18.31, N18.32

ICD-9-CM: 403, 404, 581, 582, 583, 585, 587, 588, 646.2, V13.03

SNOMED CT: 7724006, 16726004, 19034001, 32916005, 90708001, 111395007, 8501000119104, 104931000119100, 367591000119105, 709044004, 197663003, 197679002, 197707007, 236379002, 52254009, 60926001, 1187124004, 1177174007, 1187460003, 1216942009, 1220648003, 1220586000, 145781000119106, 1208934006, 1208341008, 1162265009, 11801451000119106, 1187557007, 1163491000, 1187554000, 1187555004, 1187553006, 1163484003, 1187507004, 1187546002, 11801411000119105, 1169358003, 1187464007, 1162266005, 1187040004, 1426004, 1474004, 1592005, 3321001, 3704008, 193003, 290006, 3999002, 4390004, 4451004, 4495005, 4576001, 4676006, 5397007, 7448003, 7703001, 7725007, 8199003, 8468007, 8996006, 9264002, 9298006, 9918001, 11013005, 11480007, 11659006, 12897005, 13335004, 13530005, 13886001, 14178006, 14343001, 14669001, 14973001, 15123008, 16297002, 16507009, 16652001, 16751003, 16756008, 17121006, 17380002, 17901006, 18417009, 19351000, 20341008, 20483002, 20917003, 21764004, 21952001, 22011005, 22352007, 22702000, 22846003, 22966008, 23697004, 23754003, 24790002, 25646005, 25765006, 25821008, 26121002, 26235008, 26367008, 27174002, 27810000, 28196006, 28637003, 28770003, 29908007, 30124006, 30275001, 30295007, 31005002, 31881008, 32093003, 32278006, 32659003, 32801008, 33561005, 33763006, 34165000, 80902009, 81363003, 81475007, 81896006, 81986001, 82112005, 82525005, 83563007, 83850008, 83866005, 83923004, 84121007, 85020001, 85487008, 85495007, 85901000, 86234004, 86249007, 86463003, 88102009, 88380005, 88531004, 89637003, 90241004, 90493000,

90688005, 91003006, 91195006, 92165001, 92319008, 92624000, 92697000, 92921005, 92975004, 93290000, 93425004, 93849006, 93985007, 94360002, 94514000, 94889006, 95025009, 95568003, 95570007, 95571006, 95575002, 95577005, 95581005, 95582003, 102803006, 105999006, 106000008, 109477002, 111403005, 111406002, 111407006, 112066009, 118951003, 123609007, 123610002, 123612005, 123752003, 123753008, 124147007, 126880001, 126881002, 127013003, 129128006, 1079531000119106, 1079541000119102, 1080561000119103, 1080611000119100, 1137561000, 1142048002, 1142097006, 1144897006, 1144899009, 1144903002, 1144909003, 1144941009, 1144960002, 1144961003, 1144972008, 1144973003, 1144975005, 1144979004, 1148846008, 1148852009, 1148867008, 1148869006, 1148871006, 1148873009, 1148876001, 1148881005, 1148882003, 1148883008, 1148884002, 1148885001, 1148887009, 1148888004, 1148903009, 1148908000, 1148909008, 1148915008, 1148919002, 1148921007, 1148923005, 1148931000, 1148933002, 1148935009, 1148937001, 1148939003, 1155732005, 1157178009, 1224648100119106, 15957661000119104, 15958021000119103, 15958101000119106, 15958181000119103, 15958221000119106, 15958261000119101, 15958301000119109, 15958381000119101, 15958661000119103, 15958701000119105, 15958741000119107, 15959261000119108, 16067811000119101, 16067851000119100, 16697291000119100, 352961000119103, 353111000119108, 763101411000119100, 894171571000119107, 788613004, 789660001, 792925006, 792926007, 816069008, 818952002, 829971006, 833291009, 833293007, 836486002, 838353009, 840638005, 860892003, 860893008, 870589006, 711000119100, 721000119107, 731000119105, 741000119101, 751000119104, 771000119108, 1801000119106, 4181000119109, 5941000119101, 7011000119105, 15781000119107, 15911000119103, 18521000119106, 28191000119109, 40371000119105, 40391000119106, 42231000009109, 50411000119108, 71421000119105, 71441000119104, 71701000119105, 71721000119101, 74011000119107, 76401000119105, 85381000119105, 89681000119101, 90721000119101, 90731000119103, 90741000119107, 90751000119109, 90761000119106, 90771000119100, 90781000119102, 90791000119104, 90971000119103, 94181000119108, 96441000119101, 96701000119107, 96711000119105, 96721000119103, 96731000119100, 96741000119109, 96751000119106, 98371000119102, 101711000119105, 106911000119102, 108101000119101, 108131000119108, 108201000119105, 108211000119108, 111411000119103, 111631000119106, 117681000119102, 118781000119108, 120241000119100, 120261000119101, 126551000119103, 127901000119107, 127911000119105, 127991000119101, 128001000119105, 129151000119102, 129161000119100, 129171000119106, 129181000119109, 129561000119108, 129721000119106, 130181000119100, 140031000119103, 140101000119109, 140111000119107, 140121000119100, 140131000119102, 145681000119101, 153851000119106, 153891000119101, 157141000119108, 195791000119101, 243421000119104, 284961000119106, 284971000119100, 284981000119102, 284991000119104, 285001000119105, 285011000119108, 285041000119107, 285061000119106, 285081000119102, 285101000119109, 285831000119108, 285841000119104, 285851000119102, 285861000119100, 285871000119106, 285881000119109, 285911000119109, 285921000119102, 286371000119107, 295101000119105, 295111000119108, 295121000119101, 308751000119106, 320681000119102, 351421000119105, 351611000119101, 353911000119105, 353961000119108, 354021000119100, 354121000119104, 354351000119105, 354361000119107, 367051000119105, 367481000119108, 367511000119101, 367521000119108, 367531000119106, 367541000119102, 367551000119100, 367561000119103, 367571000119109, 367581000119107, 367611000119100, 367621000119107, 367821000119106, 368871000119106, 368881000119109, 368901000119106, 368911000119109, 368921000119102, 368931000119104, 368941000119108, 368951000119105, 368961000119107, 369071000119105, 429691000124100, 429701000124100, 429711000124102, 429721000124105, 429731000124108, 431431000124100, 434431000124103, 449631000124102, 691421000119108, 1056501000112102, 1078991000119106, 1079001000119106, 1081231000119103, 1081241000119107, 1081571000119102, 1081581000119104, 1081801000112104, 1082281000119105, 1082301000119109, 1082321000119100, 1082331000119102, 1086071000119103, 1087041000119107, 1092831000119109, 10752771000119100, 10757401000119104, 10757481000119107, 11798411000119101, 11798461000119103, 11800851000119104, 11800891000119109, 11801171000119107, 11801251000119105, 14350001000004108, 15636991000119103, 15638291000119105, 15648731000119101, 15648771000119103, 15648821000119105,

15959061000119101, 15959101000119103, 16218721000119107, 16705321000119109, 16705361000119104, 18300001000004102, 22470001000004107, 1776003, 13889008, 24814002, 44785005, 50581000, 55652009, 59479006, 65520001, 68779003, 75888001, 79385002, 81987005, 204949001, 236385009, 236407003, 236411009, 236506009, 236522005, 282364005, 422593004, 707742001, 723720008, 783787000, 444558002, 444645005, 444691002, 444749006, 444794000, 444830001, 444976001, 444977005, 445119005, 445120004, 445170001, 445236007, 445258009, 445388002, 445404003, 446449009, 446923008, 446989009, 446991001, 447335007, 448215006, 449408005, 449409002, 449820008, 473392002, 609452007, 609455009, 609472002, 609482001, 609490001, 698591006, 698757009, 699055002, 699322002, 700107006, 700109009, 700111000, 700112007, 700378005, 700379002, 702391001, 702397002, 702718005, 703310005, 704055002, 704203009, 704204003, 704205002, 704206001, 704667004, 705065000, 707148007, 707208009, 707221002, 707324008, 707332000, 707530009, 707751009, 707755000, 707756004, 707770004, 708127008, 708975004, 709978007, 710565001, 710571007, 711531007, 712487000, 713313000, 713453003, 713504001, 713574000, 713695001, 713696000, 713825007, 713886006, 713887002, 714152005, 714153000, 714813000, 714814006, 714815007, 714816008, 714817004, 714818009, 716094008, 716200002, 716657000, 716864001, 716999001, 717187000, 717191005, 717263009, 717734005, 717735006, 717736007, 717742006, 717744007, 717745008, 717746009, 717748005, 717749002, 717750002, 717760006, 717766000, 717767009, 717768004, 717770008, 717776002, 717789008, 717790004, 717791000, 717952004, 718141008, 718192000, 719839000, 719840003, 720414005, 720415006, 720458005, 720519003, 720982007, 721167009, 721168004, 721169007, 721170008, 721173005, 721175003, 721191000, 721207002, 721297008, 721840000, 721862000, 721970009, 722065002, 722077007, 722078002, 722081007, 722082000, 722085003, 722086002, 722088001, 722089009, 722095005, 722096006, 722098007, 722118005, 722119002, 722120008, 722139003, 722147003, 722149000, 722150000, 722168002, 722223000, 722231005, 722278006, 722294004, 722369003, 722381004, 722431007, 722433005, 722457005, 722466009, 722467000, 722468005, 722721004, 722758004, 722759007, 722760002, 722761003, 722948009, 723074006, 723188008, 723189000, 723190009, 723333000, 723363009, 723373006, 723409007, 723440000, 723449004, 723555007, 723995003, 723999009, 724093004, 724094005, 724099000, 724282009, 724469003, 724485008, 725033008, 725592009, 725905005, 725908007, 726017001, 726018006, 726082003, 726106004, 726541005, 733089005, 733096007, 733097003, 733116005, 733137002, 733138007, 733139004, 733342000, 733453005, 733470002, 733471003, 733472005, 733603009, 733608000, 733650000, 733839001, 734015000, 734990008, 735430007, 735471001, 735474009, 735475005, 736450002, 736451003, 736640009, 736992003, 736993008, 737327002, 737562008, 762288006, 762531008, 762907005, 762908000, 762909008, 762910003, 762911004, 762912006, 762913001, 762914007, 763280005, 763891005, 764694005, 764856008, 764961009, 764990003, 765095002, 765154003, 765155002, 765330003, 765331004, 765773009, 765774003, 765775002, 765776001, 765777005, 765778000, 765779008, 765780006, 766249007, 766765009, 767094002, 770414008, 771149000, 771266007, 771447009, 771477003, 771509001, 772125001, 773647007, 773737004, 775909002, 776416004, 778025006, 782722002, 782738008, 782771007, 782942003, 783157004, 783159001, 783614008, 783620009, 155876003, 168041003, 168042005, 168044006, 168045007, 186796004, 187144000, 188250002, 188252005, 188253000, 190829000, 194779001, 194780003, 194781004, 194909006, 197579006, 197580009, 197582001, 197583006, 197584000, 197585004, 197589005, 197590001, 197591002, 197593004, 197594005, 197595006, 197596007, 197597003, 197598008, 197599000, 197600002, 197601003, 197603000, 197604006, 197605007, 197606008, 197607004, 197613008, 197616000, 197617009, 197618004, 197619007, 197626007, 197627003, 197629000, 197659005, 197660000, 197661001, 197664009, 197671004, 197681000, 197682007, 197683002, 197684008, 197685009, 197686005, 197687001, 197688006, 197689003, 197690007, 197691006, 197692004, 197693009, 197694003, 197695002, 197696001, 197697005, 197708002, 197709005, 197712008, 197713003, 197714009, 197715005, 197738008, 197739000, 197742006, 197743001, 197744007, 197745008, 197747000, 197753000, 197757004, 197760006, 197761005, 197762003, 197763008, 197764002, 197769007, 197770008, 197772000, 197782004, 197783009, 197794008, 197801000, 197802007, 197813005, 197817006,

197820003, 198708004, 198747002, 198949009, 198951008, 198952001, 198953006, 199007008, 199110003, 199132007, 199134008, 199135009, 199136005, 199137001, 200118004, 204938007, 204941003, 204942005, 204950001, 204957003, 204958008, 204984002, 204985001, 307309005, 307604008, 307618001, 307755009, 309426007, 310387003, 310647000, 310670008, 311366001, 311496007, 314967004, 359694003, 361146001, 361147005, 363224005, 363233007, 363234001, 363286005, 363287001, 363457009, 363518003, 370368007, 370493008, 370494002, 371011007, 371019009, 371020003, 373421000, 373422007, 373584008, 373599008, 373637000, 373651000, 210197003, 210203000, 210205007, 210206008, 210207004, 213150003, 213231008, 226309007, 230970001, 234348004, 234485006, 236369004, 236374007, 236375008, 236376009, 236377000, 236378005, 236380004, 236381000, 236382007, 236383002, 236384008, 236392004, 236393009, 236394003, 236395002, 236397005, 236398000, 236399008, 236400001, 236401002, 236402009, 236403004, 236404005, 236405006, 236406007, 236409000, 236410005, 236412002, 236413007, 236414001, 236415000, 236416004, 236417008, 236418003, 236419006, 236422008, 236423003, 236424009, 236426006, 236427002, 236428007, 236429004, 236430009, 236431008, 236432001, 236433006, 236434000, 236435004, 236436003, 236438002, 236441006, 236442004, 236443009, 236444003, 236445002, 236446001, 236447005, 236451007, 236452000, 236453005, 236454004, 236455003, 236456002, 236457006, 236458001, 236459009, 236460004, 236461000, 236463002, 236464008, 236466005, 236467001, 236468006, 236469003, 236470002, 236471003, 236472005, 236474006, 236475007, 236477004, 236478009, 236479001, 236480003, 236481004, 236482006, 236483001, 236484007, 236485008, 236486009, 236487000, 236495001, 236496000, 236498004, 236499007, 236500003, 236502006, 236503001, 236504007, 236505008, 236507000, 236508005, 236511006, 236512004, 236513009, 236514003, 236515002, 236516001, 236517005, 236518000, 236519008, 236520002, 236521003, 236523000, 236524006, 236525007, 236527004, 236528009, 236530006, 236531005, 236532003, 236535001, 236552002, 236569000, 236570004, 236571000, 236572007, 236573002, 236574008, 236575009, 236576005, 236577001, 236578006, 236579003, 236580000, 236581001, 236582008, 236583003, 236584009, 236586006, 236587002, 236588007, 236589004, 236590008, 236592000, 236593005, 236614007, 236708007, 236709004, 236710009, 236711008, 236713006, 237230004, 237612000, 239932005, 240317003, 253860008, 253862000, 253864004, 253865003, 253866002, 253867006, 253869009, 253872002, 253873007, 253874001, 253875000, 253876004, 253878003, 253879006, 253880009, 253881008, 253882001, 253883006, 253884000, 253885004, 253886003, 253897003, 35455006, 35546006, 36171008, 36225005, 36388008, 36402006, 36568005, 36689008, 36891003, 37061001, 37085009, 37133005, 37183000, 37497004, 37779008, 38046004, 38481006, 38898003, 39734002, 40068008, 40095003, 40233000, 40488004, 40951006, 41305006, 41729002, 41962002, 42399005, 42496002, 42927005, 43064006, 43258006, 43629001, 44323002, 44513007, 44730006, 45281005, 45646000, 45743004, 45812003, 45927004, 46177005, 46785007, 47330001, 48061001, 48631008, 48638002, 48655003, 48713002, 48796009, 49008000, 49102001, 49171007, 49220004, 49708008, 50909009, 51292008, 52042003, 52342006, 52845002, 53378008, 53556002, 54155004, 54781007, 54879000, 54967001, 55006001, 55536001, 55856005, 56108007, 56906005, 57088004, 57557005, 57684003, 57873008, 58574008, 58797008, 59400006, 59530001, 59531002, 59758007, 61165007, 61474001, 61503006, 61598006, 61852001, 62216007, 62240004, 62332007, 62853008, 63302006, 63510008, 64168005, 64212008, 64323009, 65443008, 66052004, 66451004, 66610008, 66709007, 66993009, 67132008, 68544003, 68815009, 69718008, 70092007, 71064009, 71110009, 71909003, 73030000, 73286009, 73305009, 74324004, 74594005, 75030003, 75150001, 75652008, 75712001, 76224000, 76521009, 77182004, 77186001, 77624000, 77945009, 78209002, 78311009, 78544004, 78815005, 80119007, 80321008, 399094007, 399190000, 399340005, 405573009, 405584002, 408642003, 418839003, 420279001, 420514000, 420715001, 421305000, 421893009, 421986006, 423322005, 423533009, 423919000, 424114000, 424188009, 425369003, 425384007, 425455002, 425879009, 426136000, 426598005, 427555000, 427649000, 428255004, 428720002, 429224003, 429489008, 430031008, 430035004, 430535006, 431480000, 431501001, 431855005, 431856006, 431857002, 433036004, 433144002,

433146000, 433229006, 438783006, 439990003, 440018001, 441815006, 441872001, 254092004, 254914004, 254915003, 254916002, 254917006, 254918001, 254919009, 254920003, 254921004, 254922006, 254923001, 254924007, 254925008, 254927000, 254928005, 262612001, 262891006, 262893009, 262894003, 262900003, 266549004, 266556005, 267430007, 268232000, 268234004, 268854008, 269257004, 269301005, 269489006, 269914004, 270494003, 270517006, 270910001, 271387005, 274401005, 275407001, 275408006, 275893001, 276583007, 276584001, 276585000, 276586004, 276627004, 276866009, 277010001, 277011002, 278531007, 283905005, 285639008, 298015003, 298127003, 301814009, 302849000, 302910002, 302922004, 1003426008, 1003842009, 1003861008, 111491000119107, 15957501000119101, 351391000119103, 351621000119108, 368201000119100, 368421000119108, 368431000119106, 368441000119102, 368451000119100, 368461000119103, 368471000119109, 8621100011910 2, 878805009, 890370003, 895345000, 897308007, 897310009, 897311008, 897312001, 1228881003, 1163077002, 1217070004, 1217380005

#### Preexisting bleeding disorders

ICD-9-CM: 286.0, 286.1, 286.2, 286.3, 286.4, 286.5, 286.7, 286.9, 287

ICD-10-CM: D66, D67, D68, D69, D68.0, D68.1, D68.2, D68.3, D68.31, D68.311, D68.312, D68.318, D68.32, D68.4, D68.5, D68.51, D68.52, D68.59, D68.6, D68.61, D68.62, D68.69, D68.8, D68.9, D69.0, D69.1, D69.2, D69.3, D69.4, D69.41, D69.42, D69.49, D69.5, D69.51, D69.59, D69.6, D69.8, D69.9

SNOMED CT: 4320005, 18604004, 19267009, 25904003, 33297000, 89729000, 128101008, 128105004, 10749581000119100, 10749641000119106, 28293008, 31925001, 37193007, 49762007, 88776002, 767712006, 767713001, 785308008, 161460007, 191287000, 234440005, 234454002, 234456000, 234467004, 37350004, 40855001, 43217004, 46981006, 47307007, 50189006, 55464009, 64779008, 439458000, 1177171004, 1162807005, 1162804003, 1177197005, 1286003, 1908008, 2036003, 3230006, 3760002, 4676006, 6364000, 6647006, 6935003, 9489006, 10153004, 10934005, 11013005, 12501008, 13507004, 13993001, 15132005, 16773005, 16872008, 16922007, 19307009, 19520006, 19682006, 21360006, 24149006, 24663001, 25380002, 26029002, 26843008, 27068000, 27312002, 30182008, 30479005, 32605001, 33169001, 33344008, 34395002, 34417008, 34478009, 80988005, 81783000, 85796009, 86075001, 87397002, 88540000, 90935002, 95623001, 95644001, 95839005, 95840007, 95841006, 95842004, 95843009, 95844003, 95845002, 105604006, 111452009, 111588002, 111589005, 123786007, 123787003, 123788008, 123789000, 123790009, 128088003, 128090002, 128092005, 128106003, 128107007, 128108002, 128113003, 128114009, 128115005, 1148849001, 1148857003, 1148859000, 1148862002, 1148929009, 1148930004, 1156746003, 1197595004, 789291002, 860725002, 39191000119103, 72161000119100, 97571000119109, 295101000119105, 295111000119108, 295121000119101, 10854051000119101, 1563006, 12153008, 12546009, 14230004, 33820001, 36351005, 41788008, 61551003, 65768009, 76407009, 76642003, 84048006, 86635005, 95605009, 234446004, 234447008, 234448003, 234450006, 234451005, 359700009, 421527008, 421766003, 609329007, 609456005, 609462000, 698694005, 699208000, 715559004, 716746003, 717407006, 717935003, 717936002, 717937006, 721304007, 724344004, 724356003, 724637001, 724767000, 724781003, 724854007, 725105006, 725157006, 725291001, 726513006, 733028000, 773333003, 773422002, 774084003, 782909004, 782934004, 783194008, 783250007, 783256001, 154818001, 154826009, 180481005, 181456001, 191288005, 191289002, 191290006, 191291005, 191292003, 191293008, 191296000, 191297009, 191298004, 191322006, 191323001, 196138005, 198912003, 198917009, 198918004, 200030007, 200031006, 200173001, 201436003, 307091009, 307115002, 307116001, 307342006, 307514008, 307515009, 307517001, 307518006, 309762007, 359704000, 359709005, 359711001, 359714009, 359717002, 359721009,

359723007, 359725000, 359727008, 359730001, 359732009, 371074009, 234442002, 234444001, 234445000, 234452003, 234453008, 234455001, 234457009, 234458004, 234459007, 234460002, 234461003, 234462005, 234463000, 234464006, 234465007, 234466008, 234468009, 237270006, 237271005, 237272003, 237336007, 237337003, 239886003, 239887007, 239888002, 239889005, 239890001, 239892009, 239894005, 239895006, 239897003, 35066007, 35554008, 35913006, 36402006, 38879000, 41106001, 41690001, 41816006, 43302000, 45366001, 45963004, 47546008, 48976006, 49177006, 49886003, 50770000, 51624005, 52042003, 52137009, 54072008, 58327003, 61802005, 61810006, 62410004, 62698000, 64315007, 64509006, 66909001, 67406007, 68815009, 69500007, 71723006, 73286009, 73397007, 73975000, 74576004, 76521009, 76771005, 77753005, 79674009, 402851000, 402865003, 403486000, 403487009, 403488004, 416902009, 425949001, 426199009, 438360006, 438372000, 438373005, 438599002, 438792009, 438827002, 439000005, 439001009, 439002002, 439006004, 439125003, 439126002, 439145006, 439156006, 439157002, 439274008, 439455002, 439459008, 439460003, 439698008, 439699000, 439701000, 439702007, 440820004, 440866009, 440867000, 440868005, 440924009, 440988005, 440989002, 440993008, 441001005, 441006000, 441042008, 441079006, 441101007, 441188004, 441189007, 441190003, 441191004, 441192006, 441420000, 441421001, 441697004, 441762006, 441945008, 441946009, 441990004, 442078001, 442121006, 442197003, 442363001, 442654007, 442760001, 267199000, 267272006, 278365007, 278366008, 278504009, 282707003, 296926001, 296927005, 296928000, 296929008, 296930003, 296931004, 296932006, 296933001, 296934007, 296935008, 296936009, 62751000119108, 1196942001, 1217411000, 1217410004, 1217409009

#### Placenta previa

ICD-10-CM: O44.03, O44.1, O44.23, O44.33, O44.10, O44.11, O44.12, O44.13

SNOMED CT: 7792000, 24095001, 198903000, 7266006, 27388005, 87814002, 119901000119109, 198899007, 198900002, 198905007, 198906008, 48888007, 267197003

ICD-9-CM: 641.0, 641.1

#### Multiple pregnancy

ICD-9-CM: 651, V27.2, V27.3, V27.4, V27.5, V27.6, V27.7, V91

ICD-10-CM: O30, O31, Z37.2, Z37.3, Z37.4, Z37.5, Z37.6, Z37.7, O30.0, O30.00, O30.001, O30.002, O30.003, O30.009, O30.01, O30.011, O30.012, O30.013, O30.019, O30.02, O30.021, O30.022, O30.023, O30.029, O30.03, O30.031, O30.032, O30.033, O30.039, O30.04, O30.041, O30.042, O30.043, O30.049, O30.09, O30.091, O30.092, O30.093, O30.131, O30.099, O30.1, O30.10, O30.101, O30.102, O30.103, O30.109, O30.11, O30.111, O30.112, O30.113, O30.119, O30.12, O30.121, O30.122, O30.123, O30.129, O30.13, O30.132, O30.133, O30.139, O30.19, O30.191, O30.192, O30.193, O30.199, O30.2, O30.20, O30.201, O30.202, O30.203, O30.209, O30.21, O30.211, O30.212, O30.213, O30.219, O30.22, O30.221, O30.222, O30.223, O30.229, O30.23, O30.231, O30.232, O30.233, O30.239, O30.29, O30.291, O30.292, O30.293, O30.299, O30.8, O30.80, O30.801, O30.802, O30.803, O30.809, O30.81, O30.811, O30.812, O30.813, O30.819, O30.82, O30.821, O30.822, O30.823, O30.829, O30.83, O30.831, O30.832, O30.833, O30.839, O30.89, O30.891, O30.892, O30.893, O30.899, O30.9, O30.90, O30.91, O30.92, O30.93, O31.0, O31.00, O31.00X0, O31.00X1, O31.00X2, O31.00X3, O31.00X4, O31.00X5, O31.00X9, O31.01, O31.01X0, O31.01X1, O31.01X2, O31.01X3, O31.01X4, O31.01X5, O31.01X9, O31.02, O31.02X0, O31.02X1, O31.02X2, O31.02X3, O31.02X4, O31.02X5, O31.02X9, O31.03, O31.03X0,

O31.03X1, O31.03X2, O31.03X3, O31.03X4, O31.03X5, O31.03X9, O31.1, O31.10, O31.10X0, O31.10X1, O31.10X2, O31.10X3, O31.10X4, O31.10X5, O31.10X9, O31.11, O31.11X0, O31.11X1, O31.11X2, O31.11X3, O31.11X4, O31.11X5, O31.11X9, O31.12, O31.12X0, O31.12X1, O31.12X2, O31.12X3, O31.12X4, O31.12X5, O31.12X9, O31.13, O31.13X0, O31.13X1, O31.13X2, O31.13X3, O31.13X4, O31.13X5, O31.13X9, O31.2, O31.20, O31.20X0, O31.20X1, O31.20X2, O31.20X3, O31.20X4, O31.20X5, O31.20X9, O31.21, O31.21X0, O31.21X1, O31.21X2, O31.21X3, O31.21X4, O31.21X5, O31.21X9, O31.22, O31.22X0, O31.22X1, O31.22X2, O31.22X3, O31.22X4, O31.22X5, O31.22X9, O31.23, O31.23X0, O31.23X1, O31.23X2, O31.23X3, O31.23X4, O31.23X5, O31.23X9, O31.3, O31.30, O31.30X0, O31.30X1, O31.30X2, O31.30X3, O31.30X4, O31.30X5, O31.30X9, O31.31, O31.31X0, O31.31X1, O31.31X2, O31.31X3, O31.31X4, O31.31X5, O31.31X9, O31.32, O31.32X0, O31.32X1, O31.32X2, O31.32X3, O31.32X4, O31.32X5, O31.32X9, O31.33, O31.33X0, O31.33X1, O31.33X2, O31.33X3, O31.33X4, O31.33X5, O31.33X9, O31.8, O31.8X, O31.8X1, O31.8X10, O31.8X11, O31.8X12, O31.8X13, O31.8X14, O31.8X15, O31.8X19, O31.8X2, O31.8X20, O31.8X21, O31.8X22, O31.8X23, O31.8X24, O31.8X25, O31.8X29, O31.8X3, O31.8X30, O31.8X31, O31.8X32, O31.8X33, O31.8X34, O31.8X35, O31.8X39, O31.8X9, O31.8X90, O31.8X91, O31.8X92, O31.8X93, O31.8X94, O31.8X95, O31.8X99, Z37.50, Z37.51, Z37.52, Z37.53, Z37.54, Z37.59, Z37.60, Z37.61, Z37.62, Z37.63, Z37.64, Z37.69

SNOMED CT: 199305006, 45384004, 3798002, 7888004, 8333008, 13859001, 15467003, 17333005, 20272009, 22514005, 24146004, 25192009, 28030000, 29997008, 30165006, 34089005, 34100008, 83121003, 84382006, 86803008, 87662006, 112075006, 10760661000119109, 10760701000119102, 10760741000119100, 10760781000119105, 10760821000119100, 10760861000119105, 10760901000119104, 151441000119105, 702452007, 702453002, 702454008, 762612009, 762613004, 762614005, 169828005, 169829002, 169830007, 169831006, 169832004, 169833009, 169834003, 199329004, 199330009, 199331008, 38257001, 39213008, 50758004, 52483005, 52942000, 54650005, 75697004, 281052005

#### Placental abruption

ICD-10-CM: O45, O45.0, O45.00, O45.001, O45.002, O45.003, O45.009, O45.01, O45.011, O45.012, O45.013, O45.019, O45.02, O45.021, O45.022, O45.023, O45.029, O45.09, O45.091, O45.092, O45.093, O45.099, O45.8, O45.8X, O45.8X1, O45.8X2, O45.8X3, O45.8X9, O45.9, O45.90, O45.91, O45.92, O45.93

SNOMED CT: 415105001, 10751771000119107, 198910006, 198911005, 198912003, 237273008, 237274002, 237275001, 62131008, 267197003

ICD-9-CM: 641.2

#### Preterm birth

ICD-10-CM: Z3A.20, Z3A.21, Z3A.22, Z3A.23, Z3A.24, Z3A.25, Z3A.26, Z3A.27, Z3A.28, Z3A.29, Z3A.30, Z3A.31, Z3A.32, Z3A.33, Z3A.34, Z3A.35, Z3A.36

SNOMED CT: 7707000, 13763000, 23464008, 84132007, 86883006, 90797000, 313178001, 41438001, 45139008, 46906003, 48688005, 57907009, 64920003, 65035007, 71355009, 72544005, 78395001, 6678005, 25026004, 54318006, 65683006, 72846000, 428567001

ICD-9-CM: 765.21, 765.22, 765.23, 765.24, 765.25, 765.26, 765.27, 765.28

#### Gastrointestinal disease

ICD-10-CM: K00, K01, K02, K03, K04, K05, K06, K08, K09, K11, K12, K13, K14, K20, K21, K22, K23, K25, K26, K27, K28, K29, K30, K31, K35, K36, K37, K38, K40, K41, K42, K43, K44, K45, K46, K50, K51, K52, K55, K56, K57, K58, K59, K60, K61, K62, K63, K64, K65, K66, K67, K68, K70, K71, K72, K73, K74, K75, K76, K77, K80, K81, K82, K83, K85, K86, K87, K90, K91, K92, K94, K95, O26.6, O99.6, K31.A0, K31.A22, K31.A2, K22.89, K31.A, K31.A1, K31.A19, K31.A21, K31.A13, K22.81, K31.A12, K31.A29, K31.A11, K31.A14, K22.82, K31.A15, K00.0, K00.1, K00.2, K00.3, K00.4, K00.5, K00.6, K00.7, K00.8, K00.9, K01.0, K01.1, K02.3, K02.5, K02.51, K02.52, K02.53, K02.6, K02.61, K02.62, K02.63, K02.7, K02.9, K03.0, K03.1, K03.2, K03.3, K03.4, K03.5, K03.6, K03.7, K03.8, K03.81, K03.89, K03.9, K04.0, K04.01, K04.02, K04.1, K04.2, K04.3, K04.4, K04.5, K04.6, K04.7, K04.8, K04.9, K04.90, K04.99, K05.0, K05.00, K05.01, K05.1, K05.10, K05.11, K05.2, K05.20, K05.21, K05.211, K05.212, K05.213, K05.219, K05.22, K05.221, K05.222, K05.223, K05.229, K05.3, K05.30, K05.31, K05.311, K05.312, K05.313, K05.319, K05.32, K05.321, K05.322, K05.323, K05.329, K05.4, K05.5, K05.6, K06.0, K06.01, K06.010, K06.011, K06.012, K06.013, K06.02, K06.020, K06.021, K06.022, K06.023, K06.1, K06.2, K06.3, K06.8, K06.9, K08.0, K08.1, K08.10, K08.101, K08.102, K08.103, K08.104, K08.109, K08.11, K08.111, K08.112, K08.113, K08.114, K08.119, K08.12, K08.121, K08.122, K08.123, K08.124, K08.129, K08.13, K08.131, K08.132, K08.133, K08.134, K08.139, K08.19, K08.191, K08.192, K08.193, K08.194, K08.199, K08.2, K08.20, K08.21, K08.22, K08.23, K08.24, K08.25, K08.26, K08.3, K08.4, K08.40, K08.401, K08.402, K08.403, K08.404, K08.409, K08.41, K08.411, K08.412, K08.413, K08.414, K08.419, K08.42, K08.421, K08.422, K08.423, K08.424, K08.429, K08.43, K08.431, K08.432, K08.433, K08.434, K08.439, K08.49, K08.491, K08.492, K08.493, K08.494, K08.499, K08.5, K08.50, K08.51, K08.52, K08.53, K08.530, K08.531, K08.539, K08.54, K08.55, K08.56, K08.59, K08.8, K08.81, K08.82, K08.89, K08.9, K09.0, K09.1, K09.8, K09.9, K11.0, K11.1, K11.2, K11.20, K11.21, K11.22, K11.23, K11.3, K11.4, K11.5, K11.6, K11.7, K11.8, K11.9, K12.0, K12.1, K12.2, K12.3, K12.30, K12.31, K12.32, K12.33, K12.39, K13.0, K13.1, K13.2, K13.21, K13.22, K13.23, K13.24, K13.29, K13.3, K13.4, K13.5, K13.6, K13.7, K13.70, K13.79, K14.0, K14.1, K14.2, K14.3, K14.4, K14.5, K14.6, K14.8, K14.9, K20.0, K20.8, K20.9, K21.0, K21.9, K22.0, K22.1, K22.10, K22.11, K22.2, K22.3, K22.4, K22.5, K22.6, K22.7, K22.70, K22.71, K22.710, K22.711, K22.719, K22.8, K22.9, K25.0, K25.1, K25.2, K25.3, K25.4, K25.5, K25.6, K25.7, K25.9, K26.0, K26.1, K26.2, K26.3, K26.4, K26.5, K26.6, K26.7, K26.9, K27.0, K27.1, K27.2, K27.3, K27.4, K27.5, K27.6, K27.7, K27.9, K28.0, K28.1, K28.2, K28.3, K28.4, K28.5, K28.6, K28.7, K28.9, K29.0, K29.00, K29.01, K29.2, K29.20, K29.21, K29.3, K29.30, K29.31, K29.4, K29.40, K29.41, K29.5, K29.50, K29.51, K29.6, K29.60, K29.61, K29.7, K29.70, K29.71, K29.8, K29.80, K29.81, K29.9, K29.90, K29.91, K31.0, K31.1, K31.2, K31.3, K31.4, K31.5, K31.6, K31.7, K31.8, K31.81, K31.811, K31.819, K31.82, K31.83, K31.84, K31.89, K31.9, K35.2, K35.20, K35.21, K35.3, K35.30, K35.31, K35.32, K35.33, K35.8, K35.80, K35.89, K35.890, K35.891, K38.0, K38.1, K38.2, K38.3, K38.8, K38.9, K40.0, K40.00, K40.01, K40.1, K40.10, K40.11, K40.2, K40.20, K40.21, K40.3, K40.30, K40.31, K40.4, K40.40, K40.41, K40.9, K40.90, K40.91, K41.0, K41.00, K41.01, K41.1, K41.10, K41.11, K41.2, K41.20, K41.21, K41.3, K41.30, K41.31, K41.4, K41.40, K41.41, K41.9, K41.90, K41.91, K42.0, K42.1, K42.9, K43.0, K43.1, K43.2, K43.3, K43.4, K43.5, K43.6, K43.7, K43.9, K44.0, K44.1, K44.9, K45.0, K45.1, K45.8, K46.0, K46.1, K46.9, K50.0, K50.00, K50.01, K50.011, K50.012, K50.013,

K50.014, K50.018, K50.019, K50.1, K50.10, K50.11, K50.111, K50.112, K50.113, K50.114, K50.118, K50.119, K50.8, K50.80, K50.81, K50.811, K50.812, K50.813, K50.814, K50.818, K50.819, K50.9, K50.90, K50.91, K50.911, K50.912, K50.913, K50.914, K50.918, K50.919, K51.0, K51.00, K51.01, K51.011, K51.012, K51.013, K51.014, K51.018, K51.019, K51.2, K51.20, K51.21, K51.211, K51.212, K51.213, K51.214, K51.218, K51.219, K51.3, K51.30, K51.31, K51.311, K51.312, K51.313, K51.314, K51.318, K51.319, K51.4, K51.40, K51.41, K51.411, K51.412, K51.413, K51.414, K51.418, K51.419, K51.5, K51.50, K51.51, K51.511, K51.512, K51.513, K51.514, K51.518, K51.519, K51.8, K51.80, K51.81, K51.811, K51.812, K51.813, K51.814, K51.818, K51.819, K51.9, K51.90, K51.91, K51.911, K51.912, K51.913, K51.914, K51.918, K51.919, K52.0, K52.1, K52.2, K52.21, K52.22, K52.29, K52.3, K52.8, K52.81, K52.82, K52.83, K52.831, K52.832, K52.838, K52.839, K52.89, K52.9, K55.0, K55.01, K55.011, K55.012, K55.019, K55.02, K55.021, K55.022, K55.029, K55.03, K55.031, K55.032, K55.039, K55.04, K55.041, K55.042, K55.049, K55.05, K55.051, K55.052, K55.059, K55.06, K55.061, K55.062, K55.069, K55.1, K55.2, K55.20, K55.21, K55.3, K55.30, K55.31, K55.32, K55.33, K55.8, K55.9, K56.0, K56.1, K56.2, K56.3, K56.4, K56.41, K56.49, K56.5, K56.50, K56.51, K56.52, K56.6, K56.60, K56.600, K56.601, K56.609, K56.69, K56.690, K56.691, K56.699, K56.7, K57.0, K57.00, K57.01, K57.1, K57.10, K57.11, K57.12, K57.13, K57.2, K57.20, K57.21, K57.3, K57.30, K57.31, K57.32, K57.33, K57.4, K57.40, K57.41, K57.5, K57.50, K57.51, K57.52, K57.53, K57.8, K57.80, K57.81, K57.9, K57.90, K57.91, K57.92, K57.93, K58.0, K58.1, K58.2, K58.8, K58.9, K59.0, K59.00, K59.01, K59.02, K59.03, K59.04, K59.09, K59.1, K59.2, K59.3, K59.31, K59.39, K59.4, K59.8, K59.9, K60.0, K60.1, K60.2, K60.3, K60.4, K60.5, K61.0, K61.1, K61.2, K61.3, K61.31, K61.39, K61.4, K61.5, K62.0, K62.1, K62.2, K62.3, K62.4, K62.5, K62.6, K62.7, K62.8, K62.81, K62.82, K62.89, K62.9, K63.0, K63.1, K63.2, K63.3, K63.4, K63.5, K63.8, K63.81, K63.89, K63.9, K64.0, K64.1, K64.2, K64.3, K64.4, K64.5, K64.8, K64.9, K65.0, K65.1, K65.2, K65.3, K65.4, K65.8, K65.9, K66.0, K66.1, K66.8, K66.9, K68.1, K68.11, K68.12, K68.19, K68.9, K70.0, K70.1, K70.10, K70.11, K70.2, K70.3, K70.30, K70.31, K70.4, K70.40, K70.41, K70.9, K71.0, K71.1, K71.10, K71.11, K71.2, K71.3, K71.4, K71.5, K71.50, K71.51, K71.6, K71.7, K71.8, K71.9, K72.0, K72.00, K72.01, K72.1, K72.10, K72.11, K72.9, K72.90, K72.91, K73.0, K73.1, K73.2, K73.8, K73.9, K74.0, K74.1, K74.2, K74.3, K74.4, K74.5, K74.6, K74.60, K74.69, K75.0, K75.1, K75.2, K75.3, K75.4, K75.8, K75.81, K75.89, K75.9, K76.0, K76.1, K76.2, K76.3, K76.4, K76.5, K76.6, K76.7, K76.8, K76.81, K76.89, K76.9, K80.0, K80.00, K80.01, K80.1, K80.10, K80.11, K80.12, K80.13, K80.18, K80.19, K80.2, K80.20, K80.21, K80.3, K80.30, K80.31, K80.32, K80.33, K80.34, K80.35, K80.36, K80.37, K80.4, K80.40, K80.41, K80.42, K80.43, K80.44, K80.45, K80.46, K80.47, K80.5, K80.50, K80.51, K80.6, K80.60, K80.61, K80.62, K80.63, K80.64, K80.65, K80.66, K80.67, K80.7, K80.70, K80.71, K80.8, K80.80, K80.81, K81.0, K81.1, K81.2, K81.9, K82.0, K82.1, K82.2, K82.3, K82.4, K82.8, K82.9, K82.A, K82.A1, K82.A2, K83.0, K83.01, K83.09, K83.1, K83.2, K83.3, K83.4, K83.5, K83.8, K83.9, K85.0, K85.00, K85.01, K85.02, K85.1, K85.10, K85.11, K85.12, K85.2, K85.20, K85.21, K85.22, K85.3, K85.30, K85.31, K85.32, K85.8, K85.80, K85.81, K85.82, K85.9, K85.90, K85.91, K85.92, K86.0, K86.1, K86.2, K86.3, K86.8, K86.81, K86.89, K86.9, K90.0, K90.1, K90.2, K90.3, K90.4, K90.41, K90.49, K90.8, K90.81, K90.89, K90.9, K91.0, K91.1, K91.2, K91.3, K91.30, K91.31, K91.32, K91.5, K91.6, K91.61, K91.62, K91.7, K91.71, K91.72, K91.8, K91.81, K91.82, K91.83, K91.84, K91.840, K91.841, K91.85, K91.850, K91.858, K91.86, K91.87, K91.870, K91.871, K91.872, K91.873, K91.89, K92.0, K92.1, K92.2, K92.8, K92.81, K92.89, K92.9, K94.0, K94.00, K94.01, K94.02, K94.03, K94.09, K94.1, K94.10, K94.11, K94.12, K94.13, K94.19, K94.2, K94.20, K94.21, K94.22, K94.23, K94.29, K94.3, K94.30, K94.31, K94.32, K94.33, K94.39, K95.0, K95.01, K95.09, K95.8, K95.81, K95.89, O26.61, O26.611, O26.612, O26.613, O26.619, O26.62, O26.63, O99.61, O99.611, O99.612, O99.613, O99.619, O99.62, O99.63, K20.80, K20.81, K20.90, K20.91,

K21.00, K21.01, K59.81, K59.89, K74.00, K74.01, K74.02

ICD-9-CM: 520, 521, 522, 523, 524, 525, 526, 527, 528, 529, 530, 531, 532, 533, 534, 535, 536, 537, 538, 539, 540, 541, 542, 543, 550, 551, 552, 553, 555, 556, 557, 558, 560, 562, 564, 565, 566, 567, 568, 569, 570, 571, 572, 573, 574, 575, 576, 577, 578, 579, 646.7, 648.90, 648.91, 648.93, 648.94

SNOMED CT: 3696007, 3855007, 10743008, 10890000, 13200003, 16121001, 16761005, 18526009, 81120009, 81256000, 85189001, 85919009, 105995000, 105997008, 119291004, 128071005, 128241005, 128321003, 128999004, 129129003, 328701000119101, 34000006, 63532004, 80967001, 81060008, 196546001, 197150008, 197166005, 197354009, 197456007, 197476001, 199294004, 371136004, 235595009, 235856003, 37657006, 39621005, 39839004, 41188003, 41309000, 46557008, 48661000, 50063009, 51868009, 52515009, 53619000, 59927004, 61170000, 62484002, 64766004, 66579008, 69244009, 74400008, 76581006, 76712006, 76783007, 396232000, 396347007, 397174009, 397825006, 414396006, 426867001, 266417001, 266418006, 266474003, 302918009, 1193809008, 1193808000, 1163531000, 1162590005, 1172640002, 1163001000, 551621000124109, 551611000124101, 11811541000119107, 11811581000119102, 1172589000, 1187643003, 1163088008, 1163087003, 1163117009, 1179217006, 1163080001, 1163142000, 1163144004, 1186854003, 1172840000, 1162443006, 1187632006, 1193651003, 1193652005, 1179407003, 1163113008, 1163140008, 1162565008, 1162262007, 1177039008, 1193650002, 1163143005, 1176987009, 1163116000, 1163559004, 1162855005, 1193803009, 1193738003, 1193737008, 1163555005, 1193802004, 1163498006, 1186970002, 1184703009, 1186967001, 1186969003, 1187148008, 1186964008, 1184620004, 1187638005, 1193752001, 1193754000, 1163492007, 1197732001, 1197428008, 1197754004, 1204346009, 1231176005, 515623061000119105, 527788091000119106, 253855721000119107, 1197727002, 1217373008, 1197032007, 1197693001, 1197724009, 1197691004, 1197692006, 1197688004, 1197690003, 1197685001, 1231377006, 1231379009, 1197665000, 1197659000, 1197661009, 1197359006, 1230314003, 1197686000, 1197680006, 1197682003, 1197714001, 1197771003, 1197708008, 1197706007, 1204466001, 1197707003, 1204464003, 1204461006, 1196912004, 1204467005, 1197184003, 1196966007, 1196965006, 1197016009, 1197015008, 1197013001, 1196862008, 1197743009, 1197739005, 1230342001, 1222580004, 1217010007, 1197033002, 1222643003, 1196893009, 1196891006, 1196901004, 1196888006, 1196890007, 1197288009, 1197733006, 1231378001, 1231380007, 1230140003, 1196885009, 1196887001, 1196834001, 1217490003, 1217617009, 1196825006, 1196826007, 1208985003, 1196823004, 1196824005, 1196822009, 1230002004, 1196850009, 1197662002, 1196848001, 1196845003, 1196847006, 1196842000, 1204421005, 123691000119104, 1197011004, 1197182004, 1197730009, 1197284006, 1197281003, 1197308005, 1197282005, 33971000087109, 34171000087105, 34191000087109, 1217489007, 1208744003, 1197728007, 1201779005, 1197726006, 1222660008, 1197677005, 1197679008, 1220591004, 1208349005, 1197674003, 1197723003, 1197725005, 1230291009, 1196853006, 1196854000, 1196846002, 1222675000, 1196881000, 1231153007, 1197354001, 1197355000, 1197286008, 1197283000, 1197307000, 1197279000, 1197276007, 1197342001, 1197669006, 1197670007, 1197014007, 1197150002, 1230143001, 1230142006, 1197768006, 1208488006, 1197667008, 1197668003, 1208745002, 1230009008, 1197701002, 1197527000, 1197687009, 1197683008, 1197704005, 1197684002, 1197740007, 1196900003, 1197742004, 1196896001, 1196897005, 1196894003, 1196895002, 1196889003, 1196884008, 1196886005, 1196833007, 1196835000, 1196859005, 1196861001, 1196903001, 1196905008, 1196899008, 1196830005, 10856801000119101, 1197698005, 1197700001, 1197696009, 1197697000, 1197694007, 1197695008, 1162504009, 1197741006, 1197716004, 11847611000119107, 1179293006, 1187071008, 1196876009, 1196874007, 1186965009, 1184621000, 1186631005, 1186966005, 1186599001, 1186629001, 1184704003, 1187156006, 1196816000, 1196818004, 1196855004, 1196857007, 1187194006, 1162445004, 1171358000, 1187641001, 1162578008, 1197722008, 1163357003, 1172608001, 1196858002, 1193801006, 1193800007, 1172793008, 1162771004,

1186614003, 1186630006, 1186865008, 1163127003, 1163128008, 1163125006, 1172951004, 1163110006, 1177030007, 1163107004, 1163568002, 1193736004, 1163090009, 1163145003, 1193729000, 1193731009, 1193805002, 1162501001, 1177179002, 1193804003, 1172791005, 1186652002, 1172961006, 1162599006, 1163456008, 1172358003, 1167366000, 1167367009, 1172988001, 1172387002, 1167368004, 1167369007, 1163455007, 1186735007, 1162521000, 1163488000, 1163493002, 1163502004, 1163463008, 1162506006, 1170359003, 1163408002, 1170355009, 1163109001, 1163081002, 1163464002, 1163480007, 1163467009, 1186654001, 1179071009, 1179034007, 1179073007, 1179031004, 1163462003, 1163477006, 1172958005, 1171359008, 1162728008, 1163367008, 1172591008, 1186864007, 1162865004, 1187519007, 1187521002, 1193735000, 1162589001, 1163347004, 1186869002, 1162856006, 1186814000, 1186811008, 1186805005, 1177023001, 1177072008, 1193740008, 1193739006, 1177037005, 1163494008, 1172889005, 1177073003, 1177074009, 1162597008, 1162567000, 1163589006, 1172728007, 1193799008, 1193798000, 1163108009, 1163123004, 1193742000, 1193741007, 1163126007, 1163124005, 1193744004, 1193743005, 1163122009, 1193746002, 1193745003, 1163120001, 1193748001, 1193747006, 1162561004, 1162577003, 1177070000, 1172732001, 11808651000119103, 11808731000119101, 11808211000119102, 11808251000119101, 16899621000119104, 11808771000119103, 11808691000119108, 1177026009, 1177041009, 1177021004, 1179456002, 1193750009, 1193749009, 1163121002, 1186713004, 1162777000, 1163496005, 1187039001, 1172901009, 1162587004, 1193807005, 1193806001, 1172607006, 1173099009, 1193649002, 1163146002, 1193751008, 1193753006, 1193733007, 1193732002, 1163521007, 1163114002, 1193660006, 1193658009, 1163115001, 1193682004, 1193681006, 1163112003, 1193685002, 1193684003, 1193688000, 1193687005, 1163522000, 1163111005, 1162579000, 1163356007, 1208726006, 1220580006, 335002, 407000, 450005, 517007, 536002, 643001, 652005, 658009, 675003, 703000, 755004, 799008, 845006, 964004, 975000, 981008, 984000, 1027000, 1034003, 1045000, 1051005, 1059007, 1116000, 1150009, 1208004, 1264004, 1287007, 1356007, 1367008, 1475003, 1488000, 1492007, 1512006, 1532007, 1542009, 1567007, 1698001, 1744008, 1761006, 1771008, 1778002, 1824008, 1826005, 1835003, 2043009, 2066005, 2091005, 2128005, 2129002, 2245007, 2303007, 2307008, 2365002, 2367005, 2435008, 2556008, 2624008, 2783007, 2807004, 2825006, 2901004, 2919008, 2946003, 2955000, 3004001, 3023008, 3084004, 3094009, 3097002, 3308008, 3376008, 3419005, 3482005, 3483000, 3549009, 3558002, 3560000, 3590007, 3639002, 3650004, 3703002, 3704008, 140004, 168000, 171008, 192008, 242004, 303002, 317006, 3738000, 3752003, 3762005, 3783004, 3797007, 3845008, 3886001, 3899003, 3913002, 3951002, 4195003, 4237001, 4264000, 4269005, 4356008, 4397001, 4399003, 4410001, 4494009, 4522001, 4556007, 4654002, 4661003, 4711003, 4834000, 4846001, 4896000, 4998000, 5126000, 5144008, 5153001, 5201002, 5327000, 5432003, 5492000, 5494004, 5562006, 5577003, 5596004, 5618007, 5639000, 5689008, 5753006, 5791006, 5820008, 5920007, 5964004, 5985004, 6033008, 6045004, 6066007, 6072007, 6075009, 6087002, 6121001, 6124009, 6137007, 6183001, 6215006, 6241000, 6288001, 6382002, 6441003, 6461009, 6477005, 6503008, 6659005, 6724001, 6761005, 6802007, 6838000, 6936002, 6971002, 7021009, 7024001, 7037003, 7102003, 7290007, 7346000, 7371002, 7399006, 7407001, 7425008, 7444001, 7475005, 7491008, 7522008, 7538002, 7731005, 7780000, 7815009, 7847004, 7881005, 7903009, 7920008, 7926002, 7941002, 8071005, 8090002, 8114009, 8262006, 8410006, 8421002, 8493009, 8543007, 8579004, 8587003, 8666004, 8733006, 8744003, 8765009, 8771003, 8832006, 8986002, 9058002, 9067002, 9124008, 9245008, 9273005, 9359003, 9366002, 9391002, 9473008, 9484001, 9491003, 9551004, 9563009, 9665009, 9707006, 9785001, 9815002, 9822005, 9829001, 9843006, 9914004, 9953008, 9984005, 10017004, 10184002, 10217006, 10295004, 10345003, 10351008, 10389003, 10480003, 10535001, 10665004, 10699001, 10816007, 10825001, 10866001, 10877007, 10883005, 10897002, 10920005, 10998006, 11003002, 11102005, 11178005, 11194003, 11223009, 11256005, 11266002, 11350001, 11426004, 11461005, 11552008, 11578004, 11619008, 11672007, 11683003, 11767005, 11818002, 11836002, 11938002, 11949008,

12002009, 12063002, 12104008, 12137005, 12245007, 12264001, 12269006, 12270007, 12274003, 12351004, 12355008, 12368000, 12384004, 12435008, 12463005, 12574004, 12625009, 12721007, 12776000, 12777009, 12779007, 12847006, 12932003, 13025001, 13026000, 13114007, 13140001, 13160009, 13177009, 13265006, 13267003, 13286006, 13457005, 13466009, 13468005, 13470001, 13483000, 13504006, 13516000, 13538003, 13568007, 13596001, 13630003, 13771001, 13886001, 13891000, 13916005, 13923006, 13959000, 14066009, 14094001, 14136000, 14220008, 14223005, 14311001, 14380007, 14430002, 14448006, 14465002, 14582003, 14725002, 14756005, 14808008, 14901003, 14911005, 14919007, 15033003, 15082003, 15115006, 15135007, 15151004, 15165002, 15230009, 15238002, 15270002, 15284007, 15342002, 15402006, 15419008, 15423000, 15445004, 15492000, 15511008, 15528006, 15699003, 15720001, 15733007, 15770003, 15836005, 15842009, 15848008, 15902003, 15970005, 15999000, 16000003, 16010007, 16069000, 16070004, 16101002, 16358007, 16376000, 16409009, 16459000, 16485001, 16516008, 16694003, 16714009, 16763008, 16816002, 16932000, 16934004, 16950007, 16957005, 16958000, 17067009, 17110002, 17186003, 17190001, 17206008, 17234001, 17266006, 17335003, 17346000, 17349007, 17355002, 17440005, 17442002, 17465007, 17531008, 17551007, 17552000, 17565009, 17593008, 17681007, 17709002, 17741008, 17755000, 17802000, 17890003, 17920008, 17960009, 17970006, 18028001, 18037001, 18081009, 18089006, 18116006, 18139000, 18147000, 18168004, 18169007, 18229003, 18269002, 18317005, 18342001, 18367003, 18389004, 18638007, 18665000, 18718003, 18735004, 18755003, 18773000, 18792003, 18796000, 18805001, 18846006, 18899000, 18910001, 18917003, 18935007, 19132000, 19213003, 19216006, 19263008, 19286001, 19311003, 19335008, 19345005, 19387007, 19523008, 19547001, 19569008, 19597002, 19640007, 19682006, 19742005, 19850005, 19881001, 19936001, 19943007, 19968009, 19995004, 20239009, 20306009, 20335001, 20402005, 20474007, 20509003, 20528005, 20547008, 20570000, 20607006, 20639004, 20674003, 20678000, 20725005, 20754004, 20759009, 20790006, 20813000, 20815007, 20824003, 20860008, 20915006, 20919000, 20926000, 20928004, 21366000, 21459002, 21565000, 21580006, 21692001, 21759003, 21763005, 21782001, 21809000, 21861000, 21863002, 21995002, 22149007, 22157005, 22169002, 22207007, 22231002, 22240003, 22304002, 22323009, 22361007, 22395006, 22500005, 22508003, 22511002, 22542007, 22589009, 22620000, 22711000, 22788004, 22845004, 22846003, 22883003, 22949006, 22974009, 23023009, 23062000, 23065003, 23080009, 23144006, 23166004, 23202007, 23234003, 23298002, 23344004, 23387001, 23401002, 23509002, 23512004, 23587002, 23649000, 23678004, 23693000, 23740006, 23812009, 23874000, 23913003, 23916006, 23949002, 23971007, 23997001, 24001002, 24060004, 24078009, 24087000, 24179004, 24194000, 24203005, 24291004, 24339001, 24383005, 24407009, 24412005, 24526004, 24557004, 24610009, 24617007, 24754009, 24764000, 24787008, 24789006, 24807004, 24810006, 24813008, 24829000, 25102003, 25110002, 25148007, 25273001, 25319005, 25335003, 25345001, 25349007, 25374005, 25418001, 25420003, 25458004, 25479004, 25507003, 25552000, 25554004, 25598004, 25617003, 25730006, 25744000, 25803005, 25817001, 25887009, 25896009, 25898005, 25924004, 25933002, 25942009, 25948008, 25959004, 25972003, 26006005, 26010008, 26103000, 26179002, 26221006, 26249004, 26284000, 26289005, 26315009, 26316005, 26373009, 26374003, 26421009, 26422002, 26485002, 26558007, 26597004, 26624006, 26629001, 26718008, 26748006, 26826005, 26828006, 26838001, 26874005, 26884006, 26918003, 26975007, 27002002, 27025001, 27123005, 27152008, 27156006, 27160009, 27235001, 27241008, 27281001, 27409004, 27673007, 27680009, 27697003, 27719009, 27729002, 27829006, 27858009, 27868004, 27878001, 27916005, 27943000, 27970007, 28041003, 28082003, 28132005, 28175004, 28198007, 28212002, 28358004, 28401004, 28412004, 28536002, 28545001, 28626004, 28670008, 28682004, 28698006, 28724005, 28756003, 28766006, 28826002, 28828001, 28845006, 28945005, 29026008, 29110005, 29120000, 29162007, 29256009, 29291001, 29384001, 29451002,

29479008, 29484002, 29553002, 29583006, 29632002, 29642000, 29691006, 29748005, 29755007, 29862005, 29874009, 29880001, 29886007, 29927001, 29980002, 30037006, 30074005, 30093007, 30102006, 30126008, 30140009, 30144000, 30183003, 30188007, 30265004, 30304000, 30464003, 30468000, 30477007, 30512007, 30514008, 30588004, 30657009, 30677002, 30689000, 30715007, 30719001, 30731004, 30736009, 30755009, 30811009, 30873000, 30874006, 30877004, 30888005, 30993009, 31005002, 31015008, 31155007, 31201001, 31258000, 31301004, 31309002, 31437008, 31452001, 31474007, 31499008, 31595004, 31607006, 31642005, 31686000, 31704005, 31712002, 31742004, 31841001, 31860008, 32038009, 32067005, 32084004, 32097002, 32110003, 32141002, 32161008, 32194006, 32200003, 32230006, 32236000, 32295003, 32337007, 32362007, 32450007, 32490005, 32527003, 32580004, 32614006, 32620007, 32665003, 32740005, 32883009, 32891000, 32900008, 32916005, 33020000, 33144001, 33167004, 33225004, 33257003, 33261009, 33334006, 33361006, 33370009, 33504000, 33505004, 33652004, 33687004, 33710003, 33727006, 33812003, 33838003, 33841007, 33906009, 33908005, 33931005, 33983003, 33990008, 33997006, 34021006, 34125004, 34255001, 34262005, 34282009, 34286007, 34346002, 34367002, 34570004, 1230310007, 80660001, 80700005, 80736008, 80753001, 80761006, 80770009, 80805005, 80829003, 80880002, 80953005, 81096007, 81115009, 81142005, 81159000, 81225008, 81318004, 81339006, 81363003, 81387001, 81407003, 81518000, 81543006, 81546003, 81624004, 81685000, 81704009, 81934005, 81950002, 82047000, 82090004, 82127005, 82196007, 82204006, 82212003, 82228008, 82308007, 82363001, 82368005, 82380002, 82393007, 82403002, 82770004, 82821008, 82872004, 82895008, 82908000, 82934008, 82949000, 82985000, 83028006, 83072009, 83134002, 83209008, 83256006, 83294006, 83412009, 83469008, 83605009, 83628005, 83714006, 83836003, 83864008, 83922009, 83976009, 84038009, 84046005, 84124004, 84150000, 84161008, 84233008, 84296002, 84305006, 84410009, 84534001, 84557007, 84568007, 84581000, 84621006, 84622004, 84647002, 84679006, 84681008, 84752003, 84974003, 85057007, 85187004, 85316008, 85479007, 85502002, 85521005, 85700008, 85787009, 85833008, 85836000, 85859006, 85870004, 85904008, 85920003, 85942002, 85968009, 85985006, 85991008, 86028001, 86070006, 86092005, 86247009, 86258000, 86422009, 86454000, 86507001, 86514004, 86595004, 86615009, 86709000, 86895006, 86941000, 86983005, 86993003, 87047005, 87093008, 87142002, 87177002, 87181002, 87248009, 87252009, 87282003, 87326000, 87510000, 87541004, 87551003, 87621000, 87715008, 87756006, 87763006, 87781009, 87796008, 87806008, 88035001, 88078006, 88169003, 88172005, 88178009, 88239001, 88280008, 88281007, 88411000, 88477005, 88539002, 88580009, 88659005, 88700009, 88773005, 88898002, 88940006, 88968005, 89030000, 89057003, 89166001, 89199004, 89231008, 89251007, 89308006, 89378007, 89424006, 89444000, 89452002, 89454001, 89469000, 89489004, 89499009, 89573004, 89580002, 89628003, 89662003, 89722009, 89748001, 89762008, 89789003, 89790007, 89835009, 89933001, 89954008, 89988002, 90054000, 90176007, 90257004, 90316008, 90325002, 90365003, 90377008, 90446007, 90450000, 90458007, 90489006, 90493000, 90535007, 90628007, 90669003, 90678009, 90679001, 90753004, 90782003, 90803001, 90858003, 90886006, 90891007, 90979004, 91002001, 91105001, 91109007, 91162000, 91173007, 91210006, 91313006, 91316003, 91375006, 91399006, 91446004, 91448003, 91465007, 91489000, 91580003, 91631003, 91669008, 91843000, 91844006, 91850001, 91851002, 91862002, 91863007, 91867008, 91868003, 91890003, 91924004, 91946007, 91949000, 91950000, 91958007, 91964000, 91969005, 91971005, 91972003, 91975001, 91976000, 91978004, 91981009, 91985000, 91990002, 91991003, 92018005, 92020008, 92031000, 92039003, 92040001, 92042009, 92054009, 92065004, 92066003, 92067007, 92075001, 92076000, 92078004, 92080005, 92091003, 92096008, 92109005, 92116006, 92117002, 92118007, 92119004, 92125000, 92126004, 92130001, 92133004, 92138008, 92139000, 92140003, 92151003, 92154006, 92162003, 92164002, 92170008, 92172000, 92174004, 92176002, 92177006, 92178001, 92183009,

92184003, 92185002, 92186001, 92189008, 92193002, 92205005, 92219005, 92220004, 92259008, 92263001, 92264007, 92265008, 92269002, 92270001, 92277003, 92279000, 92284006, 92287004, 92288009, 92290005, 92293007, 92300003, 92301004, 92303001, 92304007, 92310007, 92311006, 92312004, 92314003, 92315002, 92318000, 92323000, 92324006, 92330006, 92343006, 92369000, 92385005, 92386006, 92409001, 92411005, 92413008, 92415001, 92418004, 92423004, 92429000, 92432002, 92436004, 92442000, 92443005, 92444004, 92445003, 92448001, 92457007, 92462008, 92463003, 92472006, 92475008, 92477000, 92478005, 92479002, 92480004, 92481000, 92505009, 92507001, 92524007, 92529002, 92531006, 92532004, 92534003, 92535002, 92537005, 92539008, 92542002, 92544001, 92545000, 92547008, 92549006, 92551005, 92558004, 92559007, 92560002, 92563000, 92568009, 92569001, 92570000, 92574009, 92575005, 92576006, 92577002, 92585006, 92589000, 92595004, 92598002, 92599005, 92600008, 92601007, 92604004, 92605003, 92607006, 92608001, 92610004, 92611000, 92612007, 92613002, 92614008, 92617001, 92618006, 92619003, 92621008, 92623006, 92629005, 92631001, 92633003, 92636006, 92637002, 92642005, 92643000, 92644006, 92645007, 92648009, 92651002, 92658008, 92659000, 92664001, 92668003, 92671006, 92672004, 92673009, 92674003, 92678000, 92680006, 92681005, 92686000, 92687009, 92688004, 92689007, 92692006, 92693001, 92694007, 92695008, 92696009, 92700001, 92704005, 92708008, 92734000, 92750008, 92751007, 92756002, 92758001, 92759009, 92760004, 92763002, 92768006, 92769003, 92770002, 92771003, 92773000, 92776008, 92780003, 92781004, 92790006, 92792003, 92794002, 92795001, 92796000, 92797009, 92798004, 92804002, 92825002, 92864004, 92885004, 92913004, 92924002, 92934006, 93032003, 93039007, 93058003, 93060001, 93063004, 93066007, 93091007, 93154004, 93155003, 93164008, 93165009, 93166005, 93209006, 93259002, 93268000, 93324004, 93330004, 93332007, 93343004, 93369005, 93376000, 93386004, 93387008, 93391003, 93395007, 93420009, 93422001, 93426003, 93434009, 93446008, 93449001, 93462002, 93463007, 93464001, 93469006, 93556005, 93560008, 93640008, 93662008, 93667002, 93669004, 93670003, 93672006, 93674007, 93676009, 93679002, 93683002, 93687001, 93715005, 93717002, 93728003, 93738008, 93761005, 93763008, 93770008, 93771007, 93773005, 93775003, 93802007, 93809003, 93812000, 93818001, 93826009, 93829002, 93830007, 93831006, 93832004, 93835002, 93836001, 93837005, 93843007, 93846004, 93848003, 93854002, 93858004, 93860002, 93861003, 93862005, 93867004, 93868009, 93870000, 93883004, 93933005, 93939009, 93942003, 93947009, 93952004, 93955002, 93956001, 93958000, 93961004, 93967000, 93968005, 93970001, 93971002, 93976007, 93977003, 93978008, 93980002, 93981003, 93984006, 93989001, 93994001, 94006002, 94032008, 94048009, 94049001, 94072004, 94076001, 94077005, 94078000, 94082003, 94089007, 94090003, 94101009, 94102002, 94103007, 94105000, 94120006, 94129007, 94132005, 94134006, 94135007, 94138009, 94152006, 94158005, 94163009, 94164003, 94165002, 94166001, 94168000, 94169008, 94170009, 94172001, 94175004, 94179005, 94184004, 94185003, 94212002, 94214001, 94226006, 94234000, 94235004, 94237007, 94249008, 94260004, 94261000, 94262007, 94270002, 94271003, 94273000, 94275007, 94286009, 94291005, 94304008, 94311007, 94312000, 94313005, 94314004, 94320003, 94321004, 94325008, 94328005, 94332004, 94333009, 94334003, 94335002, 94341009, 94342002, 94343007, 94346004, 94349006, 94354002, 94357009, 94359007, 94367004, 94369001, 94371001, 94372008, 94373003, 94378007, 94379004, 94380001, 94381002, 94384005, 94388008, 94400003, 94414003, 94415002, 94416001, 94454001, 94458003, 94459006, 94460001, 94464005, 94465006, 94472007, 94474008, 94479003, 94482008, 94483003, 94485005, 94488007, 94495003, 94496002, 94498001, 94499009, 94505005, 94506006, 94507002, 94509004, 94510009, 94513006, 94518002, 94519005, 94525009, 94538001, 94564007, 94580002, 94581003, 94604000, 94606003, 94608002, 94610000, 94613003, 94618007, 94624001, 94625000, 94626004, 94627008, 94631002, 94637003, 94638008, 94639000, 94640003, 94643001, 94652005, 94657004, 94658009, 94667009,

94670008, 94672000, 94673005, 94674004, 94675003, 94676002, 94721002, 94727003, 94732002, 94733007, 94734001, 94735000, 94737008, 94738003, 94739006, 94741007, 94744004, 94748001, 94752001, 94756003, 94758002, 94768007, 94775008, 94777000, 94783002, 94790007, 94801006, 94802004, 94803009, 94810003, 94811004, 94813001, 94815008, 94825003, 94830004, 94842009, 94849000, 94850000, 94851001, 94852008, 94858007, 94863006, 94866003, 94869005, 94870006, 94871005, 94872003, 94875001, 94876000, 94877009, 94878004, 94880005, 94883007, 94886004, 94888003, 94894006, 94896008, 94898009, 94900006, 94901005, 94902003, 94907009, 94908004, 94909007, 94910002, 94913000, 94917004, 94935008, 94936009, 94937000, 94959003, 94973007, 94977008, 94978003, 94979006, 94982001, 94987007, 94989005, 94992009, 94995006, 94996007, 94998008, 95001001, 95007002, 95008007, 95010009, 95011008, 95016003, 95017007, 95018002, 95020004, 95021000, 95024008, 95029003, 95034004, 95046000, 95072009, 95088009, 95089001, 95112004, 95116001, 95118000, 95122005, 95128009, 95129001, 95130006, 95132003, 95135001, 95140009, 95141008, 95142001, 95143006, 95145004, 95154001, 95159006, 95160001, 95169000, 95172007, 95174008, 95175009, 95176005, 95177001, 95178006, 95203009, 95204003, 95205002, 95212006, 95214007, 95238006, 95246007, 95247003, 95248008, 95249000, 95252008, 95253003, 95254009, 95267007, 95269005, 95297004, 95303006, 95306003, 95308002, 95435007, 95446005, 95447001, 95470009, 95471008, 95472001, 95474000, 95516005, 95518006, 95519003, 95520009, 95524000, 95525004, 95527007, 95529005, 95530000, 95531001, 95532008, 95533003, 95535005, 95536006, 95537002, 95538007, 95539004, 95540002, 95544006, 95545007, 95546008, 95547004, 95548009, 95549001, 95550001, 95552009, 95554005, 95555006, 95556007, 95557003, 95558008, 95559000, 95560005, 95561009, 95562002, 95563007, 95564001, 95578000, 95579008, 95580006, 95625008, 95626009, 95879001, 95885008, 95893008, 95894002, 95897009, 102453009, 105992002, 109244004, 109246002, 109271004, 109272006, 109287001, 109288006, 109367000, 109368005, 109434003, 109435002, 109436001, 109437005, 109438000, 109439008, 109440005, 109442002, 109443007, 109444001, 109445000, 109446004, 109447008, 109448003, 109449006, 109450006, 109451005, 109452003, 109453008, 109454002, 109455001, 109456000, 109457009, 109458004, 109459007, 109460002, 109461003, 109462005, 109463000, 109464006, 109466008, 109467004, 109468009, 109470000, 109471001, 109472008, 109473003, 109474009, 109475005, 109476006, 109477002, 109478007, 109479004, 109480001, 109481002, 109482009, 109483004, 109484005, 109485006, 109486007, 109488008, 109489000, 109490009, 109491008, 109492001, 109495004, 109496003, 109497007, 109498002, 109499005, 109501002, 109505006, 109506007, 109508008, 109509000, 109510005, 109511009, 109512002, 109520000, 109521001, 109526006, 109527002, 109530009, 109531008, 109532001, 109533006, 109534000, 109536003, 109537007, 109538002, 109540007, 109541006, 109542004, 109544003, 109545002, 109549008, 109550008, 109551007, 109552000, 109554004, 109556002, 109557006, 109564008, 109566005, 109568006, 109569003, 109570002, 109571003, 109572005, 109573000, 109574006, 109575007, 109576008, 109577004, 109578009, 109580003, 109581004, 109582006, 109583001, 109584007, 109585008, 109586009, 109588005, 109590006, 109592003, 109593008, 109594002, 109597009, 109598004, 109599007, 109600005, 109601009, 109602002, 109603007, 109604001, 109606004, 109607008, 109608003, 109609006, 109610001, 109611002, 109612009, 109613004, 109614005, 109615006, 109617003, 109620006, 109621005, 109622003, 109623008, 109624002, 109625001, 109626000, 109627009, 109628004, 109629007, 109630002, 109631003, 109632005, 109633000, 109634006, 109635007, 109636008, 109637004, 109639001, 109644008, 109645009, 109646005, 109647001, 109648006, 109649003, 109650003, 109657000, 109658005, 109671008, 109672001, 109673006, 109674000, 109675004, 109676003, 109677007, 109678002, 109679005, 109680008, 109681007, 109682000, 109683005, 109684004, 109685003, 109701004, 109702006, 109703001, 109704007, 109705008, 109706009, 109707000, 109708005, 109709002, 109710007, 109711006, 109712004, 109713009, 109714003, 109737009, 109738004, 109739007, 109740009, 109746003, 109747007, 109748002, 109749005, 109750005, 109751009, 109752002, 109753007, 109754001, 109755000, 109756004,

109757008, 109758003, 109759006, 109760001, 109761002, 109762009, 109763004, 109764005, 109765006, 109766007, 109768008, 109769000, 109770004, 109773002, 109774008, 109775009, 109776005, 109777001, 109779003, 109780000, 109781001, 109782008, 109783003, 109784009, 109785005, 109787002, 109788007, 109789004, 109790008, 109793005, 109794004, 109795003, 109798001, 109799009, 109800008, 109801007, 109802000, 109803005, 109804004, 109805003, 109806002, 109807006, 109808001, 109809009, 109810004, 109811000, 109812007, 109813002, 109814008, 109817001, 109819003, 109820009, 109821008, 109822001, 109823006, 109824000, 109825004, 109826003, 109827007, 109828002, 109830000, 109832008, 109833003, 109835005, 109836006, 109837002, 109838007, 109839004, 109840002, 109841003, 109842005, 109843000, 109844006, 109847004, 109848009, 109850001, 109851002, 109852009, 109853004, 109854005, 109855006, 109856007, 109857003, 109858008, 109866004, 110013004, 110021005, 110025001, 110052006, 110053001, 110054007, 110055008, 110056009, 110084004, 110085003, 110086002, 110087006, 110088001, 110089009, 110090000, 110091001, 110092008, 110093003, 110094009, 110095005, 110096006, 110097002, 110098007, 110099004, 110100007, 110101006, 110102004, 110103009, 110104003, 110105002, 110106001, 110107005, 110108000, 110109008, 110110003, 110111004, 110112006, 110113001, 110114007, 110115008, 110116009, 110117000, 110118005, 110119002, 110120008, 110121007, 110122000, 110123005, 110129009, 110130004, 110131000, 110132007, 110133002, 110134008, 110135009, 110136005, 110137001, 110138006, 110139003, 110140001, 110141002, 110142009, 110143004, 110151001, 110152008, 110153003, 110154009, 110155005, 110156006, 110157002, 110158007, 110159004, 110160009, 110161008, 110162001, 110163006, 110167007, 110178004, 110179007, 110180005, 110181009, 110190002, 110191003, 110192005, 110193000, 110194006, 110195007, 110196008, 110197004, 110198009, 110199001, 110200003, 110201004, 110202006, 110203001, 110204007, 110205008, 110206009, 110207000, 110208005, 110209002, 110210007, 110211006, 110212004, 110213009, 110214003, 110215002, 110216001, 110217005, 110222005, 110223000, 110224006, 110225007, 110226008, 110227004, 110228009, 110229001, 110230006, 110231005, 110232003, 110233008, 110234002, 110235001, 110236000, 110242001, 110248002, 110250005, 110251009, 110252002, 110253007, 110254001, 110255000, 110256004, 110257008, 110258003, 110260001, 110261002, 110263004, 110267003, 110277001, 110315000, 110316004, 110993001, 110994007, 110995008, 111277007, 111326002, 111327006, 111328001, 111329009, 111330004, 111331000, 111342009, 111343004, 111345006, 111348008, 111350000, 111352008, 111353003, 111354009, 111356006, 111357002, 111358007, 111359004, 111360009, 111361008, 111362001, 111363006, 111365004, 111366003, 111368002, 111369005, 111371005, 111372003, 111373008, 111374002, 111386004, 111404004, 111560008, 111591002, 111683002, 111704000, 111751003, 111798006, 111816002, 111819009, 111843007, 111879004, 111891008, 111892001, 111896003, 111911008, 111912001, 111923002, 118926004, 118938008, 118942006, 119292006, 119522002, 119523007, 122449004, 122483006, 123313007, 123600006, 123601005, 123602003, 123603008, 123604002, 123605001, 123606000, 123607009, 123608004, 123662000, 123669009, 123716002, 123717006, 123762005, 123952009, 125591009, 125625000, 125626004, 125627008, 125628003, 125629006, 125630001, 125631002, 125632009, 125633004, 125634005, 125635006, 125636007, 126498005, 126514006, 126661001, 126662008, 126663003, 126664009, 126665005, 126666006, 126680004, 126681000, 126682007, 126683002, 126684008, 126685009, 126686005, 126687001, 126688006, 126690007, 126691006, 126700009, 126702001, 126764002, 126765001, 126768004, 126769007, 126770008, 126771007, 126772000, 126773005, 126774004, 126775003, 126776002, 126777006, 126778001, 126779009, 126780007, 126781006, 126782004, 126783009, 126784003, 126785002, 126786001, 126787005, 126788000, 126789008, 126790004, 126791000, 126792007, 126793002, 126794008, 126795009, 126796005, 126797001, 126798006, 126799003, 126802007, 126803002, 126804008, 126807001, 126808006, 126809003, 126810008, 126811007, 126812000, 126813005, 126815003, 126816002, 126817006, 126818001, 126819009, 126820003, 126821004, 126822006, 126823001, 126824007, 126825008, 126826009, 126827000, 126828005, 126829002, 126830007, 126831006, 126832004, 126833009, 126834003, 126835002, 126837005, 126838000, 126839008, 126840005, 126841009, 126842002, 126843007, 126844001, 126845000, 126846004, 126847008, 126848003,

126849006, 126850006, 126851005, 126852003, 126853008, 126854002, 126855001, 126856000, 126857009, 126858004, 126859007, 126860002, 126861003, 126862005, 126863000, 126864006, 126865007, 126866008, 126867004, 126868009, 126869001, 126870000, 126871001, 126873003, 126874009, 126876006, 126893006, 126917007, 126918002, 126919005, 127116008, 127141001, 127142008, 127154008, 127178003, 127227003, 127228008, 127229000, 127253006, 127271004, 127313006, 127316003, 127317007, 127318002, 127321000, 128046007, 128068002, 128264007, 128284006, 128285007, 128286008, 128287004, 128288009, 128302006, 128332003, 128334002, 128335001, 128346003, 128347007, 128348002, 128415001, 128524007, 128525008, 128545000, 128600008, 129144002, 129586002, 129587006, 129597002, 129598007, 129601002, 134402008, 135830006, 135874008, 107811000119106, 1082521000119109, 1137557006, 1137560004, 1137562007, 1137644008, 1141621000, 1141622007, 1141626005, 1141877000, 1142042001, 1142226000, 1144252005, 1144303008, 1144304002, 1144305001, 1144306000, 1144307009, 1144353002, 1144354008, 1144355009, 1144421007, 1144423005, 1144764001, 1144891007, 1144892000, 1144927000, 1144937005, 1144938000, 1144939008, 1144951005, 1144965007, 1144967004, 1144969001, 1144971001, 1144981002, 1144982009, 1144983004, 1144990009, 1145001009, 1145027009, 1145117000, 1145203001, 1145543005, 1145544004, 1148489000, 1148541006, 1148551007, 1148555003, 1148568006, 1148573000, 1148585008, 1148699004, 1148766007, 1148900007, 1148930004, 1149062005, 1149069001, 1149232006, 1149233001, 1149439000, 1149440003, 1149441004, 1149442006, 1149443001, 1149444007, 1149445008, 1149446009, 1149447000, 1149448005, 1149449002, 1149450002, 1149451003, 1149452005, 1149453000, 1149455007, 1149456008, 1149457004, 1149458009, 1149459001, 1149460006, 1153354008, 1153355009, 1153356005, 1153357001, 1153383006, 1153384000, 1153385004, 1153386003, 1153387007, 1153388002, 1153393004, 1153404003, 1153405002, 1155681001, 1155718008, 1155727009, 1155841005, 1155910005, 1155913007, 1156107009, 1156108004, 1156207004, 1156269004, 1156419009, 1156770004, 1156783003, 1156785005, 1156786006, 1156788007, 1156793005, 1156795003, 1156797006, 1156799009, 1156809009, 1156810004, 1156811000, 1156812007, 1156815009, 1156992005, 11847571000119103, 11855251000119102, 15966541000119105, 15970341000119106, 16091411000119109, 16218761000119102, 16636051000119105, 16636101000119105, 16637101000119107, 16823941000119108, 16899941000119102, 250856961000119105, 347218661000119103, 352701000119102, 471891000124101, 681681000119109, 681691000119107, 681701000119107, 681751000119106, 681781000119104, 681831000119107, 681911000119108, 681941000119107, 681971000119100, 682031000119109, 682541000119106, 682601000119105, 682611000119108, 683031000119101, 683041000119105, 683071000119103, 683241000119101, 683421000119106, 683711000119109, 683861000119103, 688671000119103, 688681000119100, 688751000119103, 688781000119105, 688811000119107, 688891000119103, 688901000119104, 688911000119101, 688931000119106, 688981000119107, 689211000119102, 689251000119101, 689281000119108, 689311000119105, 689361000119108, 689371000119102, 689401000119104, 689451000119100, 689471000119109, 689481000119107, 689491000119105, 689501000119103, 690751000119102, 8891000119108, 89600751000119100, 972604701000119104, 1187631004, 1177081002, 1179393009, 786878009, 787091002, 787410005, 787414001, 787446001, 787447005, 787448000, 787730008, 788010000, 788011001, 788012008, 788013003, 788390005, 788491008, 788492001, 788631003, 788632005, 788718000, 788869006, 788875002, 788951001, 788965004, 788982002, 788986004, 788987008, 788988003, 788989006, 788990002, 788991003, 789005009, 789006005, 789516009, 789673002, 789685005, 789697006, 792907004, 817966005, 818951009, 819953000, 822975003, 827045004, 830022001, 830074003, 830075002, 830222008, 836274002, 838303003, 838305005, 838311008, 838318002, 838349007, 838352004, 838354003, 838365008, 838372009, 838374005, 838375006, 838377003, 838380002, 840423002, 840424008, 840460003, 840499006, 840503000, 840504006, 840506008, 840591006, 840592004, 846579005, 846606007, 846685008, 860596009, 860597000, 860598005, 860750004, 860789005, 860828007, 860829004, 860830009, 860831008, 860856002, 860860004, 860861000, 860862007, 860872005, 860874006, 860878009, 860885008, 860887000, 860888005, 860889002, 860891005, 863897005, 863915002, 863957008, 865912001, 865890009, 865891008, 865951006, 866048009, 866052009, 866070007, 866071006, 866074003,

866077005, 866110001, 866119000, 866120006, 866122003, 866141000, 866142007, 866176003, 870298008, 870300008, 870327007, 870325004, 870326003, 870349001, 870351002, 870353004, 870355006, 870428003, 870517000, 870625007, 871619002, 871631000, 871633002, 871634008, 871635009, 871745001, 871901008, 831000119103, 1171000119108, 1651000119109, 1691000119104, 1701000119104, 2171000124100, 2181000124102, 2191000124104, 2201000124101, 2211000124103, 2221000124106, 2231000124109, 2241000124104, 2251000124102, 2261000124100, 2271000124107, 2281000124105, 2291000124108, 2301000124109, 2311000124107, 2321000124104, 2331000124101, 2341000124106, 2351000124108, 2361000124105, 2371000124103, 2381000124100, 2391000124102, 2401000124100, 2411000124102, 2421000124105, 2431000124108, 2441000124103, 2451000124101, 2461000124104, 2471000124106, 2481000124109, 2491000124107, 2501000124104, 2511000124101, 2521000124109, 2531000124107, 2541000124102, 2551000124100, 2561000124103, 2571000124105, 2581000124108, 2591000124106, 2601000124103, 2611000124100, 2621000124108, 2631000124106, 2641000124101, 2651000124104, 2661000124102, 2671000124109, 2681000124107, 3001000119103, 3411000119107, 4611000119106, 4661000119109, 4911000119101, 5021000119108, 5211000119107, 5221000119100, 5601000119107, 5661000124106, 5891000119102, 7071000119102, 7111000119109, 7391000119103, 7401000119101, 7941000119105, 8161000119106, 8631000119105, 8651000119104, 8661000119102, 8921000119103, 9541000119105, 10321000132105, 10331000132107, 10351000132101, 12851000119104, 13641000119106, 14511000119108, 18121000119104, 21101000119105, 21321000119104, 21811000119104, 21871000119107, 25741000119109, 29731000119103, 32201000119107, 33041000119105, 33411000119104, 33431000119109, 35151000119105, 38871000119104, 40241000119109, 40271000119102, 40331000119107, 41271000119108, 63061000119104, 67401000119103, 72701000119102, 76691000119101, 84361000119102, 88171000119100, 90041000119109, 90051000119106, 90061000119108, 90071000119102, 90091000119101, 90101000119106, 90111000119109, 90121000119102, 90971000119103, 94291000119103, 95001000119103, 96081000119101, 96141000119105, 96281000119107, 96981000119102, 97051000119105, 97761000119105, 97801000119102, 97961000119108, 98981000119103, 99111000119103, 102401000119108, 103611000119102, 108111000119103, 112721000119106, 113561000119103, 113571000119109, 116291000119103, 117631000119103, 119371000119100, 120541000119103, 121801000119105, 123121000119107, 123141000119101, 123151000119104, 123161000119102, 123171000119108, 123181000119106, 123201000119107, 123231000119100, 123241000119109, 123251000119106, 123271000119102, 123281000119104, 123411000119106, 123701000119104, 128041000119107, 129141000119104, 130191000119102, 130351000119105, 130371000119101, 130381000119103, 130431000119105, 130441000119101, 130451000119104, 130461000119102, 130471000119108, 130481000119106, 130521000119106, 130531000119109, 130571000119107, 133171000119105, 133411000119108, 133421000119101, 133751000119102, 134021000119105, 135091000119106, 136801000119102, 138561000119104, 141001000119104, 145321000119108, 146291000119108, 147881000119108, 150541000119104, 150721000119102, 153821000119103, 154211000119108, 184741000119107, 184861000119102, 184881000119106, 184891000119109, 190191000119107, 202161000119104, 205941000119107, 208061000119101, 218541000119109, 226521000119108, 236811000119101, 242951000119108, 243161000119102, 290641000119107, 291681000119108, 294001000119105, 302141000119108, 302151000119105, 302641000119102, 302651000119100, 307001000119105, 307011000119108, 307281000119108, 318161000119106, 318191000009109, 328691000119101, 347891000119103, 367181000119102, 367531000119106, 368101000119109, 429671000124101, 430961000124105, 431431000124100, 431501000124109, 431931000124108, 433981000124105, 435081000124107, 435091000124105, 435101000124104, 435111000124101, 435121000124109, 435131000124107, 435161000124103, 435191000124106, 435211000124107, 447961000124105, 450471000124106, 454521000124101, 458581000124106, 459371000124108, 459401000124106, 459491000124104, 459501000124107, 681601000119101, 681621000119105, 681631000119108, 681651000119102, 681721000119103, 682821000119102, 682831000119104, 687011000119101, 689991000119100, 690761000119100, 690771000119106, 1082371000119104, 1082381000119101, 1082431000119108, 1082441000119104, 1082551000119101, 1082631000119106, 1082661000119103, 1082701000119105,

1082711000119108, 1082751000119109, 1082761000119106, 1082771000119100, 1083751000119104, 1084511000119108, 1084521000119101, 1084751000119101, 1084831000119100, 1084841000119109, 1084891000119101, 1084901000119102, 1084911000119104, 1084931000119109, 1084971000119107, 1085021000119106, 1085121000119107, 1085131000119105, 1085151000119104, 1085161000119102, 1085171000119108, 1085181000119106, 1085201000119107, 1085211000119105, 1085221000119103, 1085231000119100, 1085251000119106, 1085261000119108, 1085271000119102, 1085291000119101, 1085301000119100, 1085331000119107, 1085341000119103, 1085351000119101, 1085391000119106, 1085411000119106, 1085421000119104, 1085431000119101, 1085441000119105, 1085451000119107, 1085461000119109, 1085471000119103, 1085481000119100, 1085491000119102, 1085501000119109, 1085511000119107, 1085521000119100, 1085531000119102, 1085541000119106, 1085551000119108, 1085561000119105, 1085571000119104, 1085581000119101, 1085591000119103, 1085601000119105, 1085611000119108, 1085621000119101, 1085631000119103, 1085651000119109, 1085681000119102, 1085751000119100, 1085771000119109, 1085781000119107, 1085791000119105, 1085801000119106, 1085821000119102, 1085831000119104, 1085841000119108, 1085871000119101, 1085881000119103, 1085891000119100, 1085931000119108, 1085941000119104, 1085951000119102, 1085961000119100, 1085971000119106, 1085981000119109, 1085991000119107, 1086001000119108, 1086031000119101, 1086041000119105, 1086111000119105, 1086151000119106, 1086191000119101, 1086231000119105, 1086241000119101, 1086281000119106, 1086291000119109, 1086331000119103, 1086341000119107, 1086381000119102, 1086441000119100, 1086461000119101, 1086481000119105, 1086491000119108, 1086541000119104, 1086551000119102, 1086561000119100, 1086571000119106, 1086581000119109, 1086591000119107, 1086601000119100, 1086751000119105, 1086761000119107, 1086771000119101, 1086791000119100, 1086811000119101, 1086821000119108, 1086831000119106, 1086841000119102, 1086951000119108, 1086961000119105, 1087001000119105, 1087091000119104, 1087101000119109, 1087111000119107, 1087161000119105, 1087271000119105, 1087811000119103, 1089981000119106, 1089991000119109, 1090051000119109, 1090061000119106, 1090071000119100, 1090081000119102, 1090091000119104, 1090101000119109, 1090111000119107, 1090121000119100, 1090131000119102, 1090141000119106, 1090151000119108, 1090161000119105, 1090171000119104, 1090181000119101, 1090191000119103, 1090211000119102, 1090221000119109, 1090321000119103, 1090361000119108, 1090391000119101, 1090401000119104, 1092381000119100, 1092801000119102, 1092841000119100, 1092861000119101, 1092871000119107, 1092881000119105, 1092901000119107, 1092941000119109, 2260001000004104, 7050001000004104, 10628871000119101, 10628911000119103, 10629231000119109, 10629271000119107, 10690671000119109, 10743231000119101, 10750161000119106, 10754781000119108, 10759111000119102, 10759151000119101, 10807061000119103, 10898091000119108, 10898171000119108, 10898371000119105, 10899011000119105, 10899131000119104, 10899251000119103, 10899371000119104, 10998911000119105, 10999201000119106, 11047881000119101, 11048011000119103, 11315361000119100, 11718971000119100, 11796681000119109, 11796731000119100, 11798951000119105, 11798991000119100, 11799361000119102, 11799401000119106, 11799811000119107, 11799851000119108, 11808291000119106, 11808371000119102, 11808411000119101, 11810901000119105, 11812221000119109, 11847161000119101, 11847211000119109, 11847251000119105, 11847291000119100, 11847411000119105, 11847781000119106, 11847941000119103, 12107971000119108, 12108021000119109, 12108141000119104, 12203031000119103, 12237271000119105, 12237311000119105, 12242631000119107, 12242671000119105, 12860001000004105, 13008751000119103, 13440001000004103, 13560001000004104, 13570001000004105, 14160001000004107, 14280001000004108, 14720231000119109, 15641141000119100, 15643101000119103, 15648731000119101, 15648771000119103, 15648821000119105, 15710201000119100, 15710241000119103, 15711921000119107, 15711961000119102, 15712001000119106, 15749591000119107, 15918061000119103, 15918101000119100, 15918221000119100, 15918261000119105, 15918301000119102, 15918341000119100, 15926411000119101,

15926471000119109, 15966341000119102, 15966381000119107, 15966461000119108, 15966501000119108, 15966701000119103, 15966741000119101, 15966781000119106, 15969461000119103, 15971181000119105, 15971541000119105, 15971701000119103, 15977991000119101, 15983591000119109, 16026071000119107, 16026111000119100, 16055151000119105, 16098491000119109, 16468941000119103, 16697021000119105, 16697101000119102, 16715971000119108, 16833871000119109, 16834011000119109, 16834071000119101, 16894791000119107, 16899261000119106, 24920001000004101, 29340001000004108, 81632821000132105, 206335051000132108, 355468491000132106, 402503301000132107, 550376571000132101, 589157321000132102, 625544171000132109, 695751021000132104, 842853311000132105, 903952141000132105, 1779005, 2707005, 3464007, 3815005, 4283007, 4416007, 5667009, 7265005, 7493006, 7620006, 7826003, 7877005, 8146009, 8933000, 9866007, 9936001, 10078003, 10679007, 11179002, 11840006, 13471002, 18849004, 19894004, 21638000, 22208002, 22905009, 22922006, 23688003, 28016005, 28599006, 28951000, 29604006, 29979000, 30493003, 30716008, 32298001, 33410002, 33995003, 34240005, 36529003, 37373007, 37832003, 38106008, 38302000, 38662009, 39755000, 40158001, 40453004, 41527003, 41545003, 42357009, 43017006, 44047000, 44553005, 45564002, 46288008, 46420000, 48241004, 48763007, 50440006, 50450007, 50584008, 50711007, 51254007, 52918004, 53132006, 53435002, 53462005, 54034003, 54266002, 54411001, 55821006, 56013008, 56200009, 56287005, 56689002, 57920007, 58037000, 59032001, 59158004, 59192001, 60570001, 60876000, 61424003, 61665008, 61977001, 62078006, 66622006, 66937008, 69163003, 69810009, 70014009, 70385007, 70622003, 71085009, 71572001, 71833008, 71913005, 72074009, 72779005, 72965009, 73146005, 75169006, 75964002, 76304001, 76341000, 76623002, 76795007, 78482005, 79962008, 80126007, 81020007, 81780002, 82385007, 82930004, 84739008, 86443005, 87352006, 88518009, 89538001, 90271007, 90281006, 90549003, 91155009, 91390005, 91970006, 92212001, 92487001, 92530007, 92656007, 92660005, 92803008, 93890009, 94144008, 94365007, 94407000, 94682004, 94930003, 95184009, 105656003, 109389001, 109487003, 109493006, 109494000, 109500001, 109502009, 109504005, 109558001, 109559009, 109778006, 109816005, 111325003, 111839008, 126766000, 126836001, 127226007, 128333008, 128398001, 163174003, 186138007, 186963008, 187716008, 187734007, 187752007, 196542004, 196592003, 196607008, 196977009, 197269008, 201517005, 204658006, 204687007, 204688002, 204739008, 204740005, 204741009, 230194002, 232374009, 232417005, 232427004, 235087000, 235112000, 235640006, 235664007, 235745006, 235758007, 235796008, 235895002, 235952002, 236025002, 236031004, 239023005, 240349003, 240351004, 240367005, 240554006, 240792005, 253017000, 253756002, 266429005, 266433003, 270908003, 275782008, 277537008, 286609007, 302914006, 303672007, 304552000, 312084009, 315058005, 320934008, 360434004, 363510005, 397172008, 397868007, 398050005, 399525009, 402378006, 402446002, 402901009, 403252006, 403730001, 403947009, 404810003, 404907009, 410485009, 414153008, 414154002, 414376003, 414530001, 414604009, 415724006, 415822001, 420321004, 420549007, 420687005, 421230000, 421454008, 421710003, 421983003, 424779008, 426507006, 428874003, 429430001, 445152004, 446122002, 446328009, 446594000, 446642005, 446672004, 446752000, 446754004, 446755003, 446756002, 446988001, 447282003, 447354007, 447372001, 447821002, 700455003, 702577006, 702969000, 703535000, 711167005, 717267005, 724437007, 733150001, 735387004, 780821007, 783787000, 103691000119106, 123721000119108, 123731000119106, 123741000119102, 123751000119100, 123761000119103, 142961000119108, 143401000119106, 192681000119104, 192701000119101, 1082721000119101, 1082731000119103, 1085851000119105, 1085901000119101, 1087081000119102, 1089971000119108, 1197717008, 444258003, 444259006, 444546002, 444548001, 444597005, 444667006, 444673007, 444707001, 444818007, 444819004, 444898006, 444926003, 445090007, 445130008, 445162006, 445163001, 445243001, 445257004, 445306000, 445507008, 445554007, 445736006, 445737002, 445848002, 445874008, 445965007, 446026002, 446027006, 446076000, 446081009, 446124001, 446189008, 446251006, 446334002, 446336000, 446390008, 446391007, 446392000, 446430005, 446431009, 446465005, 446615009, 446673009, 446698005, 446753005, 446807009, 446836009, 446896007, 446915006,

446925001, 446928004, 447004005, 447058001, 447072005, 447111007, 447109003, 447320006, 447325001, 447371008, 447416008, 447432006, 447656001, 447658000, 447707005, 447712006, 447731000, 447766003, 447781009, 447783007, 447785000, 447791003, 447805007, 447806008, 447823004, 447851008, 447886005, 447888006, 447917005, 447952002, 447990008, 447991007, 448071000, 448081001, 448131008, 448147005, 448164008, 448211002, 448214005, 448229007, 448258005, 448269008, 448281007, 448286002, 448315008, 448319002, 448354009, 448371005, 448373008, 448386004, 448428002, 448450001, 448468003, 448493001, 448495008, 448542008, 448550004, 448551000, 448555009, 448663003, 448664009, 448665005, 448667002, 448674007, 448675008, 448683002, 448709005, 448868009, 448908007, 448992002, 448994001, 448997008, 449014008, 449015009, 449016005, 449034009, 449053004, 449055006, 449058008, 449063007, 449072004, 449074003, 449075002, 449153001, 449156009, 449173006, 449174000, 449176003, 449218003, 449219006, 449222008, 449223003, 449248000, 449254004, 449259009, 449260004, 449289002, 449292003, 1197715000, 449377002, 449410007, 449416001, 449419008, 449472007, 449497006, 449516003, 449578008, 449763003, 449764009, 449768007, 449770003, 449771004, 449772006, 449773001, 449774007, 449776009, 449780004, 449782007, 449785009, 449788006, 449789003, 449791006, 449798000, 449805002, 449806001, 449807005, 449808000, 449809008, 449815008, 449833002, 449847003, 449848008, 449855005, 449874002, 449875001, 449908004, 450849003, 450878002, 450880008, 451030007, 458422009, 459054000, 460365008, 460375006, 471284004, 472978005, 473281000, 473392002, 473418001, 473419009, 609201007, 609454008, 609467006, 609468001, 609473007, 609474001, 609475000, 609501008, 609504000, 609505004, 697952006, 697960007, 697965002, 697969008, 697977007, 697991001, 698004001, 698005000, 698008003, 698010001, 698011002, 698019000, 698030002, 698032005, 698037004, 698039001, 698041000, 698042007, 698043002, 698044008, 698045009, 698048006, 698051004, 698053001, 698055008, 698070009, 698071008, 698072001, 698073006, 698182002, 698185000, 698186004, 698189006, 698192005, 698194006, 698199001, 698257008, 698287002, 698294004, 698317008, 698318003, 698352000, 698518004, 698522009, 698523004, 698524005, 698525006, 698733009, 698769001, 698771001, 698772008, 698779004, 698861005, 698862003, 698962002, 698963007, 699050007, 699129001, 699189004, 699192000, 699194004, 699197006, 699222000, 699235009, 699274002, 699276000, 699277009, 699290002, 699319004, 699323007, 699382004, 699384003, 699418008, 699421005, 699422003, 699423008, 699424002, 699444008, 699483005, 699489009, 699490000, 699491001, 699492008, 699494009, 699495005, 699683004, 699684005, 699700006, 699701005, 699718009, 699756005, 699762000, 699773007, 699777008, 699785004, 699816004, 699852000, 699853005, 700046006, 700080009, 700237005, 700238000, 700257009, 700283004, 700285006, 700423003, 700425005, 700463002, 702360007, 702399004, 702402003, 702432006, 702646000, 702712006, 702713001, 702714007, 702958003, 702959006, 703089009, 703095005, 703132007, 703134008, 703148008, 703271000, 703315000, 703339005, 703401001, 703406006, 703468005, 703525006, 703651000, 703866000, 705127009, 706984006, 707129008, 707135008, 707167006, 707183006, 707201003, 707219007, 707222009, 707235000, 707237008, 707238003, 707240008, 707243005, 707245003, 707247006, 707248001, 707249009, 707250009, 707251008, 707252001, 707253006, 707255004, 707256003, 707268007, 707296001, 707297005, 707301001, 707302008, 707310009, 707314000, 707318002, 707320004, 707341005, 707352004, 707395000, 707396004, 707397008, 707398003, 707399006, 707400004, 707401000, 707402007, 707406005, 707450006, 707481002, 707482009, 707483004, 707484005, 707485006, 707486007, 707487003, 707489000, 707490009, 707491008, 707492001, 707524009, 707525005, 707528007, 707529004, 707532001, 707535004, 707536003, 707537007, 707538002, 707539005, 707578009, 707579001, 707580003, 707581004, 707582006, 707583001, 707584007, 707585008, 707586009, 707587000, 707588005, 707589002, 707590006, 707591005, 707592003, 707593008, 707606004, 707607008, 707608003, 707609006, 707623008, 707627009, 707628004, 707679005, 707681007, 707686002, 707697002, 707703001, 707704007, 707705008, 707733008, 707753007, 707766007, 707792000, 707796002, 707985000, 707986004, 707989006, 707994006, 708013001, 708016009, 708055005, 708164002, 708165001, 708198006, 708248004, 708463003, 708494003, 708508006, 708512000, 708653004, 708654005, 708674003, 708791005, 708964003, 709003000,

709031009, 709062008, 709146000, 709147009, 709287005, 709424004, 709465004, 709466003, 709469005, 709471005, 709472003, 709517003, 709535007, 709556009, 709557000, 709561006, 709604005, 709608008, 709667001, 709668006, 709830006, 709852002, 709977002, 709999005, 710003004, 710088005, 710089002, 710090006, 710096000, 710105009, 710298005, 710572000, 710727006, 710728001, 710729009, 710735009, 710782002, 710786004, 710789006, 710903001, 710926008, 710927004, 711040002, 711100005, 711103007, 711171008, 711173006, 711174000, 711175004, 711368009, 711441008, 711497006, 711499009, 711564007, 711638008, 711639000, 711640003, 711641004, 712488005, 712509002, 712510007, 712511006, 712512004, 712513009, 712514003, 712515002, 712583003, 712646007, 712722007, 712728006, 712730008, 712731007, 712977006, 712979009, 712988000, 713034001, 713036004, 713045003, 713180002, 713181003, 713186008, 713189001, 713197008, 713203006, 713211001, 713212008, 713213003, 713214009, 713215005, 713218007, 713225000, 713229006, 713236007, 713245008, 713249002, 713250002, 713251003, 713256008, 713261005, 713277006, 713297001, 713300006, 713306000, 713343003, 713344009, 713347002, 713354008, 713356005, 713370005, 713411004, 713497004, 713505000, 713510001, 713529007, 713546005, 713570009, 713573006, 713638002, 713654004, 713691005, 713698004, 713702000, 713703005, 713704004, 713720009, 713724000, 713825007, 713879003, 713966008, 714036005, 714037001, 714038006, 714073008, 714103006, 714113003, 714126004, 714128003, 714148005, 714149002, 714249007, 714253009, 714278008, 714477000, 714478005, 714485009, 714507001, 714514004, 714522006, 714633003, 714634009, 714635005, 714636006, 714746001, 714811003, 714817004, 714818009, 715062005, 715140008, 715192004, 715201005, 715212005, 715285004, 715286003, 715287007, 715397000, 715401008, 715414009, 715527006, 715577009, 715656004, 715669000, 715830008, 715834004, 715837006, 715838001, 715839009, 715840006, 715843008, 715852004, 715858000, 715859008, 715860003, 715864007, 715866009, 715952000, 715989002, 716007007, 716008002, 716063008, 716092007, 716117004, 716180009, 716183006, 716195006, 716196007, 716203000, 716230005, 716239006, 716248001, 716277000, 716280004, 716305005, 716362006, 716364007, 716379000, 716586009, 716590006, 716648006, 716649003, 716650003, 716651004, 716652006, 716665002, 716685003, 716701004, 716714003, 716859000, 716860005, 717044000, 717045004, 717046003, 717047007, 717048002, 717062009, 717155003, 717156002, 717182006, 717186009, 717187000, 717231003, 717232005, 717329009, 717698000, 717699008, 717814004, 717822006, 717823001, 717832004, 717833009, 717835002, 717841009, 717842002, 717843007, 717845000, 717846004, 717847008, 717848003, 717849006, 717858004, 717860002, 717861003, 717862005, 717863000, 717864006, 717865007, 717866008, 717867004, 717868009, 717869001, 717870000, 717871001, 717872008, 717873003, 717874009, 717875005, 717876006, 717877002, 717878007, 717879004, 717880001, 717881002, 717882009, 717883004, 717884005, 717886007, 717888008, 717889000, 717891008, 717892001, 717916003, 717917007, 717919005, 717951006, 717956001, 718052004, 718053009, 718060003, 718061004, 718062006, 718064007, 718065008, 718066009, 718068005, 718069002, 718075006, 718077003, 718106009, 718214007, 718309005, 718346009, 718471008, 718551002, 718552009, 718573009, 718575002, 718680001, 718681002, 719042007, 719044008, 719306007, 719408007, 719452004, 719454003, 719456001, 719468005, 719501003, 719687007, 719823007, 719833004, 719844007, 719910004, 719911000, 719945007, 719972004, 719973009, 720009004, 720394008, 720401009, 720461006, 720464003, 720507006, 720511000, 720578000, 720604008, 720636001, 720640005, 720746006, 720820000, 720980004, 721085000, 721089006, 721090002, 721091003, 721095007, 721152005, 721154006, 721155007, 721159001, 721160006, 721161005, 721178001, 721179009, 721182004, 721191000, 721193002, 721194008, 721206006, 721208007, 721221000, 721222007, 721223002, 721234004, 721235003, 721236002, 721286008, 721555001, 721557009, 721558004, 721559007, 721562005, 721568009, 721569001, 721570000, 721580001, 721586007, 721588008, 721607006, 721608001, 721609009, 721611000, 721612007, 721613002, 721614008, 721615009, 721616005, 721617001, 721618006, 721619003, 721620009, 721621008, 721622001, 721623006, 721624000, 721625004, 721626003, 721627007, 721628002, 721629005, 721630000, 721632008, 721633003, 721634009, 721635005, 721636006, 721637002, 721638007, 721639004, 721640002, 721641003, 721642005, 721643000, 721644006, 721645007, 721646008,

721647004, 721648009, 721649001, 721650001, 721651002, 721652009, 721653004, 721654005, 721655006, 721656007, 721658008, 721659000, 721660005, 721661009, 721662002, 721663007, 721664001, 721665000, 721666004, 721667008, 721668003, 721669006, 721670007, 721671006, 721672004, 721673009, 721674003, 721675002, 721676001, 721677005, 721678000, 721679008, 721680006, 721681005, 721682003, 721683008, 721684002, 721685001, 721686000, 721688004, 721689007, 721690003, 721691004, 721692006, 721693001, 721694007, 721695008, 721696009, 721697000, 721698005, 721699002, 721700001, 721701002, 721702009, 721703004, 721704005, 721705006, 721706007, 721707003, 721708008, 721709000, 721710005, 721711009, 721712002, 721713007, 721715000, 721716004, 721717008, 721718003, 721719006, 721720000, 721721001, 721722008, 721723003, 721724009, 721725005, 721745002, 721771003, 721808004, 721834007, 721836009, 721843003, 721846006, 721847002, 721873007, 721880009, 722031003, 722034006, 722051004, 722060007, 722061006, 722062004, 722063009, 722064003, 722075004, 722092008, 722105002, 722106001, 722200003, 722201004, 722206009, 722207000, 722280000, 722288007, 722296002, 722377004, 722386009, 722392003, 722445001, 722449007, 722509001, 722510006, 722514002, 722516000, 722519007, 722521002, 722529000, 722530005, 722531009, 722532002, 722533007, 722534001, 722535000, 722536004, 722537008, 722538003, 722539006, 722540008, 722541007, 722542000, 722543005, 722544004, 722577000, 722672002, 722673007, 722674001, 722677008, 722713005, 722764006, 722781002, 722834005, 722841004, 722844007, 722845008, 722846009, 722847000, 722848005, 722849002, 722850002, 722852005, 722853000, 722854006, 722855007, 722856008, 722857004, 722858009, 722861005, 722864002, 722865001, 722866000, 722868004, 722869007, 722870008, 722871007, 722872000, 722873005, 722874004, 722875003, 722876002, 722877006, 722878001, 722879009, 722880007, 722881006, 722882004, 722883009, 722885002, 722886001, 722916005, 722933003, 723065009, 723079001, 723091005, 723096000, 723097009, 723098004, 723099007, 723100004, 723101000, 723102007, 723103002, 723104008, 723105009, 723106005, 723107001, 723108006, 723111007, 723177002, 723183004, 723265000, 723281005, 723360007, 723366001, 723383005, 723403008, 723442008, 723496007, 723581006, 723583009, 723584003, 723610009, 723676007, 723829000, 723882007, 723883002, 723884008, 723975009, 724000006, 724068001, 724276006, 724278007, 724361001, 724390007, 724454004, 724455003, 724457006, 724459009, 724473000, 724474006, 724476008, 724477004, 724511006, 724512004, 724514003, 724515002, 724516001, 724517005, 724519008, 724520002, 724521003, 724522005, 724523000, 724524006, 724525007, 724526008, 724527004, 724528009, 724529001, 724530006, 724531005, 724532003, 724533008, 724534002, 724535001, 724536000, 724537009, 724538004, 724540009, 724766009, 724768005, 725026008, 725027004, 725149008, 725167001, 725416005, 725591002, 725873003, 725910009, 725938001, 725939009, 725940006, 726101009, 726613003, 726654006, 726672000, 726703000, 732247000, 733034007, 733069009, 733071009, 733072002, 733145007, 733146008, 733147004, 733148009, 733149001, 733153004, 733154005, 733155006, 733156007, 733157003, 733158008, 733159000, 733160005, 733161009, 733162002, 733163007, 733164001, 733169006, 733172004, 733343005, 733344004, 733345003, 733346002, 733347006, 733348001, 733349009, 733350009, 733351008, 733352001, 733353006, 733354000, 733355004, 733356003, 733384008, 733385009, 733447005, 733468006, 733533008, 733605002, 733622000, 733628001, 733657002, 733939000, 733940003, 733941004, 733942006, 733943001, 733944007, 733945008, 733946009, 733947000, 733948005, 733949002, 733950002, 733951003, 733952005, 733954006, 733955007, 733957004, 733958009, 733961005, 733962003, 733968004, 733970008, 733971007, 733972000, 733973005, 733974004, 733975003, 733976002, 733977006, 733978001, 733980007, 733982004, 734018003, 734019006, 734024009, 734041006, 734839006, 734884008, 735129008, 735389001, 735390005, 735391009, 735425008, 735426009, 735427000, 735433009, 735451005, 735455001, 735456000, 735459007, 735467004, 735493006, 735496003, 735499005, 735514001, 735518003, 735546001, 735591005, 735592003, 735593008, 735595001, 735677007, 735718000, 735719008, 735720002, 735721003, 735722005, 735723000, 735724006, 735725007, 735726008, 735727004, 735729001, 735731005, 735732003, 735733008, 735734002, 735735001, 735736000, 735783003, 735784009, 735785005, 735786006, 735787002, 735788007, 736147006, 736434005, 736650005, 736724002, 736825009, 736826005,

736827001, 737058005, 737166003, 737187008, 737189006, 737190002, 737191003, 737192005, 737193000, 737194006, 737195007, 737196008, 737197004, 737198009, 737199001, 737200003, 737201004, 737202006, 737203001, 737204007, 737205008, 737211006, 737245004, 737265006, 737308008, 737309000, 737310005, 737311009, 737312002, 737313007, 737315000, 737316004, 737317008, 737345002, 737557007, 738791006, 738792004, 739032003, 741061001, 741515006, 741676008, 741677004, 742277008, 746223006, 758673004, 762272006, 762273001, 762274007, 762276009, 762277000, 762278005, 762279002, 762403002, 762404008, 762405009, 762406005, 762407001, 762408006, 762409003, 762410008, 762580003, 762581004, 762582006, 762583001, 762584007, 762585008, 762586009, 762587000, 762588005, 762608003, 762660007, 762665002, 763066009, 763096004, 763109002, 763134002, 763213001, 763397001, 763398006, 763498007, 763499004, 763536006, 763620003, 763630007, 763658004, 763686007, 763727005, 763728000, 763733001, 763734007, 763735008, 763753002, 763754008, 763828007, 763829004, 763833006, 763834000, 763835004, 763837007, 763891005, 764091001, 764092008, 764095005, 764487004, 764735002, 764808002, 764809005, 764845008, 764962002, 764963007, 764995008, 765480005, 765482002, 765741003, 765756007, 766249007, 766757006, 766761000, 766813000, 766942007, 766943002, 766944008, 766978002, 766979005, 766980008, 766981007, 767031008, 767032001, 767060004, 767291004, 767671002, 767676007, 767677003, 767678008, 767679000, 767680002, 767681003, 767684006, 767685007, 767686008, 767687004, 767748008, 767809001, 767810006, 767815001, 767823004, 767824005, 767826007, 767827003, 767855004, 767995003, 767996002, 768001004, 768006009, 768010007, 768012004, 768083003, 768084009, 768085005, 768086006, 768095003, 768098001, 768125005, 768126006, 768127002, 768288001, 768289009, 768639006, 768641007, 768642000, 768643005, 768646002, 768648001, 768649009, 768653006, 768654000, 768840005, 768841009, 768842002, 769014008, 769017001, 769020009, 769021008, 769023006, 769049001, 769087009, 769113004, 769114005, 769140004, 769141000, 769142007, 769143002, 769158005, 769159002, 769160007, 769161006, 770356006, 770592009, 770593004, 770602005, 770619007, 770684008, 770685009, 770779009, 770780007, 770781006, 770782004, 770900000, 770943008, 770944002, 771013004, 771149000, 771185000, 771240009, 771265006, 771333006, 771335004, 771352000, 771441005, 771446000, 771474005, 772139001, 772140004, 772141000, 772142007, 772143002, 772144008, 772149003, 772780004, 772781000, 772782007, 772783002, 772792004, 772821007, 773094000, 773095004, 773113008, 773200006, 773269009, 773270005, 773272002, 773279006, 773302000, 773327007, 773332008, 773345007, 773397000, 773415005, 773553003, 773579007, 773584001, 773620002, 773623000, 773649005, 773660001, 773661002, 773662009, 773666007, 773673002, 773692000, 773726000, 773737004, 773750003, 773770009, 774151000, 774204006, 774207004, 777998000, 781076008, 781201007, 781203005, 781382000, 782504002, 782505001, 782666006, 782697005, 782771007, 782780007, 782866008, 782935003, 782945001, 782946000, 783059004, 783155007, 783159001, 783183009, 783185002, 783187005, 783195009, 783253009, 783565007, 783704005, 783706007, 783730009, 783734000, 783737007, 783738002, 783740007, 783741006, 783767001, 783768006, 783771003, 784346006, 784429009, 785696007, 785704008, 785727000, 785798002, 785824001, 785834005, 785835006, 785879009, 786039009, 786068005, 163138007, 163139004, 163145007, 163148009, 163152009, 163160005, 163162002, 163163007, 163169006, 163170007, 163171006, 163172004, 163173009, 163255001, 163256000, 163317009, 163318004, 163319007, 163320001, 163321002, 164204004, 164256007, 164260005, 168036006, 168037002, 168038007, 168047004, 168048009, 168049001, 172697005, 173599005, 186116005, 186134009, 186136006, 186137002, 186150001, 186225008, 186272008, 186273003, 186357007, 186394003, 186395002, 186431008, 186537009, 186562009, 186626002, 186639003, 186659004, 186675001, 186698009, 186731007, 186815006, 186931002, 186939000, 187062006, 187068005, 187084006, 187093007, 187100003, 187101004, 187145004, 187171000, 187174008, 187187002, 187197006, 187222000, 187601000, 187604008, 187606005, 187608006, 187613005, 187614004, 187622006, 187631006, 187633009, 187634003, 187635002, 187637005, 187640005, 187641009, 187644001, 187652003, 187653008, 187658004, 187659007, 187660002, 187661003, 187662005, 187675005, 187681002, 187688008, 187692001, 187693006, 187694000, 187697007, 187700006, 187701005, 187708004, 187709007, 187722004,

187723009, 187724003, 187725002, 187726001, 187727005, 187732006, 187733001, 187736009, 187738005, 187740000, 187741001, 187742008, 187757001, 187760008, 187767006, 187769009, 187773007, 187776004, 187777008, 187784000, 187786003, 187791002, 187792009, 187793004, 187794005, 187798008, 187801002, 187803004, 187809000, 187810005, 187811009, 187814001, 187824009, 188030005, 188204000, 188445006, 188691005, 188777005, 188778000, 188779008, 188780006, 188781005, 188795008, 188796009, 188798005, 188800003, 188802006, 188803001, 188805008, 188838004, 188844000, 188845004, 188846003, 188848002, 188857008, 188859006, 188861002, 188862009, 188863004, 188868008, 189120005, 189212001, 189229003, 189231007, 189243008, 189246000, 189361002, 189362009, 189365006, 189371000, 189392000, 189397006, 189399009, 189401003, 189406008, 189814006, 190429008, 190437000, 190440000, 190823004, 190944000, 191296000, 191970005, 191983006, 192450008, 195289005, 195438008, 195469007, 195470008, 195474004, 195475003, 195476002, 195655000, 195656004, 195657008, 195658003, 195659006, 195660001, 195662009, 195663004, 195666007, 195667003, 195668008, 195669000, 195670004, 195671000, 195672007, 195673002, 195676005, 195677001, 195709006, 195779005, 195780008, 195782000, 195797002, 195798007, 195803003, 195804009, 195834004, 195924009, 195929004, 196273001, 196278005, 196279002, 196280004, 196286005, 196287001, 196298000, 196299008, 196301001, 196302008, 196305005, 196309004, 196317007, 196318002, 196319005, 196320004, 196333005, 196341005, 196356001, 196366009, 196367000, 196368005, 196376007, 196398003, 196403002, 196406005, 196407001, 196412000, 196413005, 196414004, 196416002, 196417006, 196419009, 196427000, 196439008, 196463000, 196474009, 196475005, 196476006, 196479004, 196485006, 196486007, 196487003, 196489000, 196491008, 196492001, 196493006, 196495004, 196496003, 196497007, 196499005, 196500001, 196501002, 196502009, 196503004, 196504005, 196517008, 196518003, 196519006, 196521001, 196522008, 196523003, 196537007, 196538002, 196548000, 196556002, 196568006, 196572005, 196575007, 196578009, 196579001, 196599007, 196601009, 196602002, 196609006, 196617003, 196623008, 196632005, 196639001, 196652006, 196658005, 196666001, 196671008, 196682000, 196687006, 196694009, 196707000, 196712004, 196719008, 196731005, 196735001, 196740009, 196746003, 196753007, 196754001, 196757008, 196760001, 196761002, 196764005, 196767003, 196775009, 196781001, 196789004, 196792000, 196807006, 196809009, 196812007, 196813002, 196835005, 196837002, 196840002, 196841003, 196856007, 196859000, 196861009, 196863007, 196864001, 196865000, 196880006, 196885001, 196890003, 196903009, 196904003, 196915008, 196916009, 196917000, 196918005, 196923005, 196928001, 196933002, 196942009, 196943004, 196950000, 196955005, 196956006, 196957002, 196976000, 196983007, 196987008, 196999001, 197000003, 197001004, 197006009, 197011006, 197012004, 197013009, 197024006, 197028009, 197030006, 197046003, 197047007, 197049005, 197050005, 197051009, 197052002, 197053007, 197054001, 197056004, 197058003, 197059006, 197060001, 197063004, 197078006, 197079003, 197083003, 197084009, 197085005, 197091007, 197092000, 197104005, 197105006, 197106007, 197107003, 197112002, 197118003, 197120000, 197125005, 197127002, 197130009, 197132001, 197139005, 197140007, 197141006, 197143009, 197151007, 197152000, 197155003, 197162007, 197163002, 197164008, 197171003, 197172005, 197185008, 197191005, 197192003, 197193008, 197194002, 197195001, 197197009, 197198004, 197199007, 197200005, 197201009, 197210001, 197213004, 197216007, 197230002, 197233000, 197238009, 197244008, 197247001, 197248006, 197250003, 197259002, 197262004, 197268000, 197270009, 197271008, 197274000, 197275004, 197279005, 197284004, 197286002, 197291001, 197293003, 197294009, 197296006, 197299004, 197300007, 197301006, 197303009, 197305002, 197310003, 197315008, 197316009, 197321007, 197324004, 197326002, 197327006, 197328001, 197329009, 197330004, 197339003, 197342009, 197344005, 197347003, 197348008, 197352008, 197355005, 197356006, 197358007, 197359004, 197360009, 197361008, 197362001, 197364000, 197367007, 197368002, 197377009, 197378004, 197389006, 197391003, 197402000, 197410004, 197416005, 197418006, 197419003, 197422001, 197426003, 197432008, 197433003, 197438007, 197439004, 197441003, 197442005, 197446008, 197453004, 197458008, 197460005, 197464001, 197484002, 197490003, 197491004, 197493001, 197494007, 197495008, 197712008, 197772000, 197808006, 197812000, 197819009, 197824007, 197861003,

198156004, 198159006, 198185005, 198251001, 198273008, 198285000, 198286004, 198626009, 199022003, 199023008, 199025001, 199027009, 199028004, 199117000, 199118005, 199203001, 199930000, 199931001, 200190007, 200191006, 200249004, 200251000, 200252007, 200253002, 200254008, 200255009, 201805000, 201807008, 203070001, 204508009, 204512003, 204608004, 204614006, 204615007, 204616008, 204617004, 204620007, 204629008, 204630003, 204631004, 204635008, 204642008, 204644009, 204647002, 204652007, 204659003, 204667006, 204670005, 204671009, 204676004, 204691002, 204695006, 204696007, 204697003, 204699000, 204700004, 204702007, 204708006, 204709003, 204711007, 204712000, 204715003, 204716002, 204717006, 204721004, 204723001, 204724007, 204728005, 204731006, 204736001, 204745000, 204748003, 204750006, 204762005, 204769001, 204781002, 204782009, 204787003, 204788008, 204790009, 204795004, 204797007, 204798002, 204799005, 204805004, 204806003, 204808002, 204826004, 204847000, 204848005, 204905003, 205016000, 205548006, 205817005, 205836000, 206242003, 206245001, 206358003, 206372007, 206406008, 206407004, 206408009, 206423004, 206424005, 206425006, 206523001, 206525008, 206528005, 206529002, 207794006, 207798009, 302961007, 302971009, 303012000, 303059007, 303062005, 303078007, 303082009, 303086007, 304068004, 304370001, 304371002, 304518003, 304519006, 304520000, 304529004, 304545002, 304984001, 304985000, 304986004, 304987008, 304988003, 304989006, 304990002, 306785007, 306852006, 306857000, 306963008, 307000000, 307131005, 307132003, 307133008, 307198001, 307216009, 307218005, 307227006, 307228001, 307233002, 307234008, 307235009, 307236005, 307239003, 307295007, 307296008, 307297004, 307403007, 307404001, 307410001, 307411002, 307418008, 307420006, 307421005, 307423008, 307424002, 307425001, 307502000, 307519003, 307601000, 307757001, 307759003, 307768001, 307772002, 308119005, 308129003, 308140006, 308212008, 308213003, 308853007, 308855000, 308856004, 308857008, 308858003, 308859006, 308860001, 308861002, 308862009, 308863004, 308864005, 308865006, 308866007, 308867003, 308868008, 308870004, 308871000, 308872007, 308873002, 308875009, 308876005, 308878006, 308879003, 308882008, 308887002, 308888007, 308904008, 309084001, 309734000, 309751007, 309752000, 309783001, 311366001, 311808009, 312088007, 312104005, 312108008, 312109000, 312110005, 312111009, 312112002, 312113007, 312114001, 312115000, 312121001, 312128007, 312129004, 312131008, 312153005, 312158001, 312387008, 312388003, 312422001, 312590001, 312607004, 312655004, 312823001, 312895004, 313256001, 313325007, 313326008, 313327004, 313359006, 313425006, 314049009, 314601002, 314602009, 314944001, 314945000, 314947008, 314949006, 314951005, 314952003, 314960002, 314961003, 314962005, 314963000, 314964006, 314965007, 314966008, 315057000, 319297003, 324618004, 326072005, 328313007, 328383001, 331987008, 332982000, 333952004, 334831000, 335443002, 337471007, 338486003, 353821005, 355736000, 359613008, 359651008, 359653006, 359656003, 359660000, 359664009, 359692004, 359757004, 359793002, 359797001, 359799003, 359804008, 360339005, 360375007, 360425009, 360428006, 360431007, 360441005, 360489001, 360491009, 360494001, 360522009, 360525006, 361141006, 361142004, 361212009, 361264003, 363067009, 363080007, 363132006, 363219007, 363348004, 363349007, 363350007, 363351006, 363352004, 363353009, 363360003, 363368005, 363369002, 363370001, 363372009, 363373004, 363374005, 363375006, 363376007, 363377003, 363378008, 363379000, 363380002, 363381003, 363382005, 363383000, 363384006, 363385007, 363386008, 363388009, 363389001, 363391009, 363392002, 363393007, 363394001, 363395000, 363398003, 363399006, 363400004, 363401000, 363402007, 363403002, 363404008, 363405009, 363406005, 363407001, 363408006, 363409003, 363410008, 363411007, 363412000, 363413005, 363414004, 363415003, 363416002, 363417006, 363418001, 363419009, 363421004, 363456000, 363485006, 363490009, 363491008, 363492001, 363505006, 363507003, 363508008, 363509000, 363745004, 363746003, 367403001, 367431005, 367520004, 367534004, 369445005, 369446006, 369447002, 369448007, 369449004, 369450004, 369451000, 369452007, 369453002, 369454008, 369455009, 369456005, 369457001, 369458006, 369459003, 369460008, 369461007, 369535006, 369536007, 369537003, 369538008, 369539000, 369540003, 370402009, 370472005, 370484007, 370485008, 370486009, 370487000, 370488005, 370489002, 370490006, 370492003, 370510007, 370518000, 370526008, 370889009, 371000004, 371067004, 371083004, 371084005,

371090009, 371101003, 371132002, 371139006, 371423007, 371568002, 371962007, 371967001, 371968006,
371970002, 371974006, 371975007, 371976008, 371977004, 371978009, 371981004, 371984007, 371990006,
371992003, 371996000, 371997009, 371998004, 371999007, 372000001, 372001002, 372003004, 372004005,
372014001, 372015000, 372016004, 372017008, 372020000, 372022008, 372023003, 372026006, 372027002,
372101000, 372119009, 372138000, 372139008, 372140005, 372142002, 372143007, 373407002, 373588006,
373610002, 373617004, 373639002, 373643003, 385525003, 386211005, 386537007, 387665005, 387666006,
387754006, 389203001, 389273002, 210087005, 210109007, 210114006, 210116008, 210128000, 210130003,
210131004, 210132006, 210133001, 210135008, 210150004, 210155009, 210156005, 210157001, 210159003,
210160008, 210174001, 210175000, 210176004, 210262008, 210344002, 210358000, 210360003, 210361004,
210363001, 210364007, 210366009, 210368005, 210370001, 210376007, 210378008, 210395000, 210804004,
210809009, 210811000, 211044004, 211060000, 211109004, 211128006, 211144002, 211626006, 211631008,
211634000, 211635004, 211636003, 211637007, 211638002, 211639005, 211640007, 211641006, 211642004,
211644003, 212032004, 212035002, 212042002, 213111002, 1197721001, 213118008, 213153001, 213227002,
213230009, 213231008, 213241006, 213293008, 213294002, 215617000, 229692002, 229694001, 229726002,
229727006, 229728001, 230314007, 230326002, 230327006, 230328001, 231719009, 232371001, 232372008,
232373003, 232388008, 232399005, 232400003, 232401004, 232402006, 232403001, 232404007, 232405008,
232406009, 232407000, 232408005, 232409002, 232410007, 232411006, 232413009, 232415002, 232416001,
232418000, 232419008, 232420002, 232422005, 232423000, 232424006, 232425007, 232805002, 233005009,
233662009, 233870001, 234041004, 234054006, 234115007, 234200002, 234367000, 234505006, 234527002,
234528007, 234689009, 234947003, 234949000, 234951001, 234952008, 234953003, 234954009, 234955005,
234958007, 234960009, 234961008, 234962001, 234963006, 234964000, 234965004, 234966003, 234967007,
234968002, 234969005, 234970006, 234971005, 234972003, 234973008, 234974002, 234975001, 234976000,
234977009, 234978004, 234979007, 234980005, 234981009, 234982002, 234983007, 234984001, 234985000,
234986004, 234988003, 234992005, 234993000, 234994006, 234995007, 234996008, 234997004, 234998009,
234999001, 235000001, 235001002, 235002009, 235003004, 235004005, 235005006, 235006007, 235007003,
235008008, 235009000, 235010005, 235011009, 235012002, 235013007, 235014001, 235015000, 235016004,
235017008, 235018003, 235019006, 235021001, 235022008, 235023003, 235025005, 235026006, 235027002,
235028007, 235029004, 235030009, 235031008, 235032001, 235033006, 235034000, 235035004, 235036003,
235037007, 235038002, 235039005, 235040007, 235041006, 235042004, 235043009, 235044003, 235045002,
235046001, 235047005, 235048000, 235049008, 235050008, 235051007, 235052000, 235053005, 235054004,
235055003, 235056002, 235057006, 235058001, 235059009, 235061000, 235062007, 235064008, 235065009,
235066005, 235067001, 235068006, 235069003, 235070002, 235071003, 235072005, 235075007, 235085008,
235086009, 235088005, 235089002, 235090006, 235091005, 235092003, 235093008, 235094002, 235095001,
235096000, 235098004, 235099007, 235100004, 235101000, 235104008, 235110008, 235111007, 235115003,
235121004, 235122006, 235123001, 235124007, 235125008, 235126009, 235127000, 235128005, 235129002,
235130007, 235131006, 235132004, 235133009, 235134003, 235135002, 235136001, 235137005, 235138000,
235139008, 235140005, 235143007, 235224000, 235494005, 235594008, 235596005, 235597001, 235598006,
235599003, 235600000, 235601001, 235602008, 235603003, 235604009, 235605005, 235606006, 235607002,
235608007, 235609004, 235610009, 235611008, 235612001, 235613006, 235614000, 235615004, 235616003,
235617007, 235618002, 235619005, 235620004, 235621000, 235622007, 235623002, 235625009, 235626005,
235627001, 235628006, 235629003, 235630008, 235631007, 235632000, 235633005, 235634004, 235635003,
235636002, 235637006, 235638001, 235639009, 235641005, 235642003, 235644002, 235645001, 235646000,
235648004, 235650007, 235651006, 235652004, 235654003, 235655002, 235656001, 235657005, 235659008,
235660003, 235661004, 235662006, 235663001, 235666009, 235667000, 235669002, 235670001, 235671002,
235672009, 235673004, 235674005, 235675006, 235676007, 235677003, 235678008, 235679000, 235680002,
235681003, 235682005, 235683000, 235684006, 235685007, 235686008, 235688009, 235689001, 235690005,

235692002, 235693007, 235694001, 235696004, 235698003, 235699006, 235700007, 235702004, 235705002, 235706001, 235707005, 235708000, 235709008, 235710003, 235712006, 235713001, 235714007, 235715008, 235720008, 235722000, 235723005, 235724004, 235725003, 235726002, 235727006, 235728001, 235729009, 235730004, 235731000, 235732007, 235733002, 235734008, 235735009, 235736005, 235737001, 235738006, 235739003, 235741002, 235742009, 235743004, 235746007, 235747003, 235748008, 235749000, 235750000, 235751001, 235753003, 235754009, 235755005, 235756006, 235757002, 235759004, 235760009, 235761008, 235762001, 235763006, 235764000, 235765004, 235766003, 235767007, 235768002, 235769005, 235770006, 235774002, 235776000, 235777009, 235778004, 235779007, 235780005, 235781009, 235782002, 235783007, 235784001, 235785000, 235786004, 235787008, 235788003, 235790002, 235793000, 235794006, 235795007, 235797004, 235798009, 235799001, 235800002, 235805007, 235806008, 235809001, 235810006, 235811005, 235812003, 235813008, 235814002, 235815001, 235816000, 235817009, 235820001, 235821002, 235822009, 235823004, 235824005, 235825006, 235826007, 235827003, 235828008, 235829000, 235830005, 235831009, 235832002, 235833007, 235834001, 235835000, 235836004, 235837008, 235838003, 235839006, 235840008, 235842000, 235843005, 235844004, 235845003, 235846002, 235847006, 235848001, 235849009, 235851008, 235852001, 235853006, 235855004, 235858002, 235859005, 235862008, 235863003, 235864009, 235865005, 235866006, 235867002, 235869004, 235873001, 235874007, 235875008, 235876009, 235877000, 235878005, 235880004, 235881000, 235884008, 235885009, 235886005, 235887001, 235888006, 235889003, 235890007, 235896001, 235897005, 235898000, 235899008, 235901004, 235902006, 235903001, 235906009, 235908005, 235909002, 235910007, 235911006, 235912004, 235913009, 235915002, 235916001, 235917005, 235918000, 235919008, 235921003, 235923000, 235924006, 235925007, 235926008, 235927004, 235928009, 235929001, 235930006, 235931005, 235932003, 235933008, 235934002, 235935001, 235936000, 235938004, 235939007, 235940009, 235941008, 235942001, 235943006, 235944000, 235945004, 235946003, 235947007, 235948002, 235949005, 235950005, 235951009, 235953007, 235954001, 235955000, 235956004, 235957008, 235958003, 235959006, 235960001, 235961002, 235962009, 235966007, 235967003, 235968008, 235969000, 235971000, 235972007, 235973002, 235974008, 235975009, 235976005, 235977001, 235978006, 235979003, 235980000, 235982008, 235983003, 235985005, 235986006, 235987002, 235988007, 235989004, 235990008, 235997006, 236001005, 236004002, 236008004, 236009007, 236012005, 236013000, 236014006, 236015007, 236016008, 236017004, 236018009, 236019001, 236021006, 236022004, 236024003, 236026001, 236030003, 236032006, 236036009, 236042008, 236043003, 236044009, 236045005, 236047002, 236048007, 236049004, 236050004, 236053002, 236054008, 236055009, 236058006, 236059003, 236060008, 236063005, 236066002, 236069009, 236071009, 236074001, 236075000, 236078003, 236083006, 236085004, 236086003, 236087007, 236088002, 236089005, 236090001, 236091002, 236092009, 236093004, 236094005, 236095006, 236096007, 236097003, 236098008, 236099000, 236100008, 236101007, 236102000, 236103005, 236104004, 236105003, 236106002, 236107006, 236108001, 236109009, 236110004, 236111000, 236112007, 236113002, 236114008, 236115009, 236116005, 236117001, 236119003, 236120009, 236121008, 236122001, 236123006, 236124000, 236125004, 236126003, 236127007, 236128002, 236129005, 236130000, 236131001, 236132008, 236134009, 236135005, 236136006, 236137002, 236482006, 236488005, 236490006, 236491005, 236492003, 236493008, 236559006, 237045001, 237062006, 237063001, 237064007, 237065008, 237066009, 237198001, 237201006, 237202004, 237205002, 237206001, 237208000, 237575002, 237578000, 237579008, 237580006, 237581005, 237582003, 237583008, 237584002, 237585001, 237586000, 237587009, 237589007, 237590003, 237591004, 237592006, 237593001, 237608006, 237610008, 237612000, 237616002, 237617006, 237638000, 237639008, 237646004, 237647008, 237832001, 237964009, 238033007, 238035000, 238056003, 238057007, 238152004, 238155002, 238412006, 238583005, 238661000, 238662007, 238663002, 238664008, 238665009, 238666005, 238667001, 238706002, 238751002, 238803001, 238850005, 238908006, 238919009, 239006001, 239007005, 239009008, 239010003, 239011004, 239012006, 239013001, 239014007, 239015008, 239018005, 239019002, 239020008, 239021007, 239022000, 239024004, 239025003, 239026002, 239027006, 239028001, 239030004, 239031000,

239032007, 239040001, 239041002, 239042009, 239043004, 239056006, 239072003, 239093000, 239099001, 240110008, 240163000, 240332005, 240333000, 240335007, 240336008, 240337004, 240344008, 240350003, 240352006, 240353001, 240354007, 240355008, 240356009, 240357000, 240358005, 240363009, 240364003, 240365002, 240368000, 240369008, 240370009, 240371008, 240372001, 240374000, 240375004, 240376003, 240384004, 240437000, 240444009, 240477008, 240526004, 240527008, 240529006, 240547000, 240597001, 240659008, 240704003, 240729009, 240734008, 240736005, 240789006, 240790002, 240791003, 240796008, 240798009, 240800002, 240801003, 240802005, 240803000, 240804006, 240806008, 240861001, 240863003, 240864009, 240865005, 240869004, 240870003, 240872006, 241154007, 241861008, 242003008, 242007009, 242862004, 243286001, 243410003, 243978007, 244815007, 245754007, 249421000, 249423002, 249446000, 249603008, 249633001, 249643003, 249658006, 250037002, 251331003, 252283005, 253002004, 253005002, 253006001, 253018005, 253333008, 253741006, 253750008, 253752000, 253753005, 253754004, 253755003, 253757006, 253758001, 253760004, 253763002, 253764008, 253767001, 253768006, 253769003, 253770002, 253771003, 253772005, 253773000, 253774006, 253778009, 253780003, 253781004, 253782006, 253783001, 253784007, 253785008, 253786009, 253789002, 253791005, 253792003, 253793008, 253794002, 253795001, 253796000, 253797009, 253799007, 253800006, 253801005, 253802003, 253804002, 253806000, 253807009, 253808004, 253809007, 253810002, 253811003, 253812005, 253813000, 253814006, 253815007, 253826001, 253899000, 253909006, 34580000, 34589004, 34602004, 34736002, 34786008, 34798003, 34821005, 34903006, 34921009, 34923007, 34964002, 35023000, 35065006, 35074008, 35095003, 35117008, 35156002, 35217003, 35223008, 35246005, 35265002, 35266001, 35298007, 35423006, 35447004, 35452009, 35517004, 35541001, 35560008, 35563005, 35571009, 35580009, 35681000, 35865007, 35974005, 36001007, 36046008, 36151004, 36155008, 36162004, 36184004, 36188001, 36202009, 36246001, 36261000, 36269003, 36304008, 36334003, 36376006, 36483003, 36619004, 36631002, 36746002, 36760000, 36789003, 36891003, 36921006, 36975000, 37054000, 37224001, 37320007, 37369009, 37372002, 37404003, 37439003, 37442009, 37502002, 37528004, 37666005, 37688005, 37693008, 37703005, 37830006, 37831005, 37871000, 37976006, 37992001, 38032004, 38059007, 38079004, 38089000, 38163003, 38205001, 38214006, 38215007, 38228000, 38285004, 38306002, 38365000, 38397000, 38420005, 38438008, 38541002, 38632003, 38685005, 38712009, 38739001, 38851006, 38856001, 38906007, 38938002, 38951007, 38972005, 38982006, 38993008, 38996000, 39170005, 39204006, 39205007, 39271004, 39273001, 39291006, 39306006, 39323002, 39326005, 39341005, 39344002, 39392002, 39400004, 39476006, 39634006, 39642007, 39719008, 39747007, 39772007, 39788007, 39963006, 39998009, 40028009, 40046003, 40102009, 40159009, 40194002, 40210001, 40214005, 40273006, 40315008, 40444006, 40468003, 40494007, 40515007, 40650009, 40663003, 40719004, 40739000, 40766000, 40775003, 40835002, 40845000, 40857009, 40890009, 40912001, 41060007, 41152008, 41279003, 41349008, 41364008, 41366005, 41565005, 41569004, 41582007, 41585009, 41626001, 41806003, 41889008, 41916005, 41918006, 41986000, 42089007, 42140004, 42141000, 42290006, 42323001, 42324007, 42338000, 42541005, 42631002, 42640003, 42681006, 42698006, 42711005, 42740008, 42744004, 42746002, 42982001, 43030007, 43035002, 43043007, 43080001, 43162008, 43240000, 43337002, 43406003, 43411001, 43528001, 43633008, 43634002, 43694004, 43752006, 43797002, 43823002, 43841005, 43878008, 43904005, 43935004, 44017002, 44018007, 44283002, 44332000, 44402007, 44516004, 44572005, 44601009, 44606004, 44635007, 44636008, 44758003, 44917000, 45019006, 45042004, 45067001, 45256007, 45281005, 45299002, 45312009, 45338009, 45365002, 45414006, 45456005, 45485004, 45517002, 45534005, 45568004, 45629003, 45640006, 45676007, 45743004, 45798009, 45845003, 45941008, 45974008, 45982008, 45989004, 45994004, 46040000, 46276006, 46341007, 46395002, 46495001, 46516001, 46523000, 46672003, 46689006, 46708007, 46763001, 46795000, 46799006, 46882000, 46907007, 46948005, 46996005, 47028006, 47064007, 47152002, 47206001, 47222000, 47312008, 47367009,

47384003, 47417003, 47468000, 47481007, 47533003, 47548009, 47609003, 47693006, 47753002, 47771009, 47803004, 47812002, 47841006, 47932002, 47941007, 47944004, 47977000, 48000002, 48036004, 48124008, 48128006, 48153009, 48169002, 48186008, 48269007, 48303008, 48336009, 48413001, 48607003, 48644003, 48653005, 48658001, 48729005, 48819008, 48839007, 48916002, 48974009, 49120005, 49123007, 49206006, 49232000, 49237006, 49286008, 49321003, 49341008, 49414002, 49464009, 49678000, 49714001, 49723003, 49781004, 49813000, 49827005, 49885004, 49916007, 49965002, 50008003, 50167007, 50257008, 50267003, 50325005, 50410009, 50565005, 50598000, 50663005, 50701000, 50764006, 50805004, 50844007, 50846009, 50869007, 50874004, 50882004, 50936006, 50941003, 50986000, 51010007, 51030006, 51036000, 51038004, 51062009, 51108008, 51118003, 51151007, 51209006, 51290000, 51292008, 51323005, 51338006, 51476001, 51499002, 51522004, 51543003, 51551000, 51677000, 51694003, 51744007, 51847008, 51854002, 51875005, 51877002, 51953009, 51960003, 52031007, 52087004, 52111006, 52159006, 52182008, 52231000, 52232007, 52305004, 52367009, 52368004, 52457000, 52506002, 52545007, 52619009, 52868006, 52890006, 52914002, 52931009, 52979002, 52994003, 53042006, 53067009, 53076002, 53089008, 53104002, 53156005, 53188002, 53190001, 53192009, 53200002, 53202005, 53206008, 53313006, 53337006, 53372009, 53425008, 53476006, 53616007, 53632005, 53633000, 53660007, 53740009, 53748002, 53771000, 53773002, 53827007, 53877005, 53886000, 53928001, 53954009, 53963006, 54051005, 54053008, 54138003, 54157007, 54176009, 54366004, 54389007, 54422002, 54463008, 54553003, 54554009, 54576000, 54597004, 54609002, 54616001, 54694004, 54711002, 54764003, 54798007, 54822007, 54842003, 54883000, 55023005, 55025003, 55056006, 55168004, 55184003, 55193002, 55197001, 55294001, 55355000, 55366007, 55448001, 55479002, 55483002, 55520003, 55525008, 55617001, 55746001, 55809007, 55860008, 55933000, 55955008, 55957000, 55960007, 55961006, 56165008, 56226004, 56385004, 56451001, 56461008, 56498009, 56512009, 56579005, 56623003, 56661000, 56676008, 56704004, 56776001, 56893005, 56905009, 57203004, 57246001, 57265009, 57339008, 57341009, 57412004, 57419008, 57433008, 57540006, 57602001, 57614009, 57650002, 57658009, 57725006, 57748001, 57771004, 57773001, 57871005, 57921006, 57940000, 58008004, 58031004, 58085004, 58103005, 58271001, 58275005, 58282009, 58358002, 58411009, 58565006, 58569000, 58710009, 58711008, 58748004, 58935006, 58994008, 58997001, 59006007, 59047005, 59049008, 59154002, 59221008, 59229005, 59253004, 59285001, 59329007, 59356009, 59423009, 59471009, 59514009, 59515005, 59547000, 59612001, 59658000, 59707005, 59771005, 59818004, 59848001, 59898000, 59913009, 59968008, 59992000, 60002000, 60037002, 60100005, 60127009, 60136008, 60203004, 60400003, 60416001, 60447007, 60454001, 60476005, 60503003, 60528006, 60531007, 60551006, 60627008, 60658009, 60680007, 60698006, 60699003, 60807005, 60871005, 60983006, 60989005, 61108006, 61164006, 61221008, 61250001, 61300005, 61347001, 61400006, 61401005, 61456005, 61564002, 61565001, 61598006, 61627001, 61704003, 61715008, 61738006, 61804006, 61823004, 61860000, 61998001, 62058003, 62151007, 62188005, 62198004, 62216007, 62224002, 62240004, 62341002, 62366003, 62385000, 62394006, 62465000, 62473009, 62477005, 62501005, 62557001, 62643005, 62647006, 62654000, 62809003, 62815003, 62838000, 62851005, 62878007, 62936002, 63040001, 63137003, 63210009, 63246000, 63253009, 63305008, 63425006, 63567004, 63597007, 63637002, 63650001, 63684002, 63691004, 63722008, 63811003, 63866002, 63890001, 63910001, 63924002, 63954007, 64094003, 64121000, 64143008, 64151006, 64153009, 64226004, 64252005, 64283009, 64286001, 64376004, 64398008, 64458007, 64534006, 64581007, 64595005, 64613007, 64638002, 64664008, 64834002, 64969001, 64976006, 64994000, 65001009, 65018008, 65029007, 65071008, 65082000, 65164000, 65191003, 65275009, 65324009, 65390006, 65498003, 65510006, 65544004, 65617004, 65619001, 65626001, 65747008, 65759007, 65785006, 65860006, 65941003, 65976001, 66063001, 66071002, 66123000, 66149005,

66160001, 66186006, 66265004, 66329006, 66367009, 66379009, 66383009, 66522001, 66556007, 66569006, 66636001, 66673003, 66727005, 66767006, 66789005, 66793004, 66865009, 66870002, 66889002, 66929000, 66948001, 66972006, 67012005, 67038005, 67045005, 67153004, 67289000, 67365005, 67375008, 67447005, 67504007, 67508005, 67529001, 67600007, 67602004, 67656006, 67745001, 67766009, 67787004, 67792002, 67795000, 67915005, 67964002, 67968004, 68072000, 68094008, 68126001, 68162000, 68301001, 68368005, 68394001, 68480001, 68496003, 68539005, 68591005, 68601009, 68627009, 68708005, 68734002, 68834009, 68914007, 69087003, 69098002, 69186005, 69248007, 69254008, 69299000, 69478001, 69518005, 69643006, 69686006, 69771008, 69776003, 69800000, 69817007, 69825009, 69850007, 69914001, 69980003, 70020005, 70153002, 70171009, 70176004, 70190001, 70257007, 70305005, 70342003, 70375006, 70418001, 70441009, 70450006, 70529004, 70636008, 70667005, 70668000, 70705008, 70737009, 70880006, 70911004, 70931000, 71009001, 71088006, 71207007, 71215005, 71226004, 71262003, 71363005, 71418005, 71419002, 71449000, 71457002, 71485000, 71583005, 71663002, 71728002, 71745007, 71801004, 71851009, 71858003, 71912000, 71953008, 72007001, 72048003, 72053008, 72061003, 72150009, 72219001, 72245005, 72332008, 72391004, 72395008, 72408002, 72430001, 72438008, 72445008, 72486001, 72519002, 72533002, 72547003, 72621003, 72681001, 72881003, 72888009, 72900001, 72925005, 72950008, 72951007, 73120006, 73125001, 73147001, 73197002, 73225005, 73235004, 73335002, 73423000, 73455000, 73475009, 73479003, 73481001, 73518002, 73651003, 73665001, 73817000, 73861008, 73871005, 73943003, 74066007, 74091007, 74162007, 74341002, 74355005, 74361008, 74372003, 74434009, 74474003, 74598008, 74621002, 74669004, 74744007, 74788000, 74797001, 74829002, 74973004, 75009007, 75037000, 75066002, 75119003, 75128002, 75198005, 75204008, 75236001, 75260002, 75342000, 75375008, 75419006, 75581001, 75589004, 75633002, 75694006, 75700000, 75721000, 75726005, 75860007, 75884004, 75891001, 75937003, 75938008, 75955007, 75965001, 75979009, 75994008, 76078009, 76118001, 76174009, 76181002, 76281005, 76301009, 76338009, 76362004, 76502001, 76641005, 76651006, 76719002, 76730004, 76796008, 76813008, 76816000, 76875008, 76940009, 76953007, 76957008, 77028001, 77097004, 77147000, 77225009, 77360001, 77410006, 77412003, 77414002, 77437008, 77480004, 77528005, 77609009, 77661009, 77713007, 77972001, 77987006, 78054007, 78128001, 78160009, 78171005, 78202006, 78208005, 78305006, 78324009, 78372005, 78373000, 78420004, 78430008, 78463001, 78494001, 78537008, 78560000, 78609007, 78625002, 78712000, 78754002, 78793006, 78809005, 78900008, 78911000, 78948009, 78960005, 78974003, 78996009, 79031007, 79037006, 79099006, 79118000, 79261008, 79369007, 79510004, 79552009, 79607001, 79615003, 79720007, 79740000, 79787007, 79806007, 79812002, 79831008, 79832001, 79977000, 79990007, 80018001, 80034003, 80060002, 80193009, 80281008, 80289005, 80340002, 80353004, 80378000, 80426004, 80446009, 80475003, 80483009, 80515008, 80527006, 80567007, 80640009, 80641008, 80654001, 1197705006, 396231007, 396335001, 396336000, 396337009, 396330006, 396331005, 396460000, 396461001, 397173003, 397575003, 397696004, 397729009, 397843005, 397869004, 397881000, 398061002, 398076005, 398132001, 398157006, 398197009, 398311004, 398314007, 398316009, 398320008, 398384001, 398570005, 398676009, 398870000, 399044006, 399049001, 399050001, 399094007, 399165002, 399172001, 399291005, 399379004, 399391008, 399422005, 399432003, 399468008, 399505005, 399563001, 399691000, 399723004, 399730005, 399924003, 399929008, 399946006, 399967006, 399974001, 400014002, 400036004, 400037008, 401312001, 401313006, 402194008, 402209004, 402235003, 402247009, 402250007, 402251006, 402252004, 402278008, 402279000, 402280002, 402281003, 402282005, 402283000, 402284006, 402285007, 402288009, 402289001, 402294001, 402316001, 402342001, 402362009, 402375009, 402376005, 402377001, 402407004, 402440008, 402441007, 402444004, 402451008, 402455004, 402479002, 402505009, 402506005, 402701002, 402702009, 402703004, 402704005, 402709000, 402757006, 402767001, 402813000, 402819001, 402886003, 402887007, 402892009,

402893004, 402900005, 402906004, 402908003, 402942007, 402944008, 402950003, 402951004, 402963009, 402964003, 402996006, 402997002, 402998007, 403210001, 403367007, 403368002, 403369005, 403383007, 403438007, 403439004, 403440002, 403441003, 403442005, 403443000, 403444006, 403445007, 403446008, 403447004, 403448009, 403449001, 403450001, 403451002, 403452009, 403453004, 403454005, 403455006, 403456007, 403460005, 403461009, 403484002, 403485001, 403495008, 403567006, 403636004, 403655004, 403663003, 403664009, 403665005, 403701008, 403702001, 403703006, 403704000, 403705004, 403725000, 403762003, 403764002, 403889000, 403926005, 403961004, 403963001, 404061009, 404090003, 404094007, 404194009, 404199004, 404811004, 404908004, 405247003, 405248008, 405257002, 405258007, 405542005, 405545007, 405558008, 405559000, 405563007, 405565000, 405567008, 405574003, 405575002, 405578000, 405579008, 405582003, 405585001, 405589007, 405590003, 405737000, 408335007, 408537003, 408539000, 408644002, 408645001, 408646000, 408647009, 408649007, 409506009, 409554001, 409563004, 409656006, 409657002, 409662001, 409705005, 409706006, 409865002, 409966000, 410066003, 410484008, 412787009, 413184002, 413197000, 413208006, 413210008, 413211007, 413212000, 413213005, 413214004, 413216002, 413217006, 413218001, 413219009, 413220003, 413221004, 413224007, 413225008, 413226009, 413227000, 413228005, 413255001, 413256000, 413258004, 413276006, 413438002, 413445002, 413446001, 413551009, 414155001, 414156000, 414342007, 414375004, 414390000, 414391001, 414397002, 414398007, 414401005, 414474001, 414475000, 414476004, 414529006, 414543001, 414544007, 414581006, 414603003, 414663001, 414923000, 414924006, 414952002, 415074004, 415148007, 415251008, 415252001, 415253006, 415254000, 415255004, 415256003, 415589002, 415594002, 415595001, 415623008, 415624002, 415650003, 415652006, 415653001, 415654007, 415655008, 415753007, 415767003, 416389007, 416482004, 416605008, 418103007, 418130002, 418171008, 418372008, 418973007, 419219000, 419240004, 419585004, 419616000, 419645003, 419728003, 419842002, 419958000, 420054005, 420120006, 420121005, 420373003, 420706008, 420910002, 421204004, 421224000, 421249001, 421429008, 421807004, 422400008, 422470007, 422541001, 422691006, 422706003, 422775003, 422833009, 422886007, 422977003, 423038006, 423106003, 423189008, 423318000, 423424005, 423464009, 423471004, 423590009, 423607006, 423615009, 423691004, 423708008, 423793008, 423843003, 424091006, 424263008, 424279009, 424302003, 424327005, 424440001, 424487008, 424567005, 424580008, 424758008, 424802006, 424849005, 425127006, 425178004, 425225007, 425318003, 425413006, 425414000, 425671009, 425698008, 425739008, 426033005, 426137009, 426364002, 426508001, 426549001, 426552009, 426965005, 427022004, 427044009, 427106002, 427151002, 427199002, 427225008, 427380004, 427399008, 427592000, 427743008, 427744002, 427758000, 427791009, 427883005, 427898007, 427910000, 428054006, 428137000, 428216000, 428472008, 428701004, 428722005, 428743006, 428867008, 428905002, 429196001, 429200006, 429274001, 429300008, 429759002, 429974006, 429975007, 430097009, 430328004, 430347001, 430349003, 430876009, 430877000, 430887001, 431037002, 431040002, 431222008, 431505005, 431506006, 431674004, 431873008, 431874002, 432294000, 432414001, 432461000, 432511006, 432772009, 432777003, 432862004, 432908002, 432990004, 432994008, 433184006, 433237003, 433592008, 433600001, 433804007, 433809002, 434202008, 434238001, 434270001, 434271002, 438495005, 438627000, 438946002, 439013004, 439108000, 439442003, 439478008, 439849002, 439955006, 440471007, 440501006, 440544005, 440630006, 440663004, 441285006, 441787004, 441798003, 441935006, 441971007, 442134007, 442159003, 442191002, 442222009, 442231009, 442267002, 442316003, 442374005, 442405009, 442483004, 442484005, 442551007, 442685003, 442692008, 442715002, 442858008, 443488001, 443775000, 443776004, 443777008, 253984004, 253989009, 253995005, 253996006, 253997002, 254000002, 254001003, 254002005, 254093009, 254177003, 254388002, 254389005, 254390001, 254392009, 254393004, 254394005, 254395006, 254397003, 254398008, 254399000, 254401006, 254402004, 254403009, 254404003, 254405002, 254406001, 254407005, 254408000, 254409008, 254410003, 254412006, 254413001, 254414007, 254415008, 254416009, 254417000, 254418005, 254419002, 254423005, 254424004, 254425003, 254426002, 254427006, 254428001, 254429009, 254430004, 254431000, 254432007, 254435009, 254436005, 254437001, 254438006, 254440001, 254441002,

254442009, 254443004, 254444005, 254445006, 254446007, 254447003, 254448008, 254449000, 254450000, 254451001, 254452008, 254453003, 254454009, 254455005, 254456006, 254457002, 254458007, 254459004, 254460009, 254462001, 254463006, 254465004, 254466003, 254489006, 254492005, 254497004, 254498009, 254501009, 254503007, 254504001, 254505000, 254506004, 254532005, 254533000, 254535007, 254539001, 254540004, 254543002, 254544008, 254547001, 254549003, 254551004, 254553001, 254555008, 254557000, 254559002, 254561006, 254563009, 254564003, 254567005, 254570009, 254573006, 254574000, 254582000, 254583005, 254586002, 254588001, 254589009, 254590000, 254591001, 254592008, 254593003, 254594009, 254595005, 254596006, 254597002, 254598007, 254599004, 254600001, 254601002, 254604005, 254609000, 254611009, 254612002, 254613007, 254614001, 254615000, 255038009, 255039001, 255040004, 255042007, 255043002, 255062004, 255064003, 255069008, 255070009, 255071008, 255072001, 255074000, 255077007, 255078002, 255081007, 255084004, 255086002, 255087006, 255088001, 255131000, 255132007, 255136005, 255137001, 255140001, 255143004, 255153003, 255154009, 255155005, 255158007, 255167007, 255168002, 255169005, 255171005, 255172003, 255173008, 255174002, 255175001, 255225007, 255988004, 259058004, 259583007, 261409005, 261827001, 262263007, 262602008, 262603003, 262604009, 262605005, 262606006, 262607002, 262608007, 262609004, 262627001, 262628006, 262629003, 262631007, 262632000, 262633005, 262635003, 262637006, 262638001, 262639009, 262648004, 262650007, 262651006, 262652004, 262653009, 262654003, 262655002, 262656001, 262658000, 262659008, 262660003, 262661004, 262662006, 262663001, 262664007, 262665008, 262668005, 262669002, 262766003, 262767007, 262768002, 262769005, 262770006, 262771005, 262772003, 262790002, 262791003, 262792005, 262793000, 262795007, 262796008, 262797004, 262798009, 262799001, 262801003, 262802005, 262803000, 262804006, 262805007, 262808009, 262809001, 262810006, 262811005, 262814002, 262815001, 262816000, 262826007, 262827003, 262828008, 262829000, 262830005, 262837008, 262838003, 262839006, 262840008, 262841007, 262842000, 262843005, 262845003, 262846002, 262848001, 262849009, 262850009, 262851008, 262852001, 262853006, 262854000, 262855004, 262856003, 262857007, 262858002, 262859005, 262860000, 262861001, 262862008, 262863003, 262864009, 262865005, 262866006, 262867002, 262868007, 262869004, 262870003, 262871004, 262872006, 262873001, 262874007, 262875008, 262876009, 262877000, 262878005, 262879002, 262880004, 262881000, 262882007, 262883002, 262884008, 262885009, 262886005, 262887001, 262888006, 262889003, 262902006, 263157002, 263158007, 263159004, 263161008, 263162001, 263173008, 263174002, 263175001, 263271000, 263272007, 263273002, 263274008, 263275009, 266071000, 266077001, 266078006, 266081001, 266090008, 266108008, 266114001, 266154004, 266157006, 266160004, 266162007, 266343004, 266412007, 266415009, 266421008, 266430000, 266434009, 266435005, 266438007, 266439004, 266451002, 266463007, 266464001, 266468003, 266469006, 266470007, 266471006, 266480006, 266519001, 266520007, 267051003, 268197001, 268201001, 268205005, 268209004, 268213006, 268219005, 268245001, 268651002, 268850004, 268938009, 269162006, 269229006, 269459004, 269460009, 269480005, 269515006, 269516007, 269533000, 269544008, 270513005, 270516002, 270909006, 271123000, 271139007, 271323007, 271324001, 271326004, 271366000, 271432005, 271440004, 271482000, 271525004, 271527007, 271568003, 271575002, 271943005, 271956003, 271959005, 271960000, 271981000, 274080003, 274081004, 274085008, 274106005, 274902006, 274944000, 274950005, 275262008, 275266006, 275296001, 275393007, 275394001, 275395000, 275396004, 275397008, 275399006, 275400004, 275402007, 275431006, 275448003, 275456000, 275486007, 275487003, 275489000, 275490009, 275491008, 275492001, 275508008, 275510005, 275744000, 275943004, 275959004, 276226001, 276229008, 276392006, 276393001, 276394007, 276464002, 276465001, 276500007, 276520008, 276521007, 276522000, 276523005, 276524004, 276525003, 276527006, 276539003, 276548008, 276551001, 276552008, 276553003, 276554009, 276563006, 276723008, 276803003, 276804009, 276805005, 276808007, 276809004, 276810009, 276811008, 276812001, 276815004, 276816003, 276818002, 276819005, 276821000, 276822007, 276851006, 276852004, 276883000, 276952000, 276953005, 276954004, 276962007, 277540008, 277563003, 277678003, 277782009, 277783004, 277807007, 277810000, 278486006, 278488007, 278521000, 278527001, 278535003,

278682004, 278714002, 278895007, 278933001, 279309008, 280356005, 280483007, 280992001, 280993006, 281095009, 281254000, 281255004, 281388009, 282007005, 282041002, 282042009, 282050000, 282051001, 282052008, 282053003, 282054009, 282055005, 282058007, 282059004, 282060009, 282061008, 282062001, 282063006, 282064000, 282065004, 282066003, 282068002, 282069005, 282070006, 282072003, 282073008, 282074002, 282075001, 282076000, 282077009, 282080005, 282082002, 282084001, 282085000, 282348002, 282429000, 282430005, 282431009, 282432002, 282434001, 282438003, 282439006, 282452001, 282453006, 282454000, 282455004, 282664001, 282754004, 282783001, 283015002, 283073002, 283121001, 283146001, 283147005, 283236008, 283237004, 283328001, 283355005, 283363006, 283406002, 283455006, 283456007, 283507009, 283555009, 283556005, 283606008, 283694003, 283744007, 283799003, 283855001, 283876006, 283877002, 283979000, 283980002, 284001007, 284002000, 284204005, 284205006, 284535003, 284574005, 284575006, 284583000, 284584006, 285307007, 285308002, 285310000, 285312008, 285338008, 285344007, 285361005, 285609003, 285610008, 285611007, 285612000, 285613005, 285614004, 285616002, 285671001, 285800001, 285801002, 285806007, 285807003, 285808008, 285834000, 286547006, 286567002, 286602003, 286622004, 286623009, 286887005, 286902000, 286967008, 286970007, 286972004, 287101003, 287326003, 288199005, 289260002, 289923007, 289999003, 290187002, 295046003, 297140006, 297158000, 297164007, 297165008, 297177003, 297189001, 297190005, 297191009, 297251003, 297252005, 297253000, 297254006, 297255007, 297288000, 298016002, 298017006, 298065007, 298066008, 298135000, 298136004, 299683005, 299684004, 299709002, 299969005, 300078006, 300258005, 300259002, 300922007, 300931007, 300932000, 300935003, 300952004, 301007008, 301008003, 301009006, 301469002, 301756000, 301779004, 301797007, 301799005, 301829006, 301836007, 302168000, 302169008, 302233006, 302323001, 302815008, 302819002, 302823005, 302824004, 302865004, 302910002, 302911003, 302916008, 302919001, 302920007, 302921006, 302948007, 302949004, 302951000, 302952007, 302953002, 1172705006, 1003388008, 1003390009, 1003397007, 1003398002, 1003400003, 1003421003, 1003433008, 1003439007, 1003468008, 1003514000, 1003556001, 1003567000, 1003569002, 1010603001, 1010607000, 1010616001, 1010624006, 1010626008, 1010633008, 1010656004, 1010669000, 1010674008, 1010675009, 1010677001, 1010679003, 1010681001, 10233431100011910 1, 1052210007, 1052237009, 1052238004, 1052296002, 1052300001, 1052321001, 1078279007, 107831000119101, 1087291000119106, 1119392002, 1119393007, 15959741000119102, 15959781000119107, 15965661000119103, 15965701000119105, 15965741000119107, 15965781000119102, 15966101000119102, 15966221000119102, 15966261000119107, 15969021000119101, 15969701000119108, 15970021000119103, 15970261000119101, 15970301000119109, 15971661000119105, 15971861000119106, 15971901000119100, 15972061000119109, 16696981000119105, 16697061000119100, 16697181000119105, 16840431000119101, 453720571000119100, 874837003, 874838008, 874914008, 874915009, 874919003, 876794002, 876807009, 876821006, 878799003, 878807001, 878818001, 878822006, 879873008, 879991002, 879992009, 880001008, 890099000, 890101007, 890102000, 890103005, 890104004, 890105003, 890118006, 890201000, 890217006, 890229002, 890365005, 890369004, 890379002, 890382007, 890383002, 890385009, 890408008, 894093007, 895086000, 895214001, 895215000, 895517007, 897004000, 897005004, 897304009, 897541006, 897589002, 897594002, 897595001, 966011731000119103, 969688801000119108, 1208933000, 1197183009, 1196904007, 610412651000119100, 1196832002, 1196828008, 1196829000, 1196827003, 1222709003, 1196819007, 1196821002, 1204371007, 1197676001, 1197678000, 1197341008, 1197338004, 1197339007, 1197675002, 1197594000, 1197344000, 1197336000, 1197337009, 1201780008, 1196875008, 1209075008, 1197672004, 1217704000, 1197673009, 1208338004, 1197713007, 1204348005, 1197712002, 1197736003, 1197738002, 1204221007, 344015141000119100, 289566561000119108, 1197711009, 1197735004, 1196836004, 898816651000119105, 1197729004, 1197666004, 1197270001, 1197660005, 1197699002, 1197710005, 1197720000, 1196880004, 1196878005, 1197204009, 1222706005, 1204179003, 1204180000, 1197369000, 1163487005, 1179027005, 1197703004, 1228875006, 1208875000, 1217380005, 1230135006

Truveta: 1215942, 1215944

#### Asthma

ICD-9-CM: 493.01, 493.02, 493.11, 493.12, 493.21, 493.22, 493.91, 493.92

ICD-10-CM: J45.21, J45.22, J45.31, J45.32, J45.4, J45.40, J45.41, J45.42, J45.5, J45.50, J45.51, J45.52, J45.901, J45.902, O99.5, O99.51, O99.511, O99.512, O99.513, O99.519, O99.52, O99.53

SNOMED CT: 1751000119100, 99031000119107, 124991000119109, 125001000119103, 10674711000119105, 10675431000119106, 10675471000119109, 10675551000119104, 10675911000119109, 10675991000119100, 10676391000119108, 10676431000119103, 10676511000119109, 10692721000119102, 707446004, 707447008, 707512002, 707513007, 707979007, 707980005, 707981009, 708090002, 708093000, 708094006, 708095007, 708096008, 370219009, 370221004, 426656000, 427295004, 442025000, 786836003, 829976001, 135171000119106, 10675391000119101, 10675751000119107, 733858005, 734904007, 782513000

#### Substance Use Disorder

ICD-10-CM: F10, F11, F12, F13, F14, F15, F16, F17, F18, F19, O99.31, O99.32, F10.1, F10.10, F10.11, F10.12, F10.120, F10.121, F10.129, F10.14, F10.15, F10.150, F10.151, F10.159, F10.18, F10.180, F10.181, F10.182, F10.188, F10.19, F10.2, F10.20, F10.21, F10.22, F10.220, F10.221, F10.229, F10.23, F10.230, F10.231, F10.232, F10.239, F10.24, F10.25, F10.250, F10.251, F10.259, F10.26, F10.27, F10.28, F10.280, F10.281, F10.282, F10.288, F10.29, F10.9, F10.92, F10.920, F10.921, F10.929, F10.94, F10.95, F10.950, F10.951, F10.959, F10.96, F10.97, F10.98, F10.980, F10.981, F10.982, F10.988, F10.99, F11.1, F11.10, F11.11, F11.12, F11.120, F11.121, F11.122, F11.129, F11.14, F11.15, F11.150, F11.151, F11.159, F11.18, F11.181, F11.182, F11.188, F11.19, F11.2, F11.20, F11.21, F11.22, F11.220, F11.221, F11.222, F11.229, F11.23, F11.24, F11.25, F11.250, F11.251, F11.259, F11.28, F11.281, F11.282, F11.288, F11.29, F11.9, F11.90, F11.92, F11.920, F11.921, F11.922, F11.929, F11.93, F11.94, F11.95, F11.950, F11.951, F11.959, F11.98, F11.981, F11.982, F11.988, F11.99, F12.1, F12.10, F12.11, F12.12, F12.120, F12.121, F12.122, F12.129, F12.15, F12.150, F12.151, F12.159, F12.18, F12.180, F12.188, F12.19, F12.2, F12.20, F12.21, F12.22, F12.220, F12.221, F12.222, F12.229, F12.23, F12.25, F12.250, F12.251, F12.259, F12.28, F12.280, F12.288, F12.29, F12.9, F12.90, F12.92, F12.920, F12.921, F12.922, F12.929, F12.93, F12.95, F12.950, F12.951, F12.959, F12.98, F12.980, F12.988, F12.99, F13.1, F13.10, F13.11, F13.12, F13.120, F13.121, F13.129, F13.14, F13.15, F13.150, F13.151, F13.159, F13.18, F13.180, F13.181, F13.182, F13.188, F13.19, F13.2, F13.20, F13.21, F13.22, F13.220, F13.221, F13.229, F13.23, F13.230, F13.231, F13.232, F13.239, F13.24, F13.25, F13.250, F13.251, F13.259, F13.26, F13.27, F13.28, F13.280, F13.281, F13.282, F14.129, F13.288, F13.29, F13.9, F13.90, F13.92, F13.920, F13.921, F13.929, F13.93, F13.930, F13.931, F13.932, F13.939, F13.94, F13.95, F13.950, F14.14, F13.951, F13.959, F13.96, F13.97, F13.98, F13.980, F13.981, F13.982, F13.988, F13.99, F14.1, F14.10, F14.11, F14.12, F14.120, F14.121, F14.122, F14.15, F14.150, F14.151, F14.159, F14.18, F14.180, F14.181, F14.182, F14.188, F14.19, F14.2, F14.20, F14.21, F14.22, F14.220, F14.221, F14.222, F14.229, F14.23, F14.24, F14.25, F14.250, F14.251, F14.259, F14.28, F14.280, F14.281, F14.282, F14.288, F14.29, F14.9, F14.90, F14.92, F14.920, F14.921, F14.922, F14.929, F14.94, F14.95, F14.950, F14.951, F14.959, F14.98, F14.980, F14.981, F14.982, F14.988, F14.99, F15.1, F15.10, F15.11, F15.12, F15.120, F15.121, F15.122, F15.129, F15.14, F15.15, F15.150,

F15.151, F15.159, F15.18, F15.180, F15.181, F15.182, F15.188, F15.19, F15.2, F15.20, F15.21, F15.22, F15.220, F15.221, F15.222, F15.229, F15.23, F15.24, F15.25, F15.250, F15.251, F15.259, F15.28, F15.280, F15.281, F15.282, F15.288, F15.29, F15.9, F15.90, F15.92, F15.920, F15.921, F15.922, F15.929, F15.93, F15.94, F15.95, F15.950, F15.951, F15.959, F15.98, F15.980, F15.981, F15.982, F15.988, F15.99, F16.1, F16.10, F16.11, F16.12, F16.120, F16.121, F16.122, F16.129, F16.14, F16.15, F16.150, F16.151, F16.159, F16.18, F16.180, F16.183, F16.188, F16.19, F16.2, F16.20, F16.21, F16.22, F16.220, F16.221, F16.229, F16.24, F16.25, F16.250, F16.251, F16.259, F16.28, F16.280, F16.283, F16.288, F16.29, F16.9, F16.90, F16.92, F16.920, F16.921, F16.929, F16.94, F16.95, F16.950, F16.951, F16.959, F16.98, F16.980, F16.983, F16.988, F16.99, F17.2, F17.20, F17.200, F17.201, F17.203, F17.208, F17.209, F17.21, F17.210, F17.211, F17.213, F17.218, F17.219, F17.22, F17.220, F17.221, F17.223, F17.228, F17.229, F17.29, F17.290, F17.291, F17.293, F17.298, F17.299, F18.1, F18.10, F18.11, F18.12, F18.120, F18.121, F18.129, F18.14, F18.15, F18.150, F18.151, F18.159, F18.17, F18.18, F18.180, F18.188, F18.19, F18.2, F18.20, F18.21, F18.22, F18.220, F18.221, F18.229, F18.24, F18.25, F18.250, F18.251, F18.259, F18.27, F18.28, F18.280, F18.288, F18.29, F18.9, F18.90, F18.92, F18.920, F18.921, F18.929, F18.94, F18.95, F18.950, F18.951, F18.959, F18.97, F18.98, F18.980, F18.988, F18.99, F19.1, F19.10, F19.11, F19.12, F19.120, F19.121, F19.122, F19.129, F19.14, F19.15, F19.150, F19.151, F19.159, F19.16, F19.17, F19.18, F19.180, F19.181, F19.182, F19.188, F19.19, F19.2, F19.20, F19.21, F19.22, F19.220, F19.221, F19.222, F19.229, F19.23, F19.230, F19.231, F19.232, F19.239, F19.24, F19.25, F19.250, F19.251, F19.259, F19.26, F19.27, F19.28, F19.280, F19.281, F19.282, F19.288, F19.29, F19.9, F19.90, F19.92, F19.920, F19.921, F19.922, F19.929, F19.93, F19.930, F19.931, F19.932, F19.939, F19.94, F19.95, F19.950, F19.951, F19.959, F19.96, F19.97, F19.98, F19.980, F19.981, F19.982, F19.988, F19.99, O99.310, O99.311, O99.312, O99.313, O99.314, O99.315, O99.320, O99.321, O99.322, O99.323, O99.324, O99.325, F10.13, F10.130, F10.131, F10.132, F10.139, F10.93, F10.930, F10.931, F10.932, F10.939, F11.13, F12.13, F13.13, F13.130, F13.131, F13.132, F13.139, F14.13, F14.93, F15.13, F19.13, F19.130, F19.131, F19.132, F19.139

ICD-9-CM: 291, 303, 304, 305, 648.3, 649.0

SNOMED CT: 1383008, 1686006, 1973000, 2403008, 281004, 5002000, 5444000, 5602001, 6348008, 7052005, 7071007, 7200002, 8635005, 8837000, 10028000, 10327003, 11061003, 11387009, 14784000, 15167005, 15277004, 18653004, 18689007, 19445006, 20385005, 20876004, 21647008, 22574000, 25753007, 26714005, 27956007, 28864000, 29212009, 29733004, 30491001, 31956009, 32009006, 32358001, 32875003, 80868005, 82339009, 83367009, 84758004, 85005007, 85561006, 86325007, 87132004, 87810006, 87858002, 88926005, 89451009, 91388009, 95661003, 817962007, 838527002, 1081000119105, 23601000119102, 34111000119108, 86391000119101, 86401000119104, 97571000119109, 125851000119106, 135301000119103, 135311000119100, 135321000119107, 144981000119109, 145101000119102, 145121000119106, 145841000119107, 154211000119108, 288021000119107, 288031000119105, 288461000119105, 288851000119106, 288861000119108, 10741871000119101, 10755041000119100, 11047881000119101, 11048011000119103, 12398281000119105, 12398651000119100, 699449003, 703845008, 703846009, 703847000, 703848005, 703849002, 703850002, 713583005, 713862009, 714829008, 719848005, 723926008, 723927004, 723928009, 723929001, 723930006, 723933008, 723936000, 723937009, 724653003, 724655005, 724656006, 724657002, 724658007, 724659004, 724660009, 724661008, 724662001, 724663006, 724664000, 724665004, 724666003, 724667007, 724668002, 724669005, 724670006, 724671005, 724672003, 724675001, 724688003, 724689006, 724690002, 724691003, 724692005, 724693000, 724695007, 724696008, 724700000, 724701001, 724702008, 724704009, 724706006, 724712001, 724714000, 724715004, 724720004, 724724008,

724725009, 724726005, 724727001, 724728006, 724732000, 735750005, 737338002, 762320004, 762321000, 762322007, 762324008, 762325009, 762326005, 762327001, 762333005, 762334004, 762335003, 762336002, 762337006, 762338001, 762339009, 762340006, 762344002, 762345001, 762346000, 762517008, 762672001, 785277001, 191475009, 191476005, 191478006, 191480000, 191802004, 191804003, 191805002, 191811004, 191812006, 191819002, 191820008, 191825003, 191826002, 191831000, 191832007, 191837001, 191838006, 191843004, 191844005, 191849000, 191850000, 191853003, 191855005, 191856006, 191865004, 191867007, 191868002, 191882002, 191883007, 191891003, 191893000, 191894006, 191899001, 191900006, 191905001, 191906000, 191909007, 191912005, 191913000, 191916008, 191918009, 191919001, 191924003, 191925002, 308742005, 312098001, 312936002, 361151007, 230443000, 231461004, 231462006, 231468005, 231469002, 231470001, 231472009, 231473004, 231474005, 231477003, 231478008, 231479000, 231480002, 34938008, 37344009, 38247002, 39807006, 39951001, 40571009, 41083005, 42344001, 43497001, 46975003, 47664006, 50320000, 50933003, 51443000, 51493001, 52866005, 53050002, 53936005, 56294008, 57588009, 59651006, 61104008, 61144001, 63649001, 63983005, 64386003, 66590003, 70328006, 70340006, 70701004, 70932007, 71328000, 73097000, 74851005, 74934004, 75122001, 75544000, 77355000, 78267003, 78358001, 416714005, 417143004, 425339005, 425885002, 426001001, 426095000, 426873000, 427205009, 427229002, 427327003, 428370001, 429692000, 441527004, 442406005, 268640002, 268645007, 268646008, 268647004, 268648009, 275471001, 284591009, 301643003, 1162823005, 1179217006, 1162820008, 1187220002, 1187219008, 1197724009, 1231319004, 1230083007, 1230087008, 1230085000, 1230080005, 1230078004, 1230076000, 1230075001, 1230081009, 1230079007, 1230077009, 1230074002, 1230089006, 1230082002, 1230084001, 1230086004, 1231336007, 1231333004, 1231331002, 1230088003, 1231162009, 1231160001, 1179385009, 1186867000, 1162818005, 1162805002, 1162824004, 1162821007, 1231158003, 1187205008, 1179728006, 1179727001, 1163367008, 1162817000, 1172690003, 1162803009, 1179725009, 1179724008, 1162505005, 1163009003, 1163013005, 1179730008, 1179732000, 1187103009, 1163008006, 1179731007, 15919911000119108, 15919991000119104, 1186983001, 1162819002, 1162806001, 1162825003, 1162822000, 520841000124109, 1163356007, 1323006, 1531000, 1647007, 1654001, 1657008, 1837006, 1848004, 1977004, 2121004, 2296009, 2638001, 2819007, 3053001, 184004, 276008, 3902000, 4223005, 4390004, 4418008, 4526003, 4688008, 4690009, 4771009, 4836003, 5189009, 5319009, 5368009, 5578008, 6024006, 6900000, 7072000, 7184000, 7201003, 7248001, 7339009, 7511009, 7538002, 7743006, 7916009, 8325007, 8481000, 8732001, 8995005, 9233004, 9291000, 9489006, 9953008, 9982009, 10385009, 10486009, 10650000, 10916006, 10984003, 11268001, 11306002, 11659006, 11862006, 11949008, 11974006, 12442008, 12777009, 12815001, 12876009, 12898000, 13054007, 13151001, 13187008, 13216001, 13503000, 13504006, 13632006, 13857004, 13993001, 14002008, 14130006, 14154001, 14607001, 14651005, 14700006, 15026003, 15244003, 15276008, 15382009, 15649002, 16248006, 16260007, 16332007, 16773005, 16800004, 16959008, 17570002, 17592003, 18051001, 18052008, 18108002, 18311006, 18323000, 18337009, 18597007, 18631001, 18768006, 18869009, 19303008, 19307009, 19920003, 20117002, 20179009, 20202008, 20215004, 20260003, 20310007, 20871009, 20881008, 21413004, 21445009, 21511000, 21673003, 21782001, 21836003, 22583005, 22706002, 22719003, 22888007, 22915003, 23142005, 23146008, 23527004, 23842007, 23877007, 24000001, 24669002, 24984009, 25277000, 25508008, 25555003, 25599007, 25702006, 25980005, 26532007, 26543004, 26617005, 26679003, 26715006, 26768000, 27064003, 27331008, 27992006, 28213007, 28224009, 28348003, 28389000, 28504009, 28563001, 28711005, 28800007, 29290000, 29602005, 30138004, 30310000, 30365001, 30857006, 30871003, 30887000, 30897009, 30984008, 31430005, 31438003, 31715000, 31752000, 32046005, 32093003, 32267003, 32287002, 32371003, 32393008, 32518004, 32552001, 32722006, 32835006, 33217001, 33845003, 33871004, 34085004, 81270000, 81459009, 81648008, 81711008, 81914009, 82047000, 82276009, 82674000, 82768008, 83150005, 83168008, 83465002, 83627000, 83864008, 84393008, 84483000, 84584008,

84811001, 85302003, 85618003, 85761009, 85871000, 85975005, 86225009, 86321003, 86624000, 86733006, 87363009, 87396006, 87816000, 88165009, 88320008, 88674007, 88993007, 89137004, 89183005, 89322006, 89535003, 89715001, 89729000, 89739006, 89765005, 90210000, 90687000, 90755006, 91014009, 91074004, 91267002, 91433009, 93432008, 93433003, 93434009, 93435005, 95239003, 95247003, 95635002, 95909001, 95910006, 95911005, 95912003, 95915001, 95916000, 95917009, 95918004, 95919007, 102448004, 102449007, 103079001, 103297000, 109254000, 109483004, 109774008, 109776005, 109804004, 109813002, 109861009, 109957002, 110281001, 110282008, 110283003, 110284009, 110285005, 110990003, 111345006, 111350000, 111460005, 111480006, 111558006, 111588002, 111755007, 111756008, 111758009, 111760006, 111762003, 111763008, 111764002, 111765001, 111766000, 111768004, 111769007, 111772000, 111779009, 111782004, 118951003, 127060006, 129576003, 129662007, 129663002, 129664008, 129666005, 1142050005, 1142059006, 1142110005, 1144368001, 1144771006, 1144776001, 1144777005, 1144778000, 1144816001, 1144817005, 1144818000, 1144820002, 1144822005, 1144823000, 1144824006, 1144826008, 1144827004, 1144828009, 1144830006, 1144831005, 1144832003, 1144834002, 1144835001, 1144836000, 1144838004, 1144839007, 1145044008, 1145046005, 1145047001, 1145050003, 1145056009, 1145057000, 1145058005, 1145065002, 1145066001, 1145067005, 1145073006, 1145075004, 1145076003, 1145077007, 1145152008, 1148522008, 1148519006, 1145546002, 1145547006, 1145548001, 1148516004, 1148533006, 1148534000, 1148536003, 1148539005, 1148542004, 1148543009, 1148544003, 1148567001, 1148569003, 1148570002, 1148571003, 1148651004, 1148653001, 1148654007, 1148655008, 1148671008, 1148673006, 1148674000, 1148696006, 1148724006, 1148725007, 1148730006, 1148731005, 1148734002, 1148751009, 1148752002, 1148753007, 1148849001, 1148859000, 1148880006, 1149139006, 1149140008, 1149145003, 1149146002, 1149147006, 1149148001, 1149228000, 1149229008, 1149235008, 1149236009, 1149239002, 1149240000, 1149241001, 1149242008, 1149328002, 1149333003, 1149353004, 1149405004, 1149406003, 1149407007, 1149408002, 1149410000, 1149411001, 1149412008, 1149470008, 1149468004, 1149469007, 1153338005, 1153364004, 1153374001, 1153419002, 1153508003, 1153538009, 1153539001, 1153540004, 1153541000, 1153542007, 1155736008, 1155737004, 1155738009, 1155739001, 1155740004, 1155741000, 1155772001, 1155990006, 1155997009, 1155999007, 1156000009, 1156001008, 1156002001, 1156013003, 1156515000, 1156517008, 1156518003, 1156734002, 1156735001, 1156746003, 1156753007, 1156769000, 1157240004, 1157257000, 136281000119109, 328371000119107, 328381000119105, 470681000124105, 470701000124108, 471751000124101, 471761000124104, 471771000124106, 471791000124107, 1231327008, 1231325000, 788863007, 788904003, 788905002, 788955005, 788956006, 788957002, 788958007, 788983007, 789046004, 789119007, 789255009, 789271009, 789272002, 789273007, 789560009, 827074008, 830280003, 830281004, 830282006, 833317005, 836439001, 838325009, 838336002, 838376007, 838382005, 838386008, 838519006, 838520000, 838521001, 838522008, 838523003, 838524009, 838525005, 838526006, 838528007, 838529004, 838530009, 838551007, 840300001, 840449007, 840589003, 840591006, 840592004, 846605006, 846646001, 846758008, 860699005, 860826006, 860915001, 863953007, 866258001, 866260004, 870367008, 870409005, 870410000, 870411001, 870412008, 870413003, 870414009, 870515008, 870621003, 870674008, 870677001, 870691007, 870715006, 870749003, 870750003, 871513009, 871521003, 871522005, 871525007, 871526008, 871529001, 871551009, 871576005, 871565006, 871569000, 871570004, 871572007, 871573002, 871574008, 871577001, 871629009, 871673008, 871674002, 871675001, 871676000, 871678004, 871679007, 871681009, 871683007, 871684001, 871685000, 871774005, 871776007, 871922002, 1461000119109, 5851000119107, 7761000119106, 21831000119109, 21841000119100, 22621000119103, 24151000119106, 27051000119102, 91411000119105, 91461000119108, 91471000119102, 91561000119109, 98811000119103, 104521000119108, 104831000119109, 118791000119106, 129141000119104, 133301000119102, 133361000119101, 135331000119105, 153491000119103, 153501000119105, 298531000119104, 298541000119108, 298551000119105, 298561000119107, 298581000119103, 298601000119107, 298611000119105, 298621000119103, 306131000119100, 306141000119109, 306151000119106, 306161000119108, 306171000119102, 306181000119104,

306191000119101, 306201000119103, 306211000119100, 306221000119107, 306371000119100, 306381000119102, 306531000119108, 306541000119104, 306551000119102, 306561000119100, 306571000119106, 306581000119109, 306591000119107, 306601000119100, 306611000119102, 306621000119109, 306631000119107, 306641000119103, 306651000119101, 306731000119104, 306811000119101, 306821000119108, 306831000119106, 315791000119107, 315801000119108, 315951000119104, 316061000119106, 328301000119102, 331981000119104, 337601000119109, 367201000119101, 367391000119102, 367421000119109, 367631000119105, 367761000119105, 368281000119108, 369091000119106, 429061000124102, 429211000124107, 432391000124104, 433231000124100, 434761000124100, 447961000124105, 453421000124103, 453431000124100, 453441000124105, 453451000124107, 455411000124107, 455421000124104, 455431000124101, 455441000124106, 455451000124108, 455461000124105, 456311000124109, 461061000124108, 461071000124101, 678401000119102, 678411000119104, 678421000119106, 680531000119103, 1078301000112109, 1089991000119109, 10625831000119107, 10625951000119109, 10692681000119108, 10755161000119102, 10838971000119103, 10853021000119107, 10854051000119101, 12398201000119102, 12398321000119100, 12398361000119105, 12398571000119105, 15735361000119109, 15735401000119100, 15911041000119105, 15911111000119105, 15919781000119104, 15920041000119105, 15921411000119106, 15921731000119106, 15993671000119108, 15993711000119107, 15993751000119108, 16031471000119107, 16076691000119100, 16077091000119107, 16077171000119107, 16228931000119102, 16236661000119100, 16238181000119101, 16238221000119109, 16238741000119105, 16270221000119108, 16623961000119100, 666000, 4863002, 7665002, 7690000, 7895008, 9408000, 10070005, 10553007, 11196001, 13889008, 15233006, 17231009, 21007002, 22681004, 23833000, 27795004, 28368009, 28926001, 30564001, 35279005, 37121000, 38751004, 41234002, 43124005, 44047000, 44969005, 45472002, 46193001, 47237001, 47362003, 48606007, 51725000, 52335006, 54531000, 56043005, 57125001, 70539005, 71320007, 72566003, 72820004, 78731006, 81119003, 85337000, 86062001, 87904007, 88031005, 105624005, 105625006, 111767009, 129660004, 129665009, 191471000, 196607008, 212718008, 216518007, 235952002, 236522005, 237694000, 237738005, 288293001, 288295008, 290424002, 290425001, 290427009, 290591004, 290592006, 290593001, 292061006, 292134008, 292168002, 292366005, 292739008, 292800009, 292856008, 292992006, 293258008, 293386002, 293544007, 295324004, 295325003, 295327006, 295427007, 295428002, 295429005, 295430000, 402748000, 403622009, 403633007, 403638003, 403644004, 403674007, 421527008, 422593004, 448090008, 448091007, 448178009, 699022007, 699687003, 435491000124103, 444271000, 445507008, 445902007, 446337009, 446468007, 448177004, 449845006, 473188002, 609437000, 609438005, 698088001, 698302009, 698321001, 698402008, 698403003, 698480002, 698481003, 698492002, 698493007, 698577000, 698609007, 698622004, 698707001, 698838008, 699011008, 699012001, 699018002, 699020004, 699021000, 699025009, 699036002, 699064007, 699065008, 699066009, 699067000, 699068005, 699071002, 699072009, 700425005, 702693007, 702702004, 702809001, 702810006, 702811005, 704055002, 704203009, 704205002, 704206001, 707524009, 708079007, 708122002, 708653004, 708655006, 709062008, 711538001, 711539009, 711545001, 712742003, 712950002, 713181003, 713186008, 713244007, 713370005, 713450000, 713496008, 713524002, 713528004, 713529007, 713542007, 713638002, 713691005, 713692003, 713693008, 713694002, 713698004, 713726003, 713728002, 713775002, 713910008, 713965007, 715058004, 715059007, 715267006, 715268001, 715269009, 715270005, 715300008, 715301007, 715302000, 715303005, 715430001, 715431002, 715971003, 716005004, 717127002, 717889000, 717947009, 720580006, 720665007, 720666008, 720672008, 720673003, 720674009, 720675005, 720676006, 720677002, 720678007, 720679004, 720680001, 720681002, 720682009, 720683004, 721710005, 721898008, 722077007, 722120008, 722139003, 722200003, 722321001, 722329004, 722536004, 723103002, 723406000, 723653009, 723882007, 723975009, 724144006, 724519008, 724523000, 724526008, 724529001, 724530006, 724558003, 724574008, 724654009, 724673008, 724674002, 724676000, 724677009, 724678004, 724679007, 724680005, 724681009, 724682002, 724683007, 724684001, 724685000, 724686004, 724687008, 724694006, 724697004, 724698009, 724699001, 724705005, 724707002, 724708007, 724710009, 724711008, 724713006, 724716003, 724717007, 724718002,

724719005, 724721000, 724722007, 724723002, 724729003, 724730008, 724833000, 724843002, 725060001, 725121001, 725287006, 725938001, 725939009, 725940006, 726543008, 735235000, 735456000, 735458004, 735493006, 735616007, 736024007, 736825009, 737226008, 737334000, 737335004, 737336003, 737337007, 737339005, 737340007, 737341006, 737342004, 737557007, 762302008, 762315004, 762323002, 762328006, 762329003, 762330008, 762331007, 762332000, 762384006, 762387004, 762388009, 762390005, 762391009, 762419009, 762420003, 762462005, 762503004, 762504005, 762505006, 762506007, 762507003, 762508008, 762509000, 762510005, 762511009, 762512002, 762513007, 762514001, 762515000, 762516004, 762518003, 762531008, 762562007, 762576008, 762578009, 762671008, 765482002, 766045006, 766054009, 766219005, 766220004, 766221000, 767013007, 770702001, 770704000, 770705004, 770707007, 770708002, 770709005, 770710000, 770711001, 770890009, 771073006, 772020009, 772133000, 772808000, 772999000, 773000004, 782579006, 782593004, 158058005, 169465000, 190282003, 190283008, 190284002, 190416008, 190447002, 190448007, 190524001, 190815001, 190965006, 191148006, 191322006, 191360002, 191390009, 191483003, 191484009, 191485005, 191486006, 191492000, 191493005, 191494004, 191495003, 191496002, 191806001, 191813001, 191816009, 191821007, 191827006, 191833002, 191839003, 191845006, 191851001, 191857002, 191869005, 191871005, 191873008, 191874002, 191875001, 191884001, 191887008, 191888003, 191889006, 191895007, 191901005, 191907009, 191914006, 191920007, 191928000, 191930003, 191931004, 191932006, 191934007, 191936009, 191937000, 191938005, 192811002, 193548006, 193609000, 194791005, 196051003, 196052005, 196053000, 196318002, 196757008, 199252002, 199253007, 199254001, 199255000, 199547006, 199549009, 199550009, 200068009, 200069001, 200070000, 200075005, 200794009, 200844002, 200852004, 200892002, 200893007, 200907003, 201146008, 201436003, 203447001, 203483005, 205788004, 205791004, 206018005, 206021007, 206022000, 206023005, 206124001, 206125000, 206126004, 206127008, 206128003, 206129006, 206130001, 206570000, 304605000, 307757001, 308374001, 308692003, 309279000, 309742004, 309783001, 310202009, 311173003, 312958000, 312959008, 312960003, 312961004, 312962006, 313260003, 313915006, 314422000, 314539001, 361128007, 361129004, 361148000, 361149008, 361268000, 361272001, 361273006, 363101005, 370511006, 371089000, 371341003, 371627004, 390833005, 395215005, 212503009, 212505002, 212509008, 212516009, 212518005, 212523005, 212533002, 212545006, 212553003, 212562001, 212563006, 212596008, 212599001, 212600003, 212602006, 212603001, 212607000, 212615002, 212616001, 212617005, 212619008, 212633008, 212636000, 212641008, 212643006, 212644000, 212658003, 212664005, 212674008, 212691007, 212694004, 212695003, 212700005, 212701009, 212704001, 212706004, 212708003, 212709006, 212712009, 212713004, 212715006, 212723008, 212725001, 212727009, 212728004, 212736008, 212737004, 212738009, 212739001, 212742007, 212748006, 212757000, 212758005, 212761006, 212763009, 212771008, 212773006, 212778002, 212785003, 212807002, 212898003, 216479006, 216480009, 216481008, 216483006, 216507007, 216475000, 216463005, 216464004, 216466002, 216467006, 216468001, 216469009, 216471009, 216472002, 216485004, 216486003, 216487007, 216488002, 216491002, 216494005, 216497003, 216498008, 216499000, 216500009, 216501008, 216502001, 216503006, 216506003, 216514009, 216519004, 216520005, 216521009, 216522002, 216523007, 216525000, 216526004, 216527008, 216528003, 216530001, 216531002, 216532009, 216533004, 216534005, 216535006, 216536007, 216539000, 216541004, 216545008, 216546009, 216547000, 216548005, 216550002, 216551003, 216554006, 216555007, 216556008, 216558009, 216559001, 216560006, 216562003, 216564002, 216569007, 216570008, 216572000, 216573005, 216576002, 216577006, 216579009, 216580007, 216582004, 216583009, 216584003, 216585002, 216586001, 216588000, 216589008, 216590004, 216591000, 216593002, 216594008, 216595009, 216596005, 216599003, 216600000, 216601001, 216603003, 216604009, 216605005, 216609004, 216610009, 216612001, 216614000, 216615004, 216616003, 216618002, 216619005, 216620004, 216621000, 216622007, 216623002, 216624008, 216627001, 216633005, 216661004, 216703009, 216709008, 216733002, 216736005, 218326006, 218356002, 218369003, 218370002, 218379001, 218382006, 218394002, 218395001, 218406006, 218408007, 218416003, 218471002, 218475006, 218481003, 218488009, 218533007, 218534001, 218566006, 218568007, 218572006, 218581000,

218586005, 218606000, 218608004, 218610002, 218613000, 218614006, 218615007, 218619001, 218620007, 218621006, 218626001, 218630003, 218634007, 218635008, 218636009, 218641001, 218643003, 218663005, 218665003, 218680009, 218696007, 218697003, 218698008, 218704008, 218705009, 218718001, 218724007, 218726009, 218727000, 218764006, 218790009, 218791008, 218792001, 218794000, 218795004, 218796003, 218799005, 218808002, 218813003, 218814009, 218845008, 218868004, 218869007, 218903005, 218916005, 218924000, 218925004, 218927007, 218928002, 218929005, 218933003, 218941003, 218943000, 218950001, 218951002, 218952009, 218953004, 218961009, 218968003, 218969006, 218972004, 218973009, 219009004, 219015004, 219019005, 219031007, 219036002, 219050007, 219063001, 230242007, 230304003, 230315008, 230316009, 230317000, 230333002, 230334008, 230346007, 230353003, 230354009, 230441003, 230665005, 230682007, 230760006, 230800004, 231451006, 231459008, 231466009, 231467000, 231481003, 231482005, 231920009, 231942005, 231962002, 231963007, 232032008, 232145009, 233637009, 233640009, 233672007, 233673002, 233687002, 233706004, 233725001, 233741000, 233945002, 234020005, 234171009, 234341005, 234366009, 234426009, 234645009, 234986004, 235022008, 235044003, 235051007, 235130007, 235619005, 235648004, 235650007, 235681003, 235705002, 235750000, 235751001, 235830005, 235837008, 235873001, 235874007, 235875008, 235876009, 235880004, 235881000, 235889003, 235942001, 235944000, 235955000, 236015007, 236074001, 236452000, 236505008, 236519008, 236520002, 236521003, 236814005, 237228001, 237449003, 237450003, 237509009, 237539004, 237545007, 237640005, 237641009, 237646004, 237665007, 237798008, 237809000, 237811009, 237844003, 237849008, 237853005, 237884007, 237908009, 238146000, 238503007, 238519000, 238635004, 238813009, 238814003, 238815002, 238894004, 238901000, 238988008, 238989000, 238990009, 238992001, 238995004, 238996003, 238997007, 238998002, 238999005, 239000007, 239845005, 239849004, 239900005, 240101000, 240102007, 240165007, 241746002, 241747006, 241748001, 241749009, 241750009, 241753006, 241754000, 241756003, 241757007, 241758002, 241761001, 241762008, 241763003, 241764009, 241767002, 241849000, 241937000, 241939002, 241947002, 241948007, 241949004, 241958006, 241959003, 241960008, 242060008, 242253008, 242255001, 242256000, 242258004, 242259007, 242260002, 242261003, 242262005, 242823008, 242824002, 242825001, 242828004, 242829007, 242830002, 242831003, 242832005, 242833000, 242834006, 242835007, 242975003, 34729003, 34738001, 34743008, 34764007, 34917002, 34958009, 35189002, 35224002, 35228004, 35528000, 35776002, 36193003, 36261000, 36265009, 36533005, 36558000, 36706008, 36784008, 37331004, 37370005, 37714008, 38149009, 38323006, 38879000, 38941006, 38968006, 39003006, 39502009, 39521001, 39679005, 39994006, 40382002, 40398009, 40550004, 40748005, 40844001, 40857009, 41047001, 41106001, 41136005, 41309000, 41390006, 41461004, 41602003, 41690001, 41839000, 42373003, 42377002, 42395004, 42524008, 42894006, 43125006, 43302000, 43599001, 43624006, 43695003, 43893007, 44003006, 44206008, 44838007, 44961008, 45088005, 45110008, 45421006, 45536007, 45701006, 45747003, 45775001, 46571000, 46604003, 46673008, 46721000, 46759008, 46878002, 47191007, 47307007, 47318007, 47546008, 47580006, 47743003, 47753002, 47774001, 47836003, 47923004, 48310002, 48445006, 48534006, 49020008, 49141004, 49450009, 49540005, 49929007, 49968000, 50026000, 50325005, 50393008, 50452004, 50529000, 50589003, 50601005, 50722006, 51160004, 51228004, 51339003, 51510002, 51814004, 51862005, 52020004, 52305004, 52367009, 52382004, 52671001, 52703008, 53001008, 53126001, 53275008, 53523003, 53556002, 53702006, 53796002, 54561008, 54594006, 54912002, 55043004, 55082002, 55410006, 55680006, 55720000, 55967005, 56194001, 56277006, 56698004, 56916002, 56918001, 57005003, 57305009, 57310008, 57319009, 57628003, 57695009, 58025002, 58104004, 58327003, 58700000, 58727001, 58960006, 59274003, 59369008, 59400006, 59826007, 60199004, 60454001, 60634005, 61351004, 61356009, 61406000, 61411003, 61438005, 61458006, 61544007, 61558009, 61704003, 61803000, 62014003, 62052002, 62276003, 62388003, 62473009, 62711001, 62942003, 63484008, 63628002, 63645007, 64415008, 64595005, 64600001, 64808005, 64921004, 65223000, 65242001, 65425009, 65435003, 65679006,

65941003, 65986000, 66351003, 66409006, 66413004, 66491001, 66595008, 66698002, 66780009, 66884007, 67438002, 67725009, 67794001, 67888001, 67893003, 67933004, 68099003, 68563002, 68629007, 68709002, 68822001, 69047008, 69434005, 69482004, 69819005, 69956002, 70273001, 70330008, 70655008, 70885001, 71277006, 72057009, 72120001, 72182007, 72527006, 72610007, 72700005, 72972005, 73108003, 73340005, 73383004, 73397007, 73692007, 73891003, 73935008, 74264003, 74265002, 74338006, 74524003, 74882009, 74902008, 74984006, 75496000, 75683007, 75831003, 76346005, 76400006, 76547000, 76636002, 76941008, 77131002, 77277003, 77607006, 77650001, 77718003, 77721001, 77857001, 78695006, 79044002, 79156004, 79815000, 79953006, 79982007, 80092001, 80258006, 80300008, 80471007, 396344000, 397696004, 397841007, 400072006, 402258000, 402260003, 402394004, 402428008, 402659000, 402662002, 402744003, 402745002, 402746001, 402747005, 402750008, 402751007, 402752000, 402753005, 402754004, 402756002, 402757006, 403265002, 403301006, 403303009, 403340001, 403606008, 403607004, 403608009, 403609001, 403610006, 403611005, 403612003, 403613008, 403614002, 403615001, 403616000, 403618004, 403619007, 403620001, 403621002, 403623004, 403625006, 403626007, 403627003, 403628008, 403629000, 403630005, 403631009, 403632002, 403635000, 403636004, 403637008, 403639006, 403640008, 403641007, 403642000, 403643005, 403645003, 403646002, 403647006, 403648001, 403649009, 403650009, 403651008, 403652001, 403653006, 403654000, 403655004, 403656003, 403657007, 403658002, 403659005, 403661001, 403662008, 403663003, 403664009, 403665005, 403666006, 403667002, 403668007, 403669004, 403670003, 403671004, 403672006, 403673001, 403675008, 403750002, 403872008, 404647000, 404694008, 405542005, 405544006, 406434004, 406447009, 406448004, 406593009, 407590002, 407674008, 408540003, 408667000, 408673004, 409564005, 410501000, 413183008, 413837004, 413851004, 414061000, 414689001, 414874007, 414996002, 415344002, 416073003, 416093006, 417229001, 417360004, 417895009, 417906002, 418108003, 418328005, 418370000, 418475009, 418505005, 418541005, 418572009, 418579000, 418603006, 418801006, 418944009, 418949004, 419007000, 419055000, 419056004, 419219000, 419253001, 419330004, 419347003, 419406002, 419476001, 419639006, 419737003, 419901001, 419914000, 420054005, 420059000, 420094007, 420545001, 421530001, 424338005, 425413006, 425581000, 425841004, 425974003, 426020004, 426364002, 426450003, 426790001, 426950009, 427046006, 427131001, 427180006, 427195008, 427399008, 427488005, 427590008, 427640001, 427670006, 427897002, 427975003, 428219007, 428383000, 428676003, 428720002, 428867008, 429299000, 429672007, 430136001, 430398007, 430400006, 430973002, 431307001, 439011002, 439014005, 439407002, 439591006, 439990003, 440385001, 441636005, 441681009, 441690002, 441858005, 442108004, 442197003, 442351006, 442654007, 442760001, 443478002, 254246009, 254247000, 254249002, 254250002, 254251003, 254252005, 254253000, 267206008, 267527002, 268796000, 268797009, 269265001, 269268004, 269269007, 269270008, 269271007, 269272000, 269688005, 269689002, 269722001, 269723006, 275387004, 275388009, 275400004, 275482009, 276581009, 276582002, 276628009, 277131000, 277804000, 277805004, 278023006, 278363000, 280955001, 280982009, 280983004, 280985006, 281510005, 287165005, 290133008, 290134002, 290135001, 290136000, 290137009, 290138004, 290139007, 290140009, 290145004, 290146003, 290147007, 290148002, 290149005, 290151009, 290152002, 290153007, 290154001, 290155000, 290156004, 290157008, 290158003, 290159006, 290160001, 290161002, 290162009, 290163004, 290164005, 290165006, 290166007, 290167003, 290168008, 290169000, 290171000, 290172007, 290173002, 290174008, 290175009, 290176005, 290177001, 290179003, 290180000, 290182008, 290183003, 290184009, 290185005, 290186006, 290188007, 290189004, 290190008, 290191007, 290193005, 290194004, 290196002, 290197006, 290198001, 290199009, 290200007, 290201006, 290202004, 290203009, 290204003, 290206001, 290207005, 290208000, 290209008, 290210003, 290211004, 290212006, 290213001, 290214007, 290215008, 290216009, 290217000, 290218005, 290219002, 290220008, 290221007, 290222000, 290226002, 290227006, 290228001, 290229009, 290230004, 290231000, 290232007, 290233002, 290234008, 290235009, 290236005, 290237001, 290238006, 290239003, 290240001, 290241002, 290242009, 290243004, 290244005, 290245006, 290247003, 290248008, 290249000, 290250000,

290251001, 290252008, 290253003, 290254009, 290255005, 290256006, 290257002, 290258007, 290260009, 290261008, 290263006, 290264000, 290265004, 290267007, 290268002, 290269005, 290270006, 290271005, 290272003, 290273008, 290274002, 290275001, 290276000, 290277009, 290278004, 290279007, 290280005, 290282002, 290283007, 290284001, 290285000, 290286004, 290287008, 290288003, 290289006, 290290002, 290291003, 290293000, 290294006, 290295007, 290297004, 290298009, 290299001, 290300009, 290301008, 290302001, 290303006, 290304000, 290305004, 290306003, 290307007, 290308002, 290309005, 290310000, 290311001, 290312008, 290313003, 290314009, 290315005, 290328003, 290332009, 290341004, 290342006, 290343001, 290344007, 290345008, 290346009, 290347000, 290348005, 290349002, 290350002, 290351003, 290352005, 290353000, 290354006, 290355007, 290356008, 290357004, 290358009, 290359001, 290360006, 290361005, 290362003, 290363008, 290364002, 290365001, 290366000, 290367009, 290368004, 290369007, 290370008, 290371007, 290372000, 290373005, 290374004, 290375003, 290376002, 290377006, 290378001, 290379009, 290380007, 290381006, 290382004, 290383009, 290384003, 290385002, 290386001, 290387005, 290388000, 290389008, 290390004, 290391000, 290392007, 290393002, 290394008, 290395009, 290396005, 290398006, 290399003, 290400005, 290401009, 290402002, 290403007, 290404001, 290405000, 290406004, 290407008, 290408003, 290409006, 290410001, 290411002, 290412009, 290413004, 290414005, 290415006, 290416007, 290417003, 290418008, 290419000, 290420006, 290421005, 290422003, 290423008, 290426000, 290428004, 290429007, 290430002, 290431003, 290432005, 290433000, 290434006, 290435007, 290436008, 290437004, 290438009, 290440004, 290441000, 290442007, 290443002, 290444008, 290445009, 290446005, 290447001, 290448006, 290449003, 290450003, 290451004, 290452006, 290453001, 290454007, 290455008, 290456009, 290457000, 290458005, 290459002, 290460007, 290461006, 290462004, 290463009, 290464003, 290465002, 290466001, 290467005, 290468000, 290469008, 290470009, 290474000, 290475004, 290476003, 290477007, 290478002, 290479005, 290480008, 290481007, 290483005, 290484004, 290485003, 290486002, 290487006, 290488001, 290489009, 290490000, 290491001, 290492008, 290493003, 290494009, 290495005, 290496006, 290497002, 290498007, 290500008, 290501007, 290502000, 290503005, 290504004, 290506002, 290507006, 290508001, 290509009, 290511000, 290513002, 290514008, 290515009, 290516005, 290517001, 290518006, 290519003, 290520009, 290521008, 290522001, 290523006, 290524000, 290525004, 290526003, 290527007, 290528002, 290529005, 290530000, 290531001, 290532008, 290533003, 290534009, 290535005, 290536006, 290537002, 290538007, 290539004, 290541003, 290542005, 290544006, 290545007, 290546008, 290547004, 290548009, 290549001, 290550001, 290551002, 290552009, 290553004, 290555006, 290556007, 290557003, 290558008, 290559000, 290560005, 290561009, 290562002, 290563007, 290564001, 290565000, 290566004, 290568003, 290569006, 290571006, 290572004, 290574003, 290575002, 290576001, 290577005, 290578000, 290579008, 290580006, 290581005, 290582003, 290583008, 290584002, 290585001, 290586000, 290587009, 290588004, 290589007, 290590003, 290594007, 290595008, 290596009, 290597000, 290598005, 290599002, 290601000, 290602007, 290603002, 290604008, 290605009, 290606005, 290607001, 290608006, 290609003, 290610008, 290611007, 290612000, 290613005, 290614004, 290615003, 290616002, 290617006, 290618001, 290619009, 290620003, 290621004, 290622006, 290623001, 290624007, 290625008, 290626009, 290627000, 290628005, 290629002, 290630007, 290631006, 290632004, 290633009, 290634003, 290635002, 290636001, 290637005, 290638000, 290639008, 290640005, 290641009, 290642002, 290643007, 290644001, 290645000, 290646004, 290647008, 290648003, 290650006, 290651005, 290652003, 290654002, 290655001, 290656000, 290657009, 290658004, 290659007, 290660002, 290661003, 290662005, 290663000, 290664006, 290665007, 290666008, 290667004, 290668009, 290669001, 290670000, 290671001, 290672008, 290673003, 290674009, 290675005, 290676006, 290677002, 290678007, 290679004, 290680001, 290681002, 290682009, 290683004, 290684005, 290685006, 290686007, 290687003, 290688008, 290689000, 290690009, 290692001, 290693006, 290694000, 290695004, 290696003, 290697007, 290698002, 290699005, 290700006, 290701005, 290702003, 290703008, 290705001, 290706000, 290707009, 290708004, 290709007, 290710002, 290711003, 290712005, 290713000, 290714006, 290715007, 290716008, 290717004, 290718009, 290719001, 290720007,

290721006, 290722004, 290723009, 290724003, 290725002, 290726001, 290727005, 290728000, 290729008, 290730003, 290731004, 290732006, 290733001, 290734007, 290735008, 290736009, 290737000, 290738005, 290739002, 290740000, 290741001, 290742008, 290743003, 290744009, 290745005, 290746006, 290748007, 290749004, 290750004, 290751000, 290752007, 290753002, 290754008, 290755009, 290756005, 290757001, 290758006, 290759003, 290760008, 290761007, 290762000, 290763005, 290764004, 290765003, 290766002, 290767006, 290768001, 290769009, 290770005, 290771009, 290772002, 290773007, 290774001, 290775000, 290776004, 290777008, 290778003, 290779006, 290781008, 290783006, 290784000, 290785004, 290786003, 290787007, 290789005, 290790001, 290791002, 290792009, 290793004, 290794005, 290795006, 290796007, 290797003, 290798008, 290799000, 290800001, 290801002, 290802009, 290803004, 290804005, 290805006, 290808008, 290809000, 290810005, 290811009, 290812002, 290813007, 290814001, 290815000, 290816004, 290817008, 290818003, 290819006, 290820000, 290821001, 290822008, 290823003, 290824009, 290825005, 290826006, 290827002, 290828007, 290829004, 290830009, 290831008, 290832001, 290833006, 290835004, 290836003, 290837007, 290838002, 290839005, 290840007, 290841006, 290842004, 290843009, 290844003, 290845002, 290846001, 290847005, 290848000, 290849008, 290850008, 290851007, 290852000, 290853005, 290854004, 290855003, 290856002, 290857006, 290858001, 290859009, 290860004, 290861000, 290862007, 290863002, 290864008, 290865009, 290866005, 290867001, 290868006, 290869003, 290870002, 290871003, 290872005, 290873000, 290874006, 290875007, 290876008, 290877004, 290878009, 290879001, 290884007, 290885008, 290886009, 290887000, 290888005, 290889002, 290890006, 290891005, 290892003, 290893008, 290894002, 290895001, 290896000, 290897009, 290898004, 290899007, 290900002, 290901003, 290902005, 290903000, 290904006, 290905007, 290906008, 290908009, 290909001, 290910006, 290911005, 290912003, 290913008, 290914002, 290915001, 290916000, 290917009, 290918004, 290919007, 290920001, 290921002, 290922009, 290923004, 290924005, 290925006, 290926007, 290927003, 290928008, 290929000, 290930005, 290931009, 290932002, 290933007, 290934001, 290935000, 290936004, 290937008, 290938003, 290939006, 290940008, 290941007, 290942000, 290943005, 290944004, 290945003, 290946002, 290947006, 290948001, 290949009, 290950009, 290951008, 290952001, 290953006, 290954000, 290956003, 290957007, 290958002, 290959005, 290960000, 290961001, 290962008, 290963003, 290964009, 290965005, 290966006, 290967002, 290968007, 290969004, 290970003, 290972006, 290973001, 290974007, 290975008, 290976009, 290977000, 290978005, 290979002, 290980004, 290981000, 290982007, 290983002, 290985009, 290986005, 290987001, 290988006, 290989003, 290990007, 290991006, 290992004, 290994003, 290995002, 290996001, 290997005, 290998000, 290999008, 291000007, 291001006, 291003009, 291004003, 291006001, 291007005, 291010003, 291011004, 291015008, 291016009, 291017000, 291018005, 291020008, 291021007, 291023005, 291024004, 291026002, 291027006, 291028001, 291029009, 291030004, 291031000, 291033002, 291034008, 291035009, 291036005, 291037001, 291038006, 291039003, 291040001, 291041002, 291042009, 291044005, 291045006, 291046007, 291047003, 291048008, 291049000, 291050000, 291051001, 291052008, 291053003, 291054009, 291055005, 291056006, 291057002, 291059004, 291060009, 291061008, 291062001, 291063006, 291064000, 291065004, 291066003, 291067007, 291068002, 291069005, 291070006, 291071005, 291072003, 291074002, 291075001, 291076000, 291077009, 291078004, 291079007, 291080005, 291081009, 291082002, 291083007, 291084001, 291085000, 291086004, 291087008, 291089006, 291090002, 291092005, 291093000, 291094006, 291095007, 291096008, 291098009, 291099001, 291100009, 291102001, 291103006, 291104000, 291105004, 291106003, 291107007, 291108002, 291110000, 291111001, 291112008, 291113003, 291114009, 291115005, 291116006, 291117002, 291118007, 291119004, 291120005, 291121009, 291122002, 291123007, 291124001, 291125000, 291126004, 291127008, 291128003, 291129006, 291130001, 291131002, 291132009, 291133004, 291134005, 291135006, 291136007, 291137003, 291138008, 291139000, 291140003, 291141004, 291142006, 291143001, 291144007, 291145008, 291147000, 291148005, 291150002, 291151003, 291152005, 291153000, 291154006, 291155007, 291156008, 291157004, 291158009, 291159001, 291160006, 291161005, 291162003, 291163008, 291164002, 291165001, 291166000, 291167009, 291168004, 291169007, 291170008, 291171007,

291172000, 291174004, 291175003, 291176002, 291177006, 291178001, 291179009, 291181006, 291182004, 291183009, 291184003, 291185002, 291186001, 291188000, 291189008, 291190004, 291191000, 291192007, 291193002, 291194008, 291195009, 291196005, 291197001, 291199003, 291200000, 291201001, 291202008, 291203003, 291204009, 291205005, 291206006, 291207002, 291208007, 291209004, 291211008, 291212001, 291213006, 291214000, 291215004, 291216003, 291217007, 291218002, 291219005, 291220004, 291221000, 291222007, 291223002, 291224008, 291225009, 291226005, 291227001, 291228006, 291229003, 291230008, 291231007, 291232000, 291233005, 291234004, 291235003, 291236002, 291237006, 291238001, 291239009, 291241005, 291242003, 291246000, 291247009, 291248004, 291253009, 291254003, 291256001, 291257005, 291261004, 291263001, 291264007, 291266009, 291267000, 291268005, 291269002, 291270001, 291271002, 291272009, 291273004, 291274005, 291275006, 291276007, 291277003, 291279000, 291280002, 291282005, 291283000, 291284006, 291285007, 291286008, 291287004, 291288009, 291289001, 291291009, 291292002, 291294001, 291295000, 291297008, 291298003, 291299006, 291300003, 291301004, 291302006, 291303001, 291304007, 291305008, 291306009, 291307000, 291308005, 291309002, 291310007, 291313009, 291314003, 291315002, 291317005, 291318000, 291320002, 291321003, 291323000, 291324006, 291326008, 291327004, 291329001, 291330006, 291331005, 291332003, 291333008, 291334002, 291337009, 291338004, 291339007, 291340009, 291341008, 291342001, 291343006, 291344000, 291345004, 291346003, 291347007, 291350005, 291351009, 291352002, 291353007, 291354001, 291355000, 291356004, 291357008, 291358003, 291359006, 291360001, 291361002, 291362009, 291363004, 291364005, 291365006, 291366007, 291367003, 291368008, 291371000, 291372007, 291373002, 291374008, 291375009, 291376005, 291377001, 291378006, 291379003, 291380000, 291381001, 291382008, 291383003, 291384009, 291388007, 291389004, 291390008, 291393005, 291395003, 291396002, 291398001, 291399009, 291400002, 291401003, 291402005, 291403000, 291404006, 291405007, 291406008, 291407004, 291408009, 291409001, 291411005, 291412003, 291413008, 291414002, 291423004, 291424005, 291425006, 291426007, 291427003, 291428008, 291429000, 291430005, 291431009, 291432002, 291433007, 291434001, 291435000, 291436004, 291437008, 291438003, 291439006, 291440008, 291441007, 291442000, 291443005, 291444004, 291445003, 291446002, 291447006, 291448001, 291449009, 291450009, 291452001, 291453006, 291454000, 291455004, 291456003, 291457007, 291458002, 291459005, 291460000, 291461001, 291462008, 291463003, 291467002, 291468007, 291469004, 291470003, 291471004, 291472006, 291473001, 291474007, 291475008, 291476009, 291477000, 291478005, 291479002, 291480004, 291481000, 291482007, 291483002, 291484008, 291485009, 291486005, 291487001, 291488006, 291489003, 291490007, 291491006, 291492004, 291493009, 291494003, 291495002, 291496001, 291497005, 291498000, 291499008, 291500004, 291501000, 291502007, 291503002, 291504008, 291505009, 291506005, 291507001, 291508006, 291509003, 291510008, 291512000, 291513005, 291514004, 291515003, 291516002, 291517006, 291518001, 291519009, 291520003, 291521004, 291522006, 291523001, 291524007, 291525008, 291526009, 291527000, 291528005, 291529002, 291530007, 291531006, 291532004, 291533009, 291534003, 291535002, 291536001, 291537005, 291538000, 291539008, 291540005, 291541009, 291542002, 291543007, 291544001, 291545000, 291546004, 291547008, 291548003, 291549006, 291550006, 291551005, 291552003, 291553008, 291554002, 291555001, 291556000, 291557009, 291558004, 291559007, 291560002, 291561003, 291562005, 291563000, 291564006, 291565007, 291566008, 291567004, 291568009, 291569001, 291570000, 291571001, 291572008, 291573003, 291574009, 291575005, 291576006, 291577002, 291578007, 291579004, 291580001, 291581002, 291582009, 291583004, 291584005, 291585006, 291586007, 291587003, 291588008, 291589000, 291590009, 291591008, 291592001, 291593006, 291594000, 291595004, 291596003, 291597007, 291598002, 291599005, 291600008, 291601007, 291602000, 291603005, 291604004, 291605003, 291606002, 291607006, 291608001, 291609009, 291610004, 291611000, 291613002, 291614008, 291615009, 291616005, 291617001, 291618006, 291619003, 291620009, 291621008, 291622001, 291623006, 291624000, 291625004, 291626003, 291627007, 291628002, 291629005, 291630000, 291631001, 291632008, 291633003, 291634009, 291635005, 291636006, 291637002, 291638007, 291639004, 291640002, 291641003, 291642005, 291643000, 291644006,

291645007, 291646008, 291647004, 291648009, 291649001, 291650001, 291651002, 291652009, 291653004, 291654005, 291655006, 291656007, 291657003, 291658008, 291659000, 291660005, 291662002, 291663007, 291664001, 291666004, 291667008, 291668003, 291669006, 291670007, 291671006, 291672004, 291673009, 291674003, 291675002, 291676001, 291677005, 291678000, 291679008, 291680006, 291681005, 291682003, 291683008, 291684002, 291685001, 291686000, 291687009, 291688004, 291689007, 291690003, 291691004, 291692006, 291693001, 291694007, 291695008, 291696009, 291697000, 291698005, 291699002, 291700001, 291782006, 291783001, 291784007, 291785008, 291786009, 291787000, 291788005, 291789002, 291791005, 291792003, 291793008, 291794002, 291795001, 291796000, 291797009, 291798004, 291799007, 291800006, 291801005, 291802003, 291803008, 291804002, 291805001, 291806000, 291807009, 291808004, 291809007, 291810002, 291811003, 291812005, 291813000, 291814006, 291815007, 291817004, 291818009, 291819001, 291820007, 291821006, 291822004, 291823009, 291824003, 291825002, 291826001, 291827005, 291828000, 291829008, 291830003, 291831004, 291832006, 291833001, 291834007, 291835008, 291836009, 291837000, 291838005, 291839002, 291840000, 291841001, 291842008, 291843003, 291844009, 291845005, 291846006, 291847002, 291848007, 291849004, 291850004, 291851000, 291852007, 291853002, 291854008, 291855009, 291856005, 291857001, 291858006, 291859003, 291860008, 291861007, 291862000, 291863005, 291865003, 291866002, 291867006, 291868001, 291869009, 291870005, 291871009, 291872002, 291873007, 291874001, 291875000, 291876004, 291877008, 291878003, 291879006, 291880009, 291881008, 291882001, 291883006, 291884000, 291885004, 291886003, 291887007, 291888002, 291889005, 291890001, 291891002, 291892009, 291893004, 291894005, 291895006, 291896007, 291897003, 291898008, 291899000, 291900005, 291901009, 291902002, 291903007, 291904001, 291905000, 291906004, 291907008, 291908003, 291909006, 291910001, 291914005, 291922003, 291923008, 291924002, 291925001, 291926000, 291927009, 291928004, 291929007, 291930002, 291931003, 291932005, 291933000, 291934006, 291940004, 291941000, 291942007, 291943002, 291944008, 291945009, 291946005, 291947001, 291948006, 291949003, 291950003, 291951004, 291952006, 291953001, 291954007, 291955008, 291956009, 291957000, 291958005, 291959002, 291960007, 291961006, 291962004, 291963009, 291964003, 291965002, 291966001, 291967005, 291968000, 291969008, 291970009, 291971008, 291972001, 291973006, 291974000, 291975004, 291976003, 291977007, 291978002, 291979005, 291980008, 291981007, 291982000, 291983005, 291984004, 291985003, 291986002, 291987006, 291988001, 291989009, 291990000, 291991001, 291992008, 291993003, 291994009, 291995005, 291996006, 291997002, 291998007, 292000005, 292001009, 292003007, 292004001, 292005000, 292006004, 292007008, 292008003, 292009006, 292010001, 292011002, 292012009, 292013004, 292014005, 292015006, 292017003, 292018008, 292023008, 292024002, 292025001, 292026000, 292027009, 292028004, 292029007, 292030002, 292031003, 292032005, 292033000, 292034006, 292035007, 292036008, 292037004, 292040004, 292041000, 292042007, 292044008, 292045009, 292046005, 292047001, 292048006, 292049003, 292050003, 292051004, 292052006, 292054007, 292055008, 292056009, 292057000, 292058005, 292059002, 292062004, 292063009, 292064003, 292065002, 292066001, 292067005, 292069008, 292070009, 292071008, 292072001, 292073006, 292075004, 292076003, 292078002, 292080008, 292082000, 292084004, 292085003, 292086002, 292088001, 292089009, 292090000, 292091001, 292101006, 292102004, 292103009, 292105002, 292106001, 292107005, 292108000, 292109008, 292110003, 292111004, 292112006, 292113001, 292114007, 292115008, 292118005, 292120008, 292121007, 292122000, 292123005, 292124004, 292125003, 292126002, 292127006, 292128001, 292129009, 292130004, 292131000, 292132007, 292133002, 292135009, 292136005, 292137001, 292138006, 292139003, 292140001, 292141002, 292142009, 292143004, 292144005, 292145006, 292146007, 292147003, 292148008, 292149000, 292150000, 292151001, 292152008, 292153003, 292154009, 292155005, 292156006, 292157002, 292158007, 292159004, 292162001, 292163006, 292164000, 292165004, 292166003, 292167007, 292169005, 292170006, 292171005, 292172003, 292173008, 292174002, 292175001, 292176000, 292177009, 292178004, 292179007, 292180005, 292181009, 292182002, 292184001, 292185000, 292186004, 292188003, 292189006, 292190002, 292191003, 292192005, 292193000, 292194006, 292195007, 292196008, 292197004, 292198009,

292199001, 292200003, 292201004, 292202006, 292203001, 292204007, 292205008, 292206009, 292207000, 292208005, 292209002, 292210007, 292212004, 292213009, 292215002, 292216001, 292217005, 292218000, 292220002, 292221003, 292222005, 292223000, 292224006, 292225007, 292226008, 292227004, 292228009, 292229001, 292230006, 292231005, 292232003, 292233008, 292234002, 292235001, 292236000, 292237009, 292238004, 292239007, 292240009, 292241008, 292242001, 292243006, 292244000, 292245004, 292247007, 292248002, 292249005, 292250005, 292251009, 292252002, 292253007, 292254001, 292255000, 292256004, 292257008, 292259006, 292260001, 292261002, 292262009, 292263004, 292264005, 292266007, 292267003, 292268008, 292269000, 292270004, 292272007, 292273002, 292274008, 292275009, 292276005, 292277001, 292278006, 292279003, 292280000, 292281001, 292282008, 292283003, 292284009, 292285005, 292286006, 292287002, 292288007, 292289004, 292290008, 292291007, 292292000, 292293005, 292294004, 292295003, 292296002, 292297006, 292298001, 292301002, 292302009, 292303004, 292304005, 292305006, 292306007, 292307003, 292308008, 292309000, 292310005, 292311009, 292312002, 292313007, 292314001, 292315000, 292316004, 292317008, 292318003, 292319006, 292320000, 292322008, 292323003, 292324009, 292325005, 292326006, 292327002, 292328007, 292329004, 292331008, 292332001, 292333006, 292334000, 292335004, 292336003, 292338002, 292339005, 292340007, 292342004, 292344003, 292345002, 292346001, 292347005, 292348000, 292349008, 292350008, 292351007, 292352000, 292353005, 292355003, 292356002, 292357006, 292358001, 292359009, 292360004, 292361000, 292362007, 292363002, 292364008, 292367001, 292368006, 292369003, 292370002, 292371003, 292372005, 292373000, 292374006, 292375007, 292376008, 292377004, 292378009, 292379001, 292380003, 292381004, 292382006, 292383001, 292384007, 292385008, 292386009, 292387000, 292388005, 292389002, 292390006, 292391005, 292392003, 292393008, 292394002, 292395001, 292396000, 292397009, 292398004, 292399007, 292400000, 292401001, 292402008, 292403003, 292404009, 292405005, 292406006, 292407002, 292408007, 292409004, 292410009, 292411008, 292412001, 292413006, 292414000, 292415004, 292417007, 292419005, 292420004, 292421000, 292424008, 292425009, 292426005, 292427001, 292428006, 292430008, 292431007, 292432000, 292433005, 292434004, 292435003, 292436002, 292437006, 292438001, 292439009, 292440006, 292441005, 292442003, 292443008, 292444002, 292445001, 292446000, 292447009, 292448004, 292449007, 292450007, 292451006, 292452004, 292453009, 292454003, 292455002, 292456001, 292457005, 292458000, 292459008, 292460003, 292461004, 292462006, 292463001, 292465008, 292466009, 292468005, 292469002, 292470001, 292471002, 292472009, 292473004, 292474005, 292475006, 292476007, 292477003, 292478008, 292479000, 292480002, 292482005, 292483000, 292484006, 292485007, 292486008, 292487004, 292488009, 292489001, 292490005, 292491009, 292492002, 292493007, 292494001, 292495000, 292496004, 292497008, 292498003, 292499006, 292500002, 292501003, 292502005, 292503000, 292504006, 292505007, 292506008, 292507004, 292508009, 292509001, 292510006, 292512003, 292514002, 292515001, 292516000, 292517009, 292518004, 292519007, 292520001, 292521002, 292522009, 292523004, 292525006, 292527003, 292528008, 292530005, 292531009, 292532002, 292533007, 292534001, 292535000, 292536004, 292537008, 292538003, 292539006, 292540008, 292541007, 292542000, 292543005, 292544004, 292545003, 292546002, 292547006, 292549009, 292551008, 292553006, 292554000, 292555004, 292556003, 292560000, 292561001, 292563003, 292564009, 292565005, 292573001, 292581000, 292587001, 292591006, 292592004, 292593009, 292598000, 292600006, 292603008, 292605001, 292606000, 292609007, 292610002, 292615007, 292616008, 292617004, 292618009, 292619001, 292623009, 292624003, 292627005, 292628000, 292629008, 292630003, 292636009, 292637000, 292641001, 292642008, 292650004, 292651000, 292659003, 292665003, 292666002, 292667006, 292668001, 292669009, 292671009, 292675000, 292676004, 292677008, 292679006, 292682001, 292684000, 292685004, 292687007, 292688002, 292689005, 292690001, 292691002, 292692009, 292693004, 292694005, 292695006, 292696007, 292699000, 292700004, 292701000, 292702007, 292704008, 292709003, 292710008, 292711007, 292712000, 292713005, 292714004, 292716002, 292717006, 292718001, 292719009, 292720003, 292723001, 292724007, 292725008, 292726009, 292727000, 292729002, 292730007, 292731006, 292732004, 292734003, 292735002, 292737005, 292738000, 292740005,

292742002, 292744001, 292745000, 292748003, 292752003, 292756000, 292759007, 292760002, 292761003, 292762005, 292763000, 292764006, 292767004, 292769001, 292771001, 292777002, 292779004, 292785006, 292786007, 292793006, 292794000, 292795004, 292796003, 292798002, 292799005, 292801008, 292802001, 292803006, 292805004, 292806003, 292808002, 292809005, 292810000, 292811001, 292812008, 292814009, 292815005, 292816006, 292817002, 292818007, 292819004, 292820005, 292821009, 292822002, 292823007, 292824001, 292825000, 292826004, 292827008, 292828003, 292829006, 292831002, 292832009, 292833004, 292834005, 292835006, 292837003, 292838008, 292839000, 292840003, 292841004, 292842006, 292843001, 292844007, 292845008, 292846009, 292847000, 292848005, 292849002, 292850002, 292851003, 292852005, 292853000, 292854006, 292855007, 292858009, 292859001, 292860006, 292863008, 292864002, 292865001, 292866000, 292867009, 292868004, 292870008, 292875003, 292877006, 292879009, 292883009, 292885002, 292886001, 292887005, 292888000, 292890004, 292891000, 292894008, 292896005, 292897001, 292898006, 292899003, 292900008, 292901007, 292902000, 292903005, 292904004, 292906002, 292907006, 292908001, 292909009, 292910004, 292911000, 292912007, 292913002, 292914008, 292915009, 292917001, 292918006, 292919003, 292920009, 292922001, 292923006, 292924000, 292925004, 292927007, 292929005, 292933003, 292934009, 292936006, 292937002, 292938007, 292939004, 292940002, 292941003, 292942005, 292943000, 292944006, 292945007, 292946008, 292947004, 292949001, 292950001, 292951002, 292952009, 292953004, 292955006, 292956007, 292957003, 292958008, 292959000, 292960005, 292961009, 292962002, 292963007, 292964001, 292965000, 292966004, 292967008, 292968003, 292969006, 292970007, 292971006, 292972004, 292973009, 292974003, 292975002, 292976001, 292977005, 292978000, 292979008, 292980006, 292981005, 292982003, 292985001, 292986000, 292988004, 292990003, 292991004, 292994007, 292995008, 292996009, 292997000, 292998005, 292999002, 293000001, 293001002, 293002009, 293003004, 293004005, 293005006, 293006007, 293007003, 293008008, 293009000, 293010005, 293011009, 293012002, 293013007, 293014001, 293015000, 293016004, 293017008, 293019006, 293020000, 293021001, 293023003, 293024009, 293025005, 293027002, 293028007, 293029004, 293030009, 293031008, 293034000, 293035004, 293036003, 293037007, 293038002, 293039005, 293040007, 293041006, 293042004, 293043009, 293044003, 293046001, 293049008, 293050008, 293052000, 293054004, 293055003, 293056002, 293057006, 293058001, 293059009, 293060004, 293062007, 293064008, 293066005, 293068006, 293069003, 293070002, 293071003, 293073000, 293074006, 293075007, 293076008, 293078009, 293079001, 293080003, 293081004, 293082006, 293084007, 293085008, 293097009, 293100004, 293101000, 293102007, 293103002, 293129002, 293131006, 293132004, 293135002, 293136001, 293138000, 293139008, 293140005, 293141009, 293142002, 293144001, 293146004, 293147008, 293148003, 293149006, 293150006, 293151005, 293152003, 293153008, 293154002, 293155001, 293156000, 293157009, 293158004, 293159007, 293160002, 293161003, 293162005, 293163000, 293164006, 293165007, 293166008, 293167004, 293169001, 293170000, 293171001, 293172008, 293173003, 293174009, 293175005, 293176006, 293177002, 293183004, 293184005, 293192001, 293193006, 293194000, 293195004, 293196003, 293197007, 293198002, 293200008, 293201007, 293203005, 293205003, 293206002, 293207006, 293208001, 293209009, 293210004, 293211000, 293212007, 293213002, 293214008, 293215009, 293216005, 293217001, 293218006, 293219003, 293220009, 293221008, 293222001, 293223006, 293224000, 293225004, 293226003, 293227007, 293228002, 293229005, 293230000, 293232008, 293234009, 293236006, 293237002, 293238007, 293239004, 293240002, 293241003, 293242005, 293243000, 293244006, 293245007, 293246008, 293247004, 293248009, 293249001, 293250001, 293251002, 293252009, 293253004, 293254005, 293255006, 293256007, 293257003, 293261009, 293262002, 293263007, 293265000, 293266004, 293268003, 293269006, 293270007, 293271006, 293274003, 293275002, 293276001, 293277005, 293278000, 293279008, 293280006, 293281005, 293282003, 293283008, 293285001, 293288004, 293289007, 293292006, 293293001, 293294007, 293295008, 293300005, 293301009, 293302002, 293303007, 293305000, 293306004, 293307008, 293308003, 293309006, 293310001, 293311002, 293312009, 293315006, 293317003, 293331003, 293332005, 293333000, 293334006, 293335007, 293336008, 293337004, 293339001, 293340004, 293341000, 293343002, 293344008, 293346005,

293347001, 293348006, 293349003, 293350003, 293351004, 293352006, 293354007, 293367005, 293370009, 293371008, 293372001, 293373006, 293374000, 293375004, 293380008, 293383005, 293384004, 293385003, 293389009, 293390000, 293392008, 293393003, 293394009, 293395005, 293396006, 293397002, 293398007, 293400006, 293401005, 293402003, 293403008, 293404002, 293406000, 293407009, 293408004, 293417004, 293418009, 293420007, 293421006, 293422004, 293423009, 293424003, 293427005, 293432006, 293433001, 293434007, 293435008, 293436009, 293437000, 293438005, 293440000, 293441001, 293442008, 293443003, 293444009, 293445005, 293446006, 293447002, 293448007, 293449004, 293450004, 293451000, 293452007, 293453002, 293454008, 293455009, 293456005, 293457001, 293458006, 293459003, 293460008, 293461007, 293462000, 293463005, 293464004, 293465003, 293466002, 293467006, 293468001, 293469009, 293470005, 293471009, 293472002, 293473007, 293482001, 293483006, 293484000, 293485004, 293487007, 293488002, 293489005, 293490001, 293491002, 293494005, 293495006, 293496007, 293497003, 293498008, 293499000, 293500009, 293501008, 293502001, 293503006, 293504000, 293505004, 293506003, 293507007, 293508002, 293509005, 293511001, 293512008, 293513003, 293515005, 293516006, 293517002, 293518007, 293519004, 293520005, 293521009, 293523007, 293524001, 293528003, 293529006, 293530001, 293531002, 293533004, 293535006, 293536007, 293537003, 293538008, 293539000, 293540003, 293542006, 293543001, 293545008, 293546009, 293547000, 293548005, 293549002, 293550002, 293551003, 293552005, 293553000, 293554006, 293555007, 293557004, 293558009, 293559001, 293560006, 293561005, 293562003, 293563008, 293565001, 293566000, 293567009, 293568004, 293571007, 293572000, 293573005, 293574004, 293575003, 293576002, 293577006, 293578001, 293579009, 295121001, 295122008, 295123003, 295124009, 295125005, 295126006, 295127002, 295128007, 295129004, 295130009, 295134000, 295135004, 295136003, 295137007, 295138002, 295139005, 295140007, 295141006, 295142004, 295143009, 295144003, 295145002, 295146001, 295147005, 295148000, 295149008, 295150008, 295151007, 295152000, 295153005, 295154004, 295155003, 295156002, 295157006, 295158001, 295159009, 295160004, 295161000, 295163002, 295164008, 295165009, 295166005, 295167001, 295168006, 295169003, 295170002, 295171003, 295172005, 295173000, 295174006, 295175007, 295176008, 295177004, 295178009, 295179001, 295180003, 295181004, 295182006, 295183001, 295184007, 295185008, 295186009, 295187000, 295188005, 295189002, 295190006, 295191005, 295192003, 295193008, 295194002, 295195001, 295196000, 295197009, 295198004, 295199007, 295200005, 295201009, 295202002, 295203007, 295204001, 295205000, 295206004, 295207008, 295208003, 295209006, 295210001, 295211002, 295212009, 295213004, 295217003, 295218008, 295219000, 295220006, 295221005, 295222003, 295223008, 295224002, 295225001, 295226000, 295227009, 295228004, 295229007, 295230002, 295231003, 295232005, 295233000, 295234006, 295235007, 295236008, 295237004, 295238009, 295239001, 295240004, 295241000, 295242007, 295243002, 295244008, 295245009, 295246005, 295247001, 295248006, 295249003, 295250003, 295251004, 295252006, 295253001, 295254007, 295255008, 295256009, 295257000, 295258005, 295259002, 295260007, 295261006, 295262004, 295264003, 295265002, 295266001, 295267005, 295268000, 295269008, 295270009, 295271008, 295272001, 295273006, 295274000, 295275004, 295276003, 295277007, 295278002, 295279005, 295280008, 295281007, 295282000, 295283005, 295284004, 295285003, 295286002, 295287006, 295288001, 295289009, 295290000, 295291001, 295292008, 295293003, 295294009, 295295005, 295296006, 295297002, 295298007, 295299004, 295300007, 295301006, 295302004, 295303009, 295304003, 295305002, 295306001, 295307005, 295308000, 295309008, 295310003, 295311004, 295312006, 295313001, 295314007, 295316009, 295317000, 295318005, 295319002, 295322000, 295328001, 295329009, 295330004, 295331000, 295332007, 295333002, 295334008, 295335009, 295336005, 295337001, 295338006, 295339003, 295340001, 295341002, 295342009, 295343004, 295344005, 295345006, 295346007, 295347003, 295348008, 295349000, 295350000, 295351001, 295352008, 295353003, 295354009, 295355005, 295356006, 295357002, 295359004, 295360009, 295361008, 295362001, 295363006, 295364000, 295365004, 295367007, 295368002, 295369005, 295370006, 295371005, 295372003, 295373008, 295374002, 295375001, 295376000, 295377009, 295378004, 295379007, 295380005, 295381009, 295382002, 295383007, 295384001, 295385000, 295386004, 295387008,

295388003, 295389006, 295390002, 295391003, 295392005, 295393000, 295394006, 295395007, 295396008, 295397004, 295398009, 295399001, 295400008, 295401007, 295402000, 295403005, 295404004, 295405003, 295406002, 295407006, 295408001, 295409009, 295410004, 295411000, 295412007, 295413002, 295414008, 295415009, 295416005, 295417001, 295418006, 295419003, 295420009, 295421008, 295422001, 295423006, 295424000, 295425004, 295426003, 295431001, 295432008, 295433003, 295434009, 295435005, 295436006, 295437002, 295438007, 295439004, 295440002, 295441003, 295442005, 295443000, 295444006, 295445007, 295446008, 295447004, 295448009, 295449001, 295450001, 295451002, 295452009, 295453004, 295454005, 295455006, 295456007, 295457003, 295458008, 295459000, 295460005, 295461009, 295462002, 295463007, 295464001, 295465000, 295466004, 295468003, 295469006, 295470007, 295471006, 295472004, 295473009, 295474003, 295475002, 295476001, 295477005, 295478000, 295479008, 295480006, 295481005, 295482003, 295483008, 295484002, 295485001, 295486000, 295487009, 295488004, 295489007, 295490003, 295491004, 295492006, 295493001, 295494007, 295495008, 295497000, 295498005, 295499002, 295500006, 295501005, 295502003, 295503008, 295504002, 295505001, 295506000, 295507009, 295508004, 295510002, 295511003, 295512005, 295513000, 295514006, 295515007, 295516008, 295517004, 295518009, 295519001, 295520007, 295521006, 295522004, 295523009, 295524003, 295525002, 295526001, 295527005, 295528000, 295529008, 295530003, 295531004, 295532006, 295533001, 295534007, 295535008, 295536009, 295537000, 295538005, 295539002, 295540000, 295541001, 295542008, 295543003, 295544009, 295545005, 295546006, 295547002, 295548007, 295549004, 295550004, 295551000, 295552007, 295553002, 295554008, 295555009, 295556005, 295557001, 295558006, 295559003, 295560008, 295561007, 295562000, 295563005, 295564004, 295565003, 295566002, 295567006, 295568001, 295570005, 295571009, 295572002, 295573007, 295574001, 295575000, 295576004, 295577008, 295578003, 295579006, 295580009, 295581008, 295582001, 295583006, 295584000, 295585004, 295586003, 295587007, 295588002, 295589005, 295590001, 295593004, 295594005, 295595006, 295596007, 295597003, 295598008, 295599000, 295600002, 295601003, 295602005, 295603000, 295604006, 295605007, 295606008, 295607004, 295608009, 295609001, 295610006, 295611005, 295612003, 295613008, 295614002, 295615001, 295616000, 295617009, 295618004, 295619007, 295620001, 295621002, 295622009, 295623004, 295624005, 295625006, 295626007, 295627003, 295628008, 295629000, 295630005, 295631009, 295632002, 295633007, 295634001, 295635000, 295636004, 295637008, 295638003, 295639006, 295640008, 295641007, 295642000, 295643005, 295644004, 295645003, 295646002, 295647006, 295648001, 295649009, 295650009, 295651008, 295652001, 295653006, 295654000, 295655004, 295656003, 295657007, 295658002, 295659005, 295660000, 295661001, 295662008, 295663003, 295664009, 295665005, 295666006, 295667002, 295668007, 295669004, 295671004, 295672006, 295673001, 295674007, 295675008, 295676009, 295677000, 295678005, 295679002, 295680004, 295681000, 295682007, 295683002, 295684008, 295685009, 295686005, 295687001, 295688006, 295689003, 295690007, 295691006, 295692004, 295693009, 295694003, 295695002, 295696001, 295697005, 295698000, 295699008, 295700009, 295701008, 295702001, 295703006, 295704000, 295705004, 295706003, 295707007, 295708002, 295709005, 295710000, 295711001, 295712008, 295713003, 295714009, 295715005, 295716006, 295717002, 295718007, 295720005, 295721009, 295722002, 295724001, 295725000, 295726004, 295727008, 295728003, 295729006, 295730001, 295731002, 295732009, 295733004, 295734005, 295735006, 295736007, 295737003, 295738008, 295739000, 295740003, 295741004, 295742006, 295743001, 295744007, 295745008, 295746009, 295747000, 295748005, 295749002, 295750002, 295751003, 295752005, 295753000, 295754006, 295755007, 295756008, 295757004, 295758009, 295759001, 295760006, 295762003, 295763008, 295764002, 295765001, 295766000, 295767009, 295768004, 295769007, 295770008, 295771007, 295772000, 295773005, 295775003, 295776002, 295777006, 295778001, 295779009, 295780007, 295781006, 295782004, 295783009, 295784003, 295785002, 295786001, 295787005, 295788000, 295789008, 295790004, 295791000, 295792007, 295793002, 295794008, 295795009, 295796005, 295797001, 295798006, 295799003, 295800004, 295801000, 295802007, 295803002, 295804008, 295805009, 295806005, 295807001, 295808006, 295809003, 295810008, 295811007, 295812000, 295813005, 295814004, 295815003, 295816002,

295817006, 295818001, 295819009, 295820003, 295821004, 295822006, 295823001, 295824007, 295825008, 295826009, 295827000, 295828005, 295829002, 295830007, 295831006, 295832004, 295833009, 295834003, 295835002, 295836001, 295837005, 295838000, 295839008, 295840005, 295841009, 295842002, 295843007, 295844001, 295845000, 295846004, 295847008, 295848003, 295849006, 295851005, 295852003, 295853008, 295854002, 295855001, 295856000, 295857009, 295858004, 295859007, 295860002, 295861003, 295862005, 295863000, 295864006, 295865007, 295866008, 295867004, 295868009, 295869001, 295870000, 295871001, 295872008, 295873003, 295874009, 295875005, 295877002, 295878007, 295879004, 295880001, 295881002, 295882009, 295883004, 295884005, 295885006, 295886007, 295887003, 295888008, 295889000, 295890009, 295891008, 295892001, 295893006, 295894000, 295895004, 295896003, 295897007, 295898002, 295899005, 295900000, 295901001, 295902008, 295903003, 295904009, 295905005, 295906006, 295907002, 295908007, 295909004, 295910009, 295911008, 295912001, 295913006, 295914000, 295915004, 295916003, 295917007, 295918002, 295919005, 295920004, 295921000, 295922007, 295923002, 295924008, 295925009, 295926005, 295927001, 295928006, 295929003, 295930008, 295931007, 295932000, 295933005, 295934004, 295935003, 295936002, 295937006, 295938001, 295939009, 295940006, 295941005, 295942003, 295943008, 295944002, 295945001, 295946000, 295947009, 295948004, 295949007, 295950007, 295951006, 295952004, 295953009, 295954003, 295955002, 295956001, 295957005, 295958000, 295959008, 295960003, 295961004, 295962006, 295963001, 295964007, 295965008, 295966009, 295967000, 295968005, 295969002, 295970001, 295971002, 295972009, 295973004, 295974005, 295975006, 295976007, 295978008, 295979000, 295980002, 295981003, 295982005, 295983000, 295984006, 295985007, 295986008, 295987004, 295988009, 295989001, 295990005, 295991009, 295992002, 295993007, 295994001, 295995000, 295996004, 295997008, 295998003, 295999006, 296000008, 296001007, 296002000, 296003005, 296004004, 296005003, 296006002, 296007006, 296008001, 296009009, 296010004, 296011000, 296012007, 296013002, 296015009, 296016005, 296017001, 296018006, 296019003, 296020009, 296021008, 296022001, 296023006, 296024000, 296025004, 296026003, 296027007, 296028002, 296029005, 296030000, 296031001, 296032008, 296033003, 296034009, 296035005, 296036006, 296037002, 296038007, 296039004, 296040002, 296041003, 296042005, 296043000, 296044006, 296045007, 296046008, 296047004, 296048009, 296049001, 296050001, 296051002, 296052009, 296053004, 296054005, 296055006, 296056007, 296057003, 296058008, 296059000, 296060005, 296061009, 296062002, 296063007, 296064001, 296065000, 296066004, 296067008, 296068003, 296069006, 296070007, 296071006, 296072004, 296073009, 296074003, 296075002, 296076001, 296077005, 296079008, 296080006, 296081005, 296082003, 296083008, 296084002, 296085001, 296086000, 296087009, 296088004, 296089007, 296090003, 296091004, 296092006, 296093001, 296094007, 296095008, 296096009, 296097000, 296098005, 296099002, 296100005, 296101009, 296102002, 296103007, 296104001, 296105000, 296106004, 296107008, 296108003, 296109006, 296110001, 296111002, 296112009, 296113004, 296114005, 296115006, 296116007, 296117003, 296118008, 296119000, 296120006, 296121005, 296122003, 296123008, 296124002, 296125001, 296126000, 296127009, 296128004, 296129007, 296130002, 296131003, 296132005, 296133000, 296134006, 296135007, 296136008, 296137004, 296138009, 296139001, 296140004, 296141000, 296142007, 296143002, 296144008, 296145009, 296146005, 296147001, 296148006, 296149003, 296150003, 296151004, 296152006, 296153001, 296154007, 296155008, 296156009, 296157000, 296158005, 296159002, 296160007, 296161006, 296162004, 296163009, 296164003, 296165002, 296166001, 296167005, 296168000, 296169008, 296170009, 296171008, 296172001, 296173006, 296174000, 296175004, 296176003, 296177007, 296178002, 296180008, 296181007, 296182000, 296183005, 296184004, 296185003, 296186002, 296187006, 296188001, 296189009, 296190000, 296191001, 296192008, 296193003, 296194009, 296195005, 296196006, 296197002, 296198007, 296199004, 296200001, 296201002, 296202009, 296203004, 296204005, 296205006, 296206007, 296207003, 296208008, 296209000, 296210005, 296211009, 296212002, 296213007, 296214001, 296215000, 296216004, 296217008, 296218003, 296219006, 296220000, 296221001, 296222008, 296223003, 296224009, 296225005, 296226006, 296227002, 296228007, 296229004, 296230009, 296231008, 296232001, 296233006, 296234000, 296235004, 296236003,

296237007, 296238002, 296239005, 296240007, 296241006, 296242004, 296243009, 296244003, 296245002, 296246001, 296247005, 296248000, 296249008, 296250008, 296251007, 296252000, 296253005, 296258001, 296259009, 296260004, 296261000, 296262007, 296263002, 296264008, 296265009, 296270002, 296271003, 296272005, 296273000, 296274006, 296275007, 296276008, 296277004, 296278009, 296279001, 296281004, 296282006, 296283001, 296284007, 296285008, 296286009, 296287000, 296288005, 296289002, 296290006, 296291005, 296292003, 296293008, 296294002, 296300004, 296301000, 296302007, 296303002, 296304008, 296305009, 296306005, 296307001, 296308006, 296309003, 296310008, 296311007, 296312000, 296313005, 296314004, 296315003, 296316002, 296317006, 296321004, 296322006, 296323001, 296324007, 296325008, 296326009, 296327000, 296328005, 296329002, 296330007, 296331006, 296333009, 296334003, 296335002, 296336001, 296337005, 296338000, 296339008, 296340005, 296341009, 296342002, 296343007, 296344001, 296345000, 296346004, 296347008, 296348003, 296349006, 296350006, 296351005, 296352003, 296353008, 296354002, 296355001, 296356000, 296357009, 296358004, 296359007, 296360002, 296361003, 296362005, 296363000, 296364006, 296365007, 296366008, 296367004, 296368009, 296369001, 296370000, 296371001, 296372008, 296373003, 296374009, 296376006, 296377002, 296378007, 296379004, 296380001, 296381002, 296382009, 296384005, 296385006, 296386007, 296387003, 296389000, 296390009, 296391008, 296392001, 296393006, 296394000, 296395004, 296396003, 296397007, 296398002, 296399005, 296400003, 296401004, 296402006, 296403001, 296404007, 296405008, 296406009, 296407000, 296408005, 296409002, 296410007, 296411006, 296412004, 296413009, 296414003, 296415002, 296416001, 296417005, 296418000, 296419008, 296420002, 296421003, 296422005, 296423000, 296424006, 296425007, 296426008, 296427004, 296428009, 296429001, 296430006, 296431005, 296432003, 296433008, 296434002, 296435001, 296436000, 296437009, 296438004, 296439007, 296440009, 296441008, 296442001, 296443006, 296444000, 296445004, 296446003, 296447007, 296448002, 296449005, 296450005, 296451009, 296452002, 296453007, 296454001, 296455000, 296456004, 296457008, 296458003, 296459006, 296460001, 296461002, 296462009, 296463004, 296464005, 296465006, 296466007, 296467003, 296468008, 296469000, 296470004, 296471000, 296472007, 296473002, 296474008, 296475009, 296476005, 296477001, 296478006, 296479003, 296480000, 296481001, 296482008, 296483003, 296485005, 296486006, 296487002, 296488007, 296489004, 296490008, 296491007, 296492000, 296493005, 296494004, 296495003, 296496002, 296497006, 296498001, 296499009, 296500000, 296501001, 296502008, 296503003, 296504009, 296505005, 296506006, 296507002, 296509004, 296510009, 296511008, 296512001, 296513006, 296514000, 296515004, 296516003, 296517007, 296518002, 296519005, 296520004, 296521000, 296522007, 296523002, 296529003, 296530008, 296531007, 296532000, 296533005, 296534004, 296535003, 296536002, 296537006, 296538001, 296539009, 296540006, 296541005, 296542003, 296543008, 296544002, 296545001, 296546000, 296547009, 296548004, 296549007, 296550007, 296551006, 296552004, 296553009, 296554003, 296555002, 296556001, 296557005, 296558000, 296559008, 296560003, 296561004, 296562006, 296563001, 296564007, 296565008, 296566009, 296567000, 296568005, 296569002, 296570001, 296571002, 296572009, 296573004, 296574005, 296575006, 296576007, 296577003, 296578008, 296579000, 296580002, 296581003, 296582005, 296583000, 296584006, 296585007, 296587004, 296588009, 296589001, 296590005, 296591009, 296592002, 296593007, 296594001, 296595000, 296596004, 296597008, 296598003, 296599006, 296600009, 296601008, 296602001, 296603006, 296604000, 296605004, 296606003, 296607007, 296608002, 296609005, 296610000, 296611001, 296612008, 296613003, 296614009, 296615005, 296616006, 296617002, 296618007, 296619004, 296620005, 296621009, 296622002, 296623007, 296624001, 296625000, 296626004, 296627008, 296628003, 296629006, 296630001, 296631002, 296632009, 296633004, 296634005, 296635006, 296636007, 296637003, 296638008, 296639000, 296640003, 296641004, 296642006, 296643001, 296644007, 296645008, 296646009, 296647000, 296648005, 296649002, 296650002, 296651003, 296652005, 296653000, 296654006, 296655007, 296656008, 296657004, 296658009, 296659001, 296660006, 296661005, 296662003, 296663008, 296664002, 296665001, 296666000, 296667009, 296668004, 296669007, 296670008, 296671007, 296672000, 296673005, 296674004, 296675003, 296676002, 296677006, 296678001, 296679009,

296680007, 296681006, 296682004, 296683009, 296684003, 296685002, 296686001, 296688000, 296689008, 296690004, 296691000, 296692007, 296693002, 296694008, 296695009, 296696005, 296697001, 296698006, 296699003, 296700002, 296701003, 296702005, 296703000, 296704006, 296705007, 296706008, 296707004, 296708009, 296709001, 296710006, 296711005, 296712003, 296713008, 296714002, 296715001, 296716000, 296717009, 296718004, 296719007, 296720001, 296721002, 296722009, 296723004, 296724005, 296725006, 296726007, 296727003, 296728008, 296729000, 296730005, 296731009, 296732002, 296733007, 296734001, 296735000, 296737008, 296738003, 296739006, 296741007, 296742000, 296743005, 296744004, 296745003, 296746002, 296747006, 296748001, 296749009, 296750009, 296751008, 296752001, 296753006, 296754000, 296755004, 296756003, 296757007, 296758002, 296759005, 296760000, 296761001, 296762008, 296763003, 296764009, 296765005, 296766006, 296767002, 296768007, 296769004, 296770003, 296771004, 296772006, 296773001, 296774007, 296775008, 296776009, 296777000, 296779002, 296780004, 296781000, 296782007, 296783002, 296784008, 296785009, 296786005, 296787001, 296788006, 296789003, 296790007, 296792004, 296793009, 296794003, 296795002, 296796001, 296797005, 296798000, 296799008, 296800007, 296801006, 296802004, 296803009, 296804003, 296805002, 296806001, 296825003, 296826002, 296827006, 296828001, 296829009, 296830004, 296831000, 296832007, 296833002, 296834008, 296835009, 296836005, 296837001, 296838006, 296839003, 296840001, 296841002, 296842009, 296843004, 296844005, 296845006, 296846007, 296847003, 296848008, 296849000, 296850000, 296851001, 296852008, 296853003, 296854009, 296855005, 296856006, 296857002, 296858007, 296859004, 296860009, 296861008, 296862001, 296863006, 296864000, 296865004, 296866003, 296868002, 296869005, 296870006, 296871005, 296872003, 296873008, 296874002, 296875001, 296876000, 296877009, 296878004, 296879007, 296880005, 296881009, 296882002, 296883007, 296884001, 296885000, 296886004, 296887008, 296888003, 296889006, 296890002, 296891003, 296892005, 296894006, 296895007, 296896008, 296897004, 296898009, 296899001, 296900006, 296901005, 296902003, 296903008, 296904002, 296905001, 296906000, 296907009, 296908004, 296909007, 296910002, 296911003, 296912005, 296913000, 296914006, 296915007, 296916008, 296917004, 296919001, 296920007, 296921006, 296922004, 296923009, 296924003, 296925002, 296926001, 296927005, 296928000, 296929008, 296930003, 296931004, 296932006, 296933001, 296934007, 296935008, 296936009, 296938005, 296939002, 296940000, 296941001, 296942008, 296943003, 296944009, 296945005, 296946006, 296947002, 296948007, 296949004, 296950004, 296951000, 296952007, 296953002, 296954008, 296955009, 296956005, 296957001, 296958006, 296959003, 296960008, 296961007, 296962000, 296963005, 296964004, 296965003, 296966002, 296967006, 296968001, 296969009, 296970005, 296971009, 296972002, 296973007, 296974001, 296975000, 296976004, 296977008, 296978003, 296979006, 296980009, 296981008, 296982001, 296983006, 296984000, 296985004, 296986003, 296987007, 296988002, 296989005, 296990001, 296991002, 296993004, 296994005, 296995006, 296996007, 296997003, 296998008, 296999000, 297000002, 297001003, 297002005, 297003000, 297004006, 297005007, 297006008, 297007004, 297008009, 297009001, 297010006, 297011005, 297012003, 297013008, 297014002, 297015001, 297016000, 297017009, 297018004, 297019007, 297020001, 297021002, 297022009, 297023004, 297024005, 297025006, 297026007, 297027003, 297028008, 297029000, 297030005, 297031009, 297032002, 297033007, 297034001, 297035000, 297036004, 297037008, 297038003, 297039006, 297040008, 297041007, 297042000, 297043005, 297044004, 297045003, 297046002, 297047006, 297048001, 297049009, 297050009, 297051008, 297052001, 297053006, 297054000, 297055004, 297056003, 297057007, 297058002, 297059005, 297060000, 297061001, 297062008, 297063003, 297064009, 297065005, 297066006, 297067002, 297068007, 297070003, 297071004, 297072006, 297073001, 297074007, 297075008, 297076009, 297077000, 297078005, 297079002, 297080004, 297081000, 297082007, 297083002, 297084008, 297085009, 297086005, 297087001, 297088006, 297089003, 297090007, 297091006, 297092004, 297094003, 297095002, 297096001, 297097005, 297098000, 297099008, 297100000, 297101001, 297102008, 297199006, 297200009, 297201008, 297202001, 297259001, 297940005, 297941009, 297942002, 299969005, 300907002, 300992002, 300993007, 302507002, 302895007, 302897004, 1003473002, 1003530008, 1003532000, 1003534004, 1003535003,

1003842009, 1017197007, 111491000119107, 15999391000119104, 16000231000119105, 16000271000119108, 16077051000119102, 367411000119102, 876800006, 878878007, 880042006, 890086003, 890087007, 890088002, 890095006, 890112007, 890202007, 890232004, 897046009, 897590006, 1230072003, 1230071005, 1230266009, 1231328003, 1231159006, 1231163004, 1231161002

#### connective tissue disorder and autoimmune conditions

ICD-10-CM: M30, M31, M32, M33, M34, M35, M36, M35.06, M35.08, M35.05, M35.07, M35.0A, M31.19, M31.10, M35.0B, M31.11, M35.0C, M30.0, M30.1, M30.2, M30.3, M30.8, M31.0, M31.1, M31.2, M31.3, M31.30, M31.31, M31.4, M31.5, M31.6, M31.7, M31.8, M31.9, M32.0, M32.1, M32.10, M32.11, M32.12, M32.13, M32.14, M32.15, M32.19, M32.8, M32.9, M33.0, M33.00, M33.01, M33.02, M33.03, M33.09, M33.1, M33.10, M33.11, M33.12, M33.13, M33.19, M33.2, M33.20, M33.21, M33.22, M33.29, M33.9, M33.90, M33.91, M33.92, M33.93, M33.99, M34.0, M34.1, M34.2, M34.8, M34.81, M34.82, M34.83, M34.89, M34.9, M35.0, M35.00, M35.01, M35.02, M35.03, M35.04, M35.09, M35.1, M35.2, M35.3, M35.4, M35.5, M35.6, M35.7, M35.8, M35.9, M36.0, M36.1, M36.2, M36.3, M36.4, M36.8, M35.81, M35.89

SNOMED CT: 11791001, 81573002, 89155008, 105969002, 155441006, 55464009, 396230008, 1162336001, 1162292003, 1163001000, 11818331000119102, 1163088008, 1163087003, 1177028005, 11818291000119108, 11818851000119104, 11817691000119106, 11817731000119104, 11817771000119101, 11817811000119101, 11819171000119108, 11817261000119102, 11860221000119107, 11857901000119102, 11857741000119103, 11828261000119106, 1197749008, 1230239009, 1078201000119106, 1078191000119108, 300491000119106, 300651000119104, 1222678003, 1222677008, 1220589007, 1197751007, 1208889005, 16052071000119107, 16052031000119109, 1196856003, 1196841007, 1196883002, 1197296004, 1197259008, 310491000119108, 310501000119101, 310511000119103, 310521000119105, 310551000119102, 310561000119100, 310571000119106, 11777561000119105, 305961000119100, 1228876007, 1217372003, 1217667002, 1208830003, 1222679006, 1197747005, 349231000119108, 15992551000119106, 790731321000119100, 561258721000119104, 15675081000119107, 15701201000119107, 11860181000119104, 11833471000119105, 11860061000119109, 11860021000119104, 11819251000119106, 11833431000119107, 675451000119106, 11818691000119105, 11818731000119103, 11771091000119101, 1172606002, 16673981000119108, 11861881000119103, 11861961000119100, 1176990003, 1187507004, 1162361003, 1162354002, 1162352003, 1162353008, 1162335002, 1187546002, 1162319009, 11771051000119106, 16674081000119106, 11861841000119108, 11861921000119105, 11861721000119108, 1162301000, 1162303002, 11859981000119102, 11859941000119107, 11833551000119100, 11833511000119101, 11860141000119109, 11860101000119107, 11859821000119109, 11859781000119104, 11819291000119101, 11817381000119102, 11817341000119107, 11818491000119107, 11818531000119107, 11818571000119105, 11842411000119108, 11842331000119109, 11767691000119107, 11777961000119100, 11782001000119109, 11781961000119100, 11767651000119102, 11778841000119109, 11778801000119107, 11777921000119105, 11857621000119102, 11857581000119102, 11761051000119100, 1162342002, 11857941000119100, 11857781000119108, 1162302007, 11828341000119101, 1177035002, 1162362005, 15675321000119106, 701003, 815008, 961007, 984000, 1212005, 1282001, 1378000, 1418007, 1539003, 1763009, 2028007, 2304001, 2585002, 2955000, 3044008, 3144005, 3199001, 3558002, 3662000, 3903005, 4106009, 4168008, 4170004, 4237001, 4410001, 4676006, 5280003, 5313005, 5327000, 5494004, 5618007, 5965003, 6033008, 6154004, 6802007, 6825008, 6838000, 6849006, 7097001, 7102003, 7253006, 7548000, 8032007, 8549006, 8733006, 8976003, 9051008, 9771006, 10033001, 10535001, 10713006, 10728006, 11013005, 11338007, 11354005, 11480007, 11654001, 11666007, 11836002, 12227007, 12371008, 12519004, 13082000, 13370002, 13596001, 13771001, 13907006, 14136000, 14552009, 14949009, 15013002,

15511008, 15550007, 15788006, 15848008, 16751003, 17157001, 17234001, 17668000, 17883008, 18035009, 18484008, 18682003, 18988001, 19494006, 19564003, 19579005, 19682006, 19726003, 20021007, 20196005, 20926000, 21316006, 21333004, 21381006, 21565000, 22125009, 22352007, 22361007, 22817005, 22888007, 22949006, 23061007, 23415000, 23698009, 23777003, 23882000, 24005006, 24129002, 24302002, 24383005, 24730004, 24787008, 24864009, 25354003, 25380002, 25756004, 25817001, 26010008, 26077002, 26121002, 26538006, 26884006, 27182002, 27302009, 27741009, 27757009, 27810000, 27911000, 27970007, 28004004, 28122003, 28167005, 28358004, 28637003, 28845006, 29026008, 29210001, 29297002, 29583006, 29642000, 30477007, 30689000, 30832001, 31166000, 31249007, 31341008, 31474007, 31798004, 31810002, 31860008, 32093003, 32128001, 32141002, 32620007, 32861005, 32931001, 33132004, 33724004, 33760009, 33763006, 33882007, 33908005, 34038005, 34125004, 34129005, 34139004, 34439005, 1197745002, 81096007, 81543006, 81823008, 81902001, 82085002, 82112005, 82204006, 82275008, 82363001, 82632005, 82788007, 83071002, 83294006, 83836003, 83922009, 83923004, 84062004, 84089009, 84097002, 84288007, 84305006, 84857004, 84970007, 85216006, 85420008, 85426002, 85551004, 85991008, 86070006, 86128003, 86365006, 86378006, 86422009, 86595004, 86667008, 86709000, 86787000, 86907008, 87510000, 87778004, 87781009, 88075009, 88178009, 88773005, 88835002, 88906006, 89030000, 89573004, 89687005, 89730005, 89807005, 89988002, 90068007, 90073001, 90448008, 90450000, 90631008, 90654004, 91195006, 91240008, 91669008, 91945006, 92031000, 92046007, 92087008, 92269002, 92270001, 92277003, 92278008, 92284006, 92287004, 92298003, 92315002, 92316001, 92317005, 92330006, 92429000, 92432002, 92463003, 92483002, 92489003, 92490007, 92491006, 92492004, 92551005, 92674003, 92677005, 92685001, 92704005, 92781004, 92800006, 92844009, 92845005, 92846006, 92847002, 92875000, 93022005, 93038004, 93441003, 93442005, 93443000, 93444006, 93728003, 93742006, 93782004, 93942003, 93947009, 93948004, 93952004, 93955002, 93966009, 93981003, 93982005, 93983000, 93994001, 94089007, 94090003, 94120006, 94140004, 94226006, 94241006, 94282006, 94464005, 94465006, 94472007, 94473002, 94479003, 94482008, 94493005, 94510009, 94511008, 94512001, 94525009, 94624001, 94625000, 94626004, 94627008, 94658009, 94678001, 94768007, 94781000, 94822000, 94982001, 94987007, 94988002, 94992009, 94995006, 95006006, 95021000, 95022007, 95023002, 95034004, 95128009, 95129001, 95130006, 95160001, 95180000, 95336007, 95337003, 95350002, 95410001, 95414005, 95416007, 95434006, 95467005, 95468000, 95488001, 95550001, 95552009, 95554005, 95577005, 95609003, 95644001, 95735008, 95737000, 95740000, 95800001, 95802009, 95833006, 105611005, 109347009, 109348004, 109370001, 109372009, 109374005, 109375006, 109378008, 109382005, 109383000, 109384006, 109419009, 109464006, 109599007, 109600005, 109601009, 109630002, 109660007, 109661006, 109662004, 109753007, 109851002, 109852009, 109853004, 109854005, 109855006, 109856007, 109857003, 109858008, 109862002, 109863007, 109866004, 111026008, 111210001, 111222003, 111224002, 111368002, 111388003, 111653006, 111684008, 111807001, 118942006, 122864009, 123662000, 126487009, 126598008, 126648001, 126649009, 126671004, 126719004, 126720005, 126721009, 126722002, 126724001, 126733004, 126735006, 126865007, 126866008, 126867004, 126868009, 126869001, 126870000, 126871001, 126873003, 126876006, 126893006, 126917007, 126918002, 126919005, 127012008, 127178003, 127271004, 127292004, 127293009, 128346003, 128421002, 128460000, 128461001, 128463003, 128464009, 128465005, 128466006, 128467002, 129129003, 129451001, 129452008, 129586002, 1142039007, 1142092000, 1142205000, 1144805008, 1144809002, 1144897006, 1144899009, 1144903002, 1144905009, 1144920003, 1144925008, 1144931006, 1144937005, 1144938000, 1144941009, 1144944001, 1144967004, 1144969001, 1144972008, 1144977002, 1144982009, 1144990009, 1145203001, 1145238004, 1145241008, 1145242001, 1145243006, 1149232006, 1149233001, 1153347002, 1153372002, 1153393004, 1153394005, 1153396007, 1156387004, 1156390005, 1156391009, 1156392002, 1156814008, 11779001000119108, 11779041000119105, 16674031000119105,

682031000119109, 91191000119108, 91211000119109, 91221000119102, 11818771000119100, 1162355001, 11817491000119108, 11817531000119108, 11817571000119106, 11817611000119102, 11817301000119105, 11817421000119106, 1197746001, 786961001, 787094005, 787730008, 788010000, 788011001, 788012008, 788013003, 788529004, 788978004, 789006005, 789054002, 789394007, 789574002, 789694004, 789754007, 836274002, 836300009, 838354003, 840338002, 840426005, 840453009, 846637007, 860596009, 860597000, 860598005, 860801002, 860836003, 860890006, 860894002, 860923004, 866081005, 866103007, 866106004, 866176003, 870329005, 870330000, 870331001, 870332008, 870333003, 870335005, 870336006, 870393001, 870394007, 870573008, 870672007, 870767005, 870768000, 871492003, 871654009, 871843002, 331000119106, 1491000119102, 1781000119107, 1791000119105, 12121000119102, 13271000119101, 13291000119100, 15911000119103, 22531000119104, 23081000119102, 32561000119105, 36241000119108, 50411000119108, 72791000119108, 89681000119101, 91181000119105, 104951000119106, 107931000119105, 110471000119102, 120401000119101, 122691000119106, 135091000119106, 135141000119104, 137791000119100, 145751000119104, 187371000119108, 236871000119109, 286011000119101, 295101000119105, 295111000119108, 295121000119101, 296121000119105, 296131000119108, 296141000119104, 296151000119102, 296161000119100, 296171000119106, 296181000119109, 296191000119107, 296201000119105, 296211000119108, 296221000119101, 297181000119107, 297191000119105, 297201000119108, 297211000119106, 300161000119106, 300501000119104, 300671000119108, 301691000119102, 301701000119102, 302981000119100, 302991000119102, 303161000119101, 303171000119107, 303191000119108, 303211000119109, 303241000119108, 303261000119107, 303281000119103, 303841000119107, 303851000119109, 305661000119106, 305671000119100, 305701000119104, 305711000119101, 305831000119104, 305981000119109, 306001000119108, 306011000119106, 306031000119101, 306051000119107, 306071000119103, 306081000119100, 306101000119107, 306121000119103, 307001000119105, 307011000119108, 307371000119102, 307391000119101, 307451000119100, 307471000119109, 308761000119108, 308771000119102, 308941000119107, 308951000119109, 308961000119106, 308971000119100, 308981000119102, 308991000119104, 309001000119104, 309011000119101, 309021000119108, 309031000119106, 309041000119102, 310381000119102, 310391000119104, 311191000119109, 311211000119105, 311221000119103, 311231000119100, 311671000119100, 313521000119101, 313531000119103, 317511000119102, 317521000119109, 317741000119107, 317751000119109, 318171000119100, 318181000119102, 318771000119104, 318781000119101, 320021000119106, 320031000119109, 320661000119106, 320671000119100, 320681000119102, 321371000119100, 321451000119103, 321521000119105, 321601000119100, 322901000119100, 322961000119104, 323001000119107, 323011000119105, 328641000119109, 328661000119108, 331351000119107, 331371000119103, 331381000119100, 333651000119103, 334421000119106, 334701000119106, 335251000119100, 335411000119100, 336961000119101, 336981000119105, 336991000119108, 339251000119103, 340021000119108, 340301000119102, 340861000119109, 341021000119101, 343071000119104, 343091000119103, 343101000119108, 345661000119104, 346161000119102, 346291000119101, 347991000119105, 348021000119105, 348061000119100, 348071000119106, 348351000119100, 348431000119100, 348461000119108, 348511000119106, 348521000119104, 348801000119105, 349061000119107, 349081000119103, 349101000119105, 349111000119108, 349301000119108, 353661000119102, 353671000119108, 354621000119105, 367661000119102, 429671000124101, 431621000124100, 431731000124106, 432131000124100, 433721000124109, 433731000124107, 433741000124102, 434301000124102, 442241000124102, 448251000124102, 448581000124108, 448591000124106, 448601000124103, 448701000124105, 450231000124109, 450691000124105, 450701000124105, 457171000124104, 460561000124109, 675461000119108, 675471000119102, 678051000119107, 678071000119103, 678091000119102, 678471000119107, 678481000119105, 679751000119101, 679761000119104, 679771000119105, 1072931000119109, 1074061000119108, 1074071000119102, 1074081000119104, 1074091000119101, 1074101000119106, 1074111000119109, 1074121000119102, 1074131000119104, 1074141000119108, 1074151000119105, 1074161000119107, 1074171000119101, 1074181000119103, 1074191000119100, 1074201000119102, 1074211000119104, 1075391000119104, 1075401000119102, 1075411000119104, 1075421000119106, 1075431000119109, 1075441000119100, 1075451000119103, 1075461000119101, 1075791000119100, 1075801000119104,

1075811000119101, 1075821000119108, 1075831000119106, 1075841000119102, 1076111000119100, 1076121000119107, 1076131000119105, 1076141000119101, 1076471000119103, 1076581000119101, 1076591000119103, 1076621000119101, 1076631000119103, 1076641000119107, 1076651000119109, 1076671000119100, 1076691000119104, 1082921000119104, 1083711000119100, 1086071000119103, 1087021000119101, 1088031000119102, 1088841000119103, 1088861000119104, 1090651000119103, 1091451000119109, 1091471000119100, 1092381000119100, 1092831000119109, 2460001000004103, 5650001000004107, 6480001000004101, 8890001000004103, 10624571000119106, 10743311000119103, 10743351000119102, 10762111000119102, 10762151000119101, 10762191000119106, 10816301000119102, 10818101000119104, 10823451000119107, 10823531000119102, 10823571000119104, 10830741000119109, 10834461000119108, 10934521000119109, 10934561000119104, 10934601000119104, 10934641000119102, 10934681000119107, 10934721000119101, 10934761000119106, 10934801000119103, 11307481000119106, 11307521000119106, 11727381000119107, 11760661000119101, 11760701000119108, 11763011000119102, 11769171000119109, 11769211000119106, 11777091000119108, 11777171000119108, 11778001000119103, 11778041000119101, 11780401000119102, 11780441000119100, 11782121000119104, 11782161000119109, 11803011000119104, 11803051000119103, 11817451000119103, 11817651000119101, 11817851000119100, 11818451000119102, 11818651000119100, 11818891000119109, 11819011000119104, 11819051000119103, 11819091000119108, 11819131000119105, 11819211000119105, 11825671000119109, 11825711000119108, 11825911000119106, 11826031000119108, 11826631000119102, 11826711000119100, 11826791000119109, 11833271000119106, 11833311000119106, 11833591000119105, 11833631000119105, 11833671000119108, 11833711000119107, 11833751000119108, 11833791000119103, 11838121000119108, 11838161000119103, 11839311000119108, 11842091000119105, 11842131000119107, 11842171000119105, 11842211000119107, 11842251000119108, 11842291000119103, 11842371000119107, 11842451000119109, 11857341000119104, 11857501000119105, 11857541000119107, 11857661000119107, 11857701000119100, 11859541000119100, 11859581000119105, 11859661000119100, 11859701000119107, 11859741000119109, 11861761000119103, 11861801000119106, 11873791000119101, 11873911000119108, 12238731000119100, 12238771000119102, 12239791000119107, 12239831000119101, 12241111000119104, 12241151000119103, 12241191000119108, 12241231000119104, 12241271000119101, 12241311000119101, 12241511000119106, 12241551000119107, 12246841000119103, 12400001000004103, 14720231000119109, 15630571000119109, 15630611000119100, 15630651000119104, 15630691000119109, 15630731000119102, 15632011000119105, 15632051000119106, 15632091000119101, 15635841000119108, 15635881000119103, 15636211000119100, 15636291000119109, 15637151000119108, 15637191000119103, 15637231000119107, 15637271000119105, 15637351000119106, 15637391000119101, 15637431000119106, 15637471000119109, 15637511000119100, 15637551000119104, 15638411000119100, 15638851000119101, 15674641000119105, 15674681000119100, 15674721000119106, 15674761000119101, 15674881000119101, 15674921000119108, 15674961000119103, 15675001000119104, 15675041000119102, 15675121000119109, 15675201000119109, 15675281000119101, 15675361000119101, 15675401000119105, 15675441000119107, 15675481000119102, 15675521000119102, 15675561000119107, 15675641000119109, 15675721000119105, 15675761000119100, 15675801000119108, 15676001000119105, 15676161000119105, 15676241000119103, 15676281000119108, 15676321000119103, 15676401000119104, 15676441000119102, 15676481000119107, 15676521000119107, 15676561000119102, 15676601000119102, 15678441000119103, 15678481000119108, 15681801000119103, 15685201000119100, 15685281000119108, 15692081000119109, 15692121000119106, 15692161000119101, 15692601000119109, 15692641000119106, 15692681000119101, 15697601000119106, 15697641000119108, 15697681000119103, 15701081000119105, 15701121000119107, 15701161000119102, 15701961000119100, 15702081000119101, 15702121000119104, 15702161000119109, 15702281000119107, 15702361000119107, 15702401000119103, 15702441000119101, 15702921000119101, 15703241000119105, 15703281000119100, 15703321000119105, 15703361000119100, 15703401000119109, 15703441000119106, 15703481000119101, 15703521000119101, 15703561000119106,

15703641000119108, 15703681000119103, 15703721000119109, 15705201000119109, 15705241000119106, 15705281000119101, 15705361000119101, 15719641000119108, 15719681000119103, 15719721000119109, 15719761000119104, 15719841000119109, 15719921000119106, 15719961000119101, 15720001000119104, 15720041000119102, 15720161000119104, 15720481000119102, 15720561000119107, 15720601000119107, 15721401000119104, 15721521000119107, 15735041000119108, 15735081000119103, 15735121000119101, 15749161000119108, 15922191000119102, 15922231000119106, 15926411000119101, 15930181000119101, 15930221000119109, 15930261000119104, 15934341000119103, 15954261000119107, 15954301000119104, 15954341000119102, 15960021000119107, 15971701000119103, 15979711000119104, 15979751000119103, 15979791000119108, 15980431000119104, 15980471000119101, 15980511000119105, 15993551000119100, 16003591000119104, 16003671000119101, 16004071000119105, 16004111000119103, 16004151000119102, 16005631000119100, 16005711000119103, 16008111000119108, 16008191000119104, 16008271000119106, 16016151000119105, 16016191000119100, 16016431000119108, 16016511000119102, 16016551000119101, 16016631000119106, 16016991000119102, 16017031000119102, 16017071000119104, 16017151000119107, 16017431000119105, 16017471000119108, 16017591000119108, 16017631000119108, 16018591000119107, 16018671000119105, 16039071000119104, 16039111000119106, 16039151000119107, 16039191000119102, 16039231000119106, 16039311000119109, 16039391000119100, 16039431000119105, 16039631000119108, 16039791000119101, 16039831000119107, 16052111000119100, 16052151000119104, 16052191000119109, 16052231000119100, 16052271000119102, 16052351000119101, 16052431000119101, 16052471000119103, 16052551000119108, 16052591000119103, 16052631000119103, 16052671000119100, 16052711000119101, 16052791000119105, 16055631000119106, 16314671000119104, 16314711000119100, 16314751000119104, 16419701000119108, 16419741000119105, 16419781000119100, 16546631000119102, 16546791000119109, 16819501000119106, 16819541000119108, 16891571000119103, 16898361000119108, 16898401000119104, 29340001000004108, 674814021000119106, 5029005, 5801006, 7720002, 12579009, 17025000, 19346006, 20766005, 25606004, 29574001, 30128009, 30652003, 31848007, 45181002, 46041001, 50801008, 51696001, 53435002, 55711009, 56212008, 58588007, 62382002, 63247009, 67202007, 71322004, 76090006, 78946008, 79935000, 83470009, 83586000, 83901003, 84739008, 86804002, 87352006, 87442008, 91155009, 109816005, 123536004, 126766000, 196137000, 203045001, 203046000, 203047009, 203049007, 203050007, 203051006, 203052004, 203053009, 203054003, 203076007, 203395009, 203418008, 234035006, 234107009, 236782005, 239087008, 239912009, 239915006, 240554006, 274142002, 312946000, 402416000, 403634001, 428883008, 429513001, 446752000, 303861000119106, 322891000119104, 322911000119102, 322921000119109, 322951000119101, 322971000119105, 322981000119108, 15632131000119104, 15992511000119105, 15992591000119101, 16003631000119104, 444133002, 444158007, 444159004, 444163006, 444448004, 444468005, 444470001, 444578005, 444971006, 444972004, 444979008, 445117007, 445253000, 445276002, 445290004, 445332006, 445335008, 445496008, 445760005, 445947008, 446026002, 446076000, 446139000, 446251006, 446313009, 446600008, 447111007, 447169001, 447781009, 447783007, 447804006, 448139005, 448258005, 448274000, 448450001, 448497000, 448554008, 448618004, 448674007, 448903003, 449040002, 449067008, 449174000, 449186002, 449228007, 449259009, 449270002, 449293008, 449308006, 449377002, 449497006, 449824004, 449866003, 449900006, 451030007, 457667005, 457672001, 457677007, 457692008, 457697002, 459054000, 460602002, 460614005, 472979002, 472980004, 473373004, 473439008, 608829007, 609468001, 609473007, 609474001, 609495006, 609499000, 609501008, 609502001, 609505004, 609511001, 697923008, 697975004, 698072001, 698073006, 698294004, 698315000, 698522009, 698558002, 698694005, 698784005, 699235009, 699312008, 699326004, 699332009, 699419000, 699456009, 699655001, 699701005, 699702003, 699814001, 699953006, 699954000, 699961001, 699962008, 699963003, 699980004, 700002003, 700249006, 700250006, 700352001, 700353006, 700354000, 700376009, 700377000, 700380004, 700381000, 700385009, 700386005, 700444003, 702359002, 702567009, 702604009, 702605005, 702691009, 702718005, 703066001, 703542000, 703966004, 704028006, 704030008, 704093007, 704183003,

704237001, 704238006, 704239003, 704345008, 707301001, 707357005, 707430007, 707433009, 707435002, 707541006, 707551007, 707755000, 707756004, 708026002, 708964003, 709604005, 710211000, 711289002, 711379004, 711435001, 711638008, 711639000, 711640003, 711641004, 712722007, 712819000, 713038003, 713225000, 713692003, 713693008, 713720009, 713853007, 715057009, 715318006, 715401008, 715439000, 715465001, 715568002, 716649003, 716650003, 717824007, 717876006, 717920004, 718211004, 718342006, 719041000, 719069008, 719096006, 719213009, 719218000, 719296002, 719501003, 720858001, 720859009, 720860004, 720861000, 721271002, 721339007, 721340009, 721562005, 721568009, 721569001, 721580001, 721725005, 721973006, 722284009, 722434004, 722445001, 722514002, 722516000, 722519007, 722677008, 722813003, 722844007, 722856008, 722874004, 722893002, 723079001, 723081004, 723367005, 723829000, 723844004, 723845003, 723847006, 723849009, 723851008, 723852001, 723854000, 723855004, 723856003, 724146008, 724152009, 724176001, 724208006, 724454004, 724455003, 724457006, 724459009, 724502006, 724578006, 724580000, 724582008, 724584009, 724588007, 724589004, 724590008, 724592000, 724603009, 724604003, 724605002, 724767000, 724781003, 724782005, 724790005, 724791009, 724836008, 724841000, 724881007, 724882000, 724884004, 724885003, 724887006, 724888001, 724889009, 724890000, 724891001, 724892008, 724894009, 724895005, 724897002, 724898007, 724900009, 724901008, 724903006, 724904000, 724906003, 724907007, 724909005, 724910000, 724914009, 724915005, 724917002, 724919004, 724920005, 724922002, 724923007, 724925000, 724926004, 724928003, 724929006, 724931002, 724932009, 724933004, 724934005, 724936007, 724937003, 724941004, 724942006, 724951003, 724952005, 724954006, 724955007, 724957004, 724958009, 724966000, 724967009, 724969007, 724970008, 724972000, 724973005, 724975003, 724976002, 724978001, 724979009, 724981006, 724982004, 724984003, 724985002, 724998006, 725004007, 725005008, 725007000, 725008005, 725014003, 725015002, 725016001, 725035001, 725101002, 725104005, 725904009, 726191001, 726192008, 726194009, 726195005, 726197002, 726198007, 726200001, 726201002, 726203004, 726204005, 726206007, 726207003, 726217008, 726218003, 726220000, 726221001, 726223003, 726224009, 726226006, 726227002, 726229004, 726230009, 726232001, 726233006, 726323001, 726324007, 726326009, 726327000, 726329002, 726330007, 726332004, 726333009, 726628003, 726674004, 733093004, 733186000, 733219009, 733284005, 733286007, 733294000, 733416004, 733453005, 733457006, 733978001, 733980007, 733982004, 734173003, 735463000, 735592003, 735599007, 735602002, 735603007, 735604001, 735605000, 735606004, 735607008, 735608003, 735609006, 735610001, 735611002, 735678002, 735736000, 735766007, 735804004, 735807006, 735808001, 735817001, 735818006, 735819003, 735820009, 735821008, 735822001, 735823006, 735824000, 735825004, 735826003, 735827007, 735828002, 735829005, 735843000, 735844006, 735846008, 735856007, 735858008, 735888004, 735891004, 735892006, 735893001, 735903009, 735914007, 735923005, 737174002, 737181009, 737182002, 737183007, 737184001, 737253007, 737256004, 737257008, 737258003, 737259006, 737260001, 762303003, 762536003, 762674000, 763320005, 763839005, 766821006, 767815001, 767855004, 770361008, 770362001, 770760006, 770791000, 771306007, 772244003, 773276004, 773328002, 773333003, 773555005, 773644000, 773994002, 774080007, 774082004, 778022009, 778068007, 781239004, 781240002, 781241003, 781242005, 781387006, 781439002, 781440000, 781442008, 781444009, 781445005, 781446006, 781447002, 781448007, 781449004, 781455009, 781456005, 782538005, 782738008, 782780007, 782866008, 782937006, 783014007, 783182004, 783187005, 783253009, 783616005, 784349004, 784381008, 785808002, 785834005, 785835006, 163990004, 164538008, 186172004, 186182003, 186200004, 186225008, 186939000, 187801002, 187809000, 187810005, 187811009, 187814001, 187829004, 187843009, 187844003, 187845002, 187846001, 187853005, 187925006, 187987001, 187991006, 187992004, 187993009, 187994003, 187995002, 187996001, 187997005, 187999008, 188002005, 188009001, 188016000, 188017009, 188020001, 188021002, 188022009, 188204000, 188235006, 188445006, 188857008, 188861002, 188862009, 188863004, 188868008, 188877001, 188878006, 188880000, 188881001, 188882008, 188934003, 189120005, 189123007, 189252004, 189253009, 189255002, 189256001, 189257005, 189399009, 189401003, 189410006, 189412003, 189414002, 189415001, 189416000, 192658007, 193119005, 194151003, 194156008,

195495009, 195896004, 196028003, 196051003, 196052005, 196053000, 196075003, 196076002, 196077006, 196079009, 196080007, 196081006, 196115007, 196116008, 196133001, 196138005, 196151000, 196152007, 196153002, 196333005, 196602002, 196781001, 196880006, 196885001, 196890003, 196923005, 196933002, 196950000, 197163002, 197171003, 197172005, 197185008, 197200005, 197201009, 197742006, 197743001, 197744007, 197745008, 197747000, 197819009, 197834003, 198066009, 198156004, 198159006, 198185005, 198251001, 198626009, 200190007, 200191006, 200936003, 201436003, 201443009, 201625003, 201627006, 201630004, 201631000, 201633002, 201634008, 201637001, 201639003, 201642009, 201643004, 201645006, 201646007, 202078001, 202079009, 202080007, 202081006, 202082004, 202083009, 202089008, 202093002, 202094008, 202095009, 202096005, 202097001, 202098006, 202099003, 202102003, 202108004, 202109007, 202110002, 202111003, 202112005, 202115007, 202116008, 202117004, 202118009, 202119001, 202120007, 202121006, 202122004, 202123009, 202124003, 202131004, 202133001, 202137000, 202138005, 202142008, 202144009, 202328006, 202333005, 202334004, 202335003, 202336002, 202707000, 202708005, 202728009, 202729001, 202730006, 202731005, 202732003, 202733008, 202734002, 202735001, 202736000, 202744000, 202745004, 202746003, 202747007, 202748002, 202749005, 202750005, 202751009, 202752002, 202753007, 202754001, 202755000, 202756004, 202757008, 202770004, 202771000, 202841003, 202856007, 202857003, 202863007, 202868003, 202872004, 202873009, 202880006, 202881005, 202882003, 202888004, 202889007, 202961008, 202962001, 202963006, 202964000, 202968002, 202971005, 202973008, 202974002, 202975001, 202976000, 202977009, 202986004, 202987008, 202988003, 202989006, 202990002, 202991003, 202992005, 203003003, 203368004, 203382004, 203521008, 204171005, 204398004, 204537008, 204558002, 204589003, 204826004, 204905003, 205016000, 205093009, 205111008, 205274004, 205480005, 205481009, 206254003, 206539008, 208122008, 208131008, 208146001, 208147005, 208155003, 208156002, 208443000, 208459000, 208476001, 208492006, 208915004, 208916003, 208917007, 208918002, 208919005, 208920004, 208921000, 208922007, 208923002, 208924008, 208925009, 208926005, 208927001, 208976007, 209120006, 209130002, 304576008, 306949002, 306950002, 306951003, 306953000, 307139007, 307423008, 307499004, 307500008, 307601000, 307731004, 307755009, 307945003, 308143008, 308154003, 308232009, 308849005, 308887002, 309744003, 309752000, 309753005, 309762007, 309780003, 309781004, 310640003, 311804006, 312225001, 312696001, 312697005, 312698000, 312847008, 312901001, 312910009, 313294001, 314042000, 314043005, 314049009, 314545009, 314546005, 314549003, 314665009, 314666005, 314883003, 360437006, 360489001, 361199007, 362992004, 363039000, 363044007, 363045008, 363055007, 363091000, 363160008, 363164004, 363173007, 363182001, 363370001, 363421004, 363431006, 363433009, 363456000, 363492001, 367356000, 369535006, 369536007, 369537003, 369538008, 369539000, 369540003, 370469003, 370484007, 370493008, 370504007, 370526008, 372015000, 372016004, 373430008, 373439009, 384710005, 385424001, 386032009, 387665005, 389264005, 389268008, 389271000, 389272007, 390909000, 16051871000119105, 209216004, 209226006, 209412004, 209413009, 209414003, 209415002, 209416001, 209417005, 209418000, 209420002, 209428009, 209429001, 209436000, 209439007, 209440009, 209441008, 209442001, 209444000, 209445004, 209446003, 209447007, 209448002, 209450005, 209451009, 209452002, 209453007, 209454001, 209455000, 209456004, 209457008, 209458003, 209459006, 209463004, 209466007, 209469000, 209470004, 209471000, 209472007, 209473002, 209474008, 209476005, 209479003, 209480000, 209481001, 209482008, 209483003, 209484009, 209485005, 209486006, 209487002, 209488007, 209489004, 209490008, 209491007, 209492000, 209506006, 209507002, 209508007, 209509004, 209517007, 209519005, 209520004, 209521000, 209522007, 209529003, 209535003, 209536002, 209540006, 209542003, 209547009, 209548004, 209549007, 209550007, 209552004, 209553009, 209557005, 209559008, 209560003, 209561004, 209565008, 209566009, 209575006, 209576007, 209577003, 209579000, 209580002, 209581003, 209583000, 209584006, 209585007, 209586008, 209587004, 209588009, 209589001, 209590005, 209591009, 209592002, 209593007, 209594001, 209595000, 209596004, 209597008, 209598003, 209600009, 209601008, 209602001, 209603006, 209604000, 209606003, 209607007, 209608002, 209609005, 209610000, 209612008, 209613003, 209614009, 209615005, 209616006,

209617002, 209618007, 209621009, 209622002, 209625000, 209626004, 209627008, 209628003, 209629006, 209632009, 209633004, 209634005, 209635006, 209637003, 209638008, 209639000, 209640003, 209641004, 209645008, 209646009, 209647000, 209648005, 209649002, 209651003, 209652005, 209653000, 209654006, 209657004, 209658009, 209659001, 209661005, 209662003, 209663008, 209668004, 209669007, 209670008, 209671007, 209674004, 209675003, 209676002, 209677006, 209678001, 209680007, 209681006, 209682004, 209683009, 209684003, 209685002, 209686001, 209689008, 209690004, 209693002, 209694008, 209695009, 209696005, 209697001, 209698006, 209701003, 209702005, 209703000, 209704006, 209705007, 209706008, 209708009, 209709001, 209710006, 209711005, 209712003, 209715001, 209716000, 209717009, 209718004, 209719007, 209720001, 209721002, 209723004, 209725006, 209729000, 209731009, 209732002, 209735000, 209736004, 209738003, 209739006, 209740008, 209741007, 209743005, 209744004, 209745003, 209747006, 209748001, 209749009, 209750009, 209753006, 209754000, 209755004, 209757007, 209761001, 209762008, 209763003, 209765005, 209767002, 209768007, 209770003, 209771004, 209772006, 209774007, 209775008, 209777000, 209779002, 209783002, 209784008, 209785009, 209793009, 209806001, 209808000, 209810003, 209812006, 209813001, 209814007, 209815008, 209816009, 209817000, 209818005, 210088000, 210094008, 210262008, 210521001, 210511009, 210512002, 210513007, 210517008, 210518003, 210520000, 210543009, 210547005, 210548000, 210549008, 210550008, 210566005, 210567001, 210568006, 210569003, 210570002, 210582006, 210583001, 210585008, 210586009, 210587000, 210588005, 210590006, 210591005, 210592003, 210594002, 210595001, 210596000, 210669008, 210670009, 210671008, 210672001, 210673006, 210674000, 210693003, 210695005, 210696006, 210697002, 210698007, 210715002, 210716001, 210717005, 210718000, 210719008, 210730006, 212399002, 212421009, 212451003, 212452005, 212453000, 212454006, 212455007, 212456008, 212457004, 212458009, 212459001, 212462003, 212464002, 212467009, 212468004, 212469007, 212470008, 212471007, 212472000, 212473005, 212475003, 212476002, 212477006, 213073009, 213075002, 213196006, 213293008, 213294002, 213386007, 230733004, 230734005, 231275009, 231872000, 231873005, 231907006, 231909009, 231931001, 231933003, 231934009, 232016005, 232017001, 232018006, 232062002, 232063007, 232064001, 232065000, 232076001, 232077005, 232366008, 232369001, 232381002, 232439007, 232460001, 233621003, 233623000, 233636000, 233637009, 233638004, 233651009, 233653007, 233655000, 233656004, 233657008, 233658003, 233659006, 233703007, 233713004, 233717003, 233788001, 233798007, 234054006, 234102003, 234108004, 234505006, 234988003, 235000001, 235006007, 235700007, 235795007, 235848001, 235983003, 235985005, 235986006, 235987002, 235988007, 235989004, 235990008, 236001005, 236008004, 236009007, 236012005, 236013000, 236014006, 236018009, 236019001, 236026001, 236028000, 236036009, 236042008, 236043003, 236053002, 236054008, 236055009, 236058006, 236302005, 236374007, 236451007, 236452000, 236453005, 236454004, 236455003, 236456002, 236457006, 236458001, 236466005, 236502006, 236503001, 236523000, 236559006, 236689006, 236775006, 237045001, 237062006, 237063001, 237064007, 237065008, 237066009, 237327008, 237608006, 237610008, 238399000, 238652000, 238675007, 238676008, 238821003, 238822005, 238850005, 238877001, 238878006, 238881001, 238882008, 238883003, 238884009, 238886006, 238887002, 238889004, 238890008, 238893005, 238894004, 238895003, 238896002, 238897006, 238898001, 238899009, 238900004, 238901000, 238903002, 238923001, 238935002, 238936001, 239094006, 239142006, 239720000, 239721001, 239723003, 239724009, 239725005, 239726006, 239727002, 239728007, 239729004, 239730009, 239731008, 239833001, 239838005, 239839002, 239840000, 239841001, 239885004, 239886003, 239887007, 239888002, 239889005, 239890001, 239901009, 239918008, 239924002, 239925001, 239926000, 239927009, 239928004, 239929007, 239930002, 239931003, 239938009, 239943002, 239949003, 239950003, 239956009, 239959002, 239962004, 239963009, 239967005, 239969008, 239970009, 239977007, 239978002, 239979005, 239980008, 239981007, 239982000, 239984004, 239985003, 239986002, 239987006, 239995005, 239996006, 240001002, 240005006, 240006007, 240009000, 240014001, 240015000, 240019006, 240020000, 240026006, 240029004, 240030009, 240037007, 240039005, 240040007, 240044003, 240134003, 240137005, 240138000, 240139008, 240140005, 240177002, 240178007, 240179004, 240180001,

240181002, 240182009, 240211000, 240212007, 240213002, 240215009, 240216005, 240261009, 240262002, 240263007, 240264001, 240265000, 240303007, 240360007, 240629003, 240657005, 246956006, 246960009, 247081001, 247182006, 247184007, 247186009, 247187000, 248315005, 251002009, 252246005, 253236002, 253376005, 253377001, 253398001, 253435000, 253461001, 253490007, 253586007, 253601007, 253733006, 253737007, 253742004, 253743009, 253748000, 253802003, 253826001, 253899000, 34840004, 34964002, 35037009, 35423006, 35726004, 35852003, 35929003, 36046008, 36071006, 36139001, 36402006, 36427004, 36746002, 36948007, 37061001, 37600001, 37632001, 37785001, 37830006, 37925008, 38086007, 38540001, 38877003, 39642007, 39662004, 39838007, 40102009, 40527005, 41022003, 41352000, 41733009, 41806003, 42290006, 42536002, 42666000, 42711005, 42786005, 42944009, 43132002, 43606009, 43633008, 43663004, 43898003, 44020005, 44274007, 44465007, 44568006, 44572005, 44606004, 44636008, 44758003, 44814008, 44957002, 44983007, 45093008, 45283008, 45365002, 45431004, 45715008, 45815001, 45855004, 45861001, 45927004, 45987002, 46365005, 46626002, 46722007, 46860007, 46960006, 46970008, 47028006, 47123000, 47330001, 47977000, 48012003, 48142003, 48153009, 48274004, 48549003, 48661000, 48732008, 48819008, 49001006, 49292002, 49303001, 49388007, 49781004, 49922003, 49923008, 50127006, 50844007, 50869007, 50927007, 50986000, 51036000, 51108008, 51877002, 51953009, 51954003, 52042003, 52111006, 52159006, 52486002, 52533003, 52890006, 53200002, 53208009, 53417006, 53556002, 53740009, 53772007, 54072008, 54138003, 54176009, 54203008, 54615002, 54879000, 54888009, 54912002, 54942004, 55031000, 55069005, 55275006, 55430005, 55898008, 56391002, 56451001, 56458007, 56472003, 56580008, 56623003, 57100005, 57201002, 57203004, 57341009, 57602001, 57658009, 57921006, 58144006, 58392004, 58474006, 58710009, 58781003, 59252009, 59399004, 59400006, 59451000, 59523007, 59658000, 59818004, 60031001, 60033003, 60058008, 60125001, 60189009, 60258001, 60485005, 60490008, 60492000, 60507002, 60627008, 60644007, 60676002, 60805002, 60926001, 60937000, 61456005, 61458006, 61951009, 62100001, 62332007, 62394006, 62557001, 62853008, 63198006, 63637002, 63885004, 63977007, 64109004, 64252005, 64286001, 64298006, 64667001, 64787009, 64954002, 64994000, 65323003, 65372005, 66259004, 66579008, 66722004, 66885008, 66993009, 67504007, 67602004, 67741005, 67782005, 67988000, 68172002, 68189005, 68675004, 68815009, 69186005, 69195002, 69688007, 69880002, 70190001, 70218004, 70558001, 70610001, 70660007, 70690000, 70704007, 70736000, 70783003, 71216006, 71325002, 71953008, 72275000, 72391004, 72634002, 72735007, 72744008, 72888009, 72898003, 73147001, 73180008, 73198007, 73286009, 73589001, 73725006, 73856006, 73873008, 74204001, 74383007, 74610006, 75037000, 75128002, 75204008, 75467001, 75659004, 75680005, 75759008, 75965001, 76097009, 76318008, 76472002, 76502001, 76521009, 76630008, 76682005, 76805007, 77138008, 77360001, 77522006, 77547008, 77609009, 77690003, 77753005, 77860008, 77881008, 78187008, 78223000, 78609007, 78675000, 78815005, 78849002, 78887005, 79158003, 79362003, 79534009, 79977000, 80258006, 80475003, 80654001, 1197705006, 11777521000119100, 397564002, 397565001, 397568004, 397570008, 397571007, 397768002, 397856003, 398021003, 398049005, 398114001, 398147005, 398316009, 398318005, 398320008, 398733004, 398878007, 399259009, 399346004, 399980009, 400138001, 402425006, 402655006, 402656007, 402657003, 402659000, 402660005, 402661009, 402662002, 402663007, 402684002, 402686000, 402687009, 402688004, 402689007, 402690003, 402691004, 402692006, 402711009, 402712002, 402782006, 402801005, 402855009, 402858006, 402859003, 402926000, 402958005, 402968000, 402969008, 402970009, 403189002, 403397004, 403398009, 403399001, 403400008, 403401007, 403404004, 403414008, 403415009, 403416005, 403417001, 403418006, 403419003, 403420009, 403421008, 403422001, 403424000, 403486000, 403487009, 403488004, 403510002, 403511003, 403518009, 403621002, 403635000, 403652001, 403657007, 403661001, 403662008, 403670003, 403672006, 403735006, 403811007, 403812000, 403813005, 403814004, 403988009, 403989001, 403990005, 404062002, 404063007, 404064001, 404066004, 404224009,

404225005, 405275001, 405549001, 405561009, 410016009, 410017000, 410074002, 410499008, 410577002, 410812005, 410813000, 413918007, 413994008, 414872006, 415574006, 415575007, 415652006, 415655008, 415737009, 415746003, 415748002, 415749005, 416050006, 416236009, 417183007, 417544004, 417939004, 418054005, 418252000, 418254004, 418435001, 418605004, 418839003, 418961007, 419039007, 419051009, 419074008, 419197009, 419378004, 419395007, 419648001, 419900000, 419982005, 420000008, 420571002, 421866006, 422061002, 422116004, 422216007, 422217003, 422947009, 423093000, 423226005, 423248004, 423417009, 423486005, 423810002, 423849004, 424010006, 424188009, 424358006, 424578002, 425772008, 425878001, 426218005, 426394008, 426437004, 426570007, 427123006, 427300002, 427683007, 427883005, 428029006, 428189005, 428228008, 428255004, 428705008, 428778006, 428861009, 429052003, 429360005, 429447000, 429632003, 429718008, 429766001, 430206002, 430377007, 430505004, 430506003, 430892004, 430938004, 431033003, 431500000, 431873008, 433162006, 438479005, 438495005, 438582003, 439052006, 439108000, 439990003, 440018001, 441737006, 441885003, 441886002, 441963008, 442009005, 442110002, 442520000, 442528007, 442884002, 442925003, 442928001, 442941002, 442942009, 443001001, 443002008, 443120002, 443142001, 443700006, 443872005, 443943005, 254044004, 254140006, 254142003, 254144002, 254220005, 254221009, 254222002, 254223007, 254522003, 254528004, 254529007, 254645002, 254646001, 254737002, 254738007, 254824006, 254825007, 254831005, 255067005, 255068000, 262823004, 262840008, 262849009, 262854000, 262872006, 262882007, 262883002, 262884008, 262885009, 262886005, 262887001, 262888006, 262889003, 262925007, 262971000, 262972007, 262975009, 262976005, 262977001, 262979003, 262981001, 262982008, 262983003, 262985005, 262986006, 262987002, 262988007, 262989004, 262990008, 262995003, 263121007, 263123005, 263125003, 263126002, 263128001, 263129009, 263130004, 263133002, 263134008, 263135009, 263136005, 263137001, 263139003, 263142009, 263143004, 263144005, 263145006, 263146007, 263257008, 263258003, 263259006, 263260001, 263275009, 264506002, 264507006, 266370006, 266564004, 267659002, 267974002, 268002004, 268022000, 268023005, 268024004, 268025003, 268100009, 268219005, 268274005, 269113006, 269134004, 269138001, 269142003, 269181003, 271018002, 271467005, 271977000, 271979002, 274137005, 274140005, 274141009, 274162005, 274174009, 274175005, 275326005, 275327001, 275330008, 275332000, 275334004, 275335003, 275336002, 275500001, 275501002, 275503004, 276421009, 276422002, 276423007, 276424001, 276425000, 276427008, 276428003, 276429006, 276430001, 276431002, 276480007, 276481006, 276522000, 276657008, 276883000, 277217005, 277218000, 277678003, 277782009, 277783004, 277844007, 278486006, 280116003, 280483007, 281358000, 281363001, 281371002, 281428005, 281429002, 281430007, 281433009, 281434003, 281464006, 281524009, 281537007, 281538002, 281539005, 281540007, 281542004, 281543009, 281544003, 281545002, 281546001, 281547005, 281549008, 281550008, 281551007, 281552000, 281553005, 281554004, 281555003, 281598004, 281599007, 281602002, 284449005, 285605009, 285616002, 285654002, 285808008, 285834000, 286905003, 287062005, 287093006, 287094000, 287980003, 288215009, 288216005, 288217001, 288218006, 289260002, 295723007, 297940005, 298016002, 298017006, 298285004, 299276009, 299683005, 299684004, 300947009, 301005000, 301453009, 301769001, 302913000, 302932006, 302933001, 302935008, 1003407000, 1003427004, 1003431005, 1003432003, 1003437009, 1003438004, 1003440009, 1010664005, 1010666007, 1010668008, 1017196003, 1017197007, 1052298001, 1052299009, 1083291000119100, 1083731000119105, 1087291000119106, 1088421000119107, 1091031000119106, 1119266006, 15675961000119102, 15959741000119102, 15959781000119107, 15965661000119103, 15965701000119105, 15965741000119107, 15965781000119102, 15971661000119105, 16020471000119100, 305991000119107, 306021000119104, 306041000119105, 306061000119109, 306091000119102, 306111000119105, 880084003, 880085002, 880088000, 880089008, 890216002, 890415000, 895215000, 897296001, 897297005, 897721003, 926335004, 15720201000119109, 15720281000119101, 16039471000119108, 16039511000119104, 300531000119106, 300571000119109, 300681000119106, 300731000119102, 1217320008, 1217670003, 15675681000119104, 1208910001, 15675601000119107, 15675921000119107, 16039591000119108, 16039671000119106, 1231448006, 1231447001, 16052391000119106, 16052311000119102, 15703601000119106,

16052751000119100, 544102301000119103

ICD-9-CM: 446, 710, 710.0, 710.1, 710.3

#### Pulmonary hypertension

ICD-9-CM: 416.0, 416.8

ICD-10-CM: I27.0, I27.2, I27.20, I27.21, I27.22, I27.23, I27.24, I27.29

SNOMED CT: 70995007, 5499009, 10964002, 11399002, 87394009, 88223008, 89420002, 840306007, 840307003, 840308008, 233948000, 697899000, 697900005, 445237003, 445928005, 472757001, 472758006, 472759003, 472760008, 472761007, 472790001, 697896007, 697897003, 697898008, 697901009, 697902002, 697903007, 697904001, 697905000, 697906004, 697907008, 697908003, 697909006, 697910001, 697911002, 697912009, 697913004, 697914005, 697915006, 697916007, 697917003, 697918008, 697919000, 697920006, 697921005, 697922003, 697923008, 697924002, 697925001, 697926000, 697927009, 697928004, 703355003, 720575002, 776416004, 360573001, 360578005, 233815004, 233941006, 233942004, 233943009, 233944003, 233945002, 233946001, 233947005, 233949008, 233950008, 233954004, 234161007, 276792008, 276793003, 276794009, 1010625007, 1010627004, 879814007, 879815008, 879816009

#### Pregestational diabetes

ICD-10-CM: E11.610, E10.10, E10.11, E10.21, E10.22, E10.29, E10.311, E10.319, E10.321, E10.3211, E10.3212, E10.3213, E10.3219, E10.329, E10.3291, E10.3292, E10.3293, E10.3299, E10.331, E10.3311, E10.3312, E10.3313, E10.3319, E10.339, E10.3391, E10.3392, E10.3393, E10.3399, E10.341, E10.3411, E10.3412, E10.3413, E10.3419, E10.349, E10.3491, E10.3492, E10.3493, E10.3499, E10.351, E10.3511, E10.3512, E10.3513, E10.3519, E10.3521, E10.3522, E10.3523, E10.3529, E10.3531, E10.3532, E10.3533, E10.3539, E10.3541, E10.3542, E10.3543, E10.3549, E10.3551, E10.3552, E10.3553, E10.3559, E10.359, E10.3591, E10.3592, E10.3593, E10.3599, E10.36, E10.37X1, E10.37X2, E10.37X3, E10.37X9, E10.39, E10.40, E10.41, E10.42, E10.43, E10.44, E10.49, E10.51, E10.52, E10.59, E10.610, E10.618, E10.620, E10.621, E10.622, E10.628, E10.630, E10.638, E10.641, E10.649, E10.65, E10.69, E10.8, E10.9, E11.00, E11.01, E11.10, E11.11, E11.21, E11.22, E11.29, E11.311, E11.319, E11.321, E11.3211, E11.3212, E11.3213, E11.3219, E11.329, E11.3291, E11.3292, E11.3293, E11.3299, E11.331, E11.3311, E11.3312, E11.3313, E11.3319, E11.339, E11.3391, E11.3392, E11.3393, E11.3399, E11.341, E11.3411, E11.3412, E11.3413, E11.3419, E11.349, E11.3491, E11.3492, E11.3493, E11.3499, E11.351, E11.3511, E11.3512, E11.3513, E11.3519, E11.3521, E11.3522, E11.3523, E11.3529, E11.3531, E11.3532, E11.3533, E11.3539, E11.3541, E11.3542, E11.3543, E11.3549, E11.3551, E11.3552, E11.3553, E11.3559, E11.359, E11.3591, E11.3592, E11.3593, E11.3599, E11.36, E11.37X1, E11.37X2, E11.37X3, E11.37X9, E11.39, E11.40, E11.41, E11.42, E11.43, E11.44, E11.49, E11.51, E11.52, E11.59, E11.618, E11.620, E11.621, E11.622, E11.628, E11.630, E11.638, E11.641, E11.649, E11.65, E11.69, E11.8, E11.9, E13.00, E13.01, E13.10, E13.11, E13.21, E13.22, E13.29, E13.311, E13.319, E13.321, E13.3211, E13.3212, E13.3213, E13.3219, E13.329, E13.3291, E13.3292, E13.3293, E13.3299, E13.331, E13.3311, E13.3312, E13.3313, E13.3319, E13.339, E13.3391, E13.3392, E13.3393, E13.3399, E13.341, E13.3411, E13.3412, E13.3413, E13.3419, E13.349, E13.3491, E13.3492, E13.3493, E13.3499, E13.351, E13.3511, E13.3512, E13.3513, E13.3519, E13.3521, E13.3522, E13.3523, E13.3529, E13.3531, E13.3532, E13.3533, E13.3539, E13.3541, E13.3542, E13.3543, E13.3549, E13.3551, E13.3552, E13.3553, E13.3559, E13.359, E13.3591,

E13.3592, E13.3593, E13.3599, E13.36, E13.37X1, E13.37X2, E13.37X3, E13.37X9, E13.39, E13.40, E13.41, E13.42, E13.43, E13.44, E13.49, E13.51, E13.52, E13.59, E13.610, E13.618, E13.620, E13.621, E13.622, E13.628, E13.630, E13.638, E13.641, E13.649, E13.65, E13.69, E13.8, E13.9, O24.011, O24.012, O24.013, O24.019, O24.02, O24.03, O24.111, O24.112, O24.113, O24.119, O24.12, O24.13, O24.311, O24.312, O24.313, O24.319, O24.32, O24.33, O24.811, O24.812, O24.813, O24.819, O24.82, O24.83

ICD-9-CM: 250.00, 250.01, 250.02, 250.03, 250.10, 250.11, 250.12, 250.13, 250.20, 250.21, 250.22, 250.23, 250.30, 250.31, 250.32, 250.33, 250.40, 250.41, 250.42, 250.43, 250.50, 250.51, 250.52, 250.53, 250.60, 250.61, 250.62, 250.63, 250.70, 250.71, 250.72, 250.73, 250.80, 250.81, 250.82, 250.83, 250.90, 250.91, 250.92, 250.93, 357.2, 362.01, 362.02, 362.03, 362.04, 362.05, 362.06, 362.07, 366.41

SNOMED CT: 9859006, 11530004, 23045005, 28032008, 81531005, 111552007, 127013003, 691000119103, 731000119105, 1481000119100, 1501000119109, 1511000119107, 1551000119108, 28331000119107, 31211000119101, 31321000119102, 41911000119107, 60951000119105, 60961000119107, 60971000119101, 60991000119100, 71441000119104, 71721000119101, 82541000119100, 82551000119103, 82571000119107, 82581000119105, 97331000119101, 102781000119107, 104941000119109, 104961000119108, 109171000119104, 110181000119105, 138881000119106, 138891000119109, 138901000119108, 138911000119106, 138921000119104, 138941000119105, 368101000119109, 368521000119107, 368581000119106, 46635009, 443694000, 770098001, 71791000119104, 87921000119104, 444073006, 609562003, 609564002, 609566000, 609567009, 712882000, 712883005, 713702000, 713703005, 713705003, 713706002, 719216001, 739681000, 190330002, 190331003, 190368000, 190372001, 190389009, 199229001, 199230006, 313435000, 313436004, 314893005, 314902007, 314903002, 359642000, 237599002, 237604008, 44054006, 420279001, 420436000, 420486006, 420789003, 420918009, 421075007, 421326000, 421365002, 421437000, 421468001, 421779007, 421847006, 421893009, 422034002, 422099009, 422166005, 427027005, 427571000, 428007007, 1217044000, 1217068008, 789542009, 792926007, 711000119100, 721000119107, 741000119101, 751000119104, 771000119108, 781000119106, 1491000119102, 18521000119106, 60981000119103, 71421000119105, 71701000119105, 71771000119100, 90721000119101, 90731000119103, 90741000119107, 90751000119109, 90761000119106, 90771000119100, 90781000119102, 90791000119104, 96441000119101, 97341000119105, 104951000119106, 126551000119103, 127991000119101, 128001000119105, 140101000119109, 140111000119107, 140121000119100, 140131000119102, 157141000119108, 243421000119104, 368591000119109, 420756003, 421920002, 769219006, 769220000, 445170001, 445353002, 703137001, 703138006, 706894000, 707221002, 722454003, 723074006, 197605007, 309426007, 310387003, 311366001, 236499007, 236500003, 237627000, 38046004, 63510008, 420514000, 420715001, 421305000, 421986006, 425455002, 426875007, 290002008, 1003605004, 16697421000119103, 16697471000119102, 16697521000119104, 16745051000119107, 16745131000119100, 16745211000119100, 16745411000119104, 16745531000119108, 16745611000119102, 16745651000119101, 16745891000119105, 16745931000119102, 16746131000119105, 16746261000119108, 16746341000119103, 16746661000119106, 16746741000119102, 16746901000119101, 16746941000119104, 16747021000119109, 16747101000119101, 16747141000119104, 16747341000119101, 16747661000119109, 16747741000119100, 16747901000119104, 16748141000119100, 16748741000119101, 16749661000119102, 16749781000119101, 368201000119100, 368421000119108, 368431000119106, 368441000119102, 368451000119100, 368461000119103, 368471000119109, 368541000119101, 1196922005, 1196923000

#### Gestational diabetes

ICD-9-CM: 648.8

ICD-10-CM: O24.410, O24.414, O24.415, O24.419, O24.420, O24.424, O24.429, O24.430, O24.434, O24.435, O24.439

SNOMED CT: 11687002, 40801000119106, 46894009, 75022004, 40791000119105, 10753491000119101

#### Neuromuscular conditions

ICD-10-CM: G40, G70, O99.35, G40.0, G40.00, G40.001, G40.009, G40.01, G40.011, G40.019, G40.1, G40.10, G40.101, G40.109, G40.11, G40.111, G40.119, G40.2, G40.20, G40.201, G40.209, G40.21, G40.50, G40.211, G40.219, G40.3, G40.30, G40.301, G40.309, G40.31, G40.311, G40.319, G40.4, G40.40, G40.401, G40.409, G40.41, G40.411, G40.419, G40.5, G40.501, G40.509, G40.8, G40.80, G40.801, G40.802, G40.803, G40.804, G40.81, G40.811, G40.812, G40.813, G40.814, G40.82, G40.821, G40.822, G40.823, G40.824, G40.89, G40.9, G40.90, G40.901, G40.909, G40.91, G40.911, G40.919, G40.A, G40.A0, G40.A01, G40.A09, G40.A1, G40.A11, G40.A19, G40.B, G40.B0, G40.B01, G40.B09, G40.B1, G40.B11, G40.B19, G70.0, G70.00, G70.01, G70.1, G70.2, G70.8, G70.80, G70.81, G70.89, G70.9, O99.350, O99.351, O99.352, O99.353, O99.354, O99.355, G40.42, G40.83, G40.833, G40.834

ICD-9-CM: 345, 358

SNOMED CT: 84757009, 199297006, 257277002, 1172593006, 1187042007, 1179294000, 1163529009, 1163527006, 1197587003, 1179359005, 1208615009, 1208939001, 1228857005, 1222659003, 1222704008, 1231282002, 1187278006, 1179282009, 1167371007, 16415791000119104, 2198002, 241006, 5127009, 6204001, 7033004, 13973009, 15523002, 19598007, 26609002, 28055006, 31839002, 80976008, 82178003, 87842000, 91637004, 95208000, 109478007, 111498005, 1156827007, 787172004, 788417006, 860804005, 860806007, 860815000, 3371000119106, 5321000124109, 8291000119107, 14401000119109, 20121000119105, 21391000119102, 47391000119107, 77461000119109, 77471000119103, 82381000119103, 84161000119100, 84171000119106, 84181000119109, 84191000119107, 84201000119105, 84211000119108, 84221000119101, 84231000119103, 100941000119100, 119001000119108, 187931000119106, 290671000119100, 290681000119102, 290691000119104, 290711000119101, 290721000119108, 290731000119106, 290741000119102, 290761000119103, 290871000119101, 290881000119103, 291311000119108, 292991000119106, 329991000119104, 430811000124100, 430821000124108, 431071000124107, 431081000124105, 431091000124108, 431101000124102, 431111000124104, 431131000124105, 431141000124100, 431151000124103, 431161000124101, 431461000124109, 431471000124102, 431961000124104, 431971000124106, 431991000124107, 432031000124101, 432151000124107, 432161000124109, 432171000124102, 432291000124105, 432461000124101, 432471000124108, 432541000124104, 434201000124105, 434211000124108, 434291000124103, 434491000124104, 434501000124107, 434511000124105, 434541000124109, 434551000124106, 435321000124106, 435331000124109, 435341000124104, 435361000124100, 436011000124108, 438411000124101, 438421000124109, 440211000124100, 442151000124108, 442161000124105, 444441000124109, 444451000124106, 10750951000119106, 15729961000119108, 15985351000119105, 16005191000119106, 16005231000119102, 322112361000132104, 29145002, 44145005, 56989000, 58756001, 230299004, 230300007, 230301006, 230387008, 230388003, 230438007, 431121000124107, 445355009, 608874000, 698021005, 698760002, 698761003, 698762005, 698763000, 698764006, 698767004, 699688008, 702326000, 703524005, 710046001, 715534008, 715629001, 716278005, 716706009, 717223008, 717225001, 717266001,

720519003, 721088003, 722386009, 722432000, 723125008, 723676007, 724274009, 724549005, 724576005, 724785007, 724786008, 724787004, 724788009, 724789001, 724988000, 724989008, 724990004, 724991000, 724992007, 725163002, 725413002, 726702005, 733031004, 733032006, 733082001, 733195008, 733623005, 734434007, 763349002, 763352005, 763534009, 763622006, 763632004, 763688008, 763802009, 763827002, 764453009, 764522009, 765093009, 765170001, 765216006, 766044005, 766815007, 767254005, 768666006, 770405003, 770431001, 770438007, 770596007, 770622009, 770623004, 770624005, 770643005, 770758009, 770898002, 771141002, 771142009, 771223000, 771261002, 771448004, 771469002, 773421009, 773497001, 773498006, 773548008, 773643006, 774153002, 774154008, 778001003, 778047006, 778063003, 780827006, 782737003, 782752005, 782772000, 782825008, 783055005, 783062001, 783064000, 783139000, 783739005, 784342008, 784372002, 784377008, 785726009, 163594004, 192845009, 192979009, 192990004, 192999003, 193000002, 193002005, 193003000, 193008009, 193009001, 193010006, 193021002, 193022009, 193206003, 193207007, 205294008, 307356008, 307357004, 352818000, 361123003, 361268000, 363222009, 230191005, 230381009, 230382002, 230383007, 230384001, 230386004, 230389006, 230390002, 230391003, 230392005, 230393000, 230394006, 230395007, 230396008, 230397004, 230398009, 230399001, 230400008, 230401007, 230403005, 230404004, 230405003, 230406002, 230407006, 230408001, 230410004, 230411000, 230412007, 230413002, 230414008, 230415009, 230416005, 230417001, 230418006, 230419003, 230420009, 230421008, 230422001, 230423006, 230425004, 230426003, 230427007, 230428002, 230429005, 230430000, 230435005, 230437002, 230439004, 230440002, 230441003, 230443000, 230444006, 230445007, 230447004, 230448009, 230450001, 230452009, 230453004, 230454005, 230455006, 230456007, 230457003, 230458008, 230459000, 230460005, 230669004, 230670003, 230671004, 230672006, 230673001, 230674007, 230675008, 230676009, 230677000, 230678005, 230679002, 230682007, 230683002, 230684008, 230685009, 230686005, 230687001, 237612000, 36803009, 37356005, 38281008, 39194005, 39745004, 40816002, 49776008, 50866000, 55051001, 63135006, 65120008, 65242001, 67725009, 71831005, 75023009, 79745005, 413101007, 414927004, 422513000, 422724001, 422873003, 423753005, 425054007, 425237009, 425349008, 441678004, 442481002, 442512002, 267581004, 267592003, 277190002, 278510009, 1222656005, 1179360000, 1230376005, 1179547007, 16005311000119104, 1222662000

##### Major mental health conditions

ICD-10-CM: F20, F21, F22, F23, F24, F25, F28, F29, F30, F31, F32, F33, F34, F39, O99.34, F32.A, F20.0, F20.1, F20.2, F20.3, F20.5, F20.8, F20.81, F20.89, F20.9, F25.0, F25.1, F25.8, F25.9, F30.1, F30.10, F30.11, F30.12, F30.13, F30.2, F30.3, F30.4, F30.8, F30.9, F31.0, F31.1, F31.10, F31.11, F31.12, F31.13, F31.2, F31.3, F31.30, F31.31, F31.32, F31.4, F31.5, F31.6, F31.60, F31.61, F31.62, F31.63, F31.64, F31.7, F31.70, F31.71, F31.72, F31.73, F31.74, F31.75, F31.76, F31.77, F31.78, F31.8, F31.81, F31.89, F31.9, F32.0, F32.1, F32.2, F32.3, F32.4, F32.5, F32.8, F32.81, F32.89, F32.9, F33.0, F33.1, F33.2, F33.3, F33.4, F33.40, F33.41, F33.42, F33.8, F33.9, F34.0, F34.1, F34.8, F34.81, F34.89, F34.9, O99.340, O99.341, O99.342, O99.343, O99.344, O99.345

ICD-9-CM: 295, 296, 297, 298

SNOMED CT: 5464005, 13746004, 31027006, 36923009, 46206005, 48500005, 58214004, 61831009, 66344007, 68890003, 69322001, 268619003, 1231503005, 596004, 832007, 1196001, 1383008, 1499003, 1973000, 2506003, 2618002, 3109008, 162004, 4441000, 4926007, 5510009, 5703000, 7025000, 7052005, 9167000, 10327003, 10875004, 10981006, 12939007, 12969000, 13313007, 13581000, 14070001, 14144000, 14183003, 14291003, 14495005, 15193003, 15639000, 16295005, 16506000, 16990005, 17782008, 18260003, 18573003, 18818009, 19300006, 19527009, 19694002, 20250007, 20385005, 20960007,

21900002, 22121000, 22407005, 22419002, 23645006, 25922000, 26025008, 26203008, 26472000, 26516009, 26530004, 27387000, 28357009, 28475009, 28663008, 28676002, 28864000, 28884001, 29599000, 29733004, 29929003, 30336007, 30491001, 30520009, 30605009, 30687003, 30935000, 31446002, 31658008, 32358001, 32552001, 33078009, 33135002, 33323008, 33380008, 33736005, 34315001, 81319007, 82218004, 82998009, 83176005, 83458005, 83501007, 83746006, 84760002, 84788008, 85080004, 85248005, 85861002, 86058007, 87203005, 87414006, 87512008, 87842000, 87950005, 88939009, 88975006, 89618007, 111482003, 111483008, 111484002, 111485001, 129602009, 129604005, 1153570009, 1153575004, 65971000052100, 788120007, 788983007, 789061003, 789119007, 838528007, 838529004, 838530009, 251000119105, 261000119107, 271000119101, 281000119103, 7761000119106, 10211000132109, 21071000119101, 21831000119109, 22981000119106, 23741000119105, 31081000119101, 60401000119104, 94631000119100, 94921000119107, 104851000119103, 133091000119105, 133121000119109, 153071000119108, 288511000119108, 288751000119101, 288851000119106, 288861000119108, 10760421000119102, 10760461000119107, 10811121000119102, 10811161000119107, 10835871000119104, 12398321000119100, 15921731000119106, 16238181000119101, 16238221000119109, 16238741000119105, 16264621000119109, 16264821000119108, 16264901000119109, 16265061000119105, 16265301000119106, 16265951000119109, 16266831000119100, 16266991000119108, 786120041000132108, 3530005, 4863002, 9340000, 17262008, 31373002, 35489007, 49468007, 55009008, 68569003, 75360000, 78269000, 83225003, 191471000, 357705009, 371596008, 450714000, 473452003, 698946008, 698951002, 698957003, 712823008, 712824002, 712850003, 715924009, 718636001, 719592004, 719593009, 719717006, 720451004, 720452006, 720453001, 720454007, 720455008, 723899008, 723900003, 723901004, 723903001, 723905008, 723912004, 723913009, 723927004, 723928009, 723929001, 723930006, 723936000, 724655005, 724673008, 724674002, 724675001, 724676000, 724677009, 724678004, 724679007, 724680005, 724681009, 724682002, 724683007, 724684001, 724689006, 724690002, 724691003, 724692005, 724696008, 724702008, 724706006, 724707002, 724718002, 724719005, 724720004, 724721000, 724729003, 724755002, 724756001, 724757005, 724758000, 724759008, 724760003, 726772006, 735750005, 737225007, 737340007, 762320004, 762321000, 762322007, 762325009, 762326005, 762327001, 762328006, 762329003, 762330008, 762335003, 762336002, 762337006, 762338001, 762339009, 762340006, 762344002, 762345001, 762346000, 762507003, 762508008, 762509000, 762510005, 762511009, 762512002, 762513007, 762514001, 765176007, 767631007, 767632000, 767633005, 767635003, 767636002, 162313000, 162722001, 191447007, 191475009, 191478006, 191483003, 191484009, 191485005, 191486006, 191494004, 191495003, 191499009, 191525009, 191526005, 191527001, 191531007, 191542003, 191547009, 191548004, 191554003, 191555002, 191559008, 191561004, 191562006, 191563001, 191564007, 191565008, 191567000, 191569002, 191570001, 191571002, 191572009, 191574005, 191577003, 191583000, 191584006, 191586008, 191588009, 191590005, 191592002, 191593007, 191595000, 191597008, 191604000, 191610000, 191611001, 191613003, 191616006, 191618007, 191620005, 191621009, 191623007, 191625000, 191627008, 191629006, 191630001, 191634005, 191636007, 191638008, 191639000, 191641004, 191643001, 191658009, 191659001, 191667009, 191668004, 191670008, 191672000, 191676002, 191678001, 191680007, 192079006, 192080009, 192362008, 307417003, 310495003, 310496002, 310497006, 319768000, 320751009, 321717001, 370143000, 371024007, 371026009, 371599001, 371600003, 371604007, 231437006, 231438001, 231443008, 231444002, 231446000, 231449007, 231450007, 231451006, 231485007, 231487004, 231489001, 231494001, 231495000, 231496004, 231499006, 231500002, 231504006, 231542000, 237349002, 237350002, 237351003, 237352005, 238972008, 238973003, 238974009, 238975005, 238977002, 238978007, 238979004, 247803002, 247804008, 35218008, 35252006, 35481005, 35722002, 35846004, 36158005, 36170009, 36474008, 36583000, 36622002, 37331004, 37739004, 38295006, 38368003, 38451003, 38547003, 38694004, 39610001, 39809009, 40379007, 40568001, 40926005, 41552001, 41832009, 41836007, 41932008, 42344001, 42594001, 42810003, 42868002, 42925002, 43568002, 43769008, 45479006, 46229002, 46244001, 47447001, 47664006, 48589009, 48937005, 49512000, 50722006,

50933003, 51133006, 51637008, 53049002, 53607008, 53936005, 54502004, 54761006, 55516002, 55736003, 58329000, 58647003, 58703003, 59617007, 60099002, 60123008, 61144001, 61403008, 63181006, 63204009, 63249007, 63412003, 63649001, 63778009, 64731001, 64905009, 65042007, 65179007, 66631006, 67002003, 67711008, 68019004, 68995007, 69392006, 69482004, 70043001, 70546001, 70747007, 70814008, 71103003, 71294008, 71336009, 71984005, 73471000, 73867007, 74266001, 74686005, 75084000, 75122001, 75752004, 75837004, 76105009, 76441001, 76566000, 77475008, 77486005, 77911002, 78640000, 78667006, 79204003, 79298009, 79584002, 79842004, 79866005, 403595006, 408858002, 416340002, 426578000, 427975003, 429672007, 430852001, 441704009, 441833000, 442057004, 268612007, 268617001, 268621008, 268622001, 268624000, 270901009, 271428004, 274948002, 274952002, 274953007, 278506006, 278508007, 278852008, 278853003, 279225001, 280949006, 300706003, 301643003, 302507002, 1204417003, 1220650006

#### Hyperthyroid

ICD-10-CM: E05.00, E05.01, E05.10, E05.11, E05.20, E05.21, E05.30, E05.31, E05.40, E05.41, E05.80, E05.81, E05.90, E05.91

ICD-9-CM: 242.00, 242.01, 242.10, 242.11, 242.20, 242.21, 242.30, 242.31, 242.40, 242.41, 242.80, 242.81, 242.90, 242.91

SNOMED CT: 4997005, 22350004, 25008002, 26389007, 87232008, 88740003, 89719007, 90739004, 27538003, 237512007, 360361000, 190255006, 190256007, 193214009, 360353005, 360358001, 237499004, 237501007, 237503005, 237505003, 237506002, 237507006, 237508001, 237509009, 237510004, 237538007, 62052002, 68437005, 73869005, 431588009, 267374005, 286909009, 1163454006, 5834008, 13795004, 15470004, 26151008, 29028009, 30985009, 67761000119108, 137421000119106, 368361000119108, 1740001000004102, 479003, 353295004, 703309000, 190241003, 190242005, 190244006, 190247004, 237498007, 237500008, 237504004, 237824009, 40607004, 55807009, 57777000, 59957008, 60216004, 60268006, 62278002, 66628005, 67723002, 69329005

#### BMI ≥40

ICD-9-CM: V85.4

ICD-10-CM: Z68.4, Z68.41, Z68.42, Z68.43, Z68.44, Z68.45

SNOMED CT: 408512008

#### Cesarean section

ICD-9-CM: 654.2

ICD-10-CM: O34.21, O34.211, O34.212, O34.219, O34.218

SNOMED CT: 38451000119105, 74961000119106, 702736005, 706890009, 161805006, 200151008, 237297004, 302254004, 725949007, 725951006

#### Bariatric surger

ICD-10-CM: O99.84, O99.840, O99.841, O99.842, O99.843, O99.844, O99.845

SNOMED CT: 100841000119109

ICD-9-CM: 649.2

#### HIV/AIDS

ICD-9-CM: 042

ICD-10-CM: B20, B97.35, O98.7, O98.71, O98.711, O98.712, O98.713, O98.719, O98.72, O98.73, Z21

SNOMED CT: 86406008, 87117006, 91947003, 91948008, 111880001, 81000119104, 80191000119101, 90681000119107, 90691000119105, 10755671000119100, 15928141000119107, 315019000, 402901009, 414376003, 414604009, 416729007, 420244003, 420281004, 420302007, 420308006, 420321004, 420403001, 420452002, 420543008, 420544002, 420549007, 420554003, 420614009, 420658009, 420691000, 420718004, 420721002, 420764009, 420774007, 420787001, 420801006, 420818005, 420877009, 420900006, 420938005, 420945005, 421023003, 421047005, 421102007, 421230000, 421272004, 421283008, 421312009, 421315006, 421394009, 421403008, 421415007, 421431004, 421454008, 421460008, 421508002, 421529006, 421571007, 421597001, 421660003, 421671002, 421706001, 421708000, 421710003, 421766003, 421827003, 421851008, 421874007, 421883002, 421929001, 421983003, 421998001, 422003001, 422012004, 422089004, 422136003, 422177004, 422189002, 422194002, 422282000, 422337001, 697904001, 697965002, 700053002, 713260006, 713275003, 713278001, 713297001, 713298006, 713299003, 713300006, 713316008, 713318009, 713320007, 713325002, 713339002, 713340000, 713341001, 713342008, 713349004, 713444005, 713445006, 713446007, 713483007, 713484001, 713487008, 713488003, 713489006, 713490002, 713491003, 713497004, 713503007, 713504001, 713505000, 713506004, 713507008, 713508003, 713510001, 713511002, 713523008, 713526000, 713527009, 713530002, 713531003, 713532005, 713533000, 713543002, 713544008, 713545009, 713546005, 713570009, 713571008, 713572001, 713695001, 713696000, 713718006, 713722001, 713729005, 713730000, 713731001, 713732008, 713733003, 713734009, 713844000, 713845004, 713880000, 713881001, 713887002, 713897006, 713964006, 713967004, 714083007, 714464009, 719522009, 721166000, 722557007, 733834006, 733835007, 735521001, 735522008, 735523003, 735524009, 735525005, 735526006, 735527002, 735528007, 765434008, 771119002, 771126002, 771127006, 186706006, 186707002, 186708007, 230180003, 230598008, 235009000, 235726002, 236406007, 240103002, 40780007, 48794007, 52079000, 62479008, 79019005, 397763006, 398329009, 405631006, 420384005, 420395004, 420524008, 421020000, 421077004, 422127002, 442134007, 442537007, 276665006, 276666007, 281388009, 1187000005, 1187001009, 95892003, 1142045004, 1142055000, 1153348007, 838377003, 840442003, 840498003, 72631000119101, 76981000119106, 76991000119109, 10746341000119109, 420687005, 72621000119104, 445945000, 230202002

#### Lupus

ICD-10-CM: M32.0, M32.10, M32.11, M32.12, M32.13, M32.14, M32.15, M32.19, M32.8, M32.9

ICD-9-CM: 710.0

SNOMED CT: 4676006, 11013005, 19682006, 22888007, 25380002, 95332009, 95644001, 196138005, 200936003, 201436003, 309762007, 239886003, 239887007, 239888002, 239889005, 239890001, 36402006, 52042003, 54072008, 54912002, 55464009, 61458006, 68815009, 73286009, 76521009, 77753005, 80258006, 403486000, 403487009, 95609003, 295101000119105, 295111000119108, 295121000119101, 698694005, 707301001, 713225000, 724767000, 724781003, 773333003, 403488004
